# An Oral SARS-CoV-2 M^pro^ Inhibitor Clinical Candidate for the Treatment of COVID-19

**DOI:** 10.1101/2021.07.28.21261232

**Authors:** Dafydd R. Owen, Charlotte M. N. Allerton, Annaliesa S. Anderson, Lisa Aschenbrenner, Melissa Avery, Simon Berritt, Britton Boras, Rhonda D. Cardin, Anthony Carlo, Karen J. Coffman, Alyssa Dantonio, Li Di, Heather Eng, RoseAnn Ferre, Ketan S. Gajiwala, Scott A. Gibson, Samantha E. Greasley, Brett L. Hurst, Eugene P. Kadar, Amit S. Kalgutkar, Jack C. Lee, Jisun Lee, Wei Liu, Stephen W. Mason, Stephen Noell, Jonathan J. Novak, R. Scott Obach, Kevin Ogilvie, Nandini C. Patel, Martin Pettersson, Devendra K. Rai, Matthew R. Reese, Matthew F. Sammons, Jean G. Sathish, Ravi Shankar P. Singh, Claire M. Steppan, Al E. Stewart, Jamison B. Tuttle, Lawrence Updyke, Patrick R. Verhoest, Liuqing Wei, Qingyi Yang, Yuao Zhu

**Author notes:** Present address: Janssen Biopharma; South San Francisco, CA 94080. Present address: Praxis Precision Medicines; Cambridge, MA 02142. Present address: GRT Therapeutics; Cambridge, MA 02142.

## Abstract

The worldwide outbreak of coronavirus disease 2019 (COVID-19) caused by severe acute respiratory syndrome coronavirus 2 (SARS-CoV-2) has become an established global pandemic. Alongside vaccines, antiviral therapeutics are an important part of the healthcare response to counter the ongoing threat presented by COVID-19. Here, we report the discovery and characterization of PF-07321332, an orally bioavailable SARS-CoV-2 main protease inhibitor with in vitro pan-human coronavirus antiviral activity, and excellent off-target selectivity and in vivo safety profiles. PF-07321332 has demonstrated oral activity in a mouse-adapted SARS-CoV-2 model and has achieved oral plasma concentrations exceeding the in vitro antiviral cell potency, in a phase I clinical trial in healthy human participants.

ClinicalTrials.gov Identifier: NCT04756531

**One-Sentence Summary:** PF-07321332 is disclosed as a novel, orally active, investigational small-molecule inhibitor of the SARS-CoV-2 main protease, which is being evaluated in clinical trials for the treatment of COVID-19.

## Main Text

Human coronavirus infections are common, with at least four examples (229E, NL63, OC43, HKU1) now considered endemic (*1*). However, the emergence within the last 20 years of SARS-CoV-1, Middle East Respiratory Syndrome (MERS-CoV) and SARS-CoV-2 as novel human coronaviruses has signaled the significant threat potential of this viral class. The catastrophic SARS-CoV-2 outbreak of 2019 has resulted in 185 million confirmed cases of COVID-19 causing over 4 million deaths globally as of July 2021. SARS-CoV-2 is a highly infectious, ribonucleic acid (RNA) beta coronavirus that can cause life-threatening viral pneumonia in the most serious cases. While effective COVID-19 vaccines have been developed within unprecedented timelines, a significant number of people are either unable, due to pre-existing medical conditions, or unwilling to be vaccinated, such that global access challenges remain.

Limited therapeutic options are available to those who are infected. Oral SARS-CoV-2 specific therapeutics that are applicable for treatment of the broad population upon COVID-19 diagnosis are urgently needed. Such a treatment approach may prevent more severe disease, hospitalization and death. Indirectly, it may also reduce further transmission from infected individuals.

Repurposing of approved drugs in the search for small molecule antiviral agents that target SARS-CoV-2 has thus far been minimally effective (*2, 3*), however viral RNA-dependent RNA polymerase inhibitors such as molnupiravir and AT-527 are currently undergoing clinical trials for the treatment of COVID-19 in patients (*4, 5*).

The SARS-CoV-2 genome encodes two polyproteins (pp1a and pp1ab) and four structural proteins (*6, 7*). These polyproteins are cleaved by the critical SARS-CoV-2 main protease (M^pro^, also referred to as 3CL protease) at eleven different sites to yield shorter, non-structural proteins vital to viral replication (*8, 9*). The coronavirus M^pro^ is a three-domain cysteine protease, which features a Cys145-His41 catalytic dyad located in the cleft between domains I and II. Several common features are shared among M^pro^ substrates, including the presence of a P1 Gln residue. No known human equivalent cysteine protease exploits a P1 Gln as the cleavage site prompt, thus offering an intriguing selectivity hypothesis for this viral target over the human proteome (*10–12*). In addition, as the SARS-CoV-2 M^pro^ and the viral spike protein are distinct protein entities within the viral proteome, the antiviral efficacy of a small-molecule M^pro^ inhibitor is not expected to be affected by spike protein variants. Viral proteases have also proved to be tractable targets for small molecule oral therapies in the treatment of human immunodeficiency (HIV) and hepatitis C (HCV) viruses (*13, 14*). Moreover, a recent report has also demonstrated oral activity of an M^pro^ inhibitor in a transgenic mouse model of SARS-CoV-2 infection (*15*). Given the pivotal role of SARS-CoV-2 M^pro^ in viral replication, its potential for mechanistic safety, and expected lack of spike protein variant resistance challenges, SARS-CoV-2 M^pro^ inhibition represents an attractive small molecule approach for an oral antiviral therapy to treat COVID-19.

As part of the response to the 2002 SARS outbreak, an effort was made to identify inhibitors of the SARS-CoV-1 M^pro^, which led to the identification of PF-00835231 (**1**, Table 1) as a potent inhibitor of recombinant SARS-CoV-1 M^pro^ in a fluorescence resonance energy transfer (FRET)- based cleavage assay (*16*). PF-00835231 also demonstrated potent inhibition (inhibition constant (K_i_) = 0.271 nM) of recombinant SARS-CoV-2 M^pro^, as the SARS-CoV-1 and -CoV-2 M^pro^ share 100% sequence homology across their respective substrate binding sites (*17*). Antiviral activity against SARS-CoV-2 was also observed with PF-00835231 (half-maximal effective concentration (EC_50_) of 231 nM) in epithelial Vero E6 cells, when co-dosed in the presence of P-glycoprotein efflux inhibitor CP-100356 (*18*). The phosphate prodrug form (PF-07304814) of PF-00835231 is currently under investigation as an intravenous treatment option for COVID-19 in hospitalized patients (*18*).

**Table 1.**
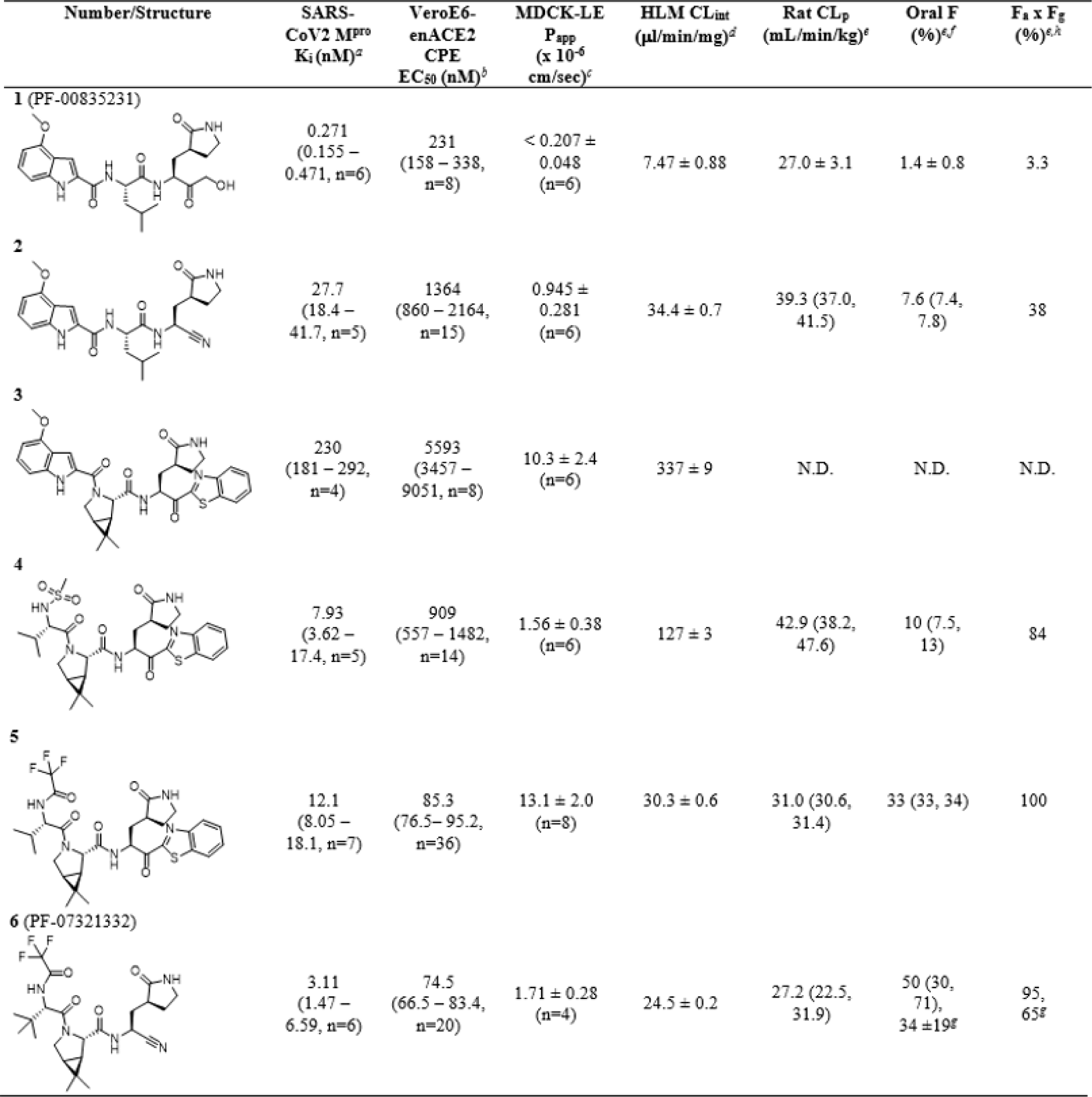
In vitro and in vivo parameters optimized in identifying oral SARS-CoV-2 M^pro^ inhibitors. ^a^Ki values were fit to the Morrison equation with substrate, Km and M^pro^ concentration parameters fixed to values described in the supplemental information. Data are geometric mean values with 95% confidence interval (CI) values and replicate numbers in parentheses. *^b^*EC50 values were calculated using data normalized to controls within the assay and fit to a 4-parameter logistic curve fit (see supplemental information for details). Data are geometric mean values with 95% CI values and replicate numbers in parentheses. *^c^*Apparent passive permeability (Papp) from apical to basolateral direction was determined in Madin-Darby canine kidney-low efflux (MDCK-LE) cells (*23*). *^d^*CLint refers to total intrinsic clearance obtained from scaling of half-lives of test compounds in NADPH-supplemented HLM (*25*). Incubations were conducted on a single day in triplicate. *^e^*Pharmacokinetic parameters were calculated from plasma concentration–time data and are reported as mean values (n= 2–3 male Wistar-Han rats/dosing route). See supplemental information for additional details. ^f^ Oral pharmacokinetics studies were conducted in the fed state. Oral bioavailability (F) is defined as the dose-normalized AUC after oral administration divided by the dose-normalized AUC after intravenous administration. ^g^Crystalline **6** was orally administered in anhydrous (Form 1) as well as anhydrous methyl-tert-butyl ether co-solvate form. ^h^The fraction of the oral dose absorbed (Fa x Fg) was estimated using the equation Fa x Fg = F/(1 – CLblood/Q) (*24*)) (see supplemental section for additional details).

| Number/Structure | SARS-CoV2 M<sup>pro</sup><br>K<sub>i</sub> (nM)<sup>a</sup> | VeroE6-enACE2<br>CPE<br>EC<sub>50</sub> (nM)<sup>b</sup> | MDCK-LE<br>P<sub>app</sub><br>(x 10<sup>-6</sup><br>cm/sec)<sup>c</sup> | HLM CL<sub>int</sub><br>(μl/min/mg)<sup>d</sup> | Rat CL<sub>p</sub><br>(mL/min/kg)<sup>e</sup> | Oral F<br>(%)<sup>e,f</sup> | F<sub>s</sub> x F<sub>g</sub><br>(%)<sup>e,h</sup> |
| --- | --- | --- | --- | --- | --- | --- | --- |
| **1** (PF-00835231) |  |  |  |  |  |  |  |
|  | 0.271<br>(0.155 –<br>0.471, n=6) | 231<br>(158 – 338,<br>n=8) | < 0.207 ±<br>0.048<br>(n=6) | 7.47 ± 0.88 | 27.0 ± 3.1 | 1.4 ± 0.8 | 3.3 |
| **2** |  |  |  |  |  |  |  |
|  | 27.7<br>(18.4 –<br>41.7, n=5) | 1364<br>(860 – 2164,<br>n=15) | 0.945 ±<br>0.281<br>(n=6) | 34.4 ± 0.7 | 39.3 (37.0,<br>41.5) | 7.6 (7.4,<br>7.8) | 38 |
| **3** |  |  |  |  |  |  |  |
|  | 230<br>(181 – 292,<br>n=4) | 5593<br>(3457 –<br>9051, n=8) | 10.3 ± 2.4<br>(n=6) | 337 ± 9 | N.D. | N.D. | N.D. |
| **4** |  |  |  |  |  |  |  |
|  | 7.93<br>(3.62 –<br>17.4, n=5) | 909<br>(557 – 1482,<br>n=14) | 1.56 ± 0.38<br>(n=6) | 127 ± 3 | 42.9 (38.2,<br>47.6) | 10 (7.5,<br>13) | 84 |
| **5** |  |  |  |  |  |  |  |
|  | 12.1<br>(8.05 –<br>18.1, n=7) | 85.3<br>(76.5 – 95.2,<br>n=36) | 13.1 ± 2.0<br>(n=8) | 30.3 ± 0.6 | 31.0 (30.6,<br>31.4) | 33 (33, 34) | 100 |
| **6** (PF-07321332) |  |  |  |  |  |  |  |
|  | 3.11<br>(1.47 –<br>6.59, n=6) | 74.5<br>(66.5 – 83.4,<br>n=20) | 1.71 ± 0.28<br>(n=4) | 24.5 ± 0.2 | 27.2 (22.5,<br>31.9) | 50 (30,<br>71),<br>34 ± 19<sup>g</sup> | 95,<br>65<sup>g</sup> |

During our oral SARS-CoV-2 M^pro^ inhibitor program, two functional groups precedented as covalent warheads for cysteine proteases were pursued in parallel: nitriles (*19, 20*) and benzothiazol-2-yl ketones (*21, 22*). These served to replace the hydrogen bond donor (HBD) of the P1’ α-hydroxymethyl ketone moiety in **1** (Fig. 1A) in order to improve upon the low passive absorptive permeability (P_app_ < 0.207 x 10^-6^ cm/s) (*23*) and poor oral absorption of **1** in animals (Table 1 and Table S1). Nitrile **2** demonstrated a significant increase in rat oral absorption (bioavailability (F) = 7.6% and fraction of oral dose absorbed (F_a_ x F_g_) = 38%) (*24*), while maintaining reasonable metabolic stability (intrinsic clearance (CL_int_)) towards oxidative metabolism in human liver microsomes (HLM) (*25*) relative to **1** (Table 1). However, the in vitro FRET M^pro^ potency (K_i_ = 27.7 nM) and SARS-CoV-2 antiviral activity (EC_50_ = 1364 nM) of **2** proved inferior in comparison to **1**. Introduction of a 6,6-dimethyl-3- azabicyclo[3.1.0]hexane as a cyclic leucine mimetic at P2 (Fig. 1B) further reduced HBD count. When combined with a P1’ benzothiazol-2-yl ketone, the resulting analog **3** (Table 1) displayed high passive absorptive permeability (P_app_ = 10.3 x 10^-6^ cm/s) The reduced biochemical SARS-CoV-2 M^pro^ inhibitory potency of **3** (K_i_ = 230 nM) relative to other reported benzothiazole-2-yl SARS-CoV-2 M^pro^ inhibitors (*26*) containing leucine P2 groups can be rationalized from the binding mode observed for **3** (Fig. 1C). While the 6,6-dimethyl-3-azabicyclo[3.1.0]hexane effectively fills the lipophilic S2 pocket formed by Met49, Met169, and His41, productive hydrogen bonding to Gln189 from a ligand backbone is no longer possible. The inferior SARS-CoV-2 M^pro^ potency and the high CL_int_ (337 ml/min/mg) precluded further investments in compound **3**. Similar to **1**, the P3 indole of **3** does not protrude into the S3 pocket (Figs. 1A and 1C). To better occupy the S3 pocket, we introduced branched, acyclic P3 groups. Methanesulfonamide **4** extends underneath Gln189, productively engaging P3 pocket residues and achieving improved hydrogen bonding interactions with the Glu166 backbone (Fig. 1D) relative to **1** and **3**. Methanesulfonamide **4** demonstrated improved SARS-CoV-2 M^pro^ biochemical potency (K_i_ = 7.93 nM), Vero E6 antiviral activity (EC_50_ = 909 nM), and HLM CL_int_ (127 ml/min/mg) relative to **3** (Table 1). Examination of the rat pharmacokinetics of **4** also revealed improvements in oral absorption (F = 10%, F_a_ x F_g_ = 84%) (Table 1). A further screen of P3 groups identified trifluoroacetamide **5**, which exhibited comparable biochemical potency to **4** (Compound **5**, K_i_ = 12.1 nM), but with greatly improved SARS-CoV-2 Vero E6 antiviral activity (EC_50_ = 85.3 nM) and passive permeability (P_app_ = 13.1 x 10^-6^ cm/sec) as well as increased metabolic stability in HLM (CL_int_ = 30.3 µl/min/mg) (Table 1). Trifluoroacetamide **5** also shows greatly improved oral pharmacokinetics in both rats (F = 33%, F_a_ x F_g_ = 100%) (Table 1) and monkeys (F = 7.9%, F_a_ x F_g_ = 66%) (Table S1). Introduction of the P1’ nitrile to this scaffold led to the identification of what would ultimately become the clinical candidate PF- 07321332 (**6**, Table 1). Compound **6** is a potent inhibitor of SARS-CoV-2 M^pro^ biochemical activity (K_i_ = 3.11 nM) with improved Vero E6 antiviral activity (EC_50_ = 74.5 nM) relative to compounds **2**-**4**. Compound **6** displays similar binding mode (Fig. 1E) relative to compound **4**. The P1’ nitrile of **6** forms a covalent thioimidate adduct with the catalytic Cys145 (Fig. 1F). To demonstrate the reversible nature of M^pro^ inhibition by **6**, we incubated SARS-CoV-2 M^pro^ (2 mM) in the presence of 2 mM of either **6** or an irreversible M^pro^ inhibitor (compound **7**, Fig. 2A) (*16*) for 30 minutes and monitored M^pro^ fractional velocity following 100-fold dilution of the incubation mixtures. No recovery of activity was observed following M^pro^ incubation with **7**. The recovery of >50% M^pro^ activity following incubation with **6** indicates that inhibition of SARS-CoV-2 M^pro^ is reversible (Fig. 2A). Faced with a choice between compounds **5** and **6**, we selected nitrile **6** as the clinical candidate (hereinafter referred to as PF-07321332) based on ease of synthetic scale-up, enhanced solubility that allowed for a simple formulation vehicle in support of pre-clinical toxicology, and reduced propensity for epimerization at the P1 stereocenter.

**Figure 1.**
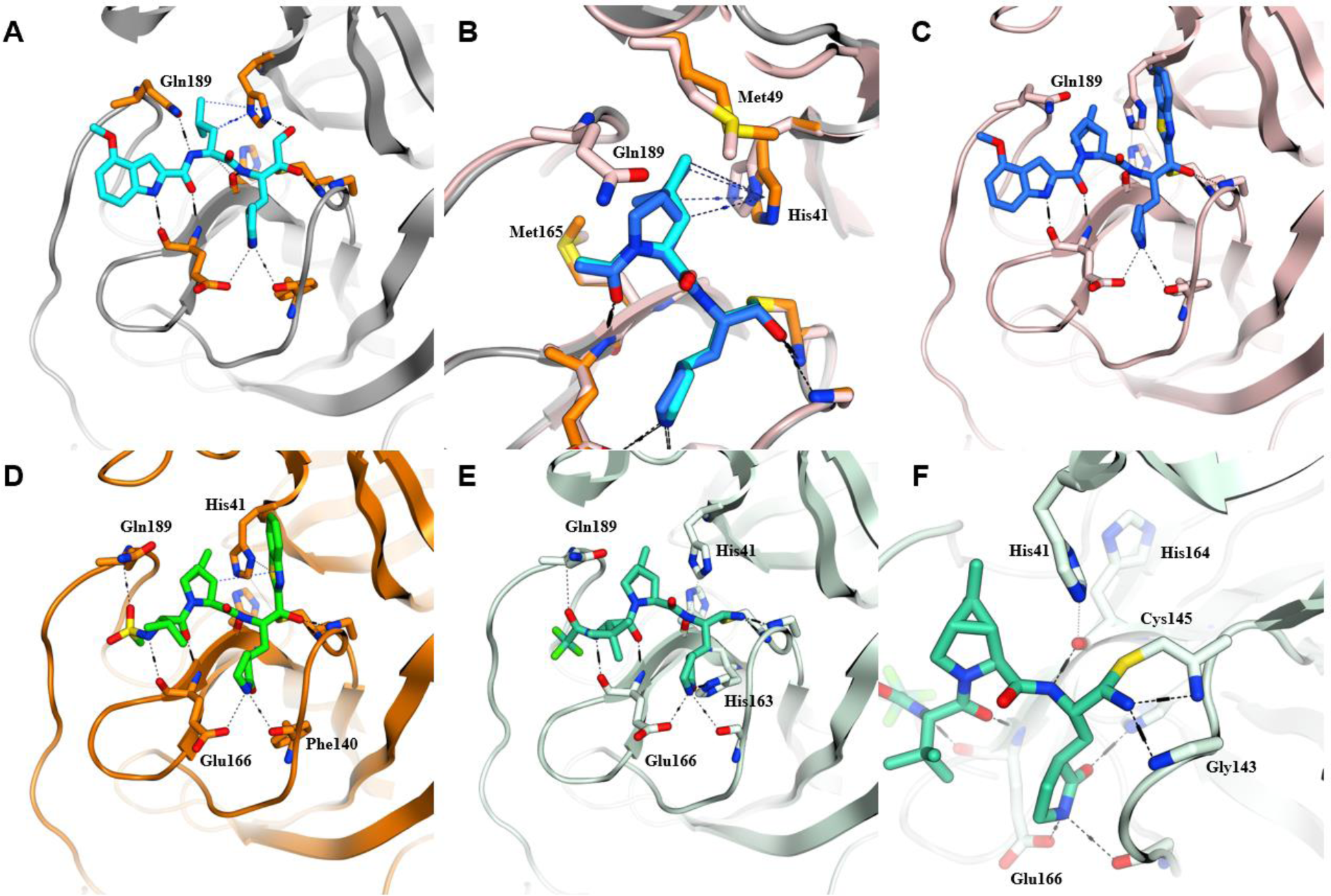
SARS-CoV-2 M^pro^ structural biology. (**A**) Key H-bond interactions of PF-00835231 (**1**). (**B**) Modelled overlap of dimethyl-bicyclo[3.1.0] proline from compound **3** (blue) as a mimic of P2 leucine residue (cyan) found in the viral polyprotein substrate and **1**. This tolerated P2 change eliminates an H-bond donor from resulting inhibitors. (**C**) Compound **3** effectively fills the lipophilic S2 pocket formed by Met49, Met165, and His41 but productive hydrogen bonding to Gln189 is no longer possible. (**D**) Compound **4** with optimized acyclic P3 group and restored Gln189 interaction. (**E**) Binding mode of clinical candidate PF-07321332 (**6**). A reversible covalent Cys145 adduct is formed with the reactive nitrile in compound **6**.

**Figure 2:**
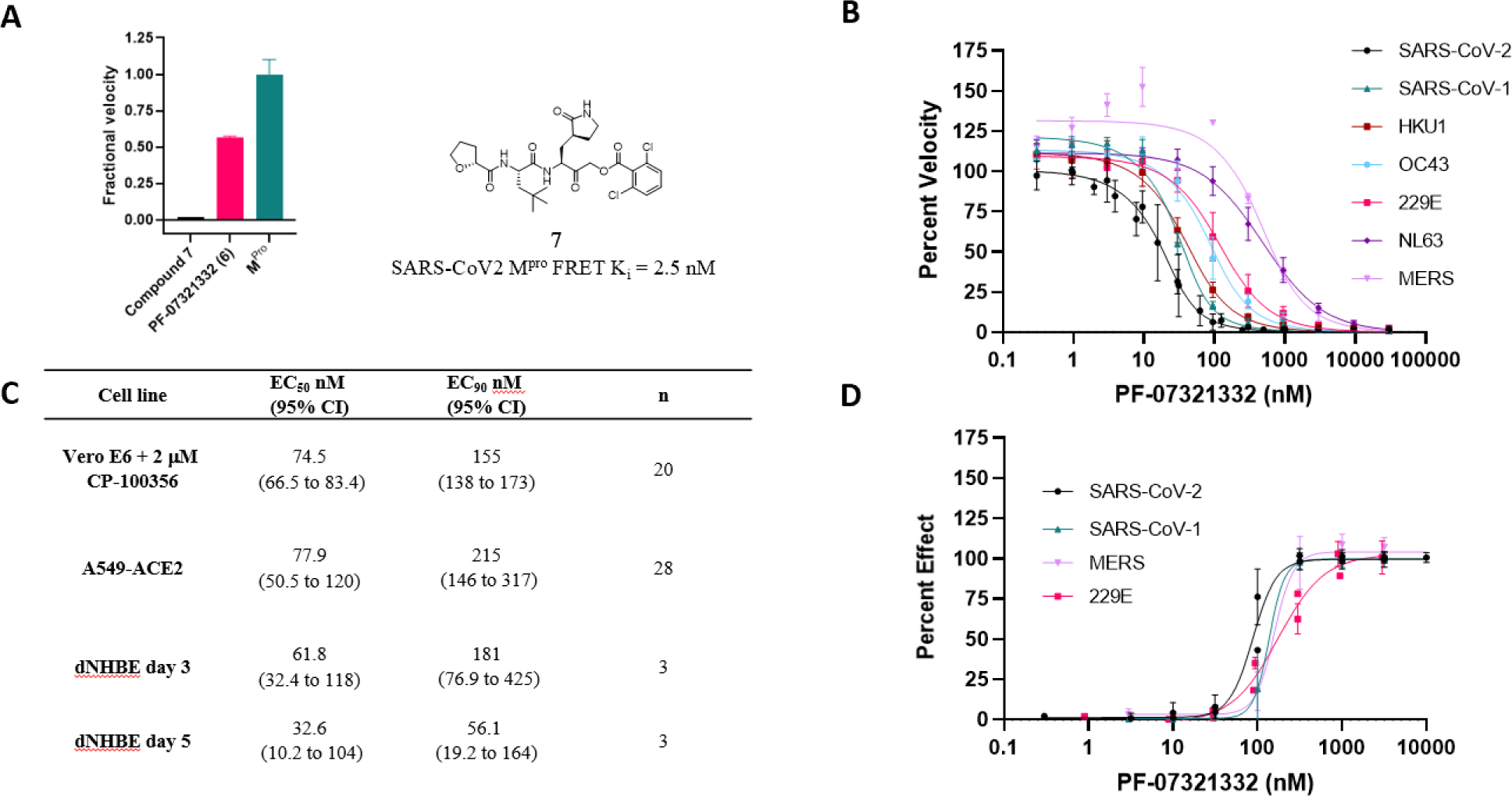
**PF-07321332 biochemical and antiviral activity**. (**A**) PF-07321332 is a reversible inhibitor as demonstrated by recovery of enzymatic activity following a 100-fold dilution of the enzyme inhibitor complex. Compound **7** (PF-00956378) an irreversible inhibitor, was included as a control. Data are representative of 3 independent experiments. (**B**) PF-07321332 is a potent inhibitor of the proteolytic activity of M^pro^ of SARS-CoV-2 as well as related coronaviruses in a FRET assay. Data shown are the mean ± SD from 3 independent experiments. (**C**) PF-07321332 inhibits SARS-CoV-2 induced cytopathic effect (CPE) in Vero E6 cells enriched for ACE2. A P-glycoprotein inhibitor, CP-100356 (efflux inhibitor, EI) was added at 2 μM to inhibit the efflux of PF-07321332. In VeroE6 cells with no EI, the EC50 (95% CI) was 4.48 μM (3.55 to 5.65 μM) (N=8, n=20). Cytotoxicity of PF- 07321332 was evaluated in non-infected cells and the CC50 was >100 μM. PF-07321332 inhibits SARS-CoV-2 replication in A549 cells expressing ACE2 (N=4). The CC50 in non-infected A549 cells was >3 μM, (N=4, n=14). In differentiated normal human bronchial epithelial (dNHBE) cells, PF-07321332 decreased SARS-CoV-2 viral replication (N=3). Data shown are the geometric mean and 95% confidence intervals (CI). (**D**) PF-07321332 inhibited SARS-CoV-1 induced CPE in Vero E6 cells (in the presence of 2 μM efflux inhibitor CP-100356), h-CoV-229E induced CPE in MRC-5 cells and MERS induced CPE in Vero 81 cells (in the presence of 1 μM efflux inhibitor CP-100356). Data shown are the mean and SD. CC50 values determined in all assays to be >100 μM.

PF-07321332 demonstrated potent inhibition in FRET M^pro^ assays representing all coronavirus types known to infect humans (*6, 7, 27*), including beta-coronaviruses (SARS-CoV-2, SARS-CoV-1, HKU1, OC43, MERS) as well as alpha-coronaviruses (229E, NL63) (Fig. 2B, Table S2). No inhibitory effects were noted against several mammalian cysteine (caspase 2), serine (chymotrypsin, elastase, thrombin) and aspartyl (cathepsin B, cathepsin D, cathepsin L, HIV-1) proteases at the highest concentration tested (100 mM ) of PF-07321332 (Table S3).

The in vitro antiviral activity of PF-07321332 was also evaluated in two physiologically relevant cellular assays, human adenocarcinoma-derived alveolar basal epithelial (A549) cells constitutively expressing ACE2 and differentiated normal human bronchial epithelial (dNHBE) cells (*28*). PF-07321332 inhibited SARS-CoV-2 replication in A549-ACE2 cells with EC_50_ and EC_90_ values of 77.9 nM and 215 nM, respectively, with no cytotoxicity detected at concentrations up to 3 µM (Fig. 2C). Treatment of dNHBE cells with varying concentrations of PF-07321332 for 3 days led to inhibition of SARS-CoV-2 viral replication with EC_50_ and EC_90_ values of 61.8 nM and 181 nM, respectively (Fig. 2C). Increasing the duration of the dNHBE study to 5 days saw viral replication inhibited further with EC_50_ and EC_90_ values of 32.6 nM and 56.1 nM (Fig. 2C). Since optimal therapeutic efficacy with marketed protease inhibitors and other antiviral agents is generally achieved when systemic unbound plasma concentrations at trough (C_min_) are maintained above cellular antiviral EC_90_ values (*29*), we selected the more conservative day 3 EC_90_ value of 181 nM for PF-07321332 in the dNHBE assay as the minimum unbound systemic concentration to be maintained at C_min_ when predicting efficacy in animal models and in the clinic. We further evaluated the in vitro cellular antiviral activity of PF-07321332 against SARS-CoV-1, MERS and human coronavirus 229E using cytopathic effect (CPE) assays. PF- 07321332 demonstrated potent antiviral activity against SARS-CoV-1 (EC_90_ = 317 nM), MERS (EC_90_ = 351 nM) and 229E (EC_90_ = 620 nM) in their respective cellular assays (Fig. 2D, Table S4).

We evaluated the in vivo antiviral activity of PF-07321332 in a mouse-adapted SARS-CoV-2 (SARS-Cov-2 MA10) model. Intranasal infection of BALB/c mice with SARS-CoV-2 MA10 leads to ∼10% body weight loss and minimal mortality in 10-week-old mice (*30*). As shown in Fig. 3A, following twice daily (BID) oral administration of PF-07321332 (300 mg/kg and 1000 mg/kg), SARS-CoV-2 MA10-infected mice were protected from weight loss versus placebo mice. At 4 days post infection, mice were sacrificed and lung viral titers were evaluated in 50% cell culture infective dose (CCID_50_) assays. Infected animals in the placebo group (n=12, two independent studies) had robust infection in the lungs (mean lung titer of 4.93 ± 0.140 CCID_50_ log_10_/ml SARS-CoV-2 MA10) (Fig. 3B) whereas virus levels in mice treated with PF-07321332 were significantly reduced (mean lung titers of 3.533 ± 0.187 and 3.02 ± 0.423 CCID_50_ log_10_/ml, respectively) for 300 mg/kg and 1000 mg/kg PF-07321332-treated groups. In a satellite group of uninfected mice, the 300 mg/kg BID dose of PF-07321332 used in the mouse-adapted viral in vivo efficacy study maintained C_min_ unbound plasma concentrations above ∼0.9 x EC_90_ (Fig. 3C) as determined in the dNHBE primary cell assay. The 1000 mg/kg BID dose of PF-07321332 maintained a C_min_ unbound plasma concentration of ∼4 x EC_90_ (Fig. 3C). This confirms that PF- 07321332 is effective at reducing SARS-CoV-2 MA10 viral load in mouse lungs at concentrations consistent with the observed in vitro anti-viral potency, and those being targeted clinically.

**Figure 3.**
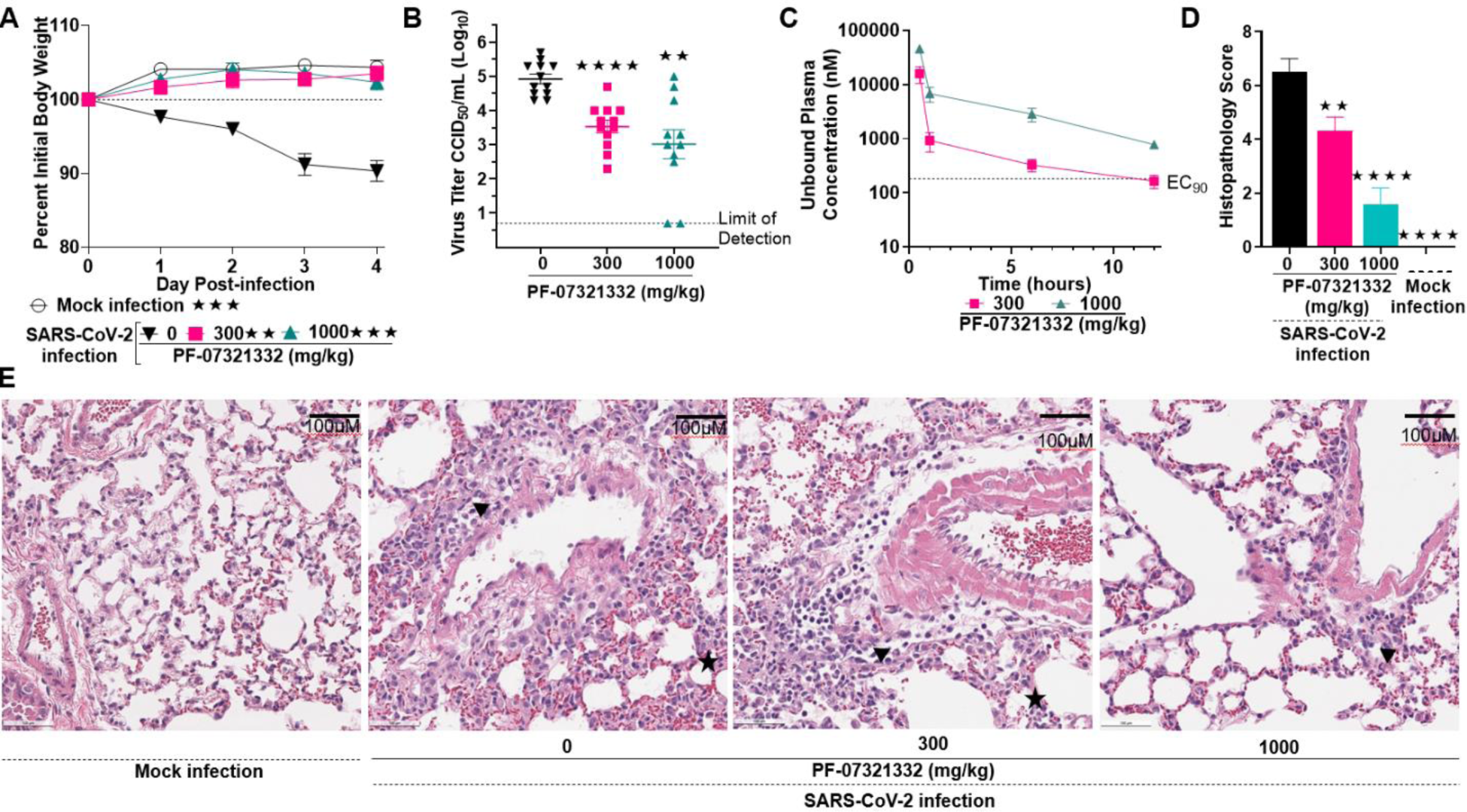
In vivo efficacy of PF-07321332 against SARS-CoV-2 MA10 infection in mice. Six-mice/group were challenged intranasally with 1 x 10^5.0^ 50% cell culture infectious doses (CCID50). Animals were orally administered 300 and 1000 mg/kg PF-07321332 or vehicle (placebo) BID beginning 4 hours post-infection. Animals were euthanized at 4 days post infection (dpi) and lungs collected for virus titers. Data for A-D are compiled from two independent studies (n=12, BALB/c mice). (**A**) Lung viral titer at 4 dpi. Lung titers are graphed as mean CCID50 log10 /ml +/- std error of mean. Dotted line represents the limit of detection for the CCID50 assay. (**B**) Weight loss during infection. Mice were weighed daily. (**C**) PF-07321332 exposure levels in uninfected, orally BID treated mice. (**D**) Histopathology scores on a scale of 0-5 where 0 is a normal healthy lung and 5 is severe coalescing areas of necrosis and confluent areas of inflammation. (**E**) Digital light microscopic scans of H&E-stained mouse lung tissue sections of mock, placebo, 300 mg/kg PF-07321332, and 1000 mg/kg PF-07321332 treated mice. Triangle indicates sites of perivascular inflammation and star indicates thickening of alveolar septum. Data in **E** are representative microscopic scans from one study. The black bar is 100 μM and magnification is 20x. For statistical analysis, one-way ANOVA with Brown-Forsythe and Welch test was performed. Groups were compared against placebo using Dunnett’s test. ** indicates p<0.01 and **** p<0.0001.

Disease in this model is manifest by weight loss due to pathological changes in the lungs of the infected mice (*30*). Histopathological analysis of lungs from the treated mice shows that PF- 07321332 protects lung tissue from damage due to virus replication and limits cellular infiltration (Figs. 3D, E). Histopathological evaluation of lungs from the untreated placebo mice demonstrates evidence of increased perivascular inflammation, bronchial or bronchiolar epithelial degeneration or necrosis, bronchial or bronchiolar inflammation, cellular debris in alveolar lumen, and alveolar inflammation and thickening of the alveolar septum compared to PF-07321332 treated mice and mock-infected mice. Most of the infected mice exhibited multifocal pulmonary lesions, which were significantly reduced in PF-07321332-treated mice.

PF-07321332 exhibited moderate plasma clearance (CL_p_) in rats and monkeys, with elimination half-lives (t_1/2_) of 5 h and <1 h, respectively, after intravenous dosing (Table S5). Following oral administration to rats, crystalline PF-07321332 (10 mg/kg) demonstrated oral F_a_ x F_g_ and F values ranging from 65-95% and 34-50% respectively, depending on the crystalline form used (Tables 1 and S5). Oral administration of PF-07321332 (10 mg/kg) to monkeys led to a relatively poor oral F of 8.5% (F_a_ x F_g_ = 20%) (Table S5), which is attributed to first-pass metabolism along the gastrointestinal tract by cytochrome P450 (CYP) enzymes, as PF-07321332 demonstrated rapid (t_1/2_ = 20.5 min, CL_int_ = 33.8 µl/min/mg) NADPH-dependent turnover in monkey intestinal microsomes (Table S6). PF-07321332 was resistant (t_1/2_ > 240 min, Cl_int_ <2.89 µl/min/mg) towards CYP-mediated metabolism in intestinal microsomes from rat and human. PF-07321332 (0.3–10 mM) exhibited concentration-independent plasma protein binding in rat (mean plasma unbound fraction (f_u,p_) = 0.478), monkey (mean f_u,p_ = 0.434), and human (mean f_u,p_ = 0.310) under equilibrium dialysis conditions (*31*).

Drug-metabolizing enzymes involved in the metabolism of PF-07321332 were also studied. In NADPH-supplemented HLM, PF-07321332 demonstrated moderate CL_int_ (24.5 ml/min/mg) (Table 1 and S7), which was significantly inhibited (≥82%) by the selective CYP3A4/5 inhibitor ketoconazole (*25*) (Table S7). Moreover, the oxidative metabolic profile of PF-07321332 in HLM, which includes modifications on the P2 6,6-dimethyl-3-azabicyclo[3.1.0]hexane, the *tert*- butyl group at the P3 position, and the P1 pyrrolidinone ring, was reproduced in incubations of PF-07321332 with recombinant human CYP3A4 (Figure S1). These in vitro studies, which established a predominant role for CYP3A4 in the metabolism of PF-07321332, also presented an opportunity to boost therapeutic concentrations of PF-07321332 in the clinic via co-dosing with the potent CYP3A4 inactivator ritonavir (RTV), which is used as a pharmacokinetic enhancer of several marketed protease inhibitors (e.g., darunavir, lopinavir, etc.) that are subject to metabolic clearance via CYP3A4 (*32, 33*).

PF-07321332 demonstrated a favorable off-target selectivity profile in a broad panel of G protein-coupled receptors, kinases, transporters and phosphodiesterase enzyme inhibitor screens, and was devoid of hERG, Ca^2+^ and Na^+^ channel binding and functional activity (Tables S8 and S9). PF-07321332 was not mutagenic or clastogenic in in vitro genetic toxicity studies, and was negative in an in vivo rat micronucleus assay (Table S10). Repeat oral dosing of PF-07321332 (40–1000 mg/kg) in 2-week regulatory toxicity studies in monkeys and rats led to dose-dependent increases in both maximal plasma concentrations (C_max_) and area-under-the-plasma concentration versus time curves (AUC) (Figs. 4A, 4B and Table S11). This resulted in unbound C_max_ and AUC margins of 273-fold and 64-fold in rats (day 14, 1000 mg/kg) and 510-fold and 245-fold in monkeys (day 15, 600 mg/kg) over the measured unbound EC_90_ value of PF- 07321332 determined in the SARS-CoV-2 dNHBE cellular assay. PF-07321332 was well tolerated with no adverse findings in either species; the corresponding no observed adverse effect levels (NOAEL) were the highest doses tested (600 mg/kg/day in monkeys and 1000 mg/kg/day in rats).

**Figure 4.**
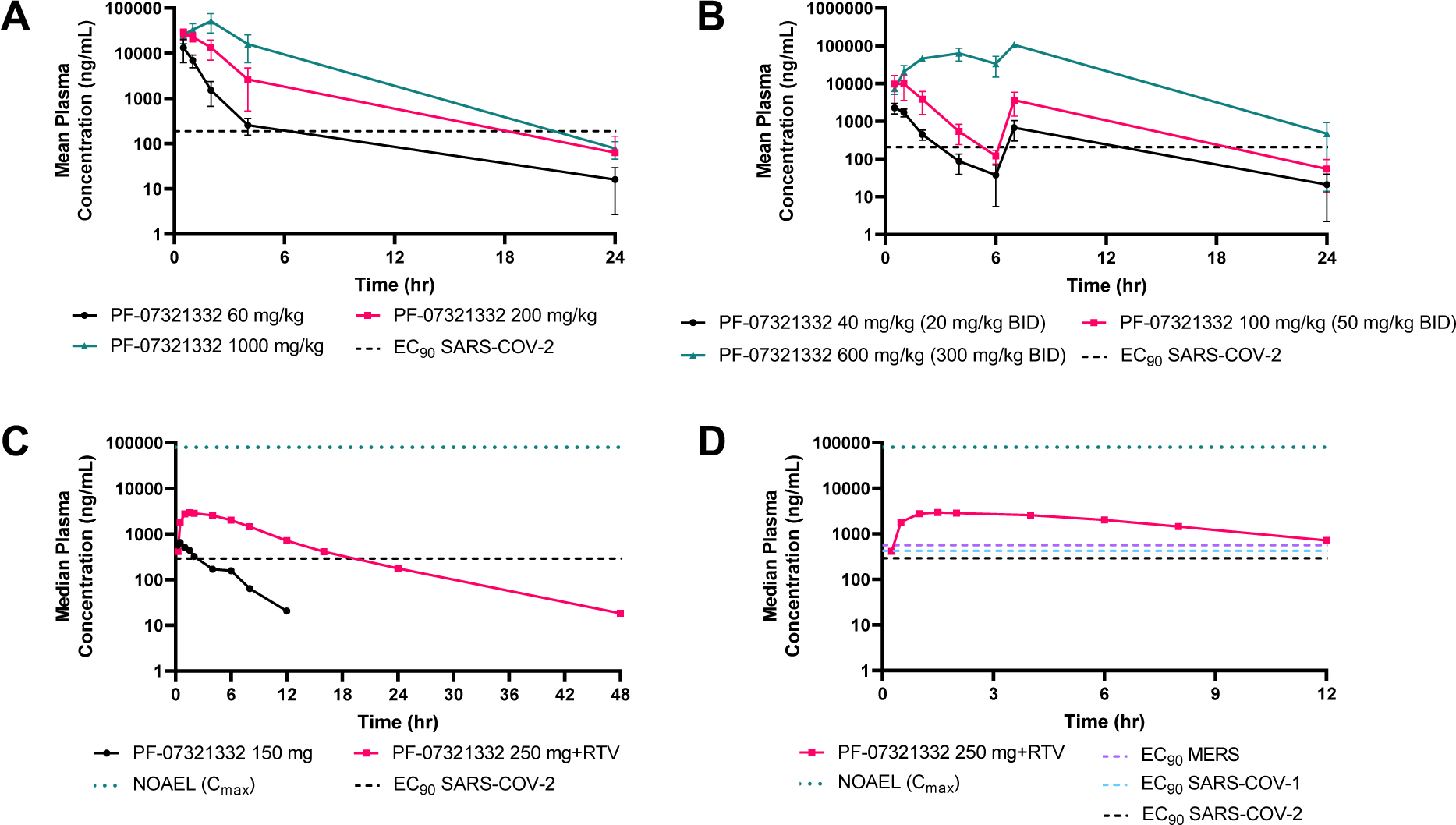
Preclinical toxicology and healthy adult participant single ascending dose study exposures for PF-07321332. (**A**) Rat oral toxicokinetic exposures (day 14) of once-daily administered PF-07321332 compared with day 3 antiviral EC90 values in dNHBE cells. (**B**) Monkey oral toxicokinetic exposures (day 15) of twice-daily administered PF-07321332 compared with day 3 antiviral EC90 in dNHBE. (**C**) Human plasma concentrations (total) versus time profile after oral administration (fasted state) of PF-07321332 (150 mg) and PF-07321332 (250 mg) with ritonavir (RTV) (100 mg at t = -12 h, 0 h and 12 h) compared with day 3 antiviral EC90 in dNHBE. (**D**) Human antiviral target coverage >EC90,u at 12 h for SARS- CoV-1, SARS-CoV-2 and MERS post oral administration of PF-07321332 (250 mg) and ritonavir (100 mg at t = -12 h, 0 h and 12 h). The in vitro unbound SARS-COV-2 EC90 of 181 nM was converted to ng/ml using a molecular weight of 499.5 g/mol. Total EC90 was calculated by dividing unbound EC90 by fu,plasma (rat 0.479, NHP 0.208, and human 0.310). This resulted in total EC90 values of 189, 208, and 292 ng/ml for rat, NHP, and human, respectively. The calculated total EC90 for MERS and SARS-COV-1 were 566 and 422 ng/ml, respectively. A human NOAEL of 79,700 ng/ml (for Cmax) was estimated from the rat Cmax value of 51500 ng/ml at the NOAEL dose of 1000 mg/kg, normalizing for plasma unbound fraction differences.

The safety, tolerability, and pharmacokinetics of PF-07321332 as a single agent and in combination with RTV are under investigation in a randomized, double-blind, placebo- controlled, single ascending dose study in healthy adult participants. At each dose tested, 4 participants were randomized to receive active treatment while 2 participants received placebo. In the PF-07321332/RTV co-administration dosing paradigm, each subject (active and placebo) received one tablet (100 mg) of RTV at -12 h, 0 h and 12 h. PF-07321332 was administered as an oral suspension under fasted conditions at 0 h (minimum fast of ∼10 h prior to treatment).

Preliminary plasma concentration versus time pharmacokinetics profile achieved from two oral doses, PF-07321332 (150 mg) alone and PF-07321332 (250 mg) with RTV are presented in Figure 4C. PF-07321332 was safe and well tolerated, and exhibited a significant boost in plasma concentrations when co-administered with RTV. Oral plasma concentrations of PF-07321332 (250 mg with RTV) were considerably above the SARS-COV-2 antiviral EC_90_ value (total EC_90_ = 292 ng/ml, unbound EC_90_ = 90.5 ng/ml, 181 nM) at 12 h post-dose enabling PF-07321332 to achieve multiples over the antiviral EC_90_ for SARS-CoV-2 and other coronaviruses, thus increasing confidence in achieving robust pan-coronavirus anti-viral activity clinically (Fig. 4D). Based on these observations and the high safety margins at the NOAEL doses in animals, the efficacy of PF-07321332 in COVID-19 patients will be assessed with a BID dosing paradigm with the potential to increase the dose of PF-07321332 as a single agent and/or co-administration with RTV.

## Data Availability

Upon request and subject to certain criteria, conditions and exceptions, Pfizer will provide access to de-identified participant data from Pfizer-sponsored global interventional clinical studies conducted for medicines, vaccines and medical devices for indications that have been approved in the US and/or the EU or in programs that have been terminated (i.e., development of all indications has been discontinued). Pfizer will also consider requests for the protocol, data dictionary, and statistical analysis plan. Data may be requested from Pfizer trials 24 months after completion. The de-identified participant data will be made available to researchers whose proposals meet the research criteria and other conditions, and for which an exception does not apply, via a secure portal. To gain access, requestors must enter into a data access agreement with Pfizer.

## Acknowledgments

The authors would like to thank the participants of the FIH study. We would also like to acknowledge the significant number of Pfizer colleagues who have contributed to the COVID-19 oral protease program across a number of disciplines. In particular, we acknowledge Sylvie Sakata, Joel Arcari, Jinzhi Zhang for external research resourcing, Kathleen Farley for NMR studies, Lorraine Lanyon for protease panel data, Yajing Lian, Erik LaChapelle, Stephen Wright, Steven O’Neil, Eddie Yang, John Humphrey, Brian Boscoe for compound synthesis, Steve Jenkinson for safety pharmacology data, Kevin Ryan for structural biology support, Elizabeth Collins, Chris Allais for FIH-enabling active pharmaceutical ingredient supply, Fran Clark for bioanalysis, Haihong Shi for clinical assay support, Gianluca Nucci, Art Bergman for first-in-human study design and clinical pharmacology, Frances Hackman for clinical statistics, Sima Toussi for medical monitoring, Karen Bartsch for clinical study leadership, Sylvester Pawlak for clinical trial leadership, Claudine Fredette for clinical trial project management, Ding Ding for regulatory documentation support, and Mikael Dolsten for scientific discussion and advice. This research used resources of the Advanced Photon Source, a U.S. Department of Energy (DOE) Office of Science User Facility, operated for the DOE Office of Science by Argonne National Laboratory under Contract No. DE-AC02-06CH11357. Extraordinary facility operations were supported in part by the DOE Office of Science through the National Virtual Biotechnology Laboratory, a consortium of DOE national laboratories focused on the response to COVID-19, with funding provided by the Coronavirus CARES Act.

## Funding

This study was sponsored by Pfizer, Inc.

## Author contributions

Conceptualization: CMNA, ASA, DRO, MP

Formal Analysis: LD, BB, SEG, QY, DKR, JJN, AD, RSO, RSPS

Investigation: MA, SB, JCL, JL, KO, LW, RF, KSG, WL, RSO, DKR, SAG, LA, SN, HE Methodology: JBT, QY, MFS, PRV, MRR, MP, DRO, NCP, AC, EPK, KJC, RSPS

Project Administration: DRO, MRR, JBT, ASK, CMS, LU, JGS, BLH, YZ, SWM, RDC, NCP

Resources: AD, JJN, KJC

Supervision: MRR, CES, AES, BLH, PRV, RDC

Visualization: SWM, DKR, SN, BB, LA, HE

Writing – original draft: DRO, ASK, MFS, CMS, RDC, MRR, JGS, CMS, YZ Writing – review & editing: DRO, ASK, MFS, ASA, CMNA

## Competing interests

DRO, CMNA, ASA, LA, MA, SB, BB, RDC, AC, KJC, AD, LD, HE, RF, KSG, SEG, EPK, ASK, JCL, JL, WL, SN, RSO, KO, NCP, DKR, MRR, MFS, JGS, RPS, CMS, AS, JBT, LU, PRV, LW, QY, YZ are employees of Pfizer and some of the authors are shareholders in Pfizer Inc. SWM, JJN, MP were employees of Pfizer Inc. during part of this study.

## Data and materials availability

Upon request and subject to certain criteria, conditions and exceptions, Pfizer will provide access to de-identified participant data from Pfizer-sponsored global interventional clinical studies conducted for medicines, vaccines and medical devices for indications that have been approved in the US and/or the EU or in programs that have been terminated (i.e., development of all indications has been discontinued). Pfizer will also consider requests for the protocol, data dictionary, and statistical analysis plan. Data may be requested from Pfizer trials 24 months after completion. The de-identified participant data will be made available to researchers whose proposals meet the research criteria and other conditions, and for which an exception does not apply, via a secure portal. To gain access, requestors must enter into a data access agreement with Pfizer. Toxicology study reports will not be publicly available until such a date that Pfizer deems them mature for release. All other non-clinical data are available in the main text or the supplementary materials. X-ray coordinates and structure factors are deposited at the RCSB Protein Data Bank under accession codes 7RFR, 7RFU, 7RFS and 7RFW. Pfizer shares compounds using requests via the ’Pure Compound Grants’ program see https://www.cybergrants.com/pfizer/Research.

**Supplementary Materials** Materials and Methods Tables S1-S11

## Supplementary Materials

### Materials and Methods

#### Abbreviations

ATP: adenosine triphosphate
APCI: atmospheric pressure chemical ionization
AUC: area under the plasma concentration-time curve
BID: bis in die (twice a day)
Boc: *tert*-butyloxycarbonyl
CCID: cell culture infective dose
CE: collision energy
CL_int_: intrinsic clearance
CL_p_: plasma clearance
CYP: cytochrome P450
DCM: dichloromethane, CH_2_Cl_2_
DIEA: N,N-diisopropylethyl amine, Hunig’s base
DMF: dimethylformamide
DMSO: dimethyl sulfoxide
DTT: dithiothreitol
EDCI: 1-[3-(dimethylamino)propyl]-3-ethylcarbodiimide hydrochloride
EDTA: ethylenediaminetetraacetic acid
ESCI: combined electrospray and atmospheric pressure ionization source
ESI: electrospray ionization
EtOAc: ethyl acetate
F_a_ x F_g_: fraction of the oral dose absorbed
FIH: first-in-human
FLIPR: fluorometric imaging plate reader
Oral F: oral bioavailability
f_u,p_: fraction unbound in plasma
GPCR: G-protein-coupled receptor
HATU: O-(7-azabenzotriazol-1-yl)-N,N,N’,N’-tetramethyluronium hexafluorophosphate
HEPES: 4-(2-hydroxyethyl)-1-piperazineethanesulfonic acid
HFIP: hexafluoroisopropanol
HLM: human liver microsomes
HOPO: 2-hydroxypyridine 1-oxide
HPLC: high performance liquid chromatography HRMS high resolution mass spectrometry
IPA: isopropanol
IPAC: isopropyl acetate
iv: intravenous
LB: Luria Broth
LCMS: liquid chromatography–mass spectrometry
LC-MS/MS: liquid chromatography tandem mass spectrometry
LLOQ: lower limit of quantitation
MeCN: acetonitrile
MEK: methyl ethyl ketone, butan-2-one
MeOH: methanol
MTBE: methyl tert-butyl ether
NADPH: nicotinamide adenine dinucleotide phosphate NMR nuclear magnetic resonance
PEG: polyethylene glycol
po: oral
PXRD: powder X ray diffraction
RTV: ritonavir
TCEP: tris(2-carboxyethyl)phosphine
TEA: triethylamine
TFA: trifluoroacetic acid
THF: tetrahydrofuran
TLC: thin layer chromatography
TOF: time-of-flight
TsOH: p-toluenesulfonic acid
UPLC: ultra-performance liquid chromatography
UV: ultraviolet
Vd_ss_: Steady state volume of distribution

#### General Methods for Compound Synthesis/Analysis

All reactants, reagents and solvents were obtained from commercial sources and used without further purification. Except where otherwise noted, reactions were run under an inert atmosphere of nitrogen gas using anhydrous solvents at room temperature (∼23 °C). The terms “concentrated” and “evaporated” refer to the removal of solvent at reduced pressure on a rotary evaporator with a water bath temperature not exceeding 60 °C. Reactions were monitored by thin layer chromatography (TLC) performed on Analtech, Inc. silica gel GF 250 μm plates or Merck silica gel plates (60 F254) and were visualized with UV light (254 nm) and/or KMnO_4_ staining or by UPLC-MS (Waters Acquity, ESCI (ESI +/-, APCI +/-)). Flash chromatography was carried out using a Combiflash system from Teledyne Isco; Biotage SNAP or Redisep Rf silica columns were used. All nuclear magnetic resonance (NMR) spectra were collected using a 600 MHz Bruker Avance III spectrometer, a 400 MHz Bruker Avance III spectrometer, or a 400 MHz JEOL ECZ spectrometer. Chemical shifts (d in ppm) for ^1^H spectra are reported relative to the residual solvent signals: 7.26 ppm for chloroform-*d*, 2.50 ppm for dimethyl sulfoxide-*d*_6_, and 3.31 ppm for methanol-*d*_4_ using the δ, multiplicity, coupling constant(s) in Hz and integration. The multiplicities are denoted as follows: s, singlet; d, doublet; t, triplet; q, quartet; m, multiplet; and br s, broad singlet. Chemical shifts (d in ppm) for ^13^C spectra are reported relative to the residual solvent signals: 39.5 ppm for dimethyl sulfoxide-*d*_6_. Chemical shifts (d in ppm) for ^19^F spectra are referenced using an external trifluoroacetic acid standard at -76.6 ppm. Residual EtOAc is present in many synthetic intermediates. High-resolution mass spectrometry (HRMS) data were gathered on a Sciex TripleTOF 5600+ (Sciex, Ontario, Canada)with DuoSpray ionization source. The LC instrument includes an Agilent (Agilent Technologies, Wilmington, DE) 1200 binary pump, Agilent 1200 autosampler, Agilent 1200 column compartment, and Agilent 1200 DAD. The instrument acquisition and data handling were done with Sciex Analyst TF version 1.7.1. Prior to acquisition instrument was calibrated with less than 5 ppm accuracy. During acquisition, a calibration run was performed initially and after every 5 injections using the Sciex positive polarity tuning mix. Elution Conditions: Column:

Waters XSelect HSS T3, 2.1x30mm, 2.5µm particle size; Column Temperature 60 °C. Solvent A: Water (0.1% formic acid), Solvent B: Acetonitrile (0.1% formic acid), Gradient: Initial 5% B, hold for 0.10 min, 5-95% B in 2.8 min, , 95-5% B in 0.20 min, 3.5 min total runtime; Flow rate 0.8 ml/min. TOF Conditions: ESI in Positive Mode; The spray chamber: Gas 1 and 2 at 60, curtain gas at 40, temperature 600 °C, IonSpray voltage 5500 V, declustering potential 100, collision energy 10. The acquisition is done in TOF MS mode with range of 100-2000 amu with accumulation time of 0.20 secs. ESI in Negative Mode; The spray chamber: Gas 1 and 2 at 60, curtain gas at 40, temperature 600 °C, IonSpray voltage -4500 V, declustering potential -100, collision energy -10. The acquisition is done in TOF MS mode with range of 100-2000 amu with accumulation time of 0.20 secs.

#### Synthesis and Characterization of Compounds **2**-**6**

Synthesis of Compound **2**

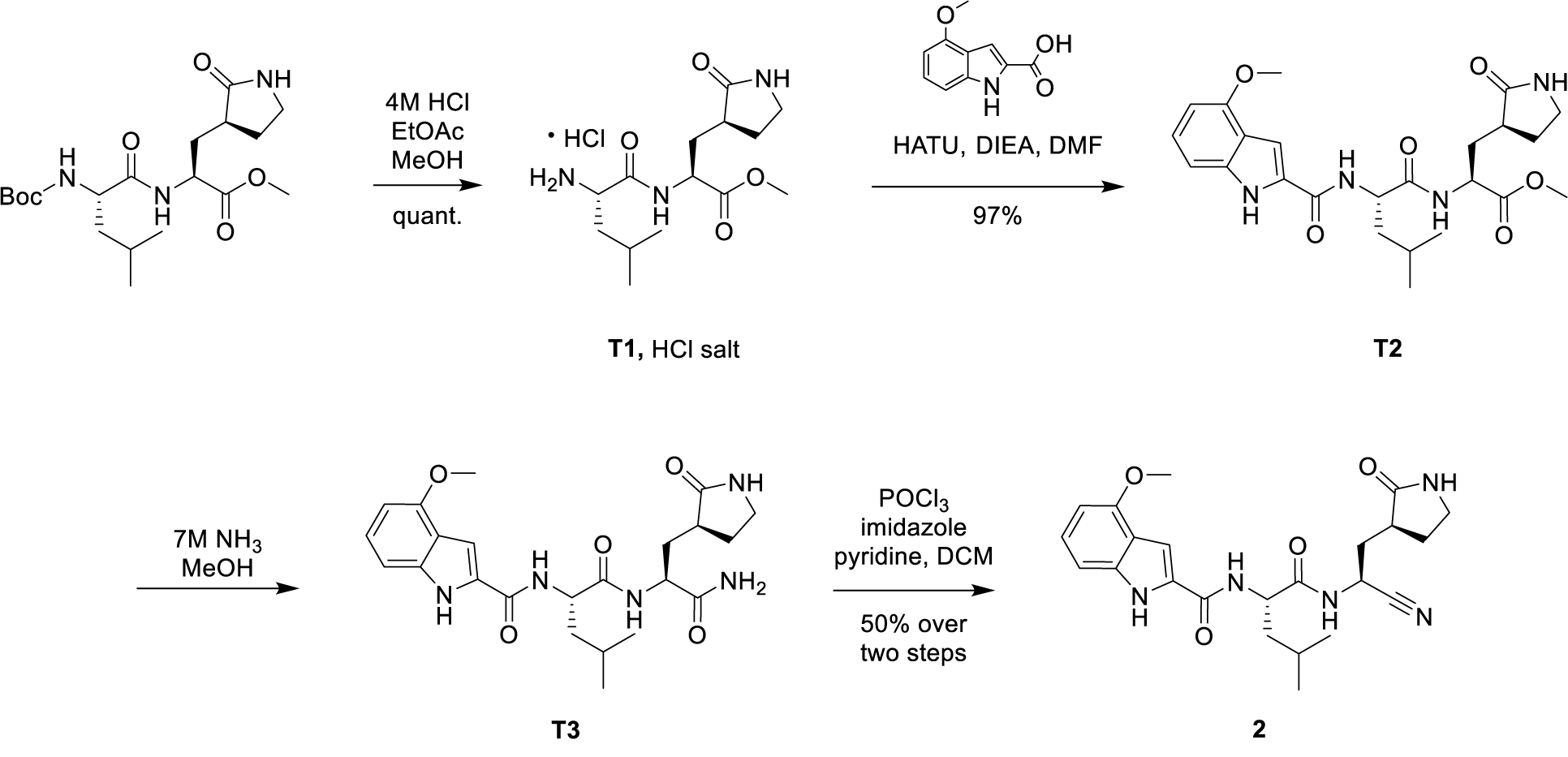

*Methyl L-leucyl-3-[(3S)-2-oxopyrrolidin-3-yl]-L-alaninate, hydrochloride salt* A solution of methyl *N*-(*tert*-butoxycarbonyl)-L-leucyl-3-[(3*S*)-2-oxopyrrolidin-3-yl]-L-alaninate (*34*) (2.0 g, 5.0 mmol) in a mixture of methanol (2 ml) and a solution of hydrogen chloride in ethyl acetate (4 M; 20 ml) was stirred at 25 °C for 1 h. Concentration afforded **T1** as a white solid (1.92 g, assumed quantitative). LCMS *m/z* 300.2 [M+H]^+^

*Methyl N-(4-methoxy-1H-indole-2-carbonyl)-L-leucyl-3-[(3S)-2-oxopyrrolidin-3-yl]-L-alaninate (**T2**)*. HATU (494 mg, 1.30 mmol) and *N,N*-diisopropylethylamine (388 mg, 3.00 mmol) were added to a 0 °C solution of **T1** (336 mg, ≤0.840 mmol) and 4-methoxy-1*H*-indole-2- carboxylic acid (159 mg, 0.832 mmol) in *N,N*-dimethylformamide (6 ml). The solution was stirred at 0 °C for 1.5 h, whereupon it was poured into water / ice (10 ml) and extracted with ethyl acetate (3 x 10 ml). The combined organic layers were washed with saturated aqueous sodium chloride solution, dried over sodium sulfate, filtered, and concentrated. Silica gel chromatography (Eluent: 10:1 dichloromethane / methanol) provided **T2** as a yellow oil. Yield: 380 mg, 0.804 mmol, 97%. ^1^H NMR (400 MHz, DMSO-*d*_6_) d 11.63 – 11.50 (m, 1H), 8.53 (d, *J* = 8.0 Hz, 1H), 8.37 (d, *J* = 8.0 Hz, 1H), 7.65 (s, 1H), 7.35 (d, *J* = 1.6 Hz, 1H), 7.09 (t, *J* = 7.9 Hz, 1H), 7.00 (d, *J* = 8.2 Hz, 1H), 6.50 (d, *J* = 7.6 Hz, 1H), 4.52 (ddd, *J* = 10.1, 8.2, 5.0 Hz, 1H), 4.36 (ddt, *J* = 11.4, 7.9, 3.3 Hz, 1H), 3.88 (s, 3H), 3.62 (s, 3H), 3.21 – 3.03 (m, 2H), 2.44 – 2.29 (m, 1H), 2.20 – 2.02 (m, 2H), 1.74 – 1.53 (m, 5H), 0.93 (d, *J* = 6.4 Hz, 3H), 0.89 (d, *J* = 6.4 Hz, 3H). LCMS *m/z* 473.2 [M+H]^+^ *N-[(4-Methoxy-1H-indol-2-yl)carbonyl]-L-leucyl-3-[(3S)-2-oxopyrrolidin-3-yl]-L-alaninamide (**T3**).* A solution of ammonia in methanol (7.0 M; 21 ml, 150 mmol) was added to a solution of **T2** (500 mg, 1.06 mmol) in methanol (2.0 ml). After the reaction mixture had been stirred at room temperature for 6 h, a solution of ammonia in methanol (7.0 M; 7.0 ml, 49 mmol) was again added, and stirring was continued overnight. A solution of ammonia in methanol (7.0 M; 7.0 ml, 49 mmol) was again added, and stirring was continued for 24 h, whereupon a final treatment with a solution of ammonia in methanol (7.0 M; 7.0 ml, 49 mmol) was carried out. The reaction mixture was stirred for one more day, at which point it was concentrated *in vacuo*. The residue was combined with the product of a similar reaction (350 mg of the 512 mg isolated) carried out using **T2** (500 mg, 1.06 mmol), and the mixture was repeatedly dissolved in ethyl acetate (5 x 10 ml) and concentrated, providing **T3** (835 mg). This material was used directly in the following step. ^1^H NMR (400 MHz, Methanol-*d*_4_) d 7.29 (d, *J* = 0.8 Hz, 1H), 7.15 (t, *J* = 8.0 Hz, 1H), 7.03 (d, *J* = 8.3 Hz, 1H), 6.51 (d, *J* = 7.7 Hz, 1H), 4.59 (dd, *J* = 9.7, 5.0 Hz, 1H), 4.45 (dd, *J* = 11.3, 4.2 Hz, 1H), 3.93 (s, 3H), 3.26 (ddd, *J* = 10.5, 7.8, 4.9 Hz, 2H), 2.52 (ddt, *J* = 14.2, 9.8, 4.6 Hz, 1H), 2.36 – 2.26 (m, 1H), 2.15 (ddd, *J* = 14.0, 11.4, 4.6 Hz, 1H), 1.84 – 1.71 (m, 5H), 1.02 (d, *J* = 6.1 Hz, 3H), 0.98 (d, *J* = 6.2 Hz, 3H). LCMS *m/z* 458.4 [M+H]^+^.

*N-[(2S)-1-({(1S)-1-Cyano-2-[(3S)-2-oxopyrrolidin-3-yl]ethyl}amino)-4-methyl-1- oxopentan-2-yl]-4-methoxy-1H-indole-2-carboxamide* (***2***). A solution of **T3** (from the previous step; 835 mg, ≤1.78 mmol) and 1*H*-imidazole (323 mg, 4.74 mmol) in a mixture of pyridine (4 ml) and dichloromethane (4 ml) was cooled to −35 °C using an acetonitrile / dry ice bath, whereupon phosphorus oxychloride (0.956 ml, 10.2 mmol) was added in a drop-wise manner over 5 min. The reaction was stirred at a temperature between −30 °C and −20 °C for about 1.5 h, then treated with hydrochloric acid (1 M; 50 ml) and stirred for 1 h. After extraction with dichloromethane (3 x 60 ml), the resulting organic layers were combined, dried over sodium sulfate, filtered, and concentrated *in vacuo*. The residue was combined with purified **2** from a different batch (75 mg, 0.17 mmol) and subjected to silica gel chromatography (Gradient: 0% to 5% methanol in ethyl acetate) to provide **2** as a solid (800 mg). This material was combined with the product (80 mg) from a similar reaction carried out using **T3** (161 mg, 0.352 mmol); the resulting material was stirred in diethyl ether (25 ml) for 3 days, whereupon it was filtered. The filter cake was washed with a mixture of diethyl ether and heptane (1:1, 4 x 2 ml) to afford *N*- [(2*S*)-1-({(1*S*)-1-cyano-2-[(3*S*)-2-oxopyrrolidin-3-yl]ethyl}amino)-4-methyl-1-oxopentan-2-yl]- 4-methoxy-1*H*-indole-2-carboxamide (**2**) as a solid. Combined yield: 519 mg, 1.18 mmol, approximately 50% over 2 steps. ^1^H NMR (600 MHz, DMSO-*d*_6_) d 11.58 (d, *J* = 2.3 Hz, 1H), 8.91 (d, *J* = 8.0 Hz, 1H), 8.47 (d, *J* = 7.7 Hz, 1H), 7.71 (s, 1H), 7.37 (dd, *J* = 2.3, 0.9 Hz, 1H), 7.09 (t, *J* = 8.0 Hz, 1H), 7.03 – 6.99 (m, 1H), 6.50 (d, *J* = 7.7 Hz, 1H), 4.98 (ddd, *J* = 9.3, 8.0, 6.7 Hz, 1H), 4.45 (ddd, *J* = 10.4, 7.7, 4.7 Hz, 1H), 3.88 (s, 3H), 3.22 – 3.05 (m, 2H), 2.42 – 2.30 (m, 1H), 2.13 (dddt, *J* = 19.4, 8.9, 6.8, 4.0 Hz, 2H), 1.86 – 1.77 (m, 1H), 1.76 – 1.65 (m, 3H), 1.53 (ddt, *J* = 12.4, 7.8, 4.4 Hz, 1H), 0.94 (d, *J* = 6.4 Hz, 3H), 0.89 (d, *J* = 6.4 Hz, 3H). ^13^C NMR (151 MHz, DMSO-*d*_6_) d 177.55, 172.53, 161.20, 153.66, 137.84, 129.75, 124.48, 119.67, 118.07, 105.41, 101.31, 99.23, 55.09, 51.40, 40.04, 39.48, 38.30, 37.12, 33.44, 27.00, 24.42, 23.02, 21.34. HRMS (ESI-TOF) *m/z* calcd. for C_23_H_29_N_5_O_4_ [M + H]^+^ 440.2293, found 440.2298.

Synthesis of Compound **3**

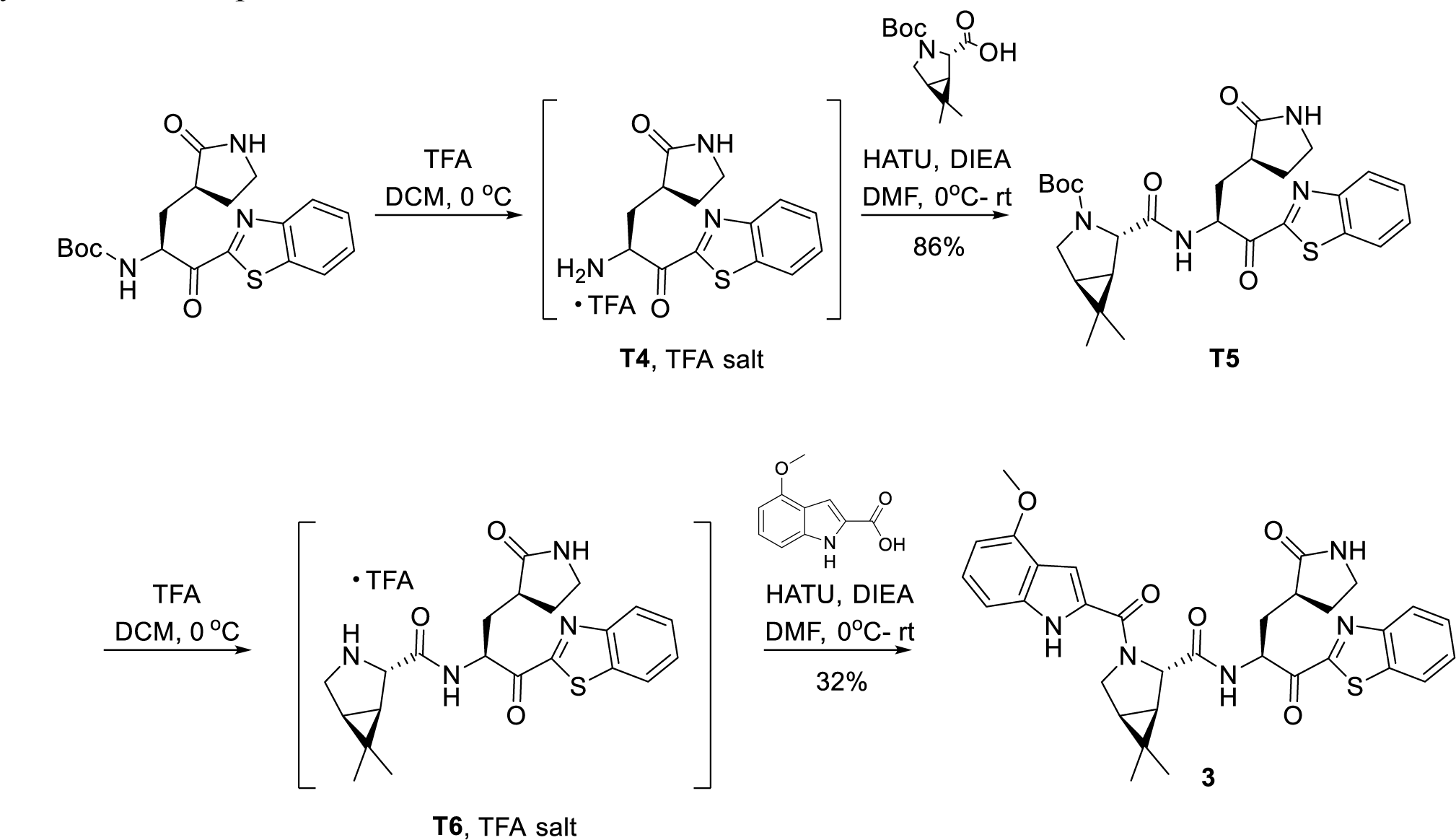

*tert-Butyl (1R,2S,5S)-2-({(2S)-1-(1,3-benzothiazol-2-yl)-1-oxo-3-[(3S)-2-oxopyrrolidin-3- yl]propan-2-yl}carbamoyl)-6,6-dimethyl-3-azabicyclo[3.1.0]hexane-3-carboxylate (**T5**)*.

Trifluoroacetic acid (0.41 ml, 5.3 mmol) was added to a 0 °C solution of tert-butyl ((S)-1- (benzo[d]thiazol-2-yl)-1-oxo-3-((S)-2-oxopyrrolidin-3-yl)propan-2-yl)carbamate (*35*) (83 mg, 0.21 mmol) in dichloromethane (1.4 ml). The reaction mixture was stirred at 0 °C for 1 h, whereupon LCMS analysis indicated that deprotection to afford **T4** was complete:

LCMS *m/z* 290.1 [M+H]^+^. Concentration provided the trifluoroacetic acid salt of the amine, which was dissolved in *N,N*-dimethylformamide (2 ml), cooled to 0 °C and treated with *N,N*- diisopropylethylamine (0.111 ml, 0.637 mmol). In a separate vial, a mixture of (1*R*,2*S*,5*S*)-3- (*tert*-butoxycarbonyl)-6,6-dimethyl-3-azabicyclo[3.1.0]hexane-2-carboxylic acid (54.4 mg, 0.213 mmol), HATU (89.1 mg, 0.234 mmol), and a drop of *N,N*-diisopropylethylamine in *N,N*- dimethylformamide (0.5 ml) was stirred at room temperature until a solution was obtained. This solution was added to the 0 °C solution of the amine salt, and the reaction mixture was allowed to warm to room temperature overnight. Ethyl acetate was added, and the resulting mixture was washed sequentially with 10% aqueous potassium bisulfate solution, 5% aqueous sodium bicarbonate solution, and saturated aqueous sodium chloride solution, dried over sodium sulfate, filtered, and concentrated. Silica gel chromatography (Gradient: 0% to 100% ethyl acetate in heptane) provided **T5** as a clear yellow oil. Yield: 93 mg, 0.18 mmol, 86%. ^1^H NMR (400 MHz, Chloroform-*d*) d 8.34 – 8.25 (m, 1H), 8.18 (dd, *J* = 8.4, 3.3 Hz, 1H), 7.98 (d, *J* = 7.3 Hz, 1H), 7.73 (d, *J* = 7.8 Hz, 0H), 7.68 – 7.48 (m, 2H), 6.24 – 5.85 (m, 1H), 5.78 (ddt, *J* = 35.8, 12.1, 4.8 Hz, 1H), 4.16 (s, 0H), 4.08 (s, 0H), 3.68 (td, *J* = 11.4, 5.3 Hz, 1H), 3.58 (d, *J* = 11.4 Hz, 1H), 3.38 (d, *J* = 6.7 Hz, 1H), 2.95 (s, 3H), 2.88 (s, 3H), 2.68 – 2.60 (m, 1H), 2.20 – 2.12 (m, 1H), 1.53 (dd, *J* = 12.3, 7.5 Hz, 1H), 1.39 (d, *J* = 4.9 Hz, 9H), 1.03 (d, *J* = 3.7 Hz, 3H), 0.92 (d, *J* = 4.9 Hz, 3H). . LCMS *m/z* 527.3 [M+H]^+^.

*(1R,2S,5S)-N-{(2S)-1-(1,3-Benzothiazol-2-yl)-1-oxo-3-[(3S)-2-oxopyrrolidin-3-yl]propan-2- yl}-3-[(4-methoxy-1H-indol-2-yl)carbonyl]-6,6-dimethyl-3-azabicyclo[3.1.0]hexane-2- carboxamide (**3**).* Trifluoroacetic acid (0.18 ml, 2.3 mmol) was added to a 0 °C solution of **T5** (50 mg, 95 µmol) in dichloromethane (1.4 ml). After the reaction mixture had been stirred at 0 °C for 1.5 h, LCMS analysis indicated the presence of the deprotected material **T6**: LCMS *m/z* 427.3 [M+H]^+^. The reaction mixture was concentrated, cooled to 0 °C, dissolved in *N,N*-dimethylformamide (2 ml) and treated with *N,N*-diisopropylethylamine (49.6 µL, 0.285 mmol). To this 0 °C mixture was added a solution of 4-methoxy-1*H*-indole-2-carboxylic acid (18.2 mg, 95.2 µmol), HATU (39.7 mg, 0.104 mmol), and a drop of *N,N*-diisopropylethylamine in *N,N*-dimethylformamide (0.5 ml). After 2 h, the reaction mixture was diluted with ethyl acetate, washed sequentially with 10% aqueous potassium bisulfate solution, 5% aqueous sodium bicarbonate solution, and saturated aqueous sodium chloride solution, dried over sodium sulfate, filtered, and concentrated under reduced pressure. Purification via reversed-phase HPLC (Column: Waters Sunfire C18, 19 x 100 mm, 5 µm; Mobile phase A: water containing 0.05%trifluoroacetic acid; Mobile phase B: acetonitrile containing 0.05% trifluoroacetic acid; Gradient: 30% to 70% B over 8.5 minutes, then 70% to 95% B over 0.5 min, then 95% B for 1.0 min; Flow rate: 25 ml/minute) afforded **3**. Yield: 18.4 mg, 30.7 µmol, 32%. ^1^H NMR (600 MHz, DMSO-*d*6 at 27 °C, presents as ∼1:1 rotamers) d 11.54 (d, *J* = 2.3 Hz, 1H), 11.50 (d, *J* = 2.3 Hz, 1H), 9.24 (d, *J* = 7.7 Hz, 1H), 9.01 (d, *J* = 7.1 Hz, 1H), 8.32 – 8.26 (m, 2H), 8.25 – 8.19 (m, 2H), 7.70 (s, 1H), 7.67 – 7.62 (m, 4H), 7.60 (s, 1H), 7.12 (td, *J* = 8.0, 5.0 Hz, 2H), 7.02 (dd, *J* = 8.3, 5.6 Hz, 2H), 6.98 – 6.94 (m, 1H), 6.90 (d, *J* = 2.3 Hz, 1H), 6.51 (d, *J* = 7.7 Hz, 1H), 6.48 (d, *J* = 7.7 Hz, 1H), 5.54 (ddd, *J* = 11.0, 7.2, 3.6 Hz, 1H), 5.41 (ddd, *J* = 12.1, 7.7, 2.9 Hz, 1H), 4.95 (s, 1H), 4.59 (s, 1H), 4.15 (dd, *J* = 10.4, 5.4 Hz, 1H), 4.03 (q, *J* = 7.1 Hz, 0H), 3.88 (s, 3H), 3.81 (s, 3H), 3.77 – 3.68 (m, 2H), 3.25 – 3.12 (m, 2H), 3.04 – 2.93 (m, 1H), 2.69 – 2.55 (m, 3H), 2.34 – 2.22 (m, 1H), 2.11 (dddd, *J* = 29.3, 13.6, 11.6, 4.0 Hz, 2H), 1.94 – 1.77 (m, 3H), 1.62 – 1.57 (m, 3H), 1.43 (dd, *J* = 7.6, 4.9 Hz, 1H), 1.35 (d, *J* = 7.6 Hz, 1H), 1.07 (s, 3H), 1.01 (s, 4H), 0.97 (s, 3H), 0.88 (s, 4H). ^1^H NMR (600 MHz, DMSO-*d*6 at 140 °C) d 10.99 (s, 1H), 8.63 (s, 1H), 8.18 (td, *J* = 7.0, 2.1 Hz, 2H), 7.75 – 7.54 (m, 2H), 7.21 – 7.02 (m, 3H), 6.92 (s, 1H), 6.52 (d, *J* = 7.5 Hz, 1H), 5.51 (s, 1H), 4.75 (s, 1H), 3.92 (s, 3H), 3.87 (d, *J* = 10.9 Hz, 1H), 3.16 (s, 2H), 2.89 (s, 2H), 2.60 – 2.51 (m, 1H), 2.22 (ddd, *J* = 14.6, 10.2, 4.7 Hz, 1H), 1.95 (q, *J* = 6.7, 5.3 Hz, 1H), 1.81 (s, 1H), 1.48 (s, 2H), 1.04 (s, 3H), 0.95 (s, 3H). ^13^C NMR (151 MHz, DMSO-*d*6 at 27 °C) d 193.33, 193.05, 178.14, 177.90, 171.38, 171.28, 164.45, 163.94, 160.49, 159.54, 153.74, 153.68, 152.90, 152.87, 137.23, 137.07, 136.38, 129.08, 129.01, 128.31, 128.19, 127.57, 127.52, 125.32, 125.21, 124.91, 124.71, 123.24, 123.22, 118.31, 118.11, 105.30, 105.22, 102.62, 100.60, 99.16, 98.94, 64.92, 61.10, 60.38, 59.76, 54.98, 54.78, 53.45, 53.33, 48.47, 48.23, 39.94, 37.92, 37.41, 33.55, 32.29, 32.13, 29.85, 27.69, 27.39, 26.30, 26.07, 25.86, 24.23, 18.81, 18.65, 12.57, 12.53. HRMS (ESI-TOF) *m/z* calcd. for C_32_H_33_N_5_O_5_S [M + H]^+^ 600.2275, found 600.2275.

Synthesis of Compound **4**

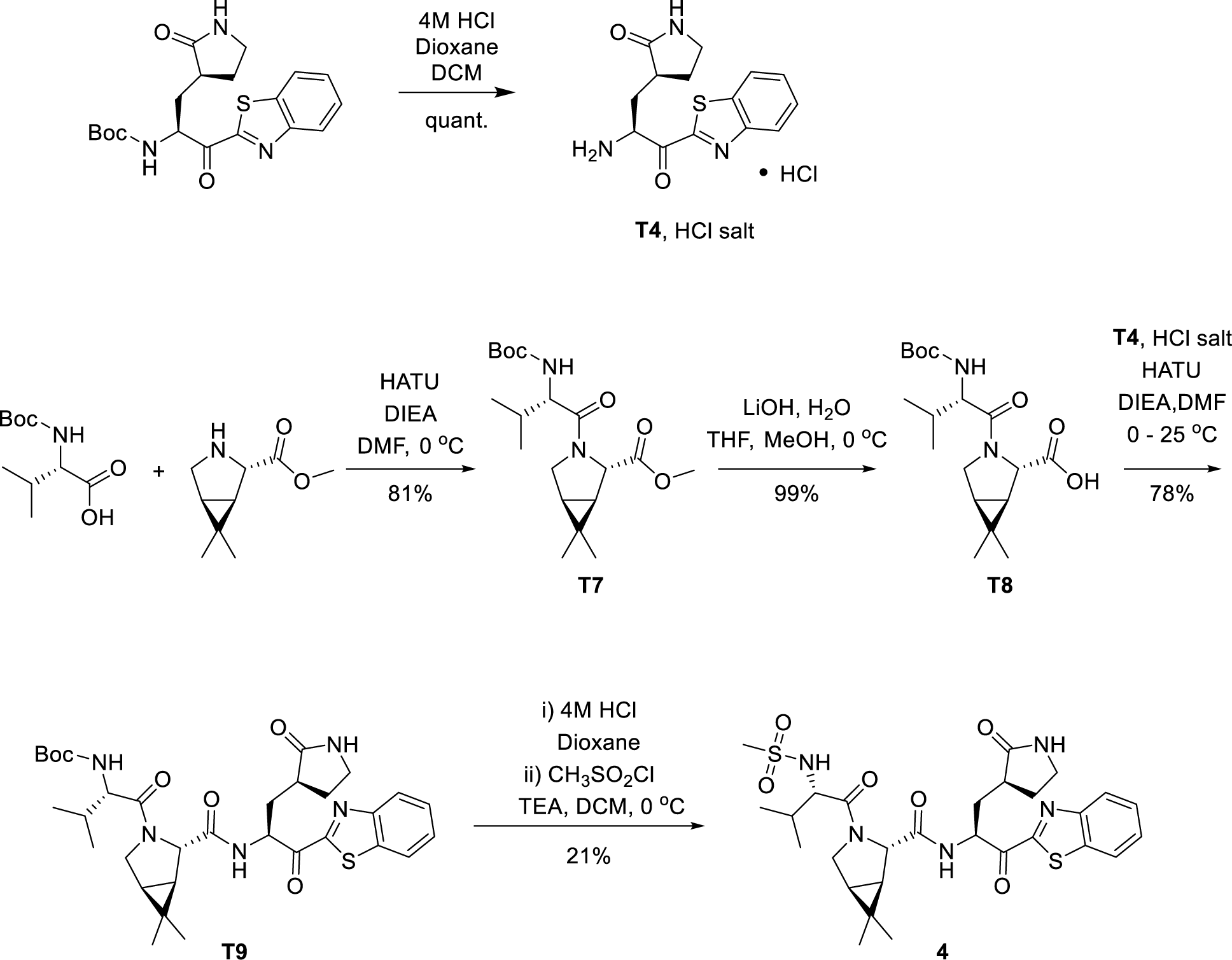

*(3S)-3-[(2S)-2-Amino-3-(1,3-benzothiazol-2-yl)-3-oxopropyl]pyrrolidin-2-one (**T4**), HCl salt.* A solution of tert-butyl ((S)-1-(benzo[d]thiazol-2-yl)-1-oxo-3-((S)-2-oxopyrrolidin-3- yl)propan-2-yl)carbamate (230 mg, 0.591 mml) in dichloromethane (4 ml) was treated with a solution of HCl in 1,4-dioxane (4 M; 1.48 ml, 5.92 mmol), followed by ethyl acetate (0.3 ml). The reaction mixture was stirred for 1 h at room temperature, whereupon it was concentrated *in vacuo*. Trituration of the residue with diethyl ether afforded the HCl salt of **T4** as a bright yellow solid, which was used in the next step without further purification. LCMS m/z 290.2 [M+H]^+^.

*Methyl (1R,2S,5S)-3-[N-(tert-butoxycarbonyl)-L-valyl]-6,6-dimethyl-3- azabicyclo[3.1.0]hexane-2-caryaboxylate (**T7**)*. A 0 °C mixture of *N*-(*tert*-butoxycarbonyl)-L-valine (4.23 g, 19.5 mmol), methyl (1*R*,2*S*,5*S*)-6,6-dimethyl-3-azabicyclo[3.1.0]hexane-2- carboxylate, hydrochloride salt (4.00 g, 19.4 mmol) and *N,N*-dimethylformamide (97 ml) was treated with HATU (8.13 g, 21.4 mmol). After the reaction mixture had been stirred for 5 minutes, *N,N*-diisopropylethylamine (8.47 ml, 48.6 mmol) was added and stirring was continued at 0 °C for 2 h. The reaction mixture was then diluted with aqueous citric acid solution (1 N; 20 ml) and water (40 ml), stirred for 2 minutes, and diluted with ethyl acetate (250 ml). The organic layer was washed with water (3 x 150 ml), and the combined aqueous layers were extracted with ethyl acetate (100 ml). The combined organic layers were then washed with saturated aqueous sodium chloride solution, dried over sodium sulfate, filtered and concentrated *in vacuo*. Silica gel chromatography (Gradient: 0% to 100% ethyl acetate in heptane) provided **T7** as a gum. Yield: 5.80 g, 15.7 mmol, 81%.^1^H NMR (400 MHz, Chloroform-*d*) d 5.06 (d, *J* = 9.6 Hz, 1H), 4.45 (s, 1H), 4.14 – 4.08 (m, 1H), 3.95 (d, *J* = 10.2 Hz, 1H), 3.86 (dd, *J* = 10.1, 4.8 Hz, 1H), 3.74 (s, 3H), 1.99 (dq, *J* = 13.6, 6.7 Hz, 1H), 1.48 – 1.44 (m, 2H), 1.40 (s, 9H), 1.05 (s, 3H), 1.00 (d, *J* = 6.7 Hz, 3H), 0.95 (d, *J* = 6.8 Hz, 3H), 0.93 (s, 3H). LCMS *m/z* 369.3 [M+H]^+^.

*(1R,2S,5S)-3-[N-(tert-Butoxycarbonyl)-L-valyl]-6,6-dimethyl-3-azabicyclo[3.1.0]hexane-2- carboxylic acid (**T8**)*. Aqueous lithium hydroxide solution (1 M; 8.14 ml, 8.14 mmol) was added dropwise to a 0 °C solution of **T7** (2.00 g, 5.43 mmol) in a mixture of tetrahydrofuran and methanol (1:1, 30 ml). The reaction mixture was stirred at 0 °C for 2 h, and then at room temperature for 4 h, whereupon aqueous lithium hydroxide solution (1 M; 1.67 ml, 1.67 mmol) was added and stirring was continued for 15 min. Aqueous lithium hydroxide solution (1 M; 3 ml, 3 mmol) was added once more; after a further 15 min, the reaction pH was adjusted to 3 by addition of 1 M HCl. The resulting mixture was diluted with water (30 ml), and the aqueous layer was extracted with ethyl acetate (2 x 75 ml). The combined organic layers were dried over sodium sulfate, filtered, and concentrated *in vacuo* to afford **T8** as an off-white solid. Yield: 1.90 g, 5.36 mmol, 99%. ^1^H NMR (400 MHz, DMSO-*d*_6_) d 12.59 (s, 1H), 7.03 (d, *J* = 8.6 Hz, 1H), 4.10 (s, 1H), 3.98 (d, *J* = 10.4 Hz, 1H), 3.80 (t, *J* = 9.1 Hz, 1H), 3.74 (dd, *J* = 10.2, 5.3 Hz, 1H), 1.95 – 1.79 (m, 1H), 1.55 – 1.46 (m, 1H), 1.39 (d, *J* = 4.7 Hz, 1H), 1.34 (s, 9H), 1.01 (s, 3H), 0.89 – 0.84 (m, 9H). LCMS *m/z* 355.3 [M+H]^+^.

*tert-Butyl {(2S)-1-[(1R,2S,5S)-2-({(2S)-1-(1,3-benzothiazol-2-yl)-1-oxo-3-[(3S)-2- oxopyrrolidin-3-yl]propan-2-yl}carbamoyl)-6,6-dimethyl-3-azabicyclo[3.1.0] hexan-3-yl]-3- methyl-1-oxobutan-2-yl}carbamate (**T9**).* To a 0 °C solution of **T8** (80.8 mg, 0.228 mmol), HATU (86.7 mg, 0.228 mmol), and *N,N*-diisopropylethylamine (54 µL, 0.31 mmol) in *N,N*-dimethylformamide (2 ml) was added **T4**, hydrochloride salt (60 mg, 0.18 mmol). DIEA (54 µL, 0.31 mmol) was added, and the reaction mixture was allowed to stir and warm to room temperature overnight. It was then diluted with EtOAc, washed sequentially with 10% aqueous potassium bisulfate solution, 5% aqueous sodium bicarbonate solution, and saturated aqueous sodium chloride solution, dried over sodium sulfate, filtered, and concentrated. Chromatography on silica gel (Gradient: 0% to 100% EtOAc in heptane) afforded **T9** as an oil. Yield: 91 mg, 0.14 mmol, 78%. ^1^H NMR (400 MHz, Chloroform-*d*) d 8.17 (d, *J* = 7.6 Hz, 1H), 7.98 (d, *J* = 8.1 Hz, 1H), 7.78 (d, *J* = 7.0 Hz, 1H), 7.62 – 7.50 (m, 2H), 5.83 (dt, *J* = 14.5, 6.1 Hz, 2H), 3.91 (dd, *J* = 10.7, 4.5 Hz, 2H), 3.41 – 3.30 (m, 2H), 2.79 – 2.65 (m, 1H), 2.58 (d, *J* = 6.6 Hz, 1H), 2.23 – 2.06 (m, 3H), 1.98 (dd, *J* = 13.5, 6.7 Hz, 1H), 1.40 (s, 9H), 1.04 (d, *J* = 5.3 Hz, 3H), 0.98 – 0.90 (m, 9H). Exchangeable protons not observed. LCMS *m/z* 626.4 [M+H]^+^.

*(1R,2S,5S)-N-{(2S)-1-(1,3-Benzothiazol-2-yl)-1-oxo-3-[(3S)-2-oxopyrrolidin-3-yl]propan-2- yl}-6,6-dimethyl-3-[N-(methylsulfonyl)-L-valyl]-3-azabicyclo[3.1.0]hexane-2 carboxamide (**4**)*. A solution of HCl in 1,4-dioxane (4 M; 0.364 ml, 1.46 mmol) was added to a solution of **T9** (91 mg, 0.14 mmol) in DCM(1 ml), and the reaction mixture was stirred at room temperature for 2 h, whereupon a solution of hydrogen chloride in 1,4-dioxane (4 M, 0.1 ml, 0.4 mmol) was again added. Stirring was continued for an additional 2 h, and then the reaction mixture was concentrated, dissolved in methylene chloride (1 ml), and cooled to 0 °C. To this was added triethylamine (60.5 µl, 0.434 mmol), followed by methanesulfonyl chloride (12.4 µl, 0.160 mmol), and the reaction mixture was stirred at 0 °C for 2.5 h. It was then partitioned between 10% aqueous potassium bisulfate solution and ethyl acetate; the organic layer was washed with saturated aqueous sodium chloride solution, dried over sodium sulfate, filtered, and concentrated. Silica gel chromatography (Gradient: 0% to 100% ethyl acetate in heptane) afforded **4** as a white solid. Yield: 18 mg, 30 µmol, 21%. ^1^H NMR (600 MHz, DMSO-*d*_6_ at 27 °C, presents as ∼3:1 rotamers) d 9.30 (d, *J* = 6.7 Hz, 1H), 8.89 (d, *J* = 7.9 Hz, 3H), 8.28 (ddd, *J* = 8.2, 5.1, 1.8 Hz, 5H), 8.25 – 8.20 (m, 3H), 7.73 – 7.60 (m, 13H), 7.35 (d, *J* = 9.0 Hz, 3H), 6.96 (d, *J* = 9.9 Hz, 1H), 5.60 (ddd, *J* = 11.5, 7.9, 3.3 Hz, 3H), 5.51 (ddd, *J* = 11.4, 6.7, 3.0 Hz, 1H), 4.42 (s, 1H), 4.32 (s, 3H), 3.78 (dd, *J* = 10.3, 5.4 Hz, 3H), 3.74 (dd, *J* = 9.0, 7.8 Hz, 4H), 3.69 (d, *J* = 10.4 Hz, 3H), 3.61 – 3.55 (m, 2H), 3.24 – 3.18 (m, 4H), 3.15 (td, *J* = 9.4, 7.0 Hz, 0H), 3.10 (td, *J* = 9.4, 7.0 Hz, 3H), 2.80 (s, 10H), 2.70 (s, 3H), 2.59 (tdd, *J* = 11.5, 8.6, 3.6 Hz, 3H), 2.33 – 2.24 (m, 3H), 2.20 – 2.03 (m, 4H), 1.94 (ddd, *J* = 13.8, 10.6, 3.1 Hz, 1H), 1.91 – 1.76 (m, 11H), 1.50 (dd, *J* = 7.7, 5.4 Hz, 3H), 1.42 (dd, *J* = 7.6, 5.2 Hz, 1H), 1.30 (d, *J* = 7.7 Hz, 3H), 1.03 (s, 3H), 0.97 (s, 9H), 0.96 (s, 3H), 0.94 (s, 8H), 0.93 (d, *J* = 3.6 Hz, 16H), 0.92 (s, 2H), 0.90 (d, *J* = 6.7 Hz, 11H), 0.72 (d, *J* = 6.8 Hz, 3H). ^13^C NMR (151 MHz, DMSO-*d*_6_ at 27 °C) d 193.03, 192.86, 178.23, 177.97, 171.45, 171.06, 169.78, 169.21, 164.38, 164.01, 163.06, 152.92, 136.40, 128.34, 128.24, 127.63, 127.57, 125.33, 125.20, 123.26, 60.00, 59.35, 59.27, 59.10, 54.18, 52.95, 47.35, 46.98, 40.83, 40.06, 39.99, 39.44, 38.07, 37.63, 33.01, 32.52, 32.27, 30.61, 30.46, 29.44, 27.55, 27.29, 27.20, 25.98, 25.97, 24.89, 20.04, 19.00, 18.76, 18.71, 18.35, 12.75, 12.72. HRMS (ESI-TOF) *m/z* calcd. for C_28_H_37_N_5_O_6_S_2_ [M + H]^+^ 604.2258, found 604.2232.

Synthesis of compound **5**

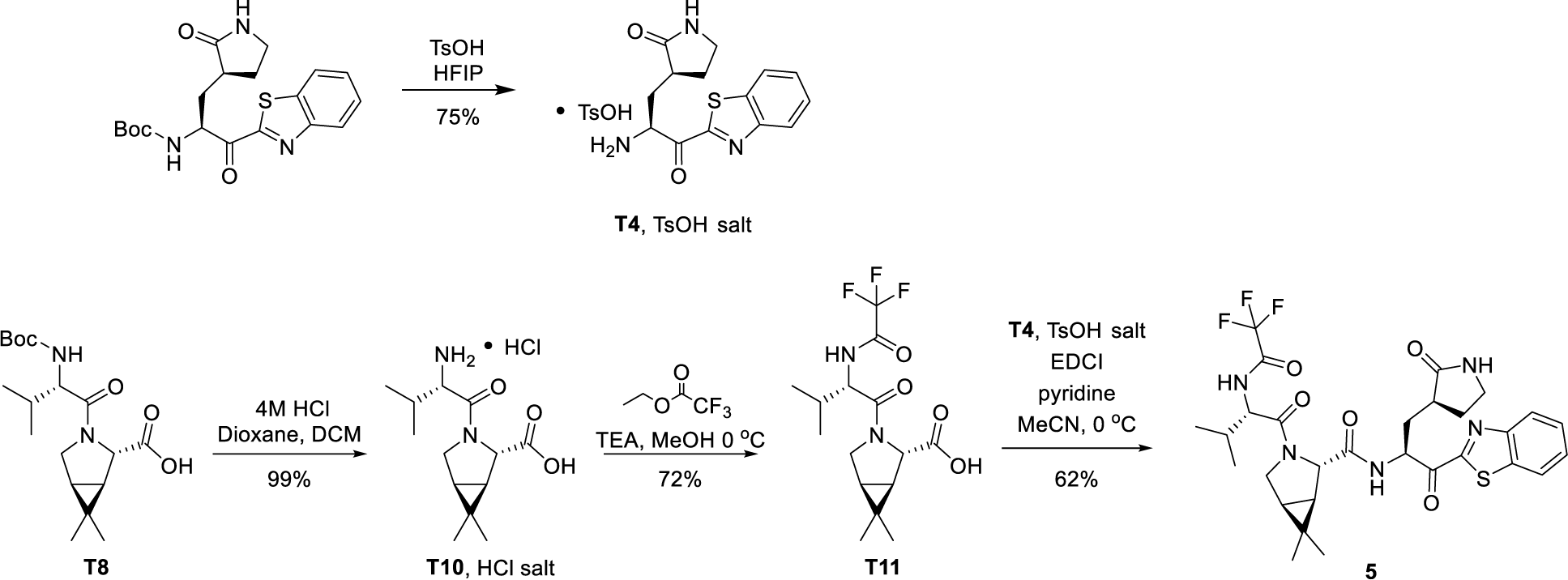

*3S)-3-[(2S)-2-Amino-3-(1,3-benzothiazol-2-yl)-3-oxopropyl]pyrrolidin-2-one (**T4**), para- toluenesulfonic salt.* A solution of *tert*-butyl ((S)-1-(benzo[d]thiazol-2-yl)-1-oxo-3-((S)-2- oxopyrrolidin-3-yl)propan-2-yl)carbamate (15.4 g, 39.5 mmol) in hexafluoroisopropanol (155 ml) was cooled in an ice bath and stirred for 15 min. The solution was then treated with *para*- toluenesulfonic acid (7.90 g, 41.5 mmol) and stirred for another 5 min before removing the ice bath. After the reaction mixture had been stirred at room temperature for 45 min, it was concentrated; trituration of the residue with ethyl acetate (50 ml) provided **T4** *para*-toluenesulfonate salt ^1^H NMR (400 MHz, Methanol-*d*4) d 8.31 – 8.23 (m, 1H), 8.23 – 8.14 (m, 1H), 7.77 – 7.62 (m, 4H), 7.22 (d, *J* = 7.9 Hz, 2H), 5.34 (dd, *J* = 9.9, 2.0 Hz, 1H), 3.55 – 3.32 (m, 2H), 3.03 (dtd, *J* = 10.8, 9.0, 4.9 Hz, 1H), 2.55 (ddd, *J* = 15.2, 4.8, 2.3 Hz, 1H), 2.46 (dddd, *J* = 12.6, 8.6, 6.3, 2.3 Hz, 1H), 2.36 (s, 3H), 2.15 – 1.88 (m, 2H). LCMS *m/z* 290.1 [M+H]^+^.

*(1R,2S,5S)-3-(L-Valyl)-6,6-dimethyl-3-azabicyclo[3.1.0]hexane-2-carboxylic acid (**T10**), HCl salt.* A solution of HCl in 1,4-dioxane (4 M; 423 ml, 1690 mmol) was added to a solution of **T8** (120 g, 339 mmol) in DCM(169 ml), and the reaction mixture was stirred at room temperature for 18 h. Removal of solvents and trituration with diethyl ether afforded the HCl salt of **T10** as a white solid (98 g, 99%). This material was used directly in the following step. ^1^H NMR (400 MHz, Methanol-*d*_4_) d 4.42 (s, 1H), 4.05 (d, *J* = 4.8 Hz, 1H), 3.89 (dd, *J* = 10.5, 5.2 Hz, 1H), 3.74 (d, *J* = 10.5 Hz, 1H), 2.38 – 2.26 (m, 1H), 1.66 – 1.53 (m, 2H), 1.16 (d, *J* = 7.0 Hz, 3H), 1.10 (s, 3H), 1.04 (d, *J* = 6.9 Hz, 3H), 1.01 (s, 3H). LCMS *m/z* 255.2 [M+H]^+^.

*(1R,2S,5S)-6,6-Dimethyl-3-[N-(trifluoroacetyl)-L-valyl]-3-azabicyclo[3.1.0]hexane-2- carboxylic acid (**T11**).* To a 0 °C solution of the HCl salt of **T10**, (40 g, 140 mmol) in methanol (138 ml) was added triethylamine (39.3 ml, 282 mmol), whereupon the reaction mixture was allowed to warm to room temperature. Ethyl trifluoroacetate (25.4 g, 21.3 ml, 179 mmol) was then added and the reaction mixture was stirred for 1.5 h. It was then concentrated *in vacuo* and the residue was diluted with water (250 ml) and adjusted to a pH of 3 to 4 by addition of 1 N HCl (250 ml) followed by conc. HCl. After extraction of the aqueous layer with ethyl acetate (3 x 250 ml), the combined organic layers were washed with saturated aqueous NaCl solution (300 ml), dried over sodium sulfate, filtered, and concentrated to afford **T11** as a white solid (34.5 g, 72 %). ^1^H NMR (400 MHz, Methanol-*d*_4_) d 9.42 (d, *J* = 7.6 Hz, 1H), 4.35 – 4.26 (m, 2H), 4.03 (d, *J* = 10.4 Hz, 1H), 3.93 (dd, *J* = 10.4, 5.3 Hz, 1H), 2.17 (dt, *J* = 9.6, 6.7 Hz, 1H), 1.59 (dd, *J* = 7.4, 5.2 Hz, 1H), 1.51 (d, *J* = 7.6 Hz, 1H), 1.08 (s, 3H), 1.06 (d, *J* = 6.7 Hz, 3H), 0.99 (d, *J* = 6.6 Hz, 3H), 0.94 (s, 3H). One exchangeable observed. LCMS *m/z* 351.2 [M+H]^+^.

*(1R,2S,5S)-N-{(2S)-1-(1,3-Benzothiazol-2-yl)-1-oxo-3-[(3S)-2-oxopyrrolidin-3-yl]propan-2- yl}-6,6-dimethyl-3-[N-(trifluoroacetyl)-L-valyl]-3-azabicyclo[3.1.0]hexane-2-carboxamide (**5**).* To a solution of **T11** (20 g, 57 mmol) and **T4**, *para-*toluenesulfonate salt (26.3 g, 57.1 mmol) in anhydrous acetonitrile (228 ml) at 0 °C was added EDCI (10.9 g, 57.1 mmol). To the stirring reaction mixture was added pyridine (13.9 ml, 171 mmol) dropwise. The resulting reaction mixture was stirred at 0 °C for 2.5 h, whereupon it was diluted with ethyl acetate (500 ml) and washed with 1N hydrochloric acid (200 ml), 5% aqueous copper sulfate (200 ml), saturated sodium bicarbonate (200 ml), and saturated sodium chloride (175 ml). The separated organic layer was then dried over sodium sulfate, filtered, and concentrated to afford an off white solid. The solid was then stirred in tetrahydrofuran (80 ml) and heptanes (120 ml) for 16 h. The resulting solid was filtered and further purified using a silica gel plug eluting with 40–60% ethyl acetate/dichloromethane. The filtrate was concentrated, whereupon the resulting solid was stirred in tetrahydrofuran (60 ml) and heptanes (120 ml) for 16 h. Filtration afforded **5** as a white solid (25 g, 71 %, 88.2 % qNMR potency, 9.5 (wt/wt)% THF). ^1^H NMR (600 MHz, DMSO-*d*_6_) d 9.82 (d, *J* = 7.7 Hz, 1H), 8.90 (d, *J* = 7.9 Hz, 1H), 8.31 – 8.25 (m, 1H), 8.25 – 8.20 (m, 1H), 7.74 – 7.58 (m, 3H), 5.60 (ddd, *J* = 11.5, 7.8, 3.4 Hz, 1H), 4.29 (s, 1H), 4.12 (dd, *J* = 10.0, 7.7 Hz, 1H), 3.94 – 3.76 (m, 2H), 3.25 – 3.18 (m, 1H), 3.10 (td, *J* = 9.4, 7.0 Hz, 1H), 2.66 – 2.55 (m, 1H), 2.34 – 2.22 (m, 1H), 2.20 – 2.03 (m, 2H), 1.81 (ddt, *J* = 14.2, 8.4, 3.5 Hz, 2H), 1.51 (dd, *J* = 7.8, 5.2 Hz, 1H), 1.30 (d, *J* = 7.7 Hz, 1H), 0.98 (s, 3H), 0.94 (d, *J* = 6.8 Hz, 3H), 0.89 (d, *J* = 6.6 Hz, 3H), 0.86 (s, 3H). ^13^C NMR (151 MHz, DMSO-*d*_6_) d 193.03, 178.21, 170.92, 168.35, 164.38, 156.49 (q, *J* = 36.8 Hz), 152.92, 136.41, 128.26, 127.59, 125.21, 123.28, 115.86 (q, *J* = 288.4 Hz), 59.98, 57.38, 52.96, 47.22, 40.06, 37.61, 32.49, 30.62, 29.46, 27.58, 27.08, 25.93, 18.89, 18.78, 18.35, 12.45. ^19^F NMR (376 MHz, DMSO-*d*_6_) d -73.55. HRMS (ESI-TOF) *m/z* calcd. for C_29_H_34_F_3_N_5_O_5_S [M + H]^+^ 622.2306, found 622.2300.

Synthesis of PF-07321332 (Compound **6**): Anhydrous, MTBE solvate form

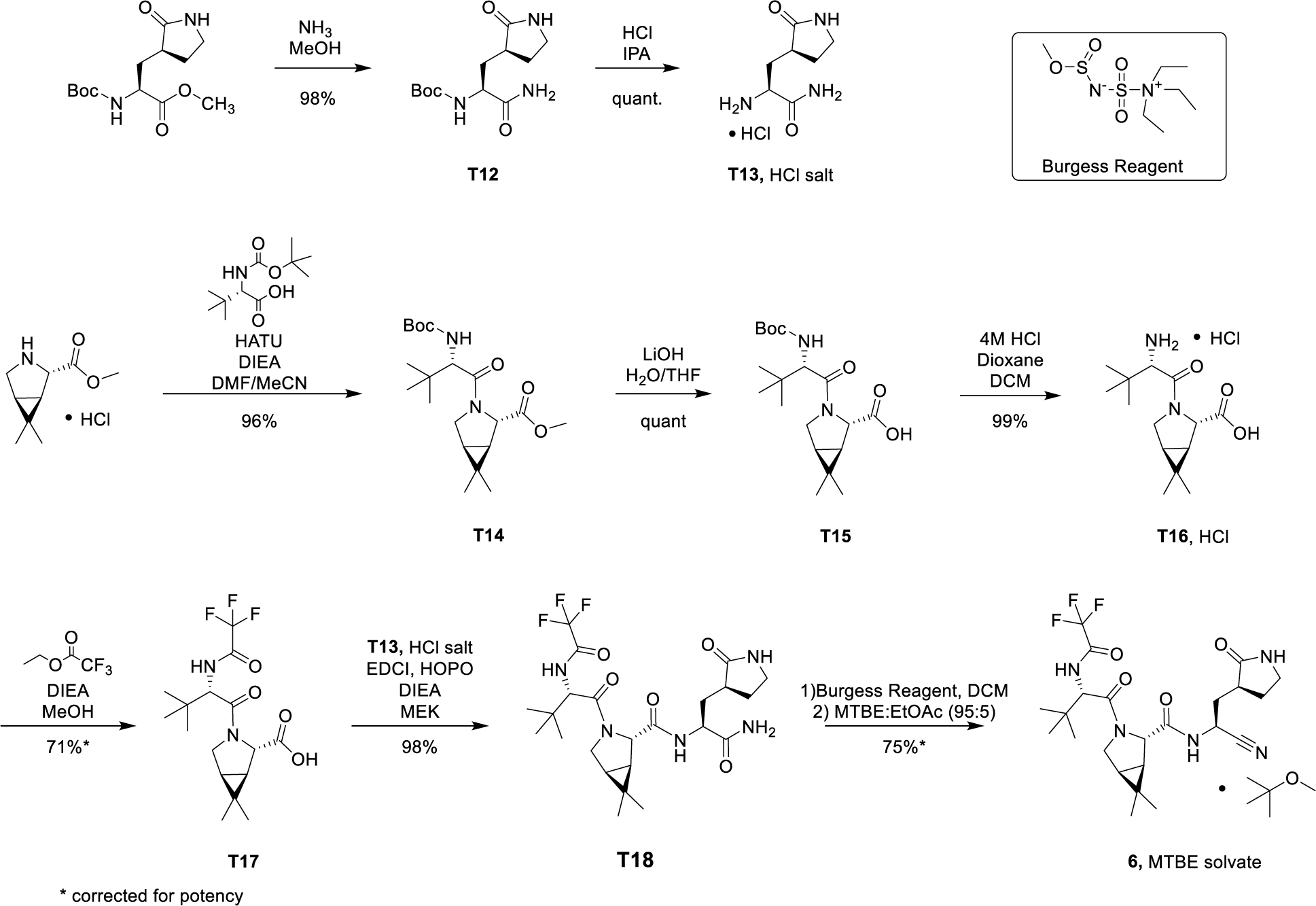

*tert-Butyl {(2S)-1-amino-1-oxo-3-[(3S)-2-oxopyrrolidin-3-yl]propan-2 yl}carbamate (**T12**)*. A solution of ammonia in methanol (7 M; 0.53 L, 3.68 mol) was added to methyl *N*-(*tert*-butoxycarbonyl)-3-[(3*S*)-2-oxopyrrolidin-3-yl]-L-alaninate (62 g, 0.22 mol) and the reaction mixture was stirred at 25 °C for 48 h. Concentration provided **T12** as a yellow solid (57.5 g, 98%). ^1^H NMR (400 MHz, Methanol-*d*_4_) d 4.10 (dd, *J* = 11.0, 3.8 Hz, 1H), 3.40 – 3.31 (m, 2H), 2.48 (dh, *J* = 9.8, 4.5 Hz, 1H), 2.41 – 2.28 (m, 1H), 2.04 (ddd, *J* = 14.1, 11.1, 4.4 Hz, 1H), 1.87 (dq, *J* = 12.5, 8.8 Hz, 1H), 1.74 (ddd, *J* = 14.0, 10.3, 4.1 Hz, 1H), 1.45 (s, 9H). LCMS *m/z* 272. 3 [M+H]^+^.

*3-[(3S)-2-Oxopyrrolidin-3-yl]-L-alaninamide (**T13**), HCl salt*. To a 0 °C solution of **T12** (57.5 g, 0.18 mol) in isopropanol (500 ml) was added a solution of HCl in isopropanol (5.5 M; 163 ml, 895 mmol). The reaction mixture was stirred at 50 °C for 4 h, cooled down to room temperature, stirred at room temperature overnight, whereupon it was concentrated to afford the HCl salt of **T13** as a white solid (37 g, 100%). ^1^H NMR (400 MHz, Methanol-*d*_4_) d 4.04 (dd, *J* = 9.2, 4.4 Hz, 1H), 3.39 (dd, *J* = 9.0, 4.5 Hz, 2H), 2.90 – 2.68 (m, 1H), 2.43 (ddt, *J* = 12.8, 8.9, 4.6 Hz, 1H), 2.21 – 1.96 (m, 2H), 1.88 (dq, *J* = 12.6, 9.1 Hz, 1H). LCMS *m/z* 172. 2 [M+H]^+^.

*Methyl (1R,2S,5S)-3-[N-(tert-butoxycarbonyl)-3-methyl-L-valyl]-6,6-dimethyl-3- azabicyclo[3.1.0]hexane-2-carboxylate (**T14).*** To a 0 °C solution of methyl (1*R*,2*S*,5*S*)-6,6- dimethyl-3-azabicyclo[3.1.0]hexane-2-carboxylate, HCl salt (50 g, 243 mmol) and *N*-(*tert*- butoxycarbonyl)-3-methyl-L-valine (61.8 g, 267 mmol) in a mixture of *N,N*-dimethylformamide (97 ml) and acetonitrile (875 ml) was added *O*HATU (102 g, 267 mmol), followed by drop-wise addition of *N,N*-diisopropylethylamine (127 ml, 729 mmol). The reaction mixture was then allowed to warm to 25 °C and was stirred for 16 h, whereupon it was concentrated. The resulting residue was treated with ethyl acetate (200 ml), washed by water (200 ml). The separated aqueous phase was extracted by ethyl acetate (2 x 200 ml). The combined organic phases were washed by water (200 ml) and HCl (1 M; 100 ml), saturated aqueous sodium chloride solution (100 ml), dried over sodium sulfate, filtered, and concentrated. The residue was purified using silica gel chromatography (Gradient: 30% to 50% ethyl acetate in heptane) to afford **T14** as a colorless oil (89.5 g, 96%). ^1^H NMR (400 MHz, DMSO-*d*_6_) d 6.73 (d, *J* = 9.3 Hz, 1H), 4.21 (s, 1H), 4.05 (d, *J* = 9.4 Hz, 1H), 3.93 (d, *J* = 10.4 Hz, 1H), 3.79 (dd, *J* = 10.3, 5.3 Hz, 1H), 3.65 (s, 3H), 1.55 – 1.49 (m, 1H), 1.41 (d, *J* = 7.5 Hz, 1H), 1.35 (s, 9H), 1.01 (s, 3H), 0.93 (s, 9H), 0.85 (s, 3H). LCMS *m/z* 383.3 [M+H]^+^.

*(1R,2S,5S)-3-[N-(tert-Butoxycarbonyl)-3-methyl-L-valyl]-6,6-dimethyl-3- azabicyclo[3.1.0]hexane-2-carboxylic acid (**T15**)*. To a solution of **T14** (88.2 g, 231 mmol) in tetrahydrofuran (231 ml) was added lithium hydroxide (16.6 g, 692 mmol) and water (50 ml). After the reaction mixture had been stirred at 25 °C for 2 h, it was concentrated to remove most of the tetrahydrofuran; the residue was then adjusted to pH 2 by addition of 1 M HCl. The resulting mixture was extracted with ethyl acetate (2 x 150 ml), and the combined organic layers were washed with saturated aqueous sodium chloride solution (500 ml), dried over sodium sulfate, filtered, and concentrated to provide **T15** as a white solid (85 g, 100%). This material was used directly in the following step. ^1^H NMR (400 MHz, DMSO-*d*_6_) d 12.54 (s, 1H), 6.67 (d, *J* = 9.4 Hz, 1H), 4.13 (s, 1H), 4.04 (s, 1H), 3.91 (d, *J* = 10.4 Hz, 1H), 3.77 (dd, *J* = 10.2, 5.3 Hz, 1H), 1.54 – 1.46 (m, 1H), 1.40 (s, 1H), 1.35 (s, 9H), 1.01 (s, 3H), 0.93 (s, 9H), 0.84 (s, 3H). LCMS *m/z* 369.3 [M+H]^+^.

*(1R,2S,5S)-6,6-Dimethyl-3-(3-methyl-L-valyl)-3-azabicyclo[3.1.0]hexane-2-carboxylic acid (**T16**), HCl salt*. A solution of HCl in 1,4-dioxane (4 M; 275 ml, 1.1 mol) was added to a solution of **T15** (81 g, 219 mmol) in dichloromethane (220 ml), and the reaction mixture was stirred at 25 °C for 16 h. Removal of solvents afforded HCl salt of **T16** as a white solid (66.5 g, 99%). This material was used directly in the following step. ^1^H NMR (400 MHz, DMSO-*d*_6_) d 12.81 (m, 1H), 8.18 (br s, 2H), 4.18 (s, 1H), 3.86 – 3.74 (m, 2H), 3.71 (d, *J* = 10.9 Hz, 1H), 1.60 – 1.51 (m, 1H), 1.46 (d, *J* = 7.7 Hz, 1H), 1.03 (d, *J* = 2.8 Hz, 12H), 0.96 (s, 3H). LCMS *m/z* 269.3 [M+H]^+^.

*(1R,2S,5S)-6,6-Dimethyl-3-[3-methyl-N-(trifluoroacetyl)-L-valyl]-3- azabicyclo[3.1.0]hexane-2-carboxylic acid (**T17**)*. To a 0 °C solution of the HCl salt of **T16** (55 g, 280 mmol) in methanol (180 ml) was added triethylamine (151 ml, 1.08 mol), followed by ethyl trifluoroacetate (64.1 g, 451 mmol), whereupon the reaction mixture was allowed to warm to 50 °C, and was stirred for 16 h. It was then concentrated *in vacuo* at 50 °C, and the residue was diluted with water (250 ml) and adjusted to a pH of 3 to 4 by addition of 1 M HCl (250 ml) followed by conc. HCl. After extraction of the aqueous layer with ethyl acetate (3 x 250 ml), the combined organic layers were washed with saturated aqueous sodium chloride solution (300 ml), dried over sodium sulfate, filtered, and concentrated to afford **T17** as a white solid (70 g, 71% corrected for residual solvent (qNMR potency: 0.817)). ^1^H NMR (400 MHz, DMSO-*d*_6_) d 12.99 – 12.47 (m, 1H), 9.43 (d, *J* = 8.5 Hz, 1H), 4.44 (d, *J* = 8.5 Hz, 1H), 4.15 (s, 1H), 3.85 (dd, *J* = 10.5, 5.3 Hz, 1H), 3.72 (d, *J* = 10.5 Hz, 1H), 1.53 (dd, *J* = 7.4, 5.2 Hz, 1H), 1.43 (d, *J* = 7.6 Hz, 1H), 1.01 (d, *J* = 3.3 Hz, 12H), 0.82 (s, 3H). LCMS *m/z* 365.1 [M+H]^+^.

*(1R,2S,5S)-N-{(2S)-1-Amino-1-oxo-3-[(3S)-2-oxopyrrolidin-3-yl]propan-2-yl}-6,6- dimethyl-3-[3-methyl-N-(trifluoroacetyl)-L-valyl]-3-azabicyclo[3.1.0]hexane-2-carboxamide (**T18**)*. 2-Hydroxypyridine 1-oxide ( 3.7 g, 34 mmol) was added to a solution of **T17** (59.9 g, 134 mmol, 81.7wt%) and the HCl salt of **T13** (33.4 g, 152 mol) in butan-2-one (540 ml), and the mixture was cooled to 0 °C. *N,N*-Diisopropylethylamine (70 ml, 403 mmol) was then added, followed by the addition of EDCI(30.9 g, 161 mmol). The reaction mixture was stirred at 25 °C for 16 h, whereupon it was diluted with ethyl acetate/ *tert*-butyl methyl ether (1:1, 500 ml) and washed by a mixture of water (200 ml) and saturated aqueous sodium chloride solution (100 ml). The separated organic layer was washed with saturated aqueous sodium chloride solution (300 ml), followed by a mixture of HCl (1 M; 200 ml, 200 mmol) and saturated aqueous sodium chloride solution (100 ml), then saturated aqueous sodium chloride solution (300 ml), dried over magnesium sulfate, filtered, and concentrated to afford **T18** as a white solid (71.4 g, 98% corrected for residual solvent). ^1^H NMR (400 MHz, DMSO-*d*_6_) d 9.41 (d, *J* = 8.6 Hz, 1H), 8.29 (d, *J* = 8.9 Hz, 1H), 7.55 (s, 1H), 7.31 (br s, 1H), 7.03 (br s, 1H), 4.43 (d, *J* = 8.6 Hz, 1H), 4.35 – 4.25 (m, 2H), 3.91 – 3.84 (m, 1H), 3.67 (d, *J* = 10.4 Hz, 1H), 3.13 (t, *J* = 9.0 Hz, 1H), 3.06 – 2.97 (m, 1H), 2.45 – 2.34 (m, 1H), 2.14 (dt, *J* = 10.5, 7.4 Hz, 1H), 1.97 – 1.86 (m, 1H), 1.70 – 1.57 (m, 1H), 1.54 – 1.45 (m, 2H), 1.38 (d, *J* = 7.7 Hz, 1H), 1.10 (s, 3H), 0.98 (s, 9H), 0.84 (s, 3H). LCMS *m/z* 518.4 [M+H]^+^.

*(1R,2S,5S)-N-{(1S)-1-Cyano-2-[(3S)-2-oxopyrrolidin-3-yl]ethyl}-6,6-dimethyl-3-[3-methyl- N-(trifluoroacetyl)-L-valyl]-3-azabicyclo[3.1.0]hexane-2-carboxamide (1 eq tert-butyl methyl ether solvate) (**6,** MTBE solvate)*. This experiment was carried out in 2 parallel batches.

Methyl *N*-(triethylammoniosulfonyl)carbamate, inner salt (Burgess reagent; 69.3 g, 276 mmol) was added to a solution of **T18** (61 g, 111 mmol) in dichloromethane (550 ml). After the reaction mixture had been stirred at 25 °C for 1 h. The reaction mixture was quenched by a mixture of saturated aqueous sodium bicarbonate solution (200 ml) and saturated aqueous sodium chloride solution (100 ml). The separated organic phase was concentrated. The resulting residue was dissolved in 50% ethyl acetate/ *tert*-butyl methyl ether (600 ml), washed by a mixture of saturated aqueous sodium bicarbonate solution (200 ml) and saturated aqueous sodium chloride solution (100 ml) twice, saturated aqueous sodium chloride solution (200 ml), a mixture of HCl (1 M; 200 ml) and saturated aqueous sodium chloride solution (100 ml) twice. The organic layer was then dried over magnesium sulfate, filtered, and concentrated. The residue was treated with a mixture of ethyl acetate and *tert*-butyl methyl ether (1:10, 400 ml) and heated to 50 °C; after stirring for 1 hour at 50 °C, it was cooled to 25 °C and stirred overnight. The solid was collected via filtration, dissolved in dichloromethane (100 ml) and filtered through silica gel (200 g); the silica gel was then washed with ethyl acetate (1 Liter), 10% methanol in ethyl acetate (2 Liters). The combined eluates were concentrated. The 2 batches were combined, taken up in a mixture of ethyl acetate and *tert*-butyl methyl ether (5:95, 550 ml). This mixture was heated to 50 °C for 1 h, cooled to 25 °C, and stirred overnight. Filtration afforded **6**, MTBE solvate, as a white solid. Yield: 104 g, 75 %. ^1^H NMR (600 MHz, DMSO-*d*_6_) δ 9.43 (d, *J* = 8.4 Hz, 1H), 9.03 (d, *J* = 8.6 Hz, 1H), 7.68 (s, 1H), 4.97 (ddd, *J* = 10.9, 8.6, 5.1 Hz, 1H), 4.41 (d, *J* = 8.4 Hz, 1H), 4.15 (s, 1H), 3.91 (dd, *J* = 10.4, 5.5 Hz, 1H), 3.69 (d, *J* = 10.4 Hz, 1H), 3.17 – 3.11 (m, 1H), 3.07 (s, 3H, MTBE), 3.04 (td, *J* = 9.4, 7.1 Hz, 1H), 2.40 (tdd, *J* = 10.4, 8.4, 4.4 Hz, 1H), 2.14 (ddd, *J* = 13.4, 10.9, 4.4 Hz, 1H), 2.11 – 2.03 (m, 1H), 1.76 – 1.65 (m, 2H), 1.57 (dd, *J* = 7.6, 5.5 Hz, 1H), 1.32 (d, *J* = 7.6 Hz, 1H), 1.10 (s, 9H, MTBE), 1.03 (s, 3H), 0.98 (s, 9H), 0.85 (s, 3H).

Recrystallization of PF-7321332 (**6**): Generation of Anhydrous ’Form 1’

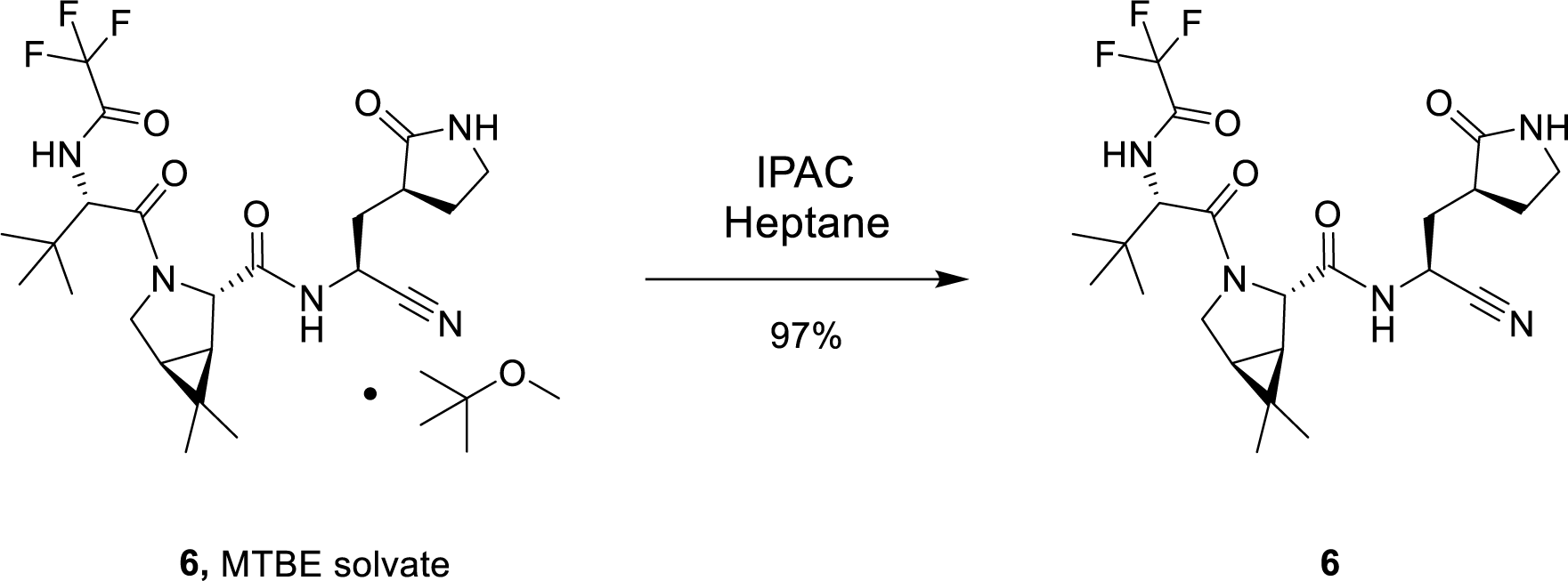

Compound **6** (anhydrous MTBE solvate, 200 g, 332.8 mmol, 83.11 mass%) was charged into a reactor with overhead half-moon stirring at 350 rpm. Heptane (1000 ml) was charged, followed by isopropyl acetate (1000 ml) and the stirring was continued at 20 °C overnight. Additional heptane (1000 ml) was charged over 120 minutes. The reaction vessel was then cooled to 10 °C over 30 min and stirred at that temp for 3 days. The solid was filtered, washing with a mixture of isopropyl acetate (80 ml) and heptane (320 ml). It was then dried under vacuum at 50 °C to provide **6**, anhydrous ’Form 1’, as a white crystalline solid. Yield: 160.93 g, 322 mmol, 97%. ^1^H NMR (600 MHz, DMSO-*d*_6_) δ 9.43 (d, *J* = 8.4 Hz, 1H), 9.03 (d, *J* = 8.6 Hz, 1H), 7.68 (s, 1H), 4.97 (ddd, *J* = 10.9, 8.6, 5.1 Hz, 1H), 4.41 (d, *J* = 8.4 Hz, 1H), 4.15 (s, 1H), 3.91 (dd, *J* = 10.4, 5.5 Hz, 1H), 3.69 (d, *J* = 10.4 Hz, 1H), 3.17 – 3.11 (m, 1H), 3.04 (td, *J* = 9.4, 7.1 Hz, 1H), 2.40 (tdd, *J* = 10.4, 8.4, 4.4 Hz, 1H), 2.14 (ddd, *J* = 13.4, 10.9, 4.4 Hz, 1H), 2.11 – 2.03 (m, 1H), 1.76 – 1.65 (m, 2H), 1.57 (dd, *J* = 7.6, 5.5 Hz, 1H), 1.32 (d, *J* = 7.6 Hz, 1H), 1.03 (s, 3H), 0.98 (s, 9H), 0.85 (s, 3H). ^13^C NMR (151 MHz, DMSO-*d*_6_) δ 177.50, 170.72, 167.45, 156.95 (q, *J* = 37.0 Hz), 119.65, 115.84 (q, *J* = 286.9 Hz), 60.08, 58.19, 47.63, 37.77, 36.72, 34.60, 34.15, 30.28, 27.34, 26.86, 26.26, 25.72, 18.86, 12.34. ^19^F NMR (376 MHz, DMSO-*d*_6_) δ -72.94. HRMS (ESI-TOF) *m/z* calcd. for C_23_H_33_F_3_N_5_O_4_ [M + H]^+^ 500.2474, found 500.2472.

#### NMR Spectra

^1^H Spectrum of **T3** in methanol-*d*_4_ at 25 °C.

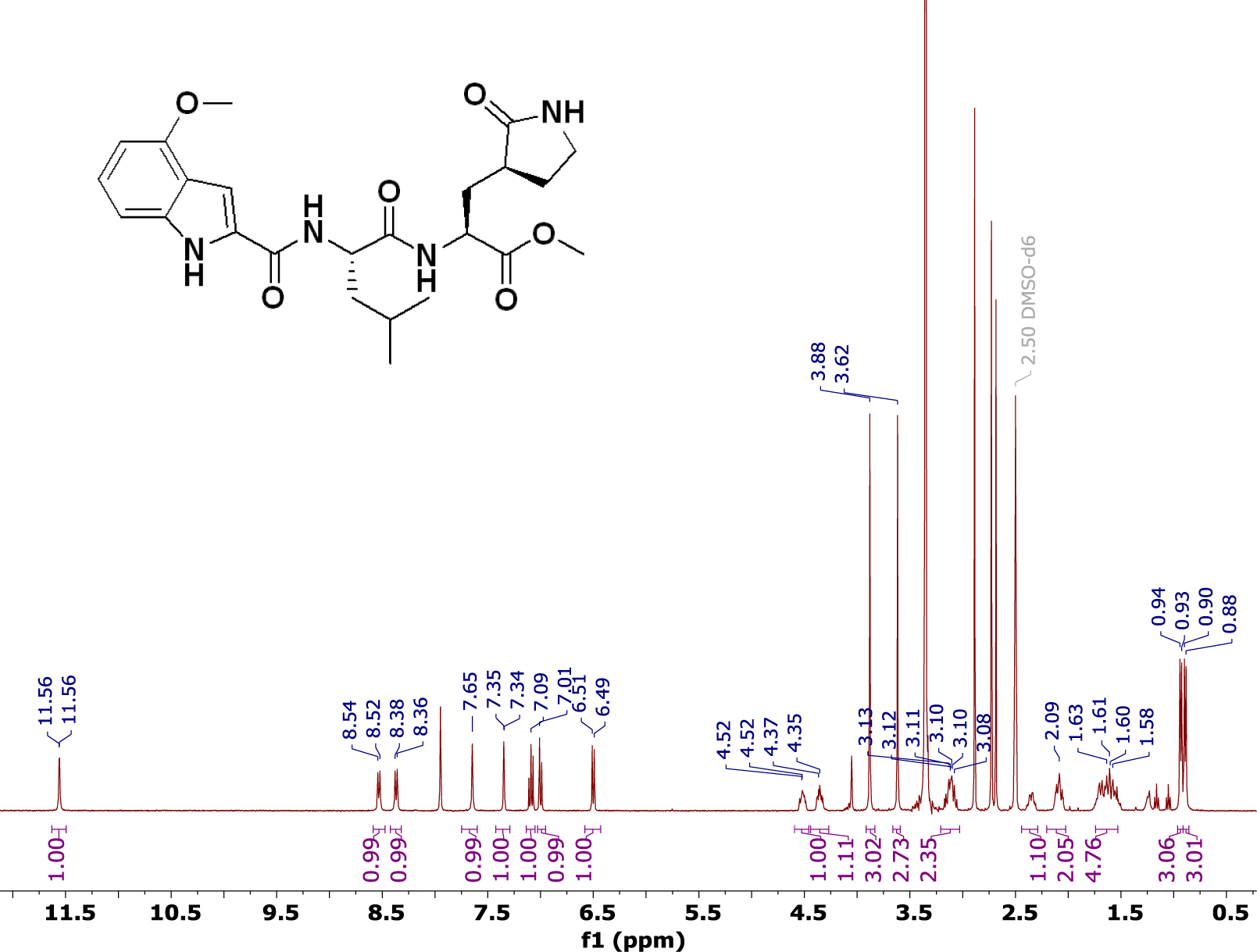

^1^H Spectrum of Compound **2** in DMSO-*d*6 at 27 °C.

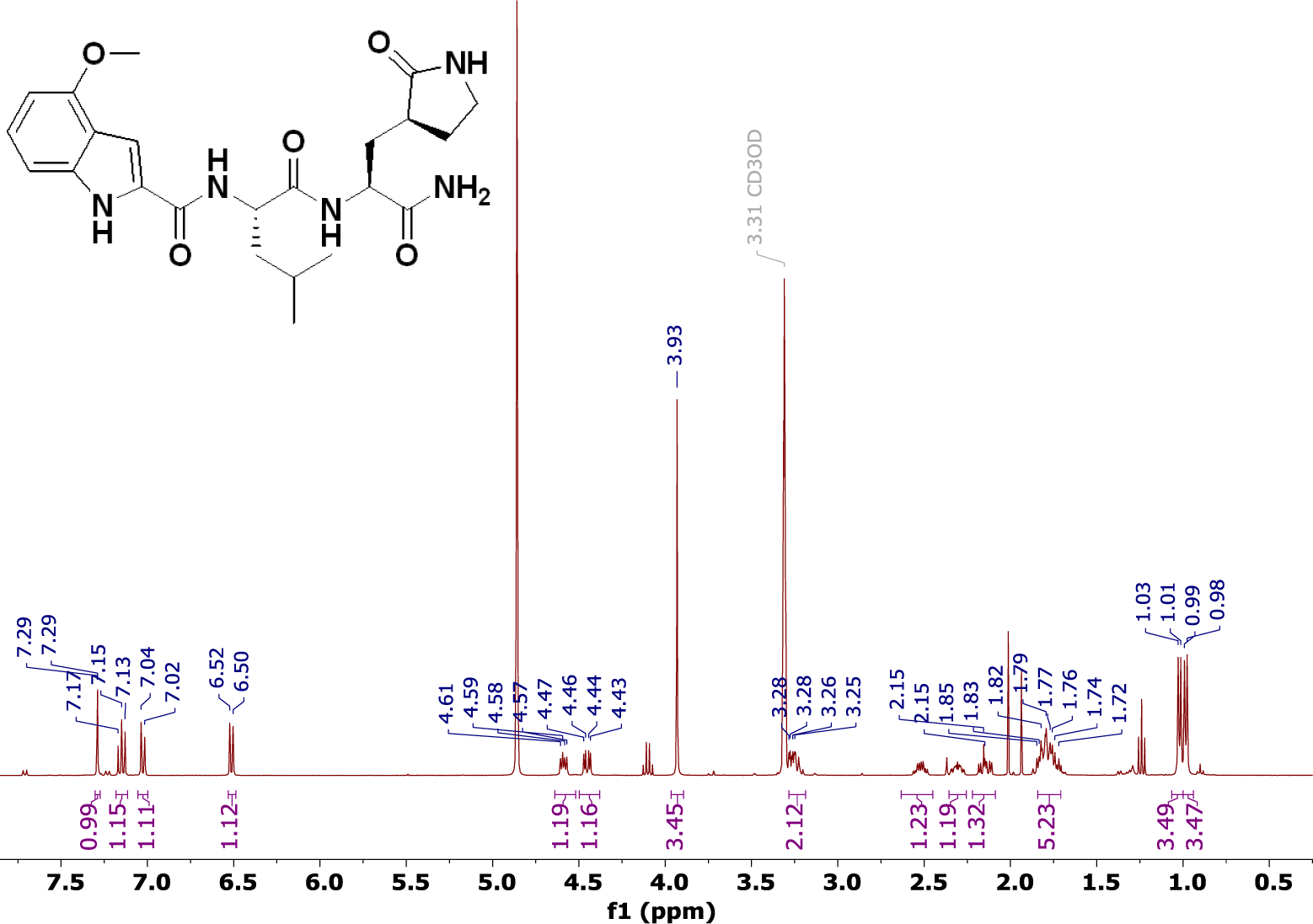

^13^C Spectrum of Compound **2** in DMSO-*d*6 at 27 °C.

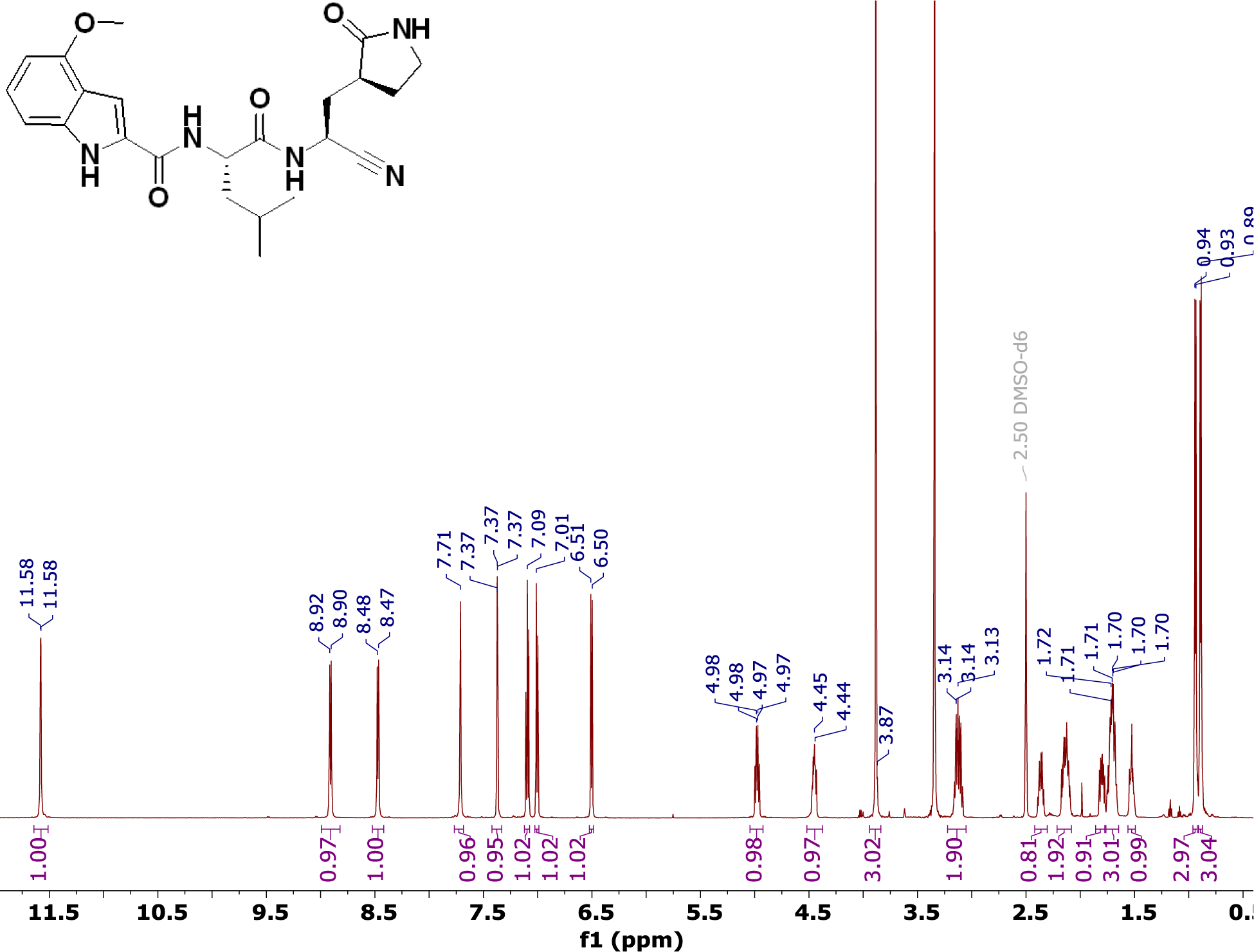

^1^H Spectrum of T4 in methanol-*d*4 at 25 °C.

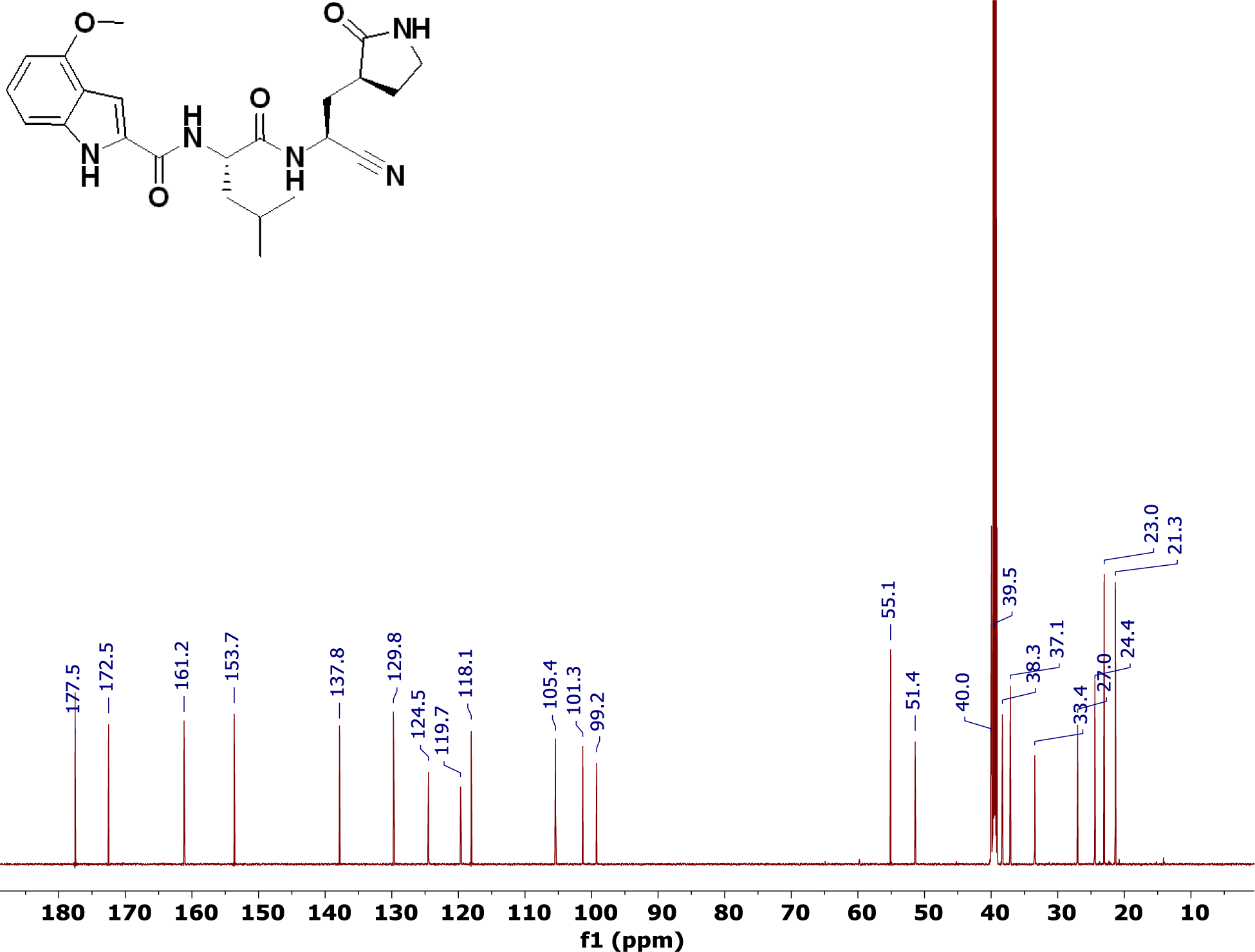

^1^H Spectrum of **T5** in chloroform-*d* at 25 °C.

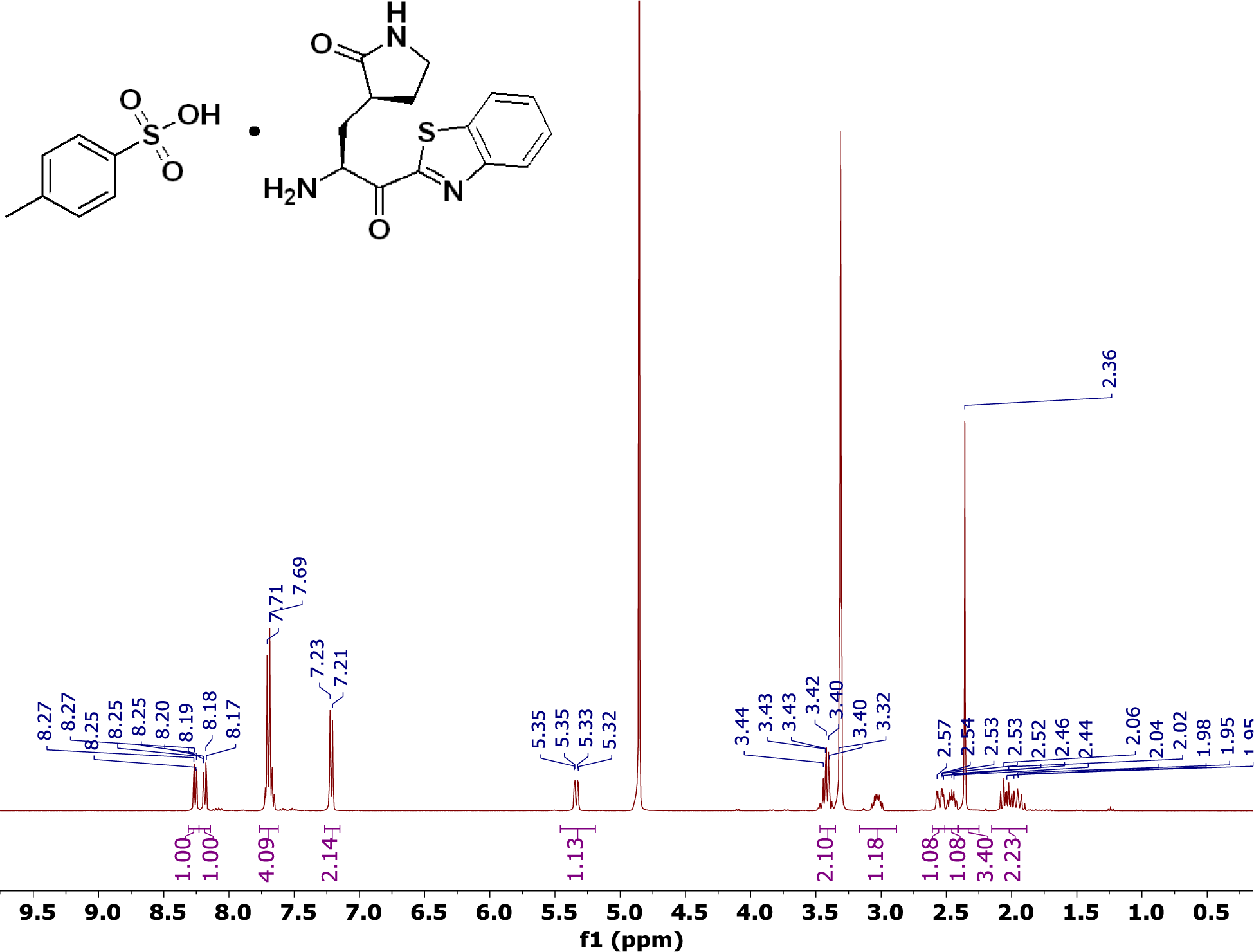

^1^H Spectrum of Compound **3** in DMSO-*d*6 at 27 °C. This compound exists as a mixture of rotamers at room temperature and resonances are doubled at ∼ 1:1 ratio.

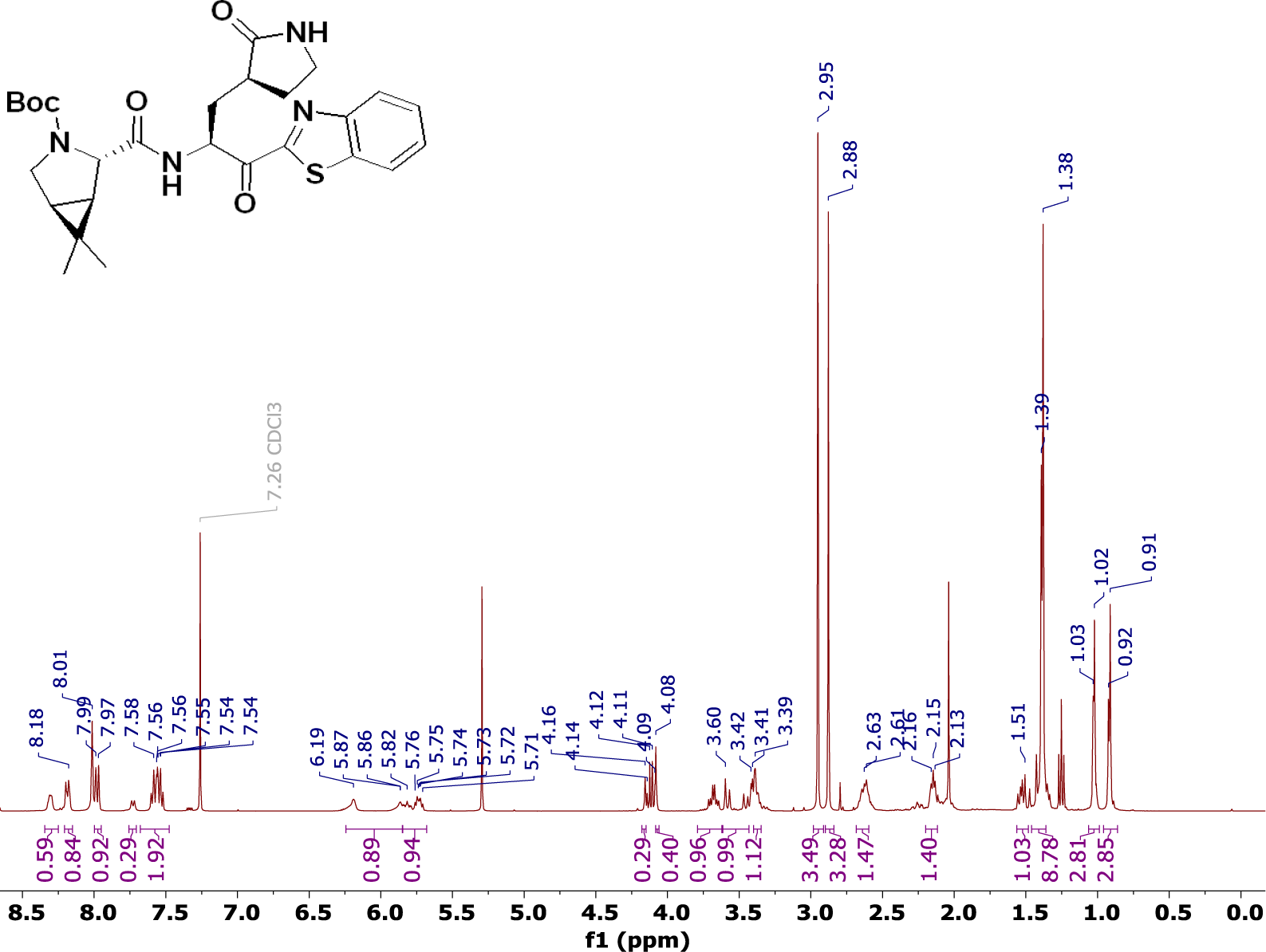

^13^C Spectrum of Compound **3** in DMSO-*d*6 at 27 °C. This compound exists as a mixture of rotamers at room temperature and resonances are doubled.

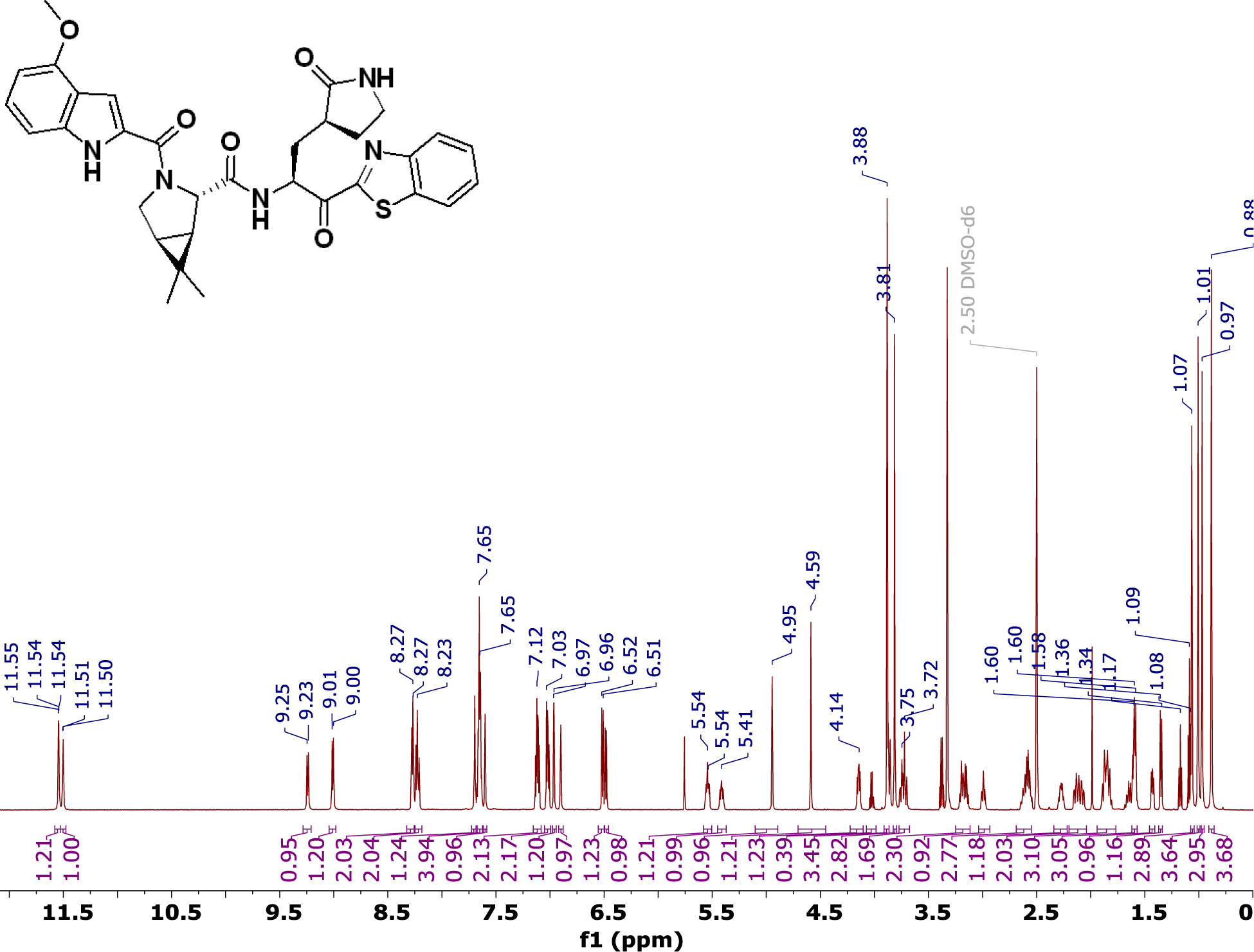

The ^1^H spectra of Compound **3** in DMSO-*d*6 at increasing temperatures. At room temperature, many of the resonances have restricted rotation and are observed as two distinct resonances (rotamers). At 140 °C, the rotation is no longer restricted and the resonances appear as a single peak. For example, the resonances at δ 4.9 and 4.6 ppm observed at 27 °C coalesce to a single resonance at δ 4.75 ppm at 140 °C.

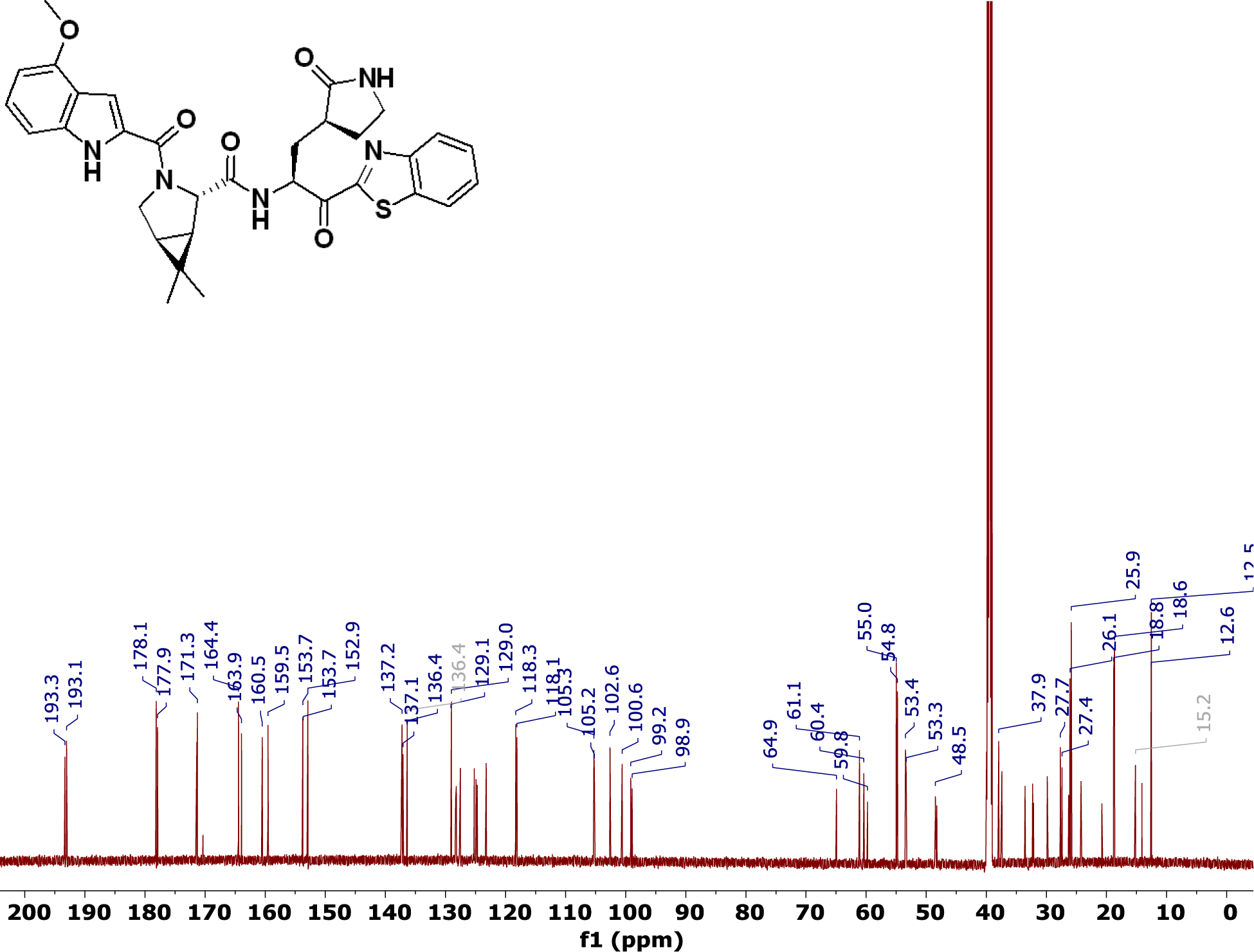

The ^1^H-^1^H NOESY spectra of Compound **3** in DMSO-*d*6 at 60°C. The resonances with restricted rotation are in chemical exchange and confirm the compound is a rotamer. The exchange peaks observed between the resonances with restricted rotation have the same sign (orange correlations) as the diagonal consistent with a transfer NOE. Actual through space NOEs in this experiment have the opposite sign as the diagonal and appear as blue correlations.

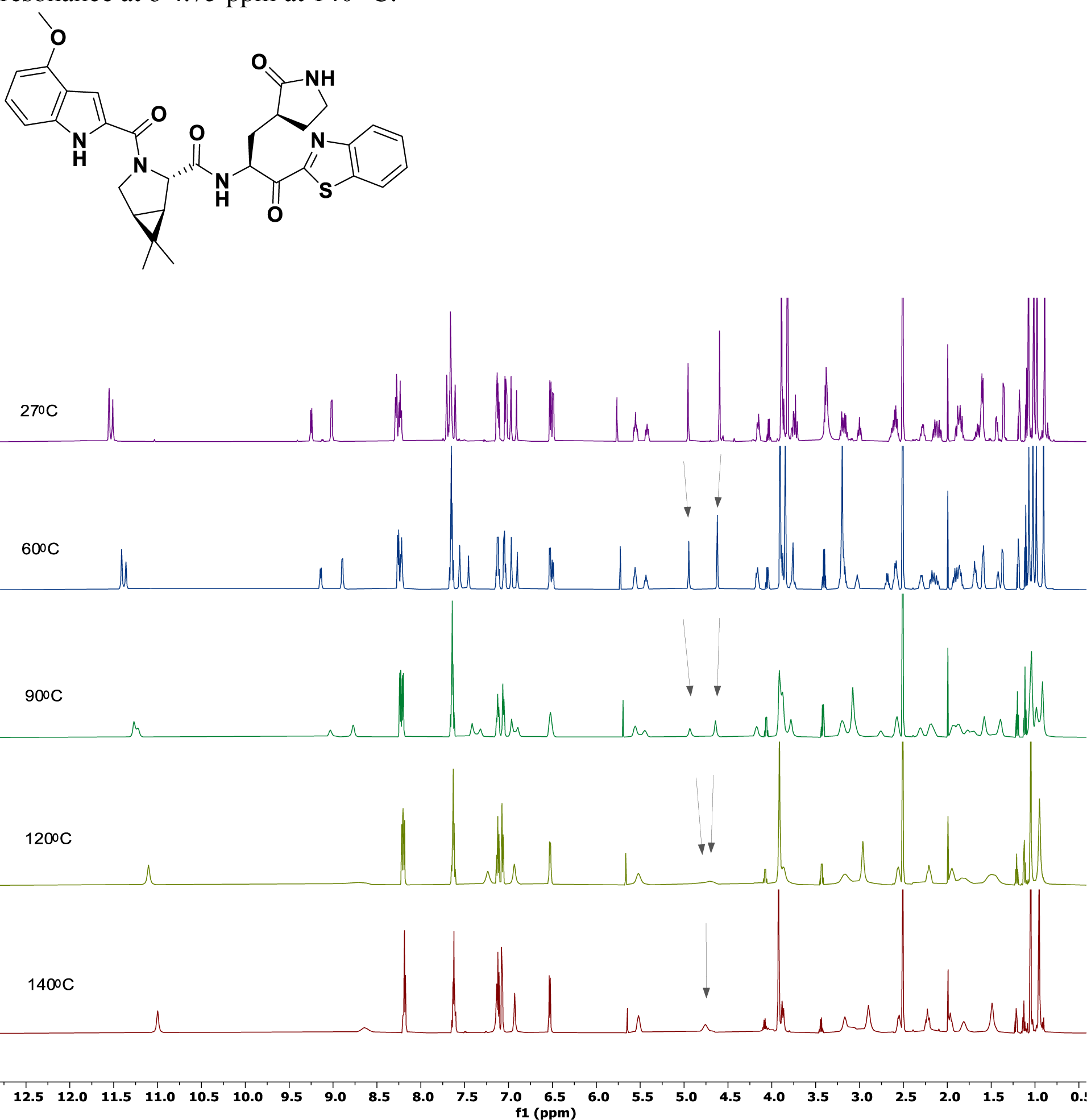

^1^H Spectrum of **T7** in chloroform-*d* at 25 °C.

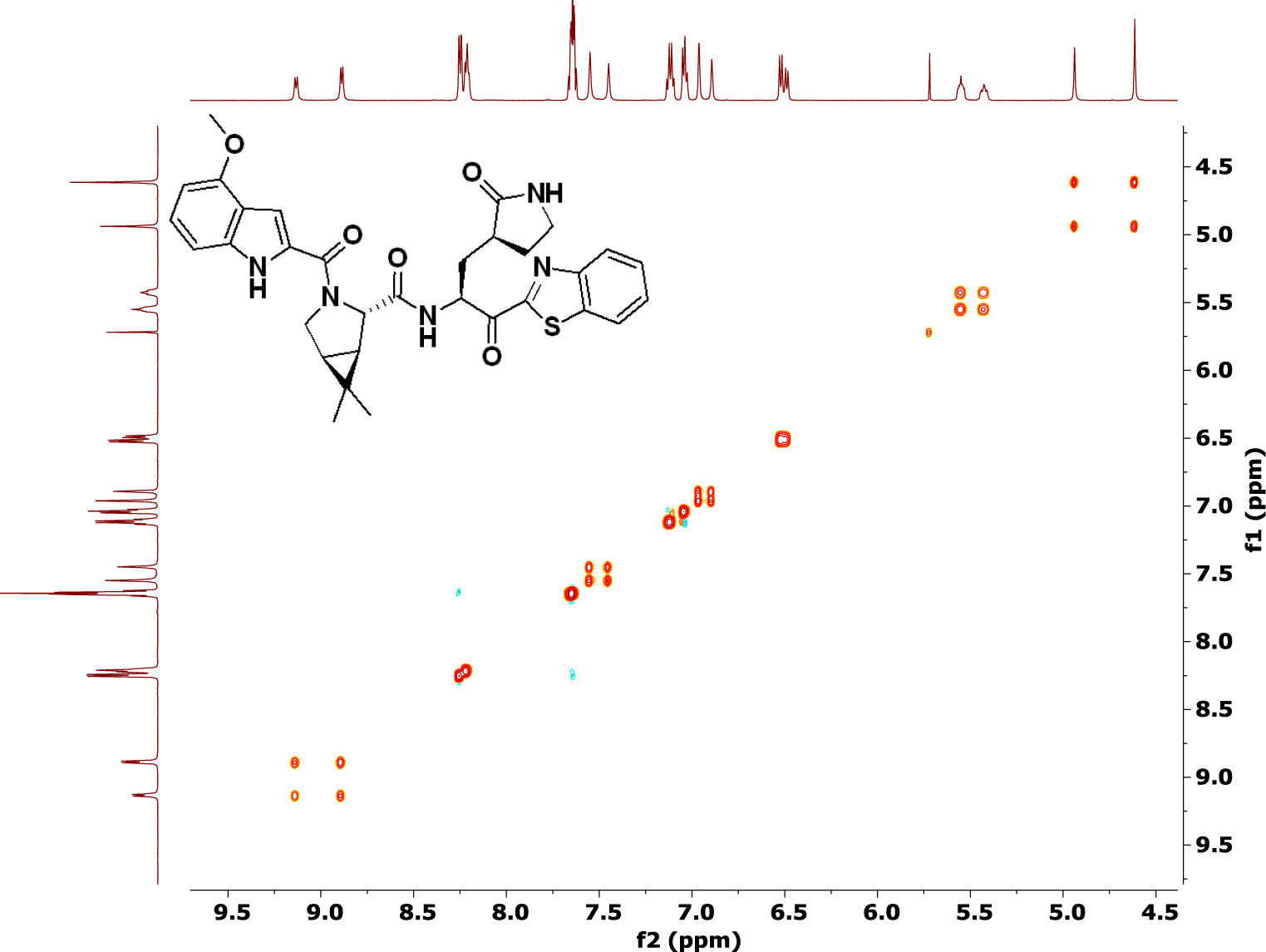

^1^H Spectrum of **T8** in DMSO-*d*6 at 25 °C.

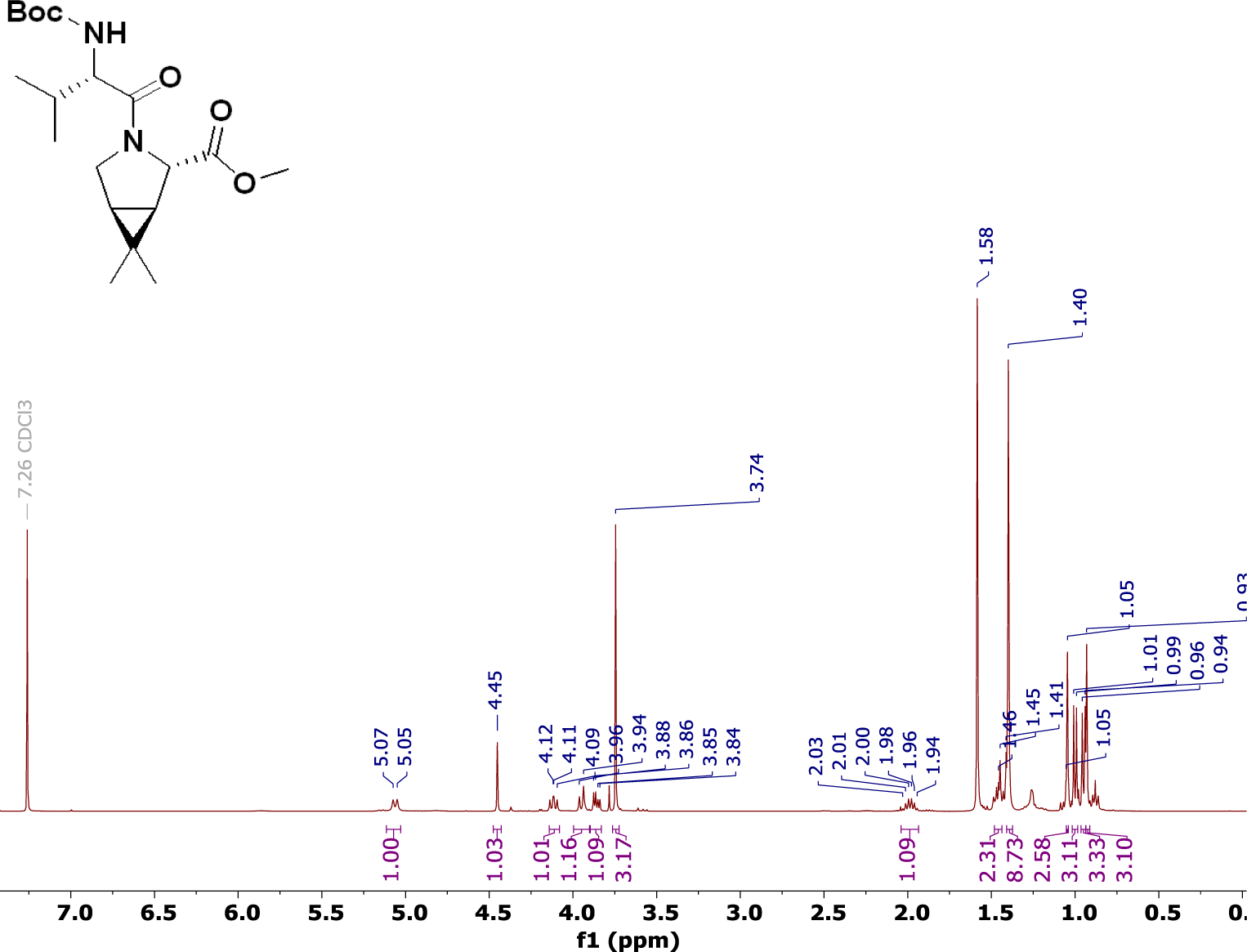

^1^H Spectrum of **T9** in chloroform-*d* at 25 °C.

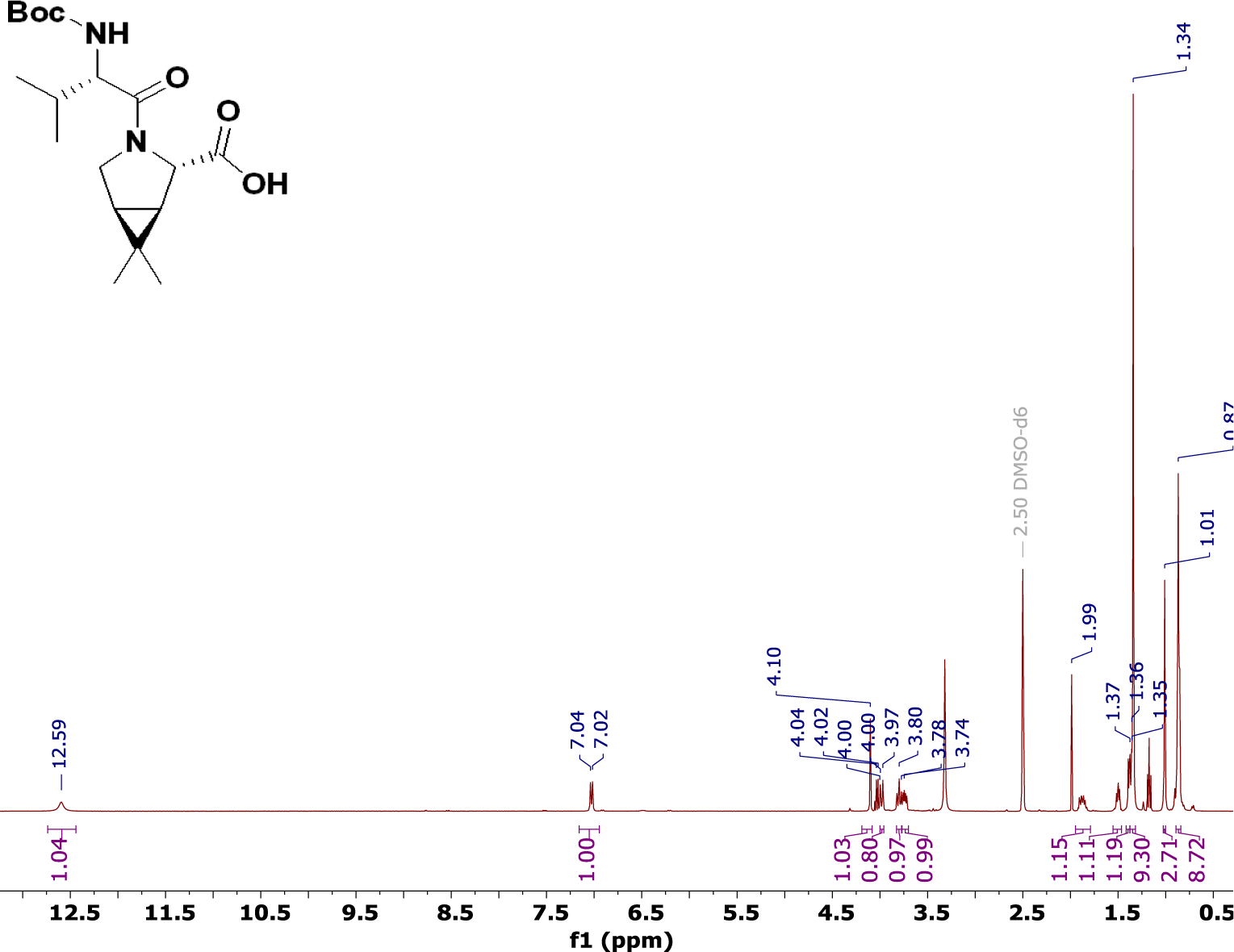

^1^H Spectrum of Compound **4** in DMSO-*d*6 at 27 °C. This compound exists as a mixture of rotamers at room temperature and resonances are doubled at ∼3:1 ratio.

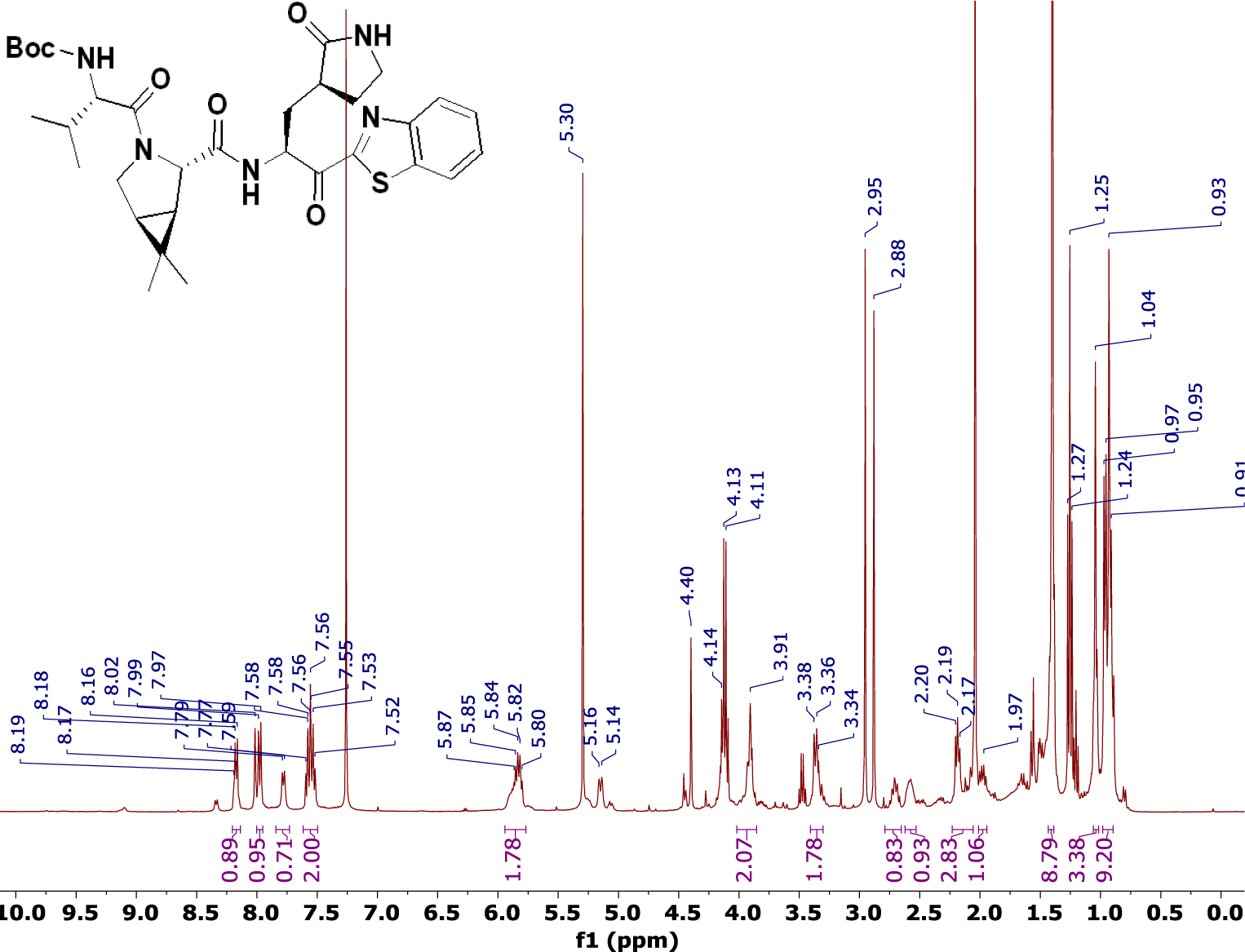

^13^C Spectrum of Compound **4** in DMSO-*d*6 at 27 °C. This compound exists as a mixture of rotamers at room temperature and resonances are doubled.

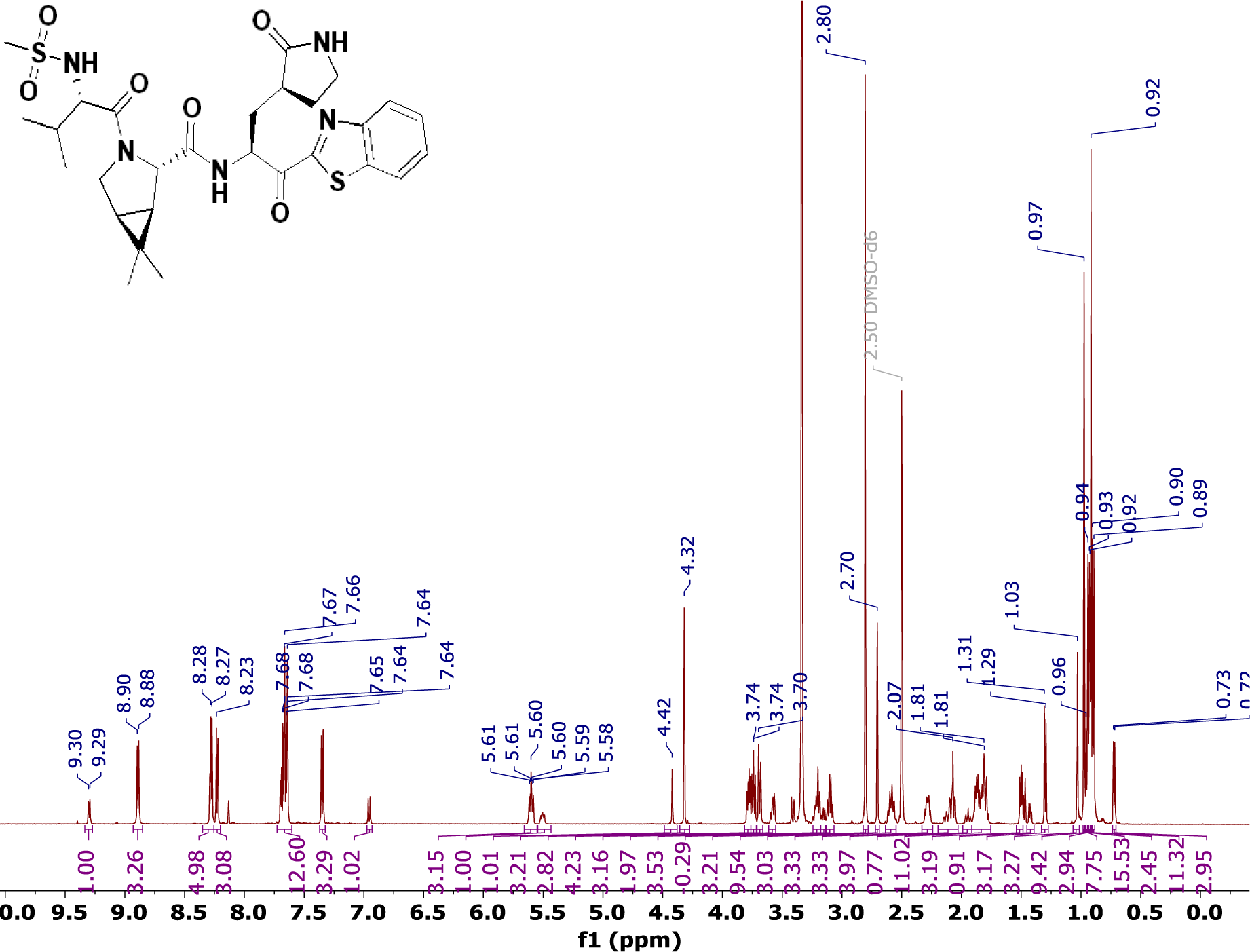

^1^H Spectrum of **T10** in methanol-*d*4 at 25 °C.

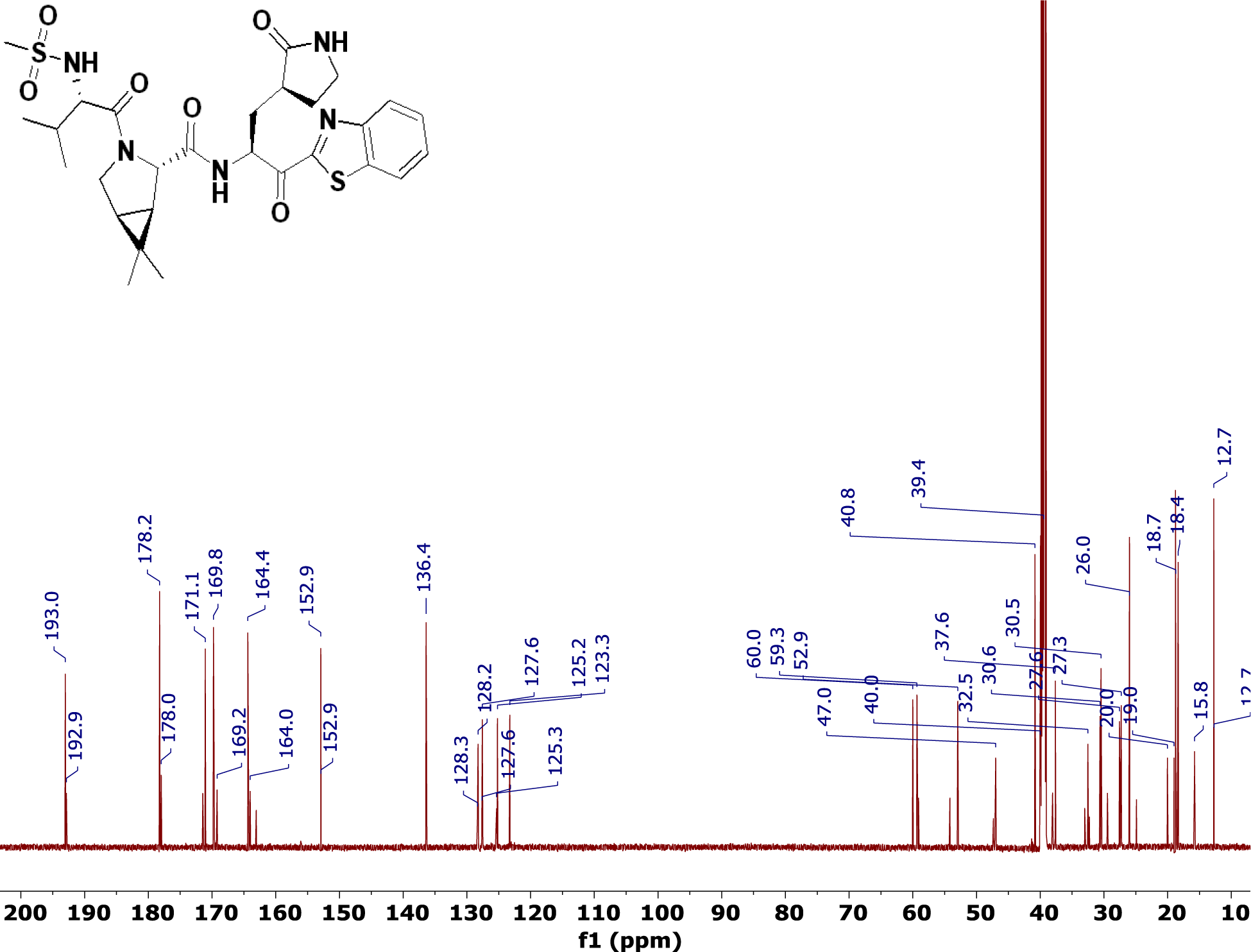

^1^H Spectrum of **T11** in methanol-*d*4 at 25 °C

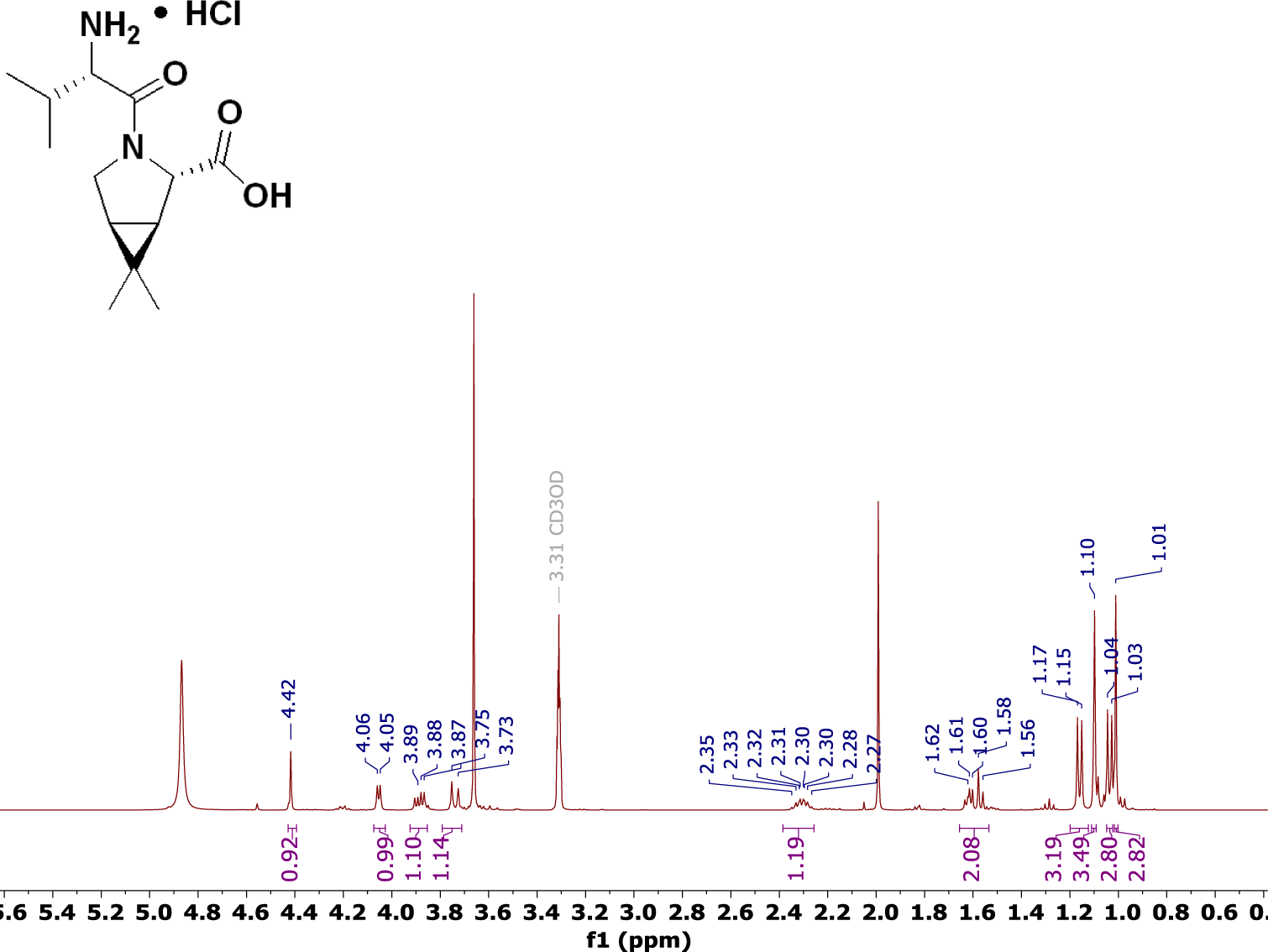

^1^H Spectrum of Compound **5** in DMSO-*d*6 at 25 °C.

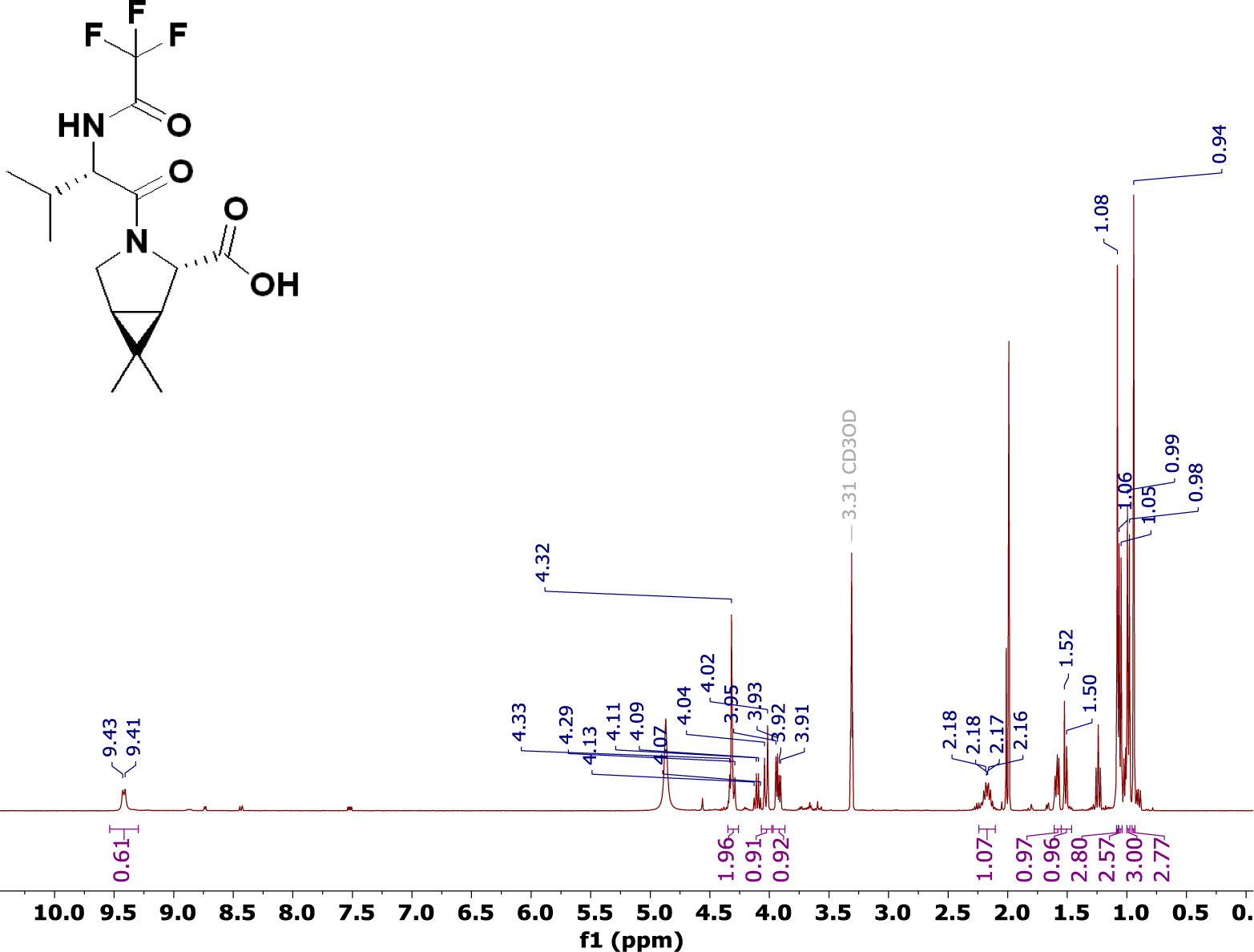

^13^C Spectrum of Compound **5** in DMSO-*d*6 at 25 °C.

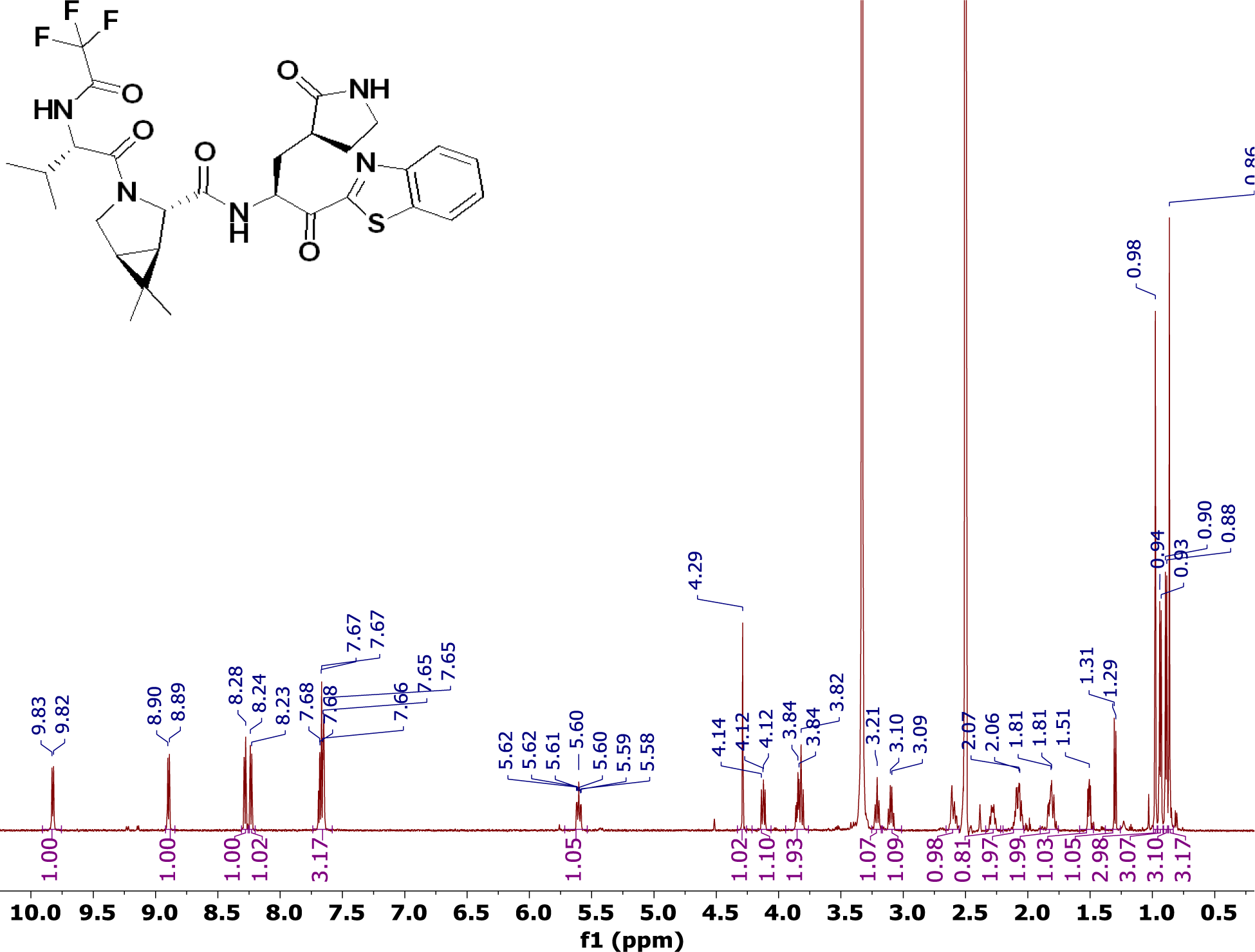

^19^F Spectrum of Compound **5** in DMSO-*d*6 at 25 °C.

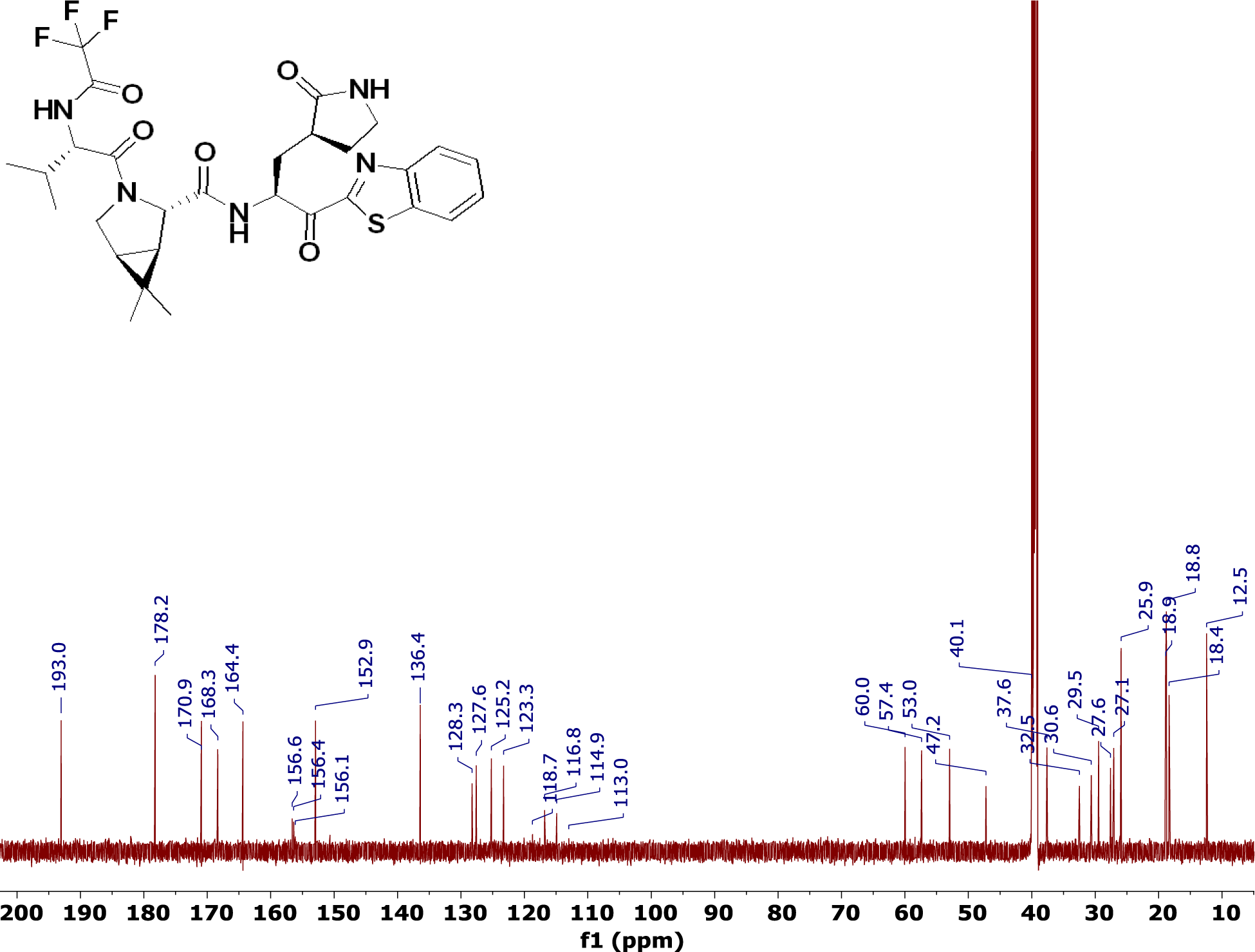

^1^H Spectrum of **T12** in methanol-*d*4 at 25 °C.

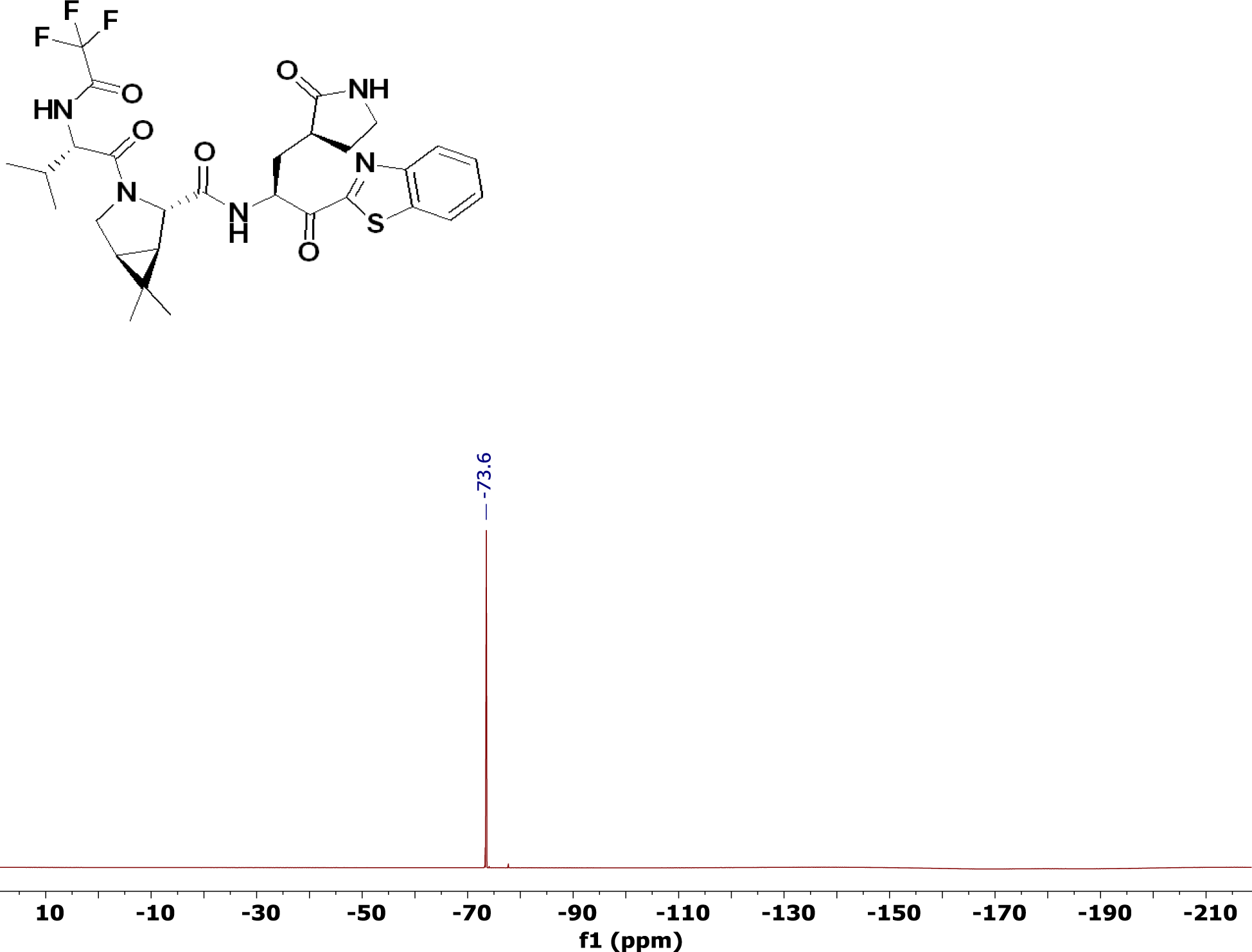

^1^H Spectrum of **T13** in methanol-*d*4 at 25 °C.

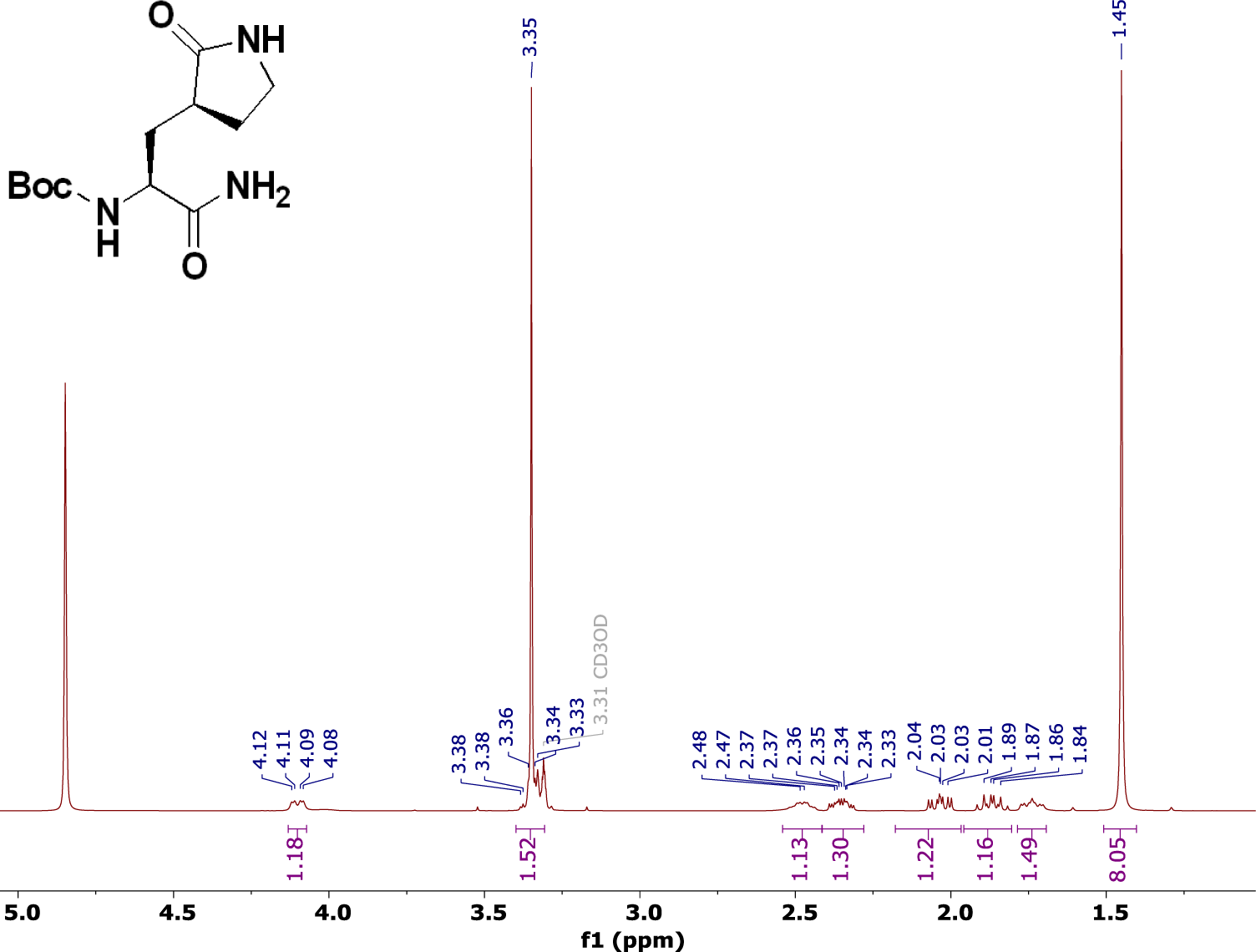

^1^H Spectrum of **T14** in DMSO-*d*6 at 25 °C.

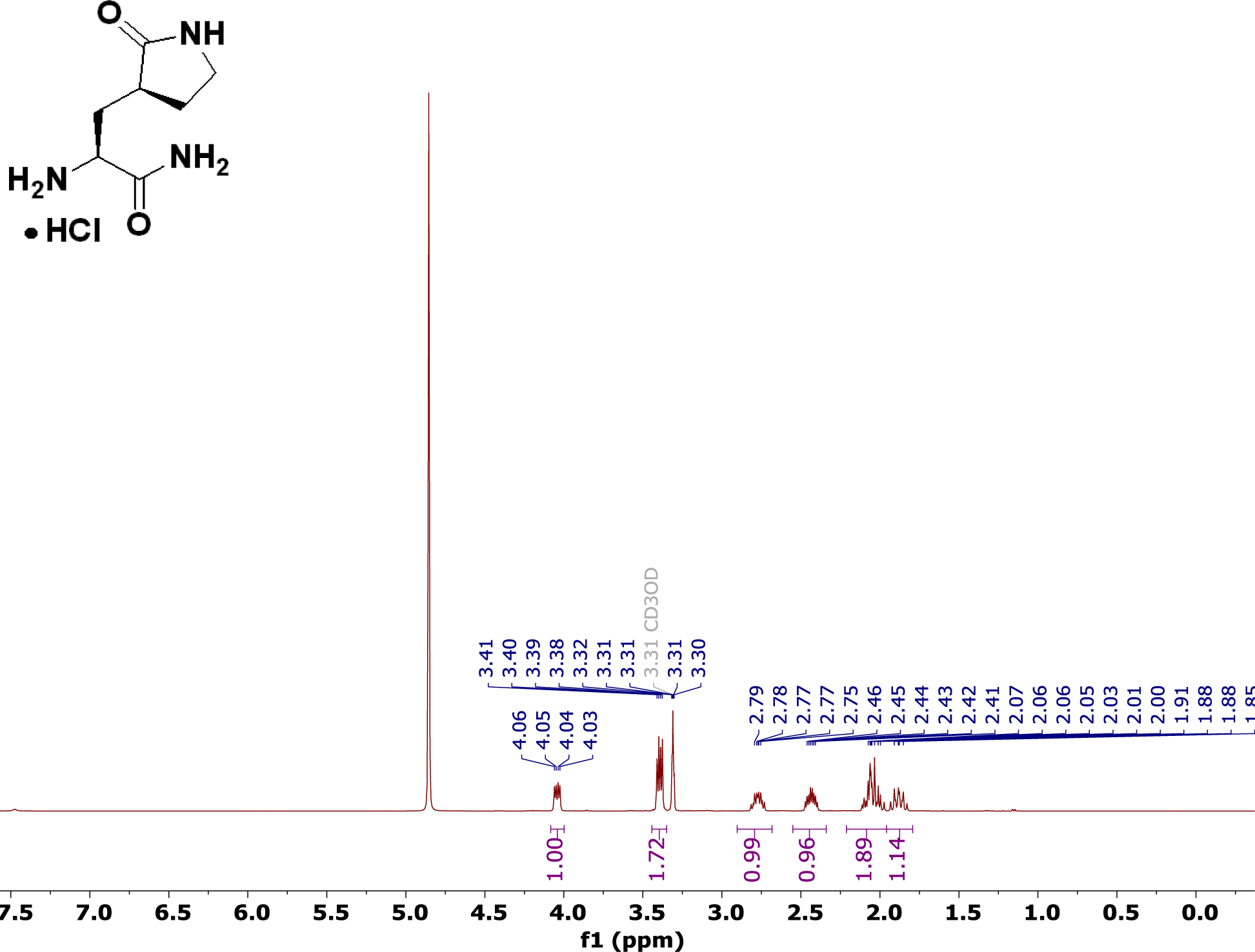

^1^H Spectrum of **T15** in DMSO-*d*6 at 25 °C.

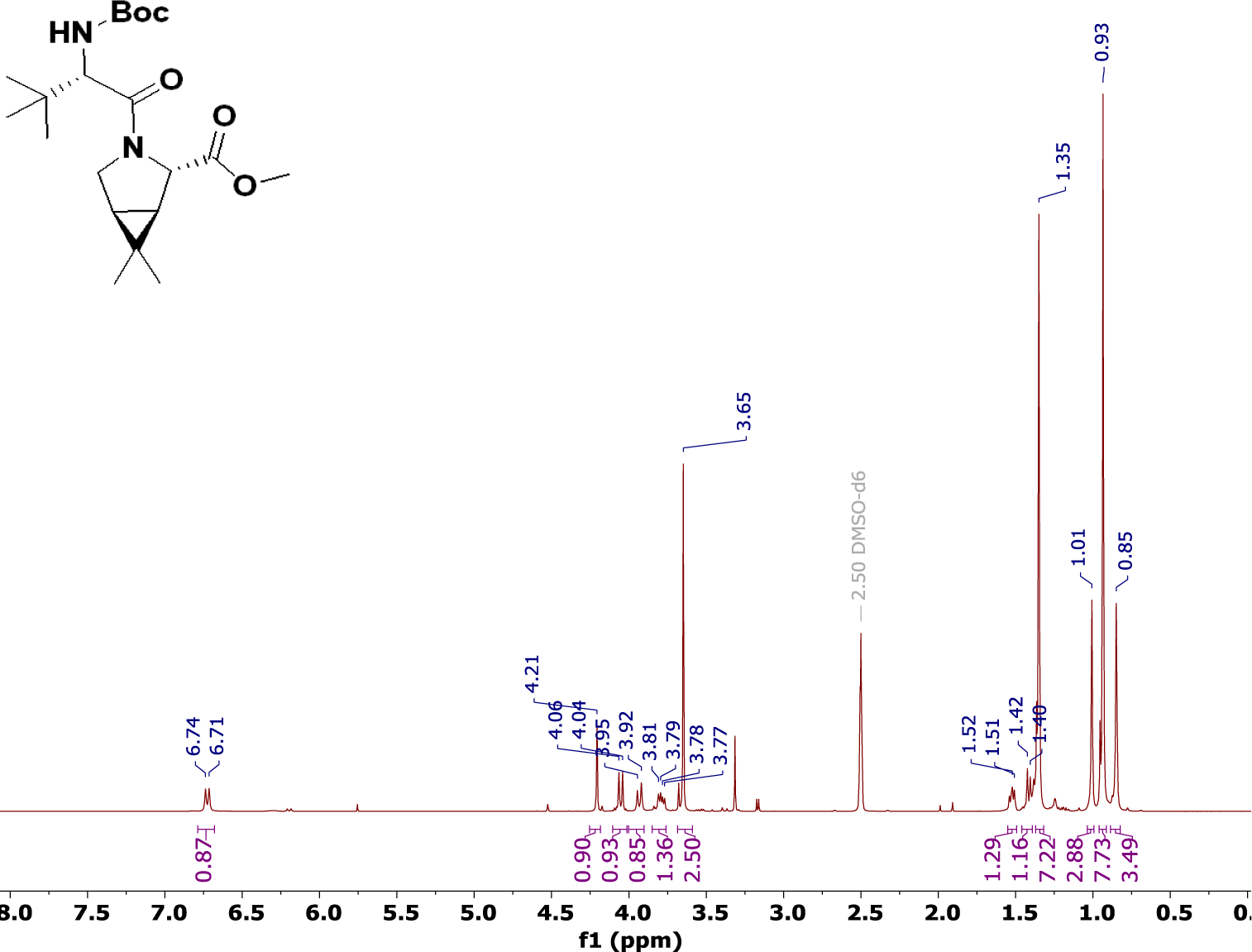

^1^H Spectrum of **T16** in DMSO-*d*6 at 25 °C.

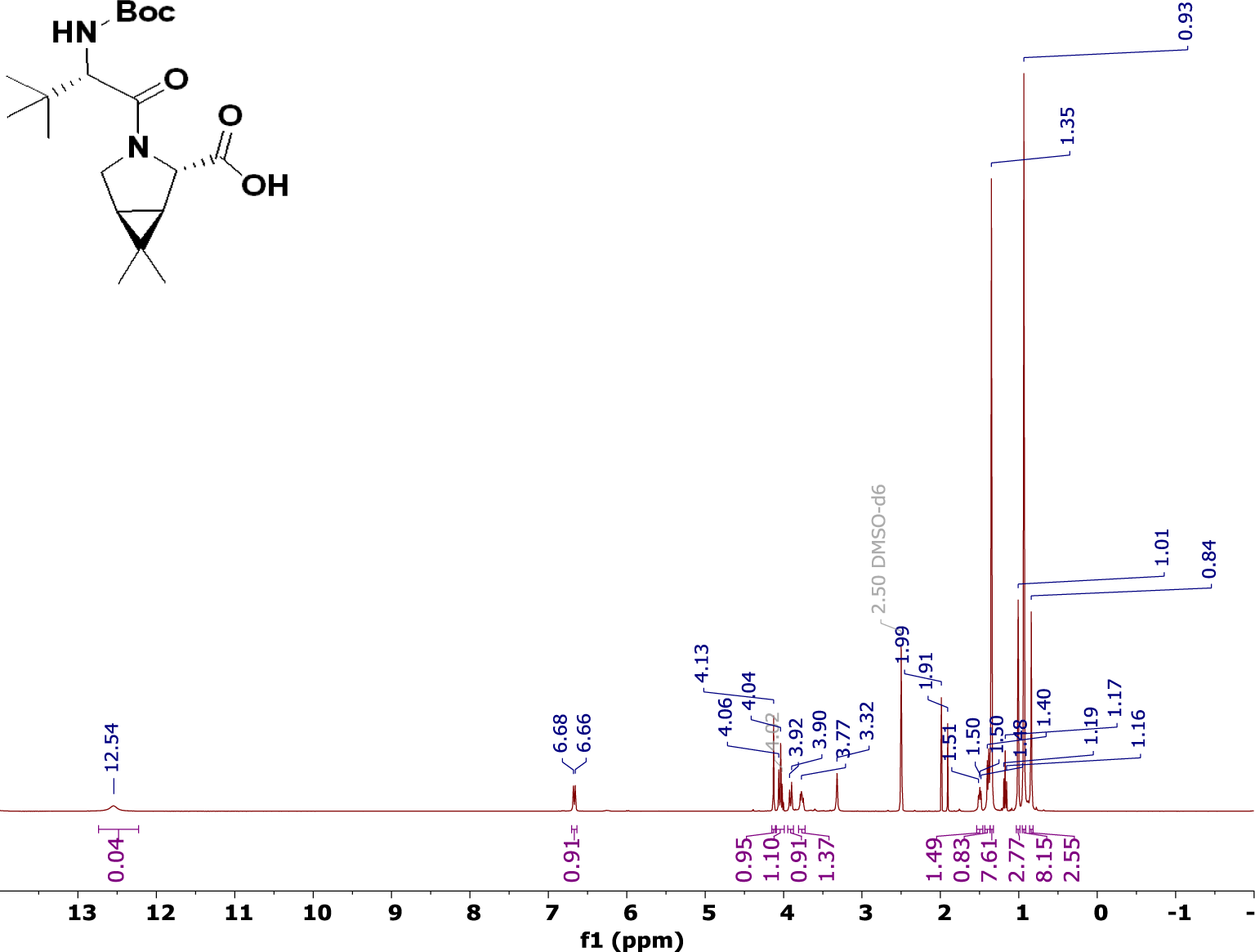

^1^H Spectrum of **T17** in DMSO-*d*6 at 25 °C.

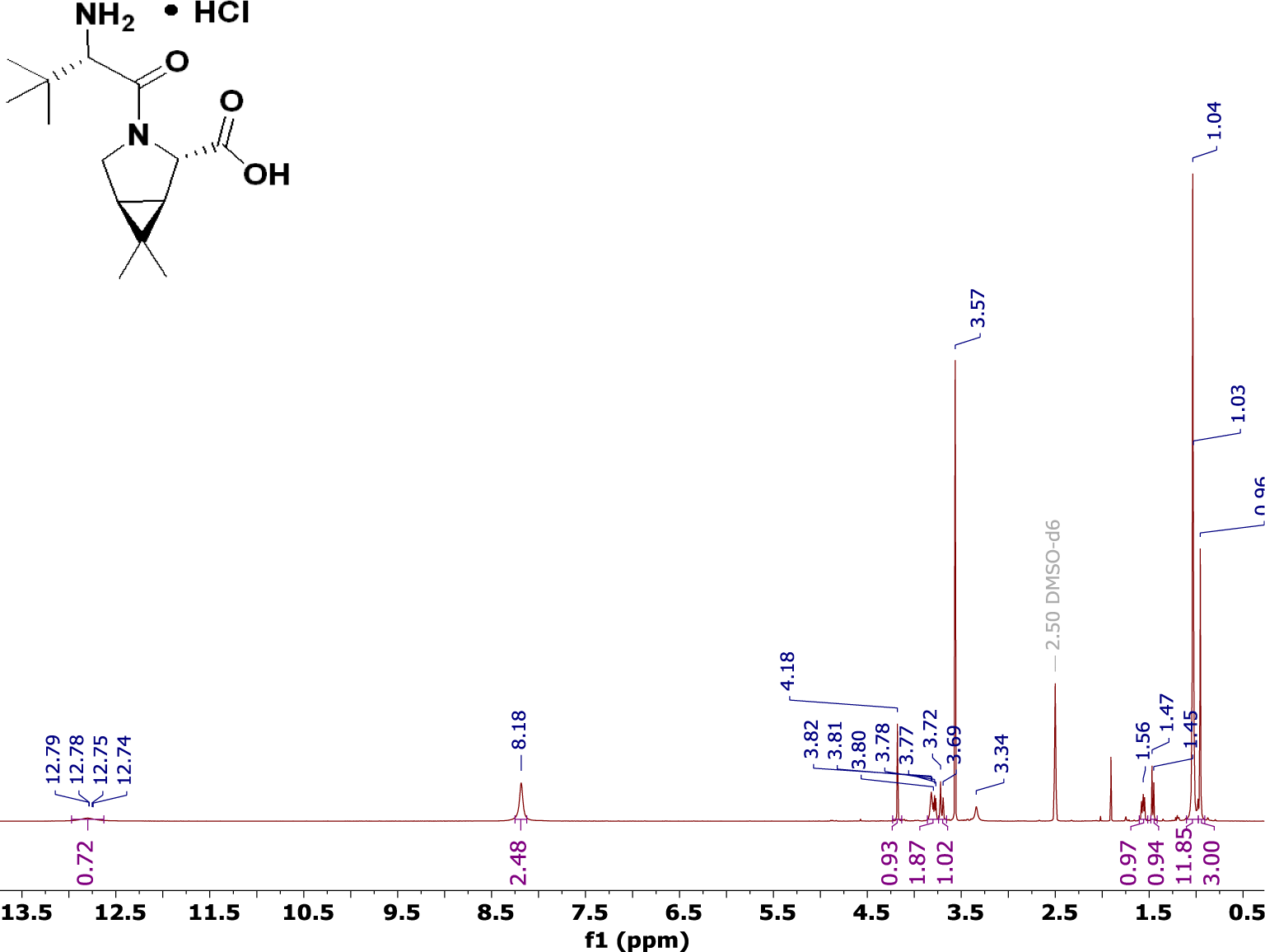

^1^H Spectrum of **T18** in DMSO-*d*6 at 25 °C.

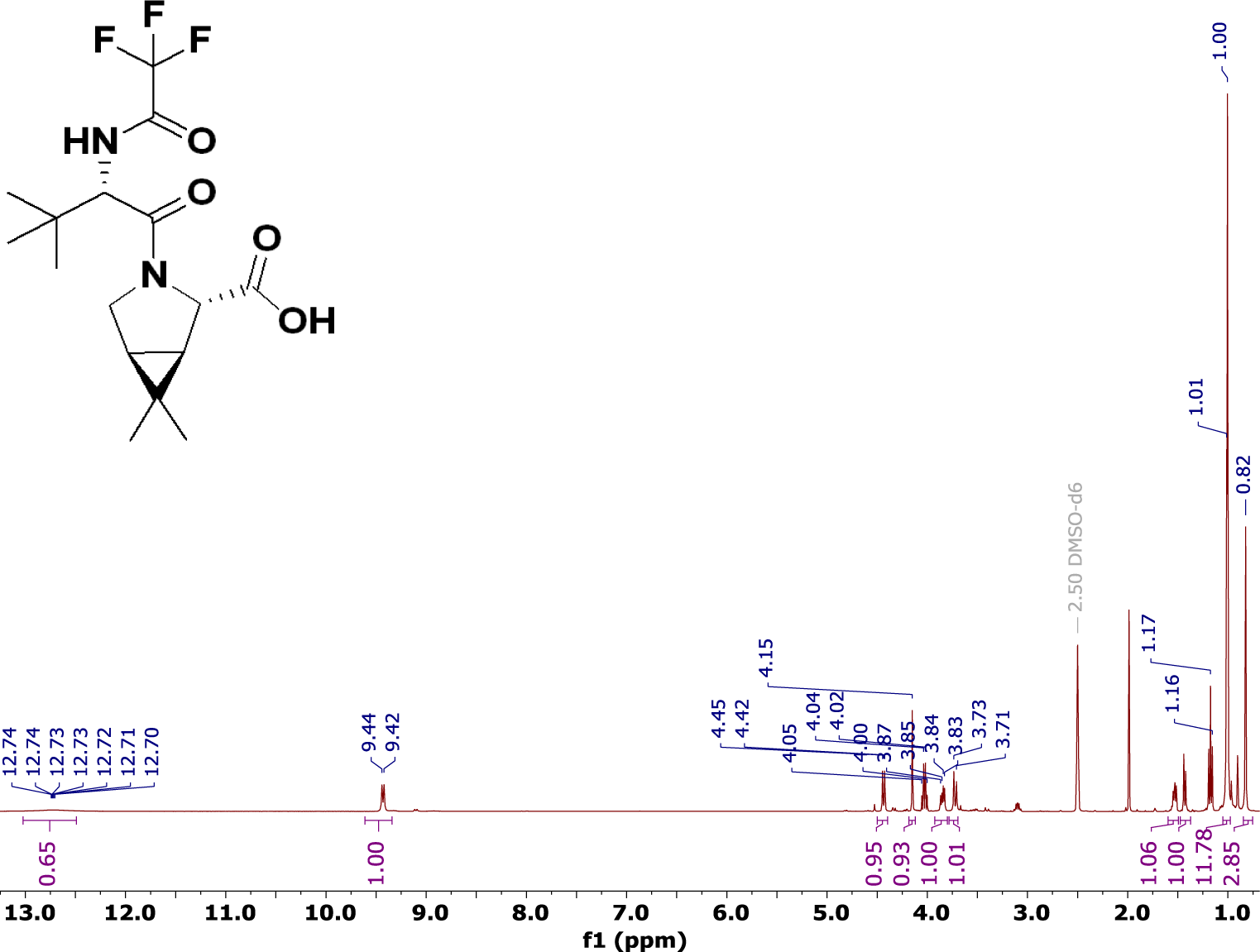

^1^H Spectrum of PF-07321332 (**6**), MTBE solvate in DMSO-*d*6 at 27 °C.

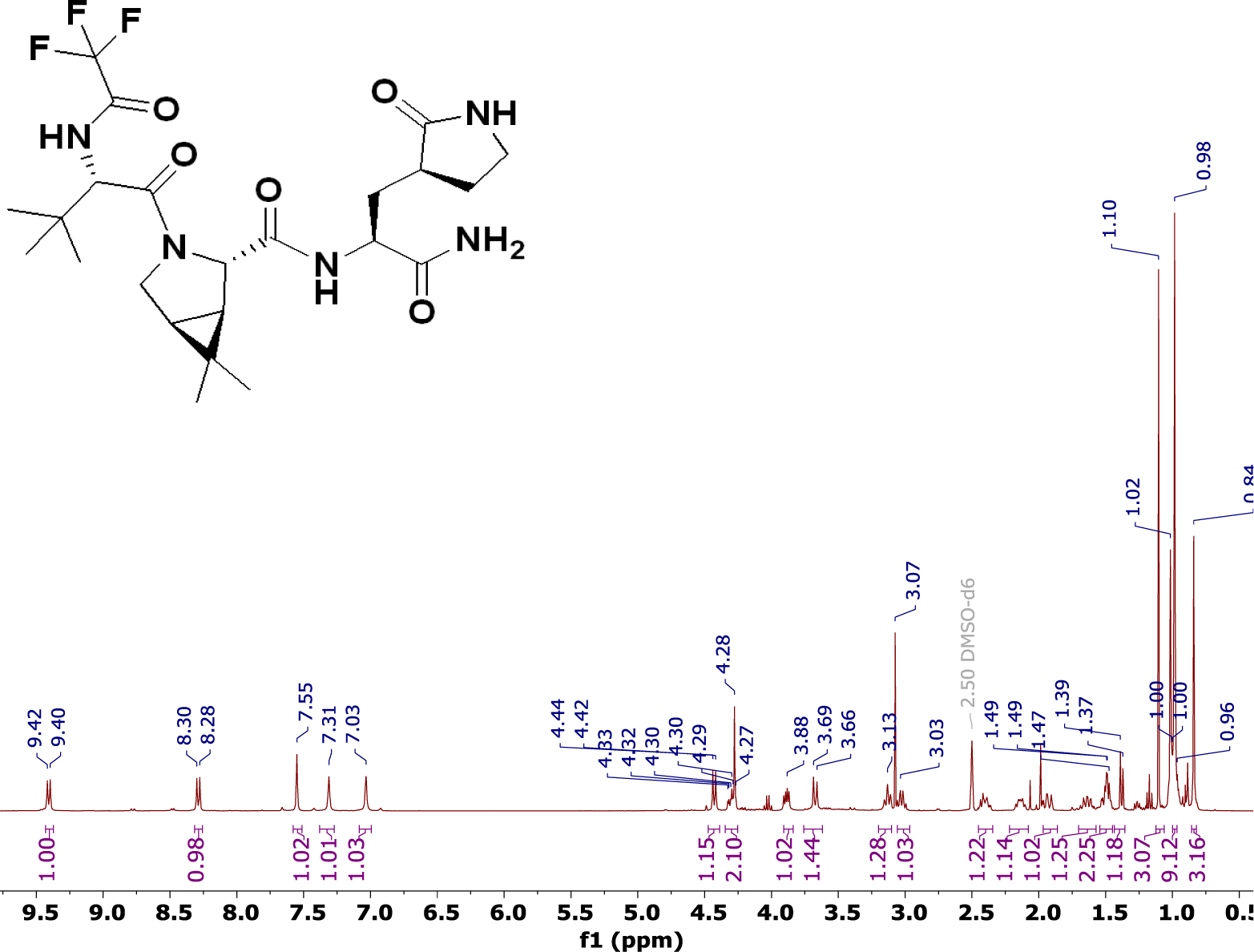

^1^H, ^13^C, ^19^F Resonance Assignments of PF-07321332 (**6**) in DMSO-*d*6 at 27°C *

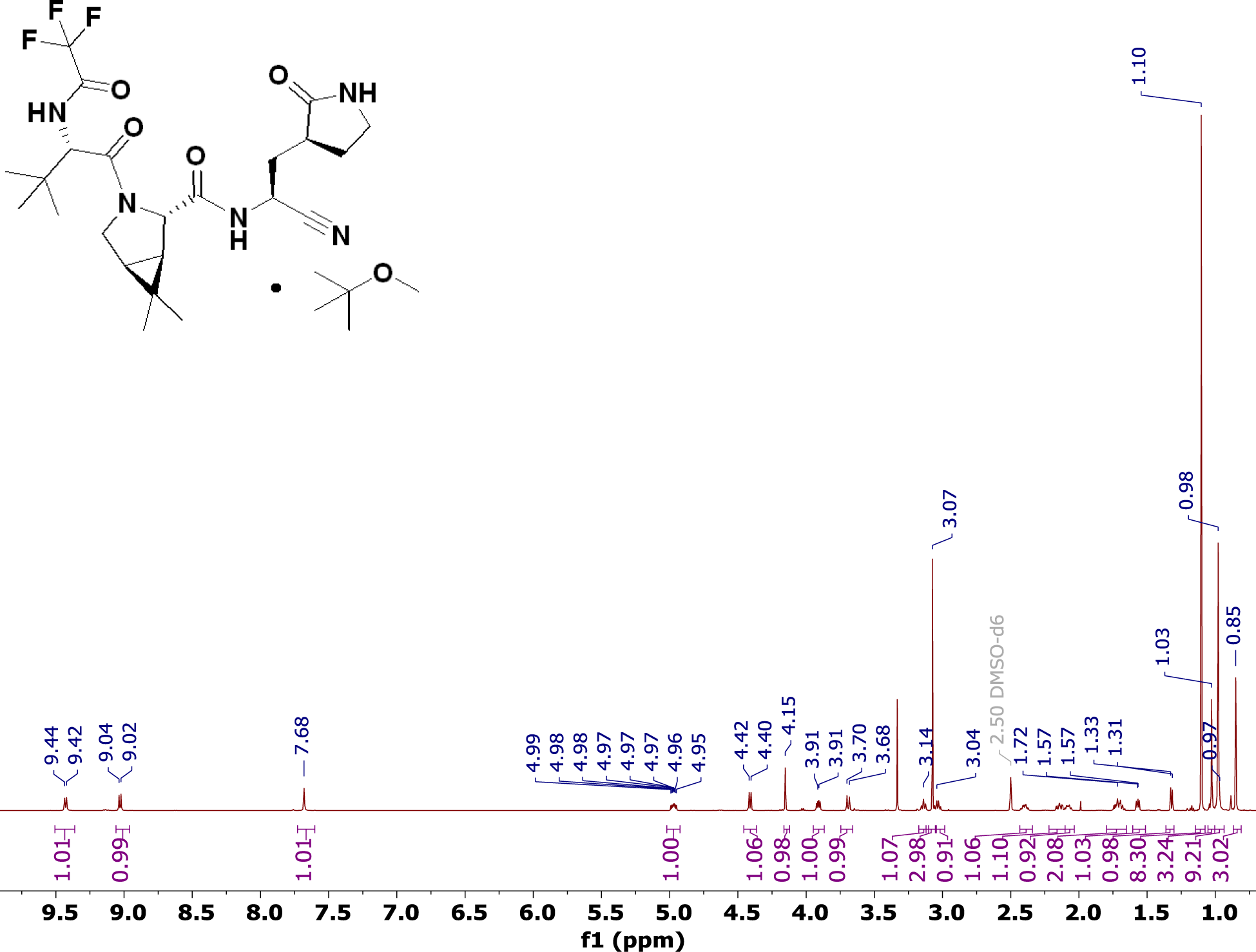

MHz for ^13^C. The ^19^F spectra are referenced using a trifluoroacetic acid standard (-76.6 ppm) at 376 MHz.

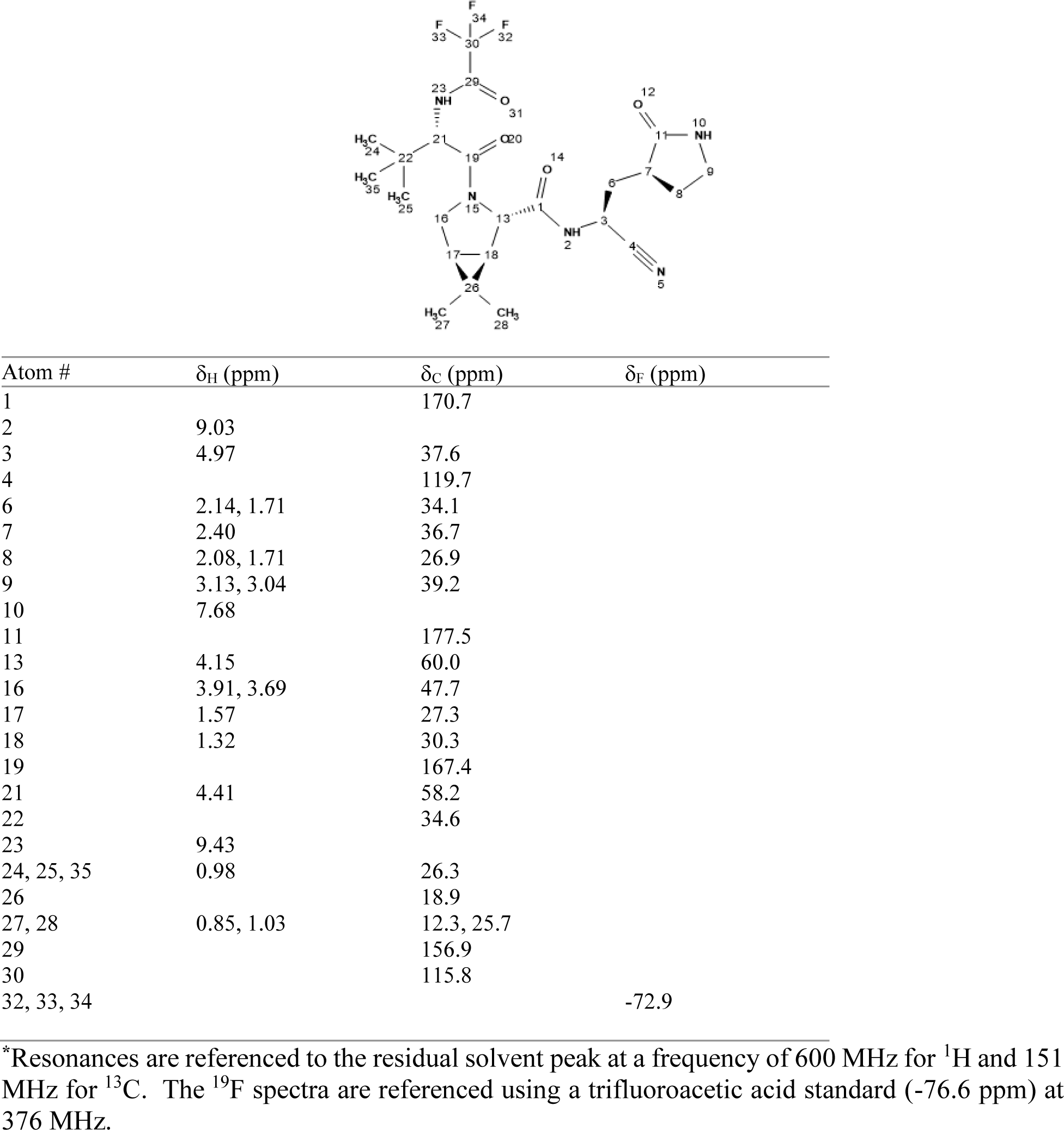

Assigned ^1^H Spectrum of PF-07321332 (Compound **6**) in DMSO-*d*6 at 27 °C.

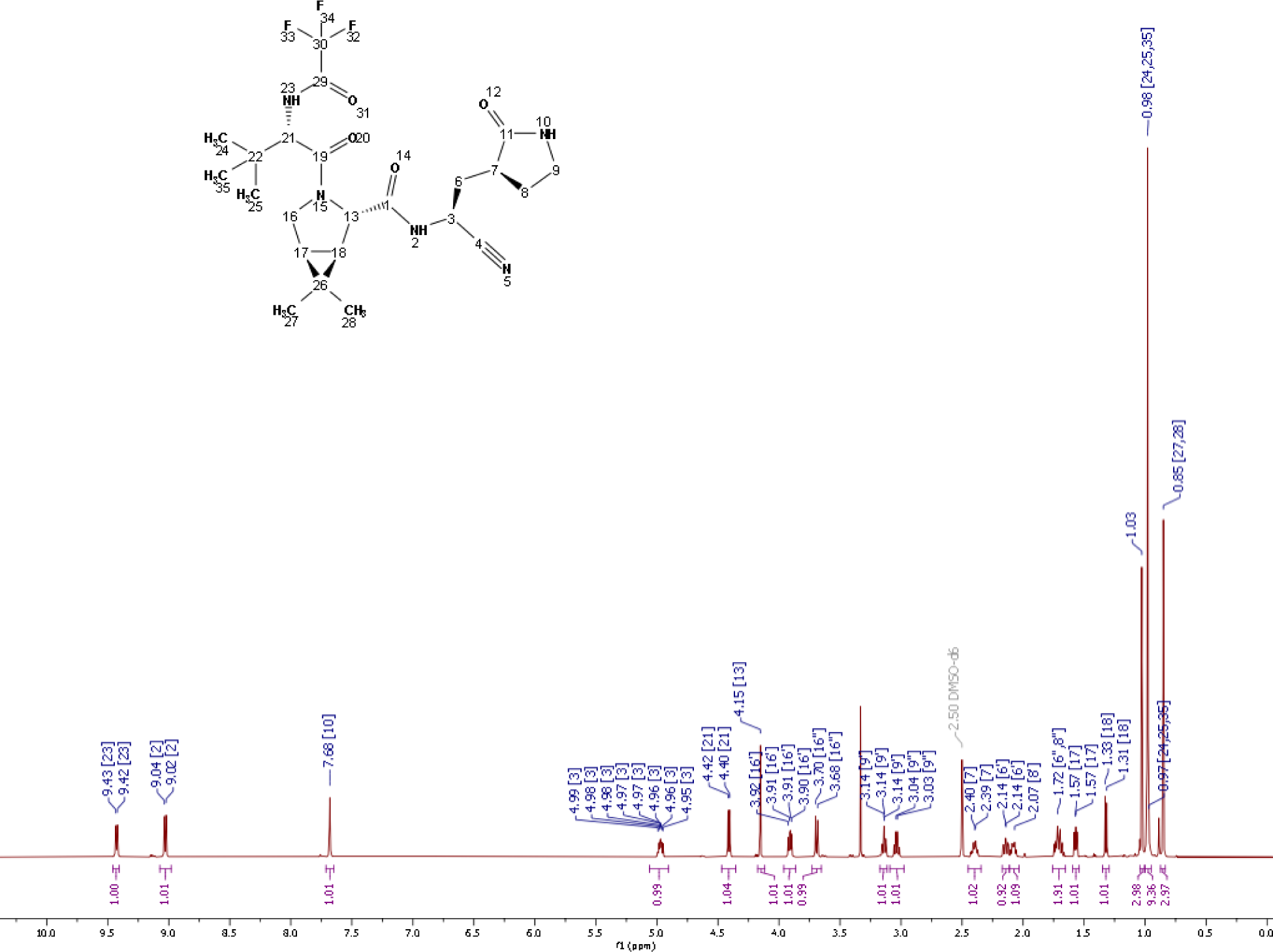

Assigned ^13^C Spectrum of PF-07321332 (Compound **6**) in DMSO-*d*6 at 27 °C.

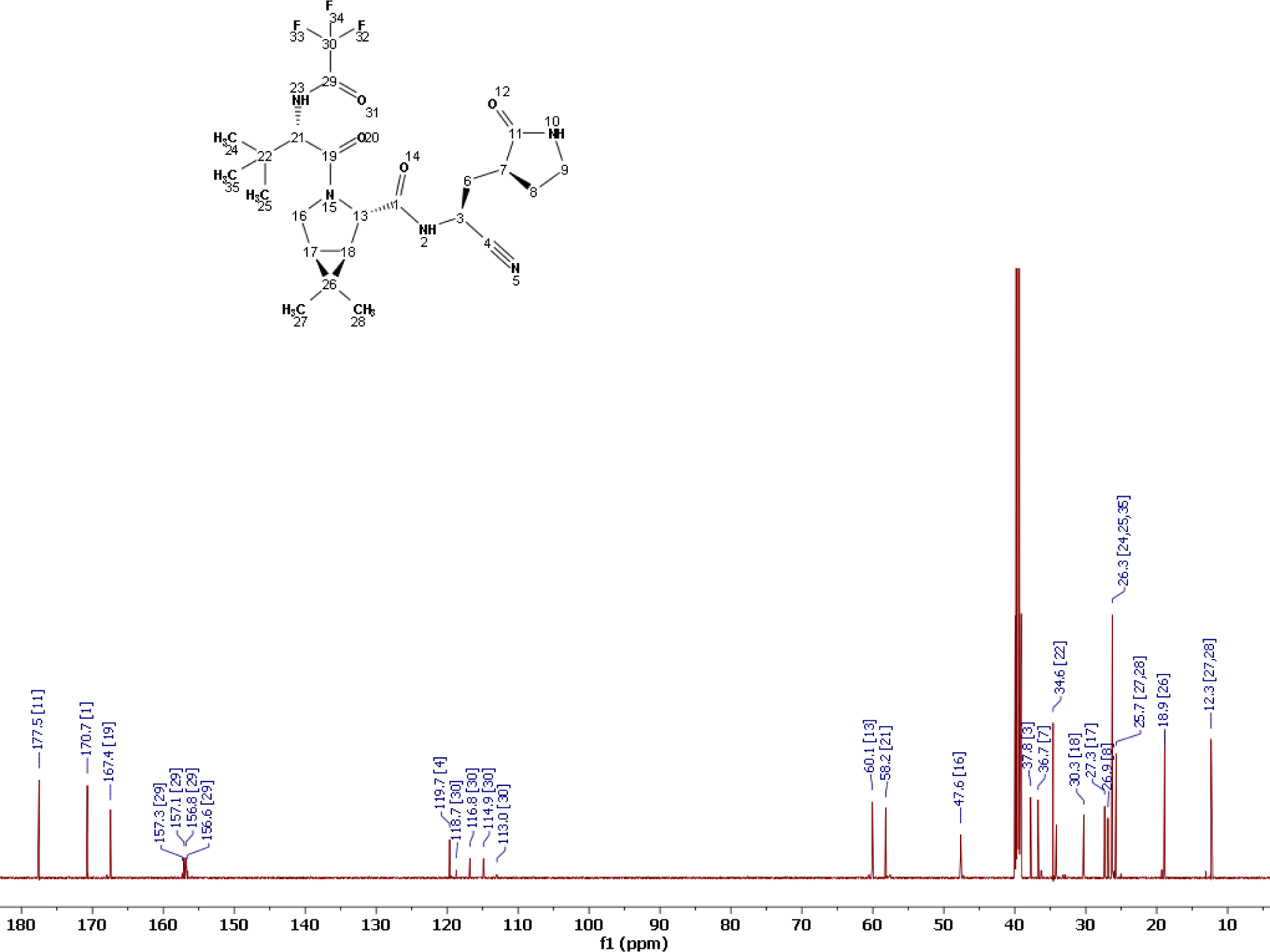

Assigned ^19^F Spectrum of PF-07321332 (Compound **6**) in DMSO-*d*6 at 27 °C.

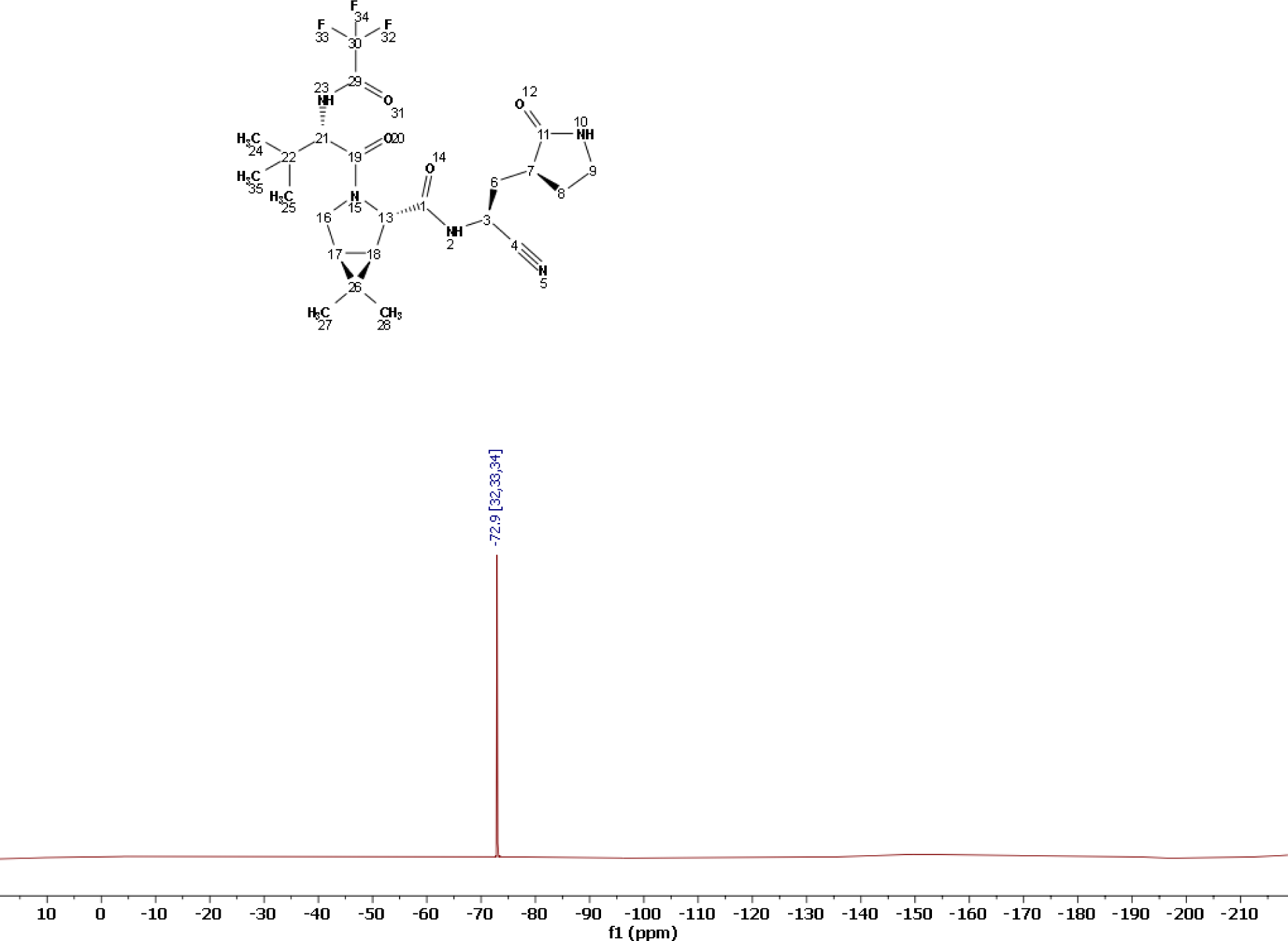

1D Rotating Frame Overhauser Effect (ROE) Spectrum of PF-07321332 (**6**)

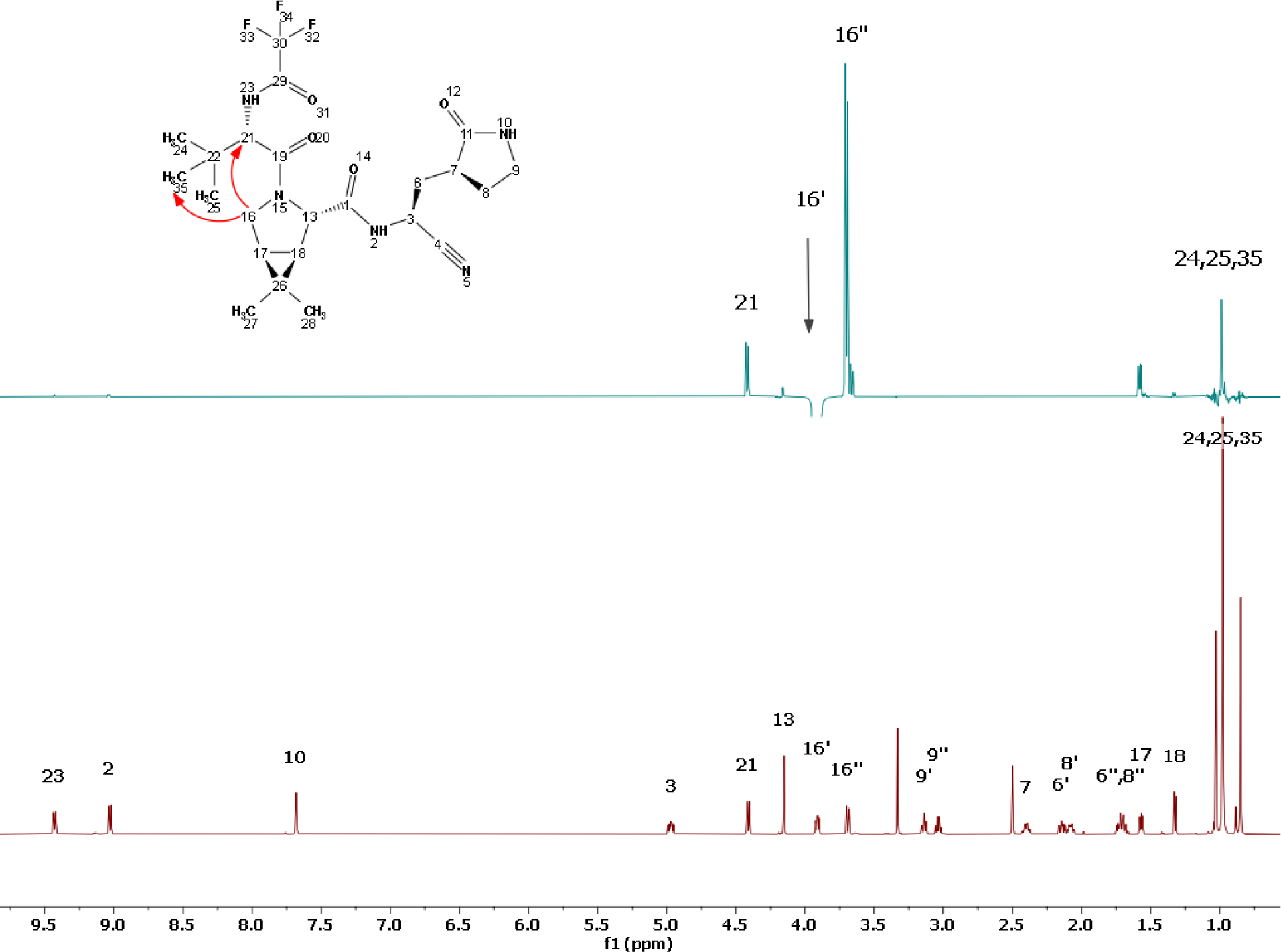

Top panel: PF-07321332 (**6**) in DMSO-*d*6 with irradiation of 16’-CH (black arrow). Through space correlations (red arrows) are observed to 21-CH and 24,25,35 CH3 consistent with the *syn* rotamer being the major conformation in solution. Bottom panel: Assigned ^1^H Spectrum of PF-07321332 (**6**) in dimethyl sulfoxide- *d*6 for comparison.

#### High Resolution Mass Spectra for compounds **2-6**

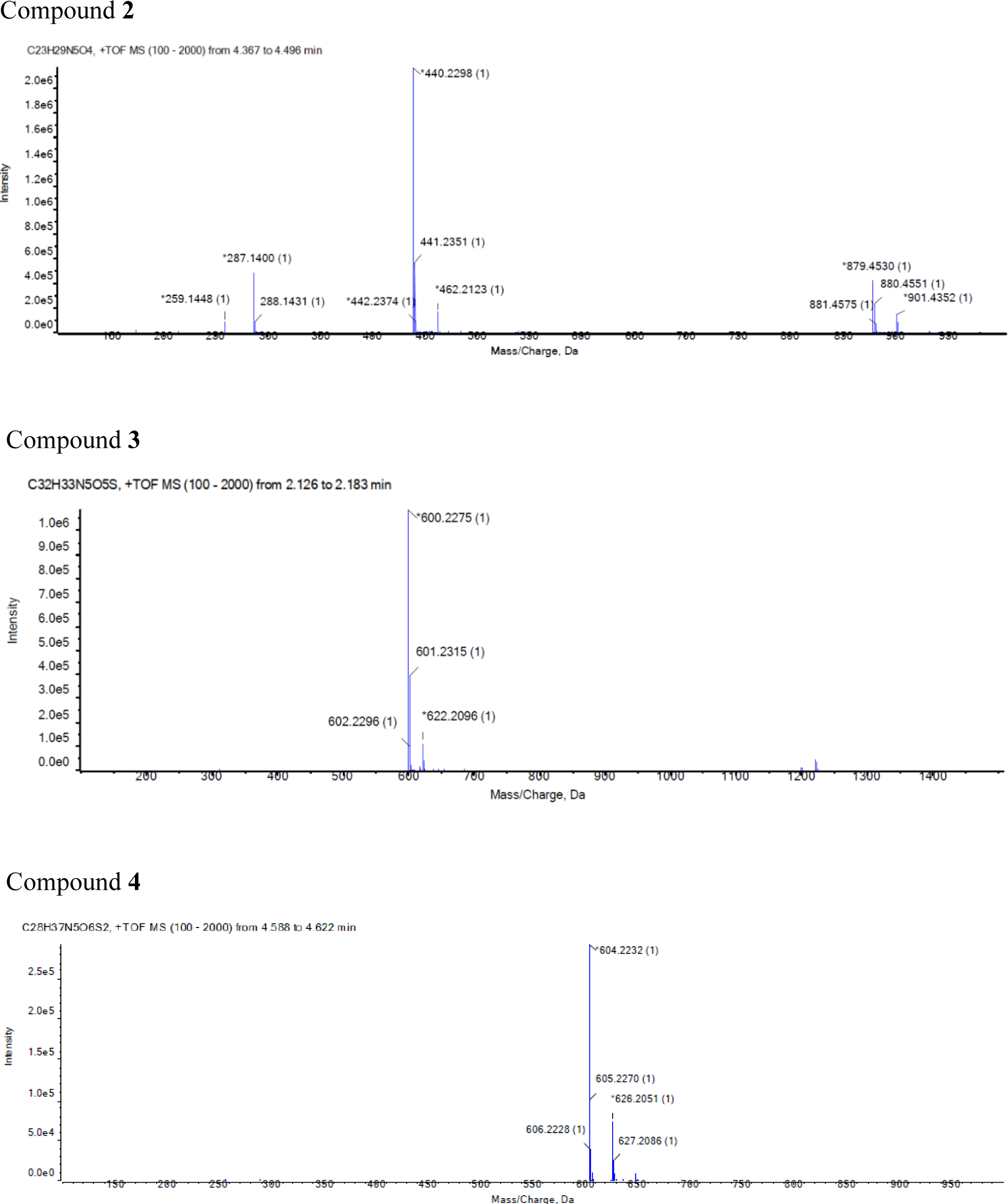
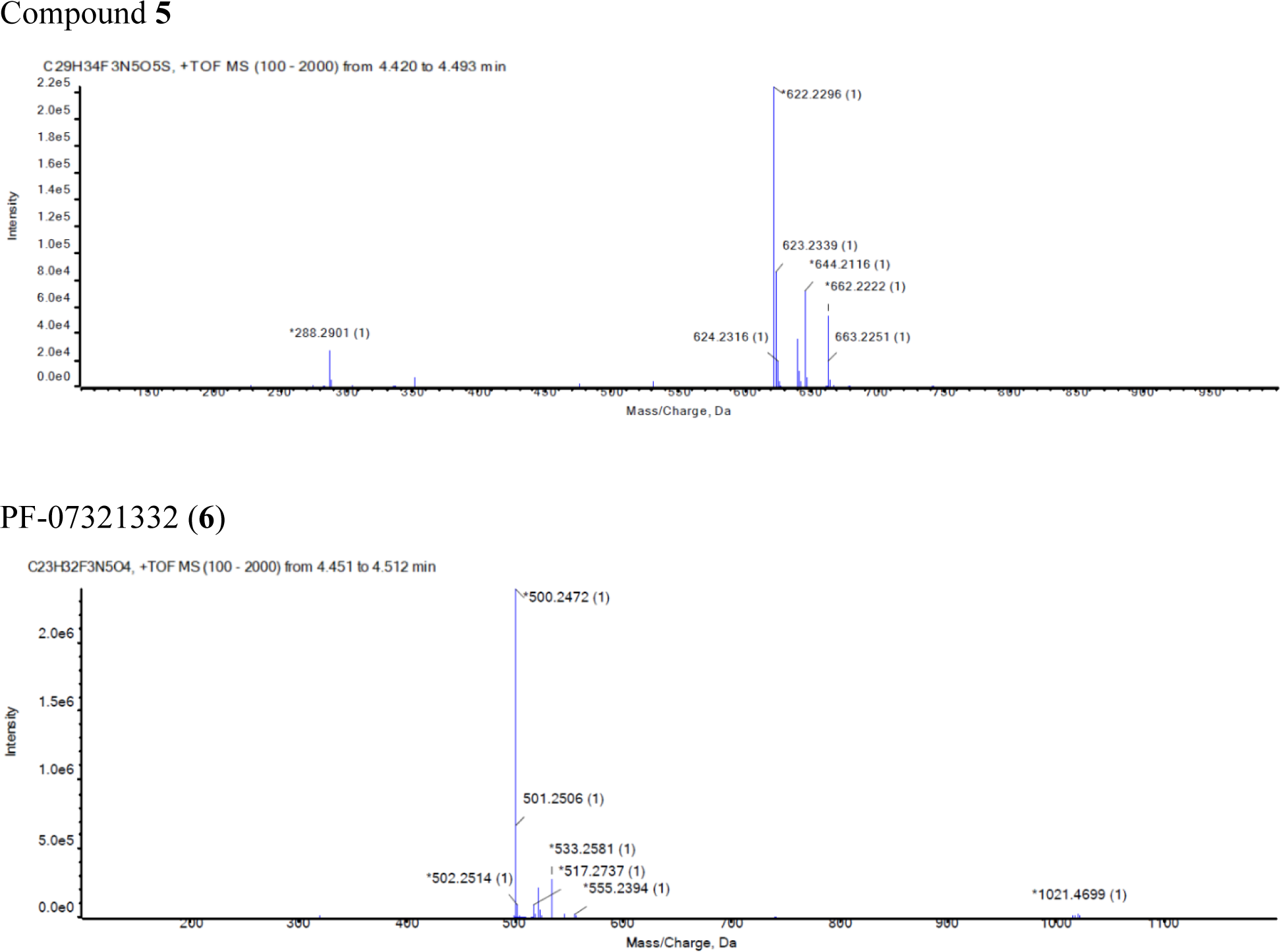

#### Powder X-Ray Diffraction Pattern of PF-07321332 (**6**)

Powder X-ray diffraction analysis was conducted using a Bruker AXS D8 Endeavor diffractometer equipped with a Cu radiation source (K-α average). The divergence slit was set at 15 mm continuous illumination. Diffracted radiation was detected by a PSD-Lynx Eye detector, with the detector PSD opening set at 2.99 degrees. The X-ray tube voltage and amperage were set to 40 kV and 40 mA respectively. Data was collected in the Theta-Theta goniometer at the Cu wavelength from 3.0 to 40.0 degrees 2-Theta using a step size of 0.00998 degrees and a step time of 1.0 second. The antiscatter screen was set to a fixed distance of 1.5 mm. Samples were rotated at 15/minute during collection. Samples were prepared by placing them in a silicon low-background sample holder and rotated during collection. Data were collected using Bruker DIFFRAC Plus software and analysis was performed by EVA DIFFRAC Plus software. Using the peak search algorithm in the EVA software, peaks selected with a threshold value of 1 were used to make preliminary peak assignments. To ensure validity, adjustments were manually made; the output of automated assignments was visually checked, and peak positions were adjusted to the peak maximum. Peaks with relative intensity of ≥ 3% were generally chosen. The peaks which were not resolved or were consistent with noise were not selected. A typical error associated with the peak position from PXRD stated in USP is up to +/- 0.2° 2- Theta (USP-941).

Powder X-ray diffraction and selected peaks for PF-07321332 (**6**), anhydrous MTBE solvate.

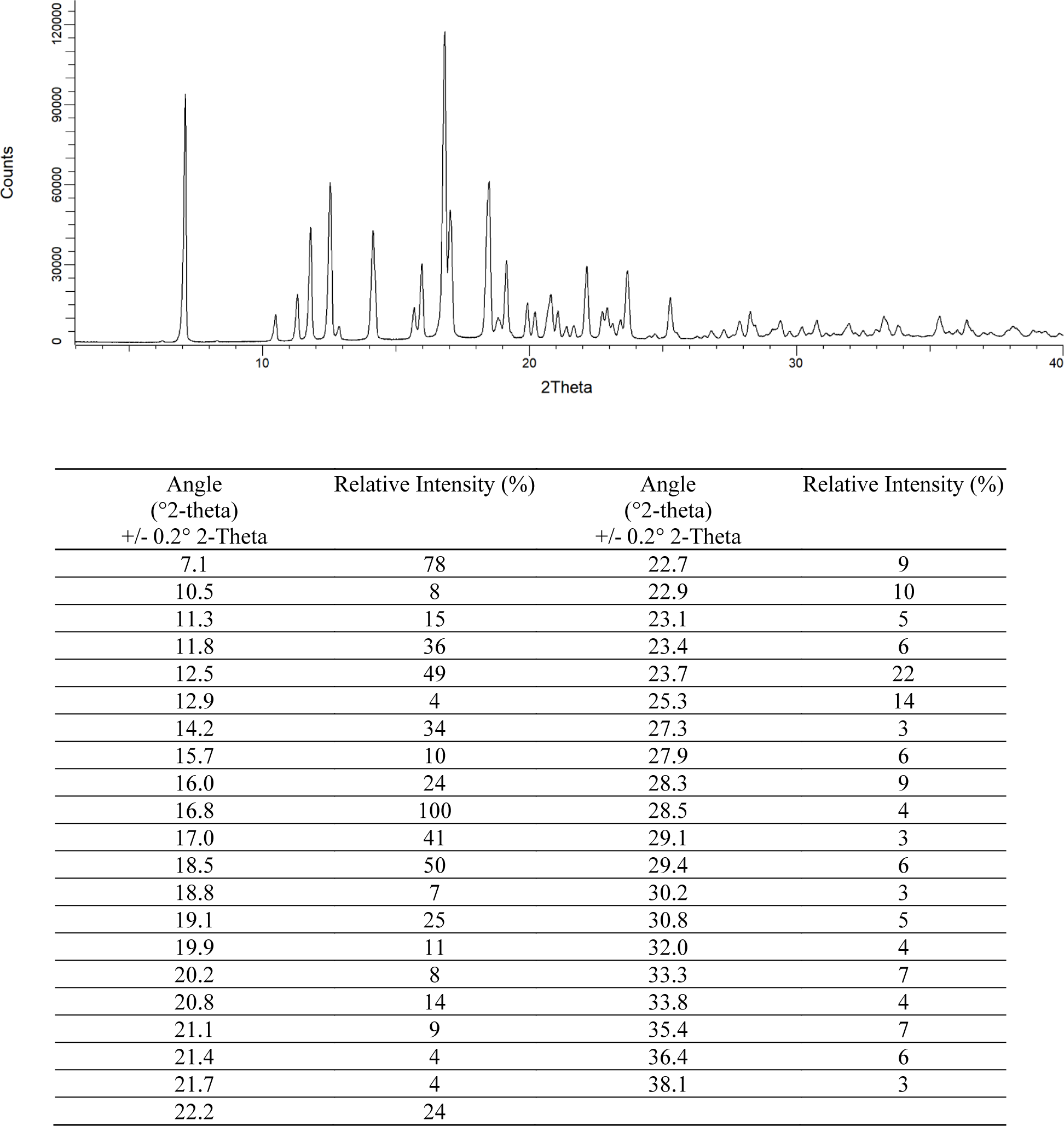

Powder X-ray diffraction and selected peaks for PF-07321332 (**6**), anhydrous ’Form 1’

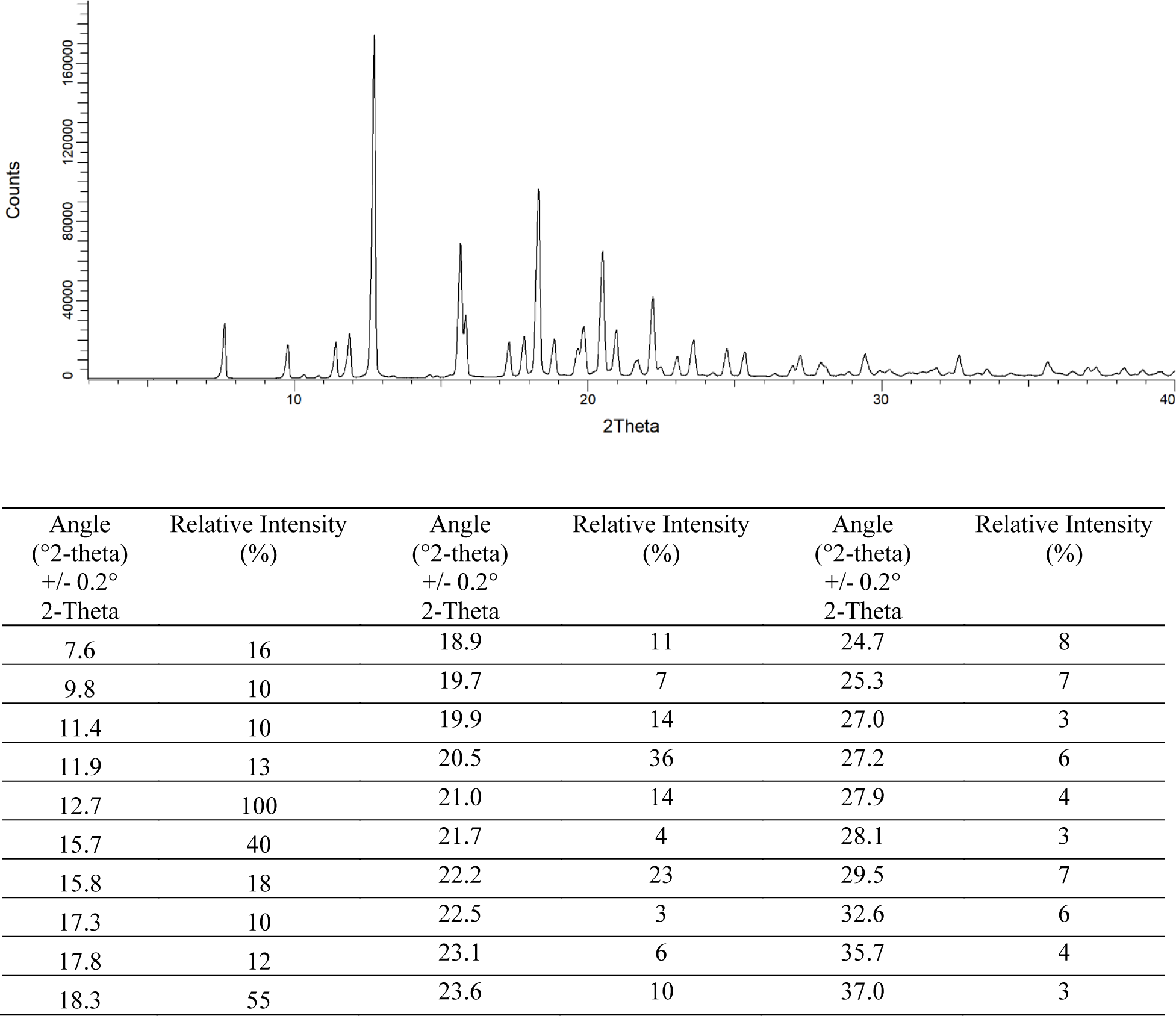

#### Recombinant SARS-CoV-2 M^pro^ Protease Production

Optimized synthetic genes coding for an *Escherichia coli (E. Coli)* expression M^pro^ protease enzyme from the SARS-CoV-2 virus (Wuhan-Hu-1 isolate; accession number MN908947) were designed and ordered from Genscript and IDT/BATJ. Two *E. Coli* expression constructs were prepared – SARS-CoV-2 M^pro^ protease (fully mature, authentic) and SARS-CoV-2 M^pro^+G (SARS-CoV-2 M^pro^ protease with an additional Glycine at its N-terminus). The SARS-CoV-2 M^pro^ construct contains both N and C-terminal His tags. The N-terminal hexa-histidine tag followed with TEV cleavage-site (TTENLYFQ↓SGFRK, arrow indicates the cleavage site), was autocleaved by SARS-CoV-2 M^pro^ protease during expression to generate the mature N- terminus At the C-terminus, the construct contained a GP hexa-histidine affinity tag, SGVTFQ↓ GP, which is a modified PreScission cleavage site that was removed during the purification with PreScission protease. The SARS-CoV-2 M^pro^+G construct contains an N-terminal hexa-histidine affinity tag with TEV cleavage-site (TTENLYFQ↓GSGFRK), which was removed during purification with TEV, leaving an extra glycine at the N-terminus *E. Coli* BL21(DE3) cells harboring the SARS-CoV-2 M^pro^ expression vector were grown in multiples of 500 ml of LB in 1 liter shake flasks for 5 hours post induction at 16 °C. *E. Coli* BL21(DE3) cells harboring the SARS-CoV-2 M^pro^+G expression vector were grown in 6 L of Terrific Broth for 5 h post induction at 30 °C in a high density shake flask. Cell pellets were stored at -80 °C until purification.

#### Purification of SARS-CoV-2 M^pro^

Cell pellets were resuspended and lysed in 50 mM tris(hydroxymethyl)aminomethane (Tris) pH 8, 250 mM NaCl, 10 mM imidazole, 0.25 mM TCEP (Buffer A) via microfluidization and clarified by centrifugation at 29,400 g for 60 min at 4 °C. Cleared lysate was added to Ni- probond resin and incubated at 4 °C for 2 h. Ni-resin was loaded on a gravity column and after 15 column volumes washes in Buffer A, protein was eluted in 50 mM Tris pH 8, 250 mM NaCl, 200 mM imidazole, 0.25 mM TCEP. Eluted protein was incubated with TEV/Precision protease and dialyzed overnight. The dialyzed and tag-removed protein was filtered and run over 5 ml nickel column to remove the affinity tag, and the flow through was further purified loading on a Superdex-200 26/60 column equilibrated with 25 mM Tris pH 7.5, 150 mM NaCl, 1 mM EDTA, 1 mM DTT. Pooled fractions were concentrated to 7.10 mg/ml and aliquots were flash-frozen in liquid nitrogen and stored at -80 °C until crystallization.

#### Purification of SARS-CoV-2 M^pro^+G

Cells were lysed via microfluidization and clarified by centrifugation at 38,400 g for 60 min at 4 °C. Lysate was loaded onto a 5 ml HisTrap HP column in 50 mM Tris (pH 8.0), 500 mM NaCl, 20 mM imidazole, pH 8.0, 14 mM β-mercaptoethanol (β−ME) (buffer B). The column was washed with buffer B before eluting with 20 column volumes of 20-300 mM imidazole (0-75% buffer C: 50 mM Tris-HCl, pH 8.0, 500 mM NaCl, 400 mM Imidazole, pH 8.0, 14 mM β−ME) and 10 column volumes 400 mM imidazole (100% buffer C). The nickel eluate was incubated with TEV protease to cleave the histidine tag and dialyzed overnight at 4 °C in 50 mM Tris-HCl (pH 8.0), 200 mM NaCl, 14 mM β−ME. The dialyzed and tag-removed protein was filtered and run over 5 ml nickel column to remove the affinity tag, and the flow through was loaded onto a 2 x 53 ml HiPrep Desalting column in 25 mM Tris-HCl (pH 8.0), 50 mM NaCl, 14 mM β−ME (Q buffer A). Pooled buffer-exchanged fractions were loaded onto a 10 ml Q column. Q flow-through fractions containing the SARS-CoV2 M^pro^ enzyme were concentrated and loaded onto a 320 ml Superdex-200 gel filtration column in 25 mM Tris (pH 7.5), 150 mM NaCl, 1 mM ethylenediaminetetraacetic acid (EDTA), and 1 mM DTT. Pooled fractions were concentrated to 11.77 mg/ml and filtered through a 0.2 μM filter. Aliquots were flash-frozen in liquid nitrogen and stored at -80 °C until crystallization.

#### Co-crystallization of SARS-CoV-2 M^pro^ with Compound **3**

SARS-CoV-2 M^pro^ protein (7.10 mg/ml) was incubated with 1.5 mM compound **3** (in 100% DMSO) for a molar ratio of approximately 1:7, for 18 h at 4 °C. The complex was then passed through a 0.45 μM cellulose-acetate spin filter and set up for crystallization using an NT-8 crystallization robot (Formulatrix). Using MRC-2 crystallization plates, wells containing 40 μl of 0.1 M MES pH 6.00, 20.0 %w/v PEG 6000, and 0.2 M sodium chloride were dispensed, and then sitting drops consisting of 0.3 μl protein were set up against 0.3 μl well buffer. Crystallization plates were incubated at 20 °C, and crystals measuring 0.05 x 0.25 x 0.35 mm grew overnight. Crystals were flash cooled in liquid nitrogen after being passed through a cryoprotectant consisting of well buffer containing 20% ethylene glycol Co-crystallization of SARS-CoV-2 M^pro^ with Compound **4** SARS-CoV-2 M^pro^ protein (7.10 mg/ml) was incubated with 1.0 mM compound **4** (in 100% DMSO) for a molar ratio of approximately 1:5, for 18 h at 4 °C. The complex was then passed through a 0.45 μM cellulose-acetate spin filter and set up for crystallization using an NT-8 crystallization robot (Formulatrix). Using MRC-2 crystallization plates, wells containing 40 μl of 20.0 %w/v PEG 6000, 0.1 M HEPES at pH 7.00, and 0.2 M sodium chloride were dispensed, and then sitting drops consisting of 0.3 μl protein were set up against 0.3 μl well buffer. Crystallization plates were incubated at 2 °C, and rod-shaped crystals measuring 0.3 x 0.02 x 0.02 mm grew overnight. Crystals were flash cooled in liquid nitrogen after being passed through a cryoprotectant consisting of well buffer containing 20% ethylene glycol

#### Co-crystallization of SARS-CoV-2 M^pro^+G with PF-07321332 (**6**)

SARS-CoV-2 M^pro^+G protein (11.77 mg/ml) was incubated with a three-fold molar excess of PF-07321332 (**6**) for 24 hours at 4 °C. The complex was then passed through a 0.45 μM cellulose-acetate spin filter and set up for crystallization using an NT-8 crystallization robot (Formulatrix). Using MRC-2 crystallization plates, wells containing 40 μl of 25.0 %w/v (25.0 μl of stock 50.0 %w/v) PEG 1500, 0.1 M (5.0 μl of stock 1.0 M) DL-Malic acid/MES monohydrate/Tris buffer at pH 6.00 were dispensed, and then sitting drops consisting of 0.3 μl protein and 0.3 μl well buffer were set up. Crystallization plates were incubated at 21°C, and very small but well-formed needle-like crystals grew in 3 days. Crystals were flash cooled in liquid nitrogen after being passed through a cryoprotectant consisting of well buffer containing 20% ethylene glycol.

#### Crystals of SARS-CoV-2 M^pro^ in complex with PF-07321332 (**6**)

Crystals of SARS-CoV-2 M^pro^ protein in complex with PF-07321332 were prepared by soaking compound into SARS-CoV-2 M^pro^ apo crystals prepared as follows: SARS-CoV-2 M^pro^ at 7.1 mg/ml was passed through a 0.45 μM cellulose-acetate spin filter and set up for apo crystallization using an NT-8 crystallization robot (Formulatrix). Using MRC-2 crystallization plates, wells containing 40 μl of 25.0 %w/v PEG 1500 and 0.1 M MIB (sodium malonate, imidazole, and boric acid) at pH 6.00 were dispensed, and then sitting drops consisting of 0.3 μl protein were set up against 0.3 μl well buffer. Crystallization plates were incubated at 13 °C, and mosaic clusters of crystals grew in about 3 days. PF-07321332 (1 μl of 100 mM stock solution in 100% DMSO) was added to the 40 μl well containing PEG 1500 and MIB, mixed, and then 0.3 μl of the solution was added to the crystallization drop (approximately 1 mM final concentration). The crystals were allowed to soak in the compound solution, undisturbed, at 13 °C for three days. Soaked crystals were teased apart from the crystal clusters and then flash cooled in liquid nitrogen after being passed through a cryoprotectant consisting of well buffer containing 20% ethylene glycol.

#### Diffraction Data, Structure Determination and Refinement

X-ray diffraction data sets were collected at IMCA-CAT 17-ID beamline (*36, 37*) of the Advanced Photon Source (APS) at Argonne National Labs (*38*) using the Eiger 2 x 9M detector (Dectris) and processed using autoPROC from Global Phasing (*39*). Anisotropic datasets used for structure determination and refinement were scaled with STARANISO (*40*). Structures of SARS-CoV-2 M^pro^ with compound **4** and SARS-CoV-2 M^pro^+G with PF-07321332 (**6**) were determined by difference Fourier in BUSTER (*41*) using the apo SARS-CoV-2 M^pro^ as the input model. This search model was also used for structure determination by molecular replacement, using either Phaser (SARS-CoV-2 M^pro^ + PF-07311342) or MolRep (SARS-CoV-2 M^pro^ + PF- 07321332) in the CCP4 suite (*42–44*). In each case, the inhibitor was built into the Fo-Fc electron density using AFITT (*45, 46*). All structures were refined iteratively by manual model building in Coot (*47*) followed by refinement in BUSTER. Statistics of diffraction data processing and the model refinement are given below.

#### Diffraction Data and Refinement Statistics

| Compound | **3** | **4** | **PF-07321332 (6)** | **PF-07321332 (6)** |
| --- | --- | --- | --- | --- |
| Protein | SARS-CoV-2 M<sup>pro</sup> | SARS-CoV-2 M<sup>pro</sup> | SARS-CoV-2 M<sup>pro</sup> | SARS-CoV-2 M<sup>pro</sup>+G |
| PDB entry ID | 7RFR | 7RFU | 7RFS | 7RFW |
| Data Statistics |  |  |  |  |
| Wavelength (Å) | 1.0 | 1.0 | 1.0 | 1.0 |
| Resolution | 72.1 – 1.63 | 55.6 – 2.50 | 55.74 – 1.91 | 55.93 – 1.73 |
| Space group | P2<sub>1</sub>2<sub>1</sub>2<sub>1</sub> | C2 | C2 | C2 |
| Unit cell dimensions [Å] | a = 67.8, b = 100.6, c = 103.4, | a = 113.8, b = 53.5, c = 45.9 | a = 114.2, b = 53.0, c = 45.7 | a = 114.6, b = 53.1, c = 45.5 |
| Unit cell dimensions [°] | $\alpha = \beta = \gamma = 90.0$ | $\alpha = \gamma = 90.0$,<br>$\beta = 102.2$ | $\alpha = \gamma = 90.0$,<br>$\beta = 102.5$ | $\alpha = \gamma = 90.0$,<br>$\beta = 102.6$ |
| Total number of reflections<sup>a</sup> | 391982 (19580) | 21559 (1032) | 46602 (1853) | 65390 (2568) |
| Unique reflections<sup>a</sup> | 59000 (2951) | 6299 (316) | 13776 (689) | 19549 (978) |
| Multiplicity<sup>a</sup> | 6.6 (6.6) | 3.4 (3.3) | 3.4 (2.7) | 3.3 (2.6) |
| Completeness (%), spherical<sup>a</sup> | 65.8 (19.2) | 66.2 (14.5) | 66.1 (12.3) | 69.8 (16.1) |
| Completeness<br>(%), ellipsoidal<sup>a</sup> | 90.7 (77.4) | 84.7 (39.7) | 75.2 (21.0) | 79.1 (33.2) |
| Mean $I/\sigma(I)$<sup>a</sup> | 12.5 (1.7) | 9.7 (1.0) | 10.7 (1.2) | 10.5 (1.3) |
| $R_{\text{merge}}$<sup>b</sup> | 0.099 (1.234) | 0.076 (1.173) | 0.071 (0.895) | 0.072 (0.804) |
| $R_{\text{pim}}$<sup>c</sup> | 0.042 (0.517) | 0.048 (0.756) | 0.046 (0.642) | 0.046 (0.597) |
| $CC_{1/2d}$ | 0.998 (0.564) | 0.998 (0.246) | 0.997 (0.493) | 0.998 (0.426) |
| **Refinement Statistics** |  |  |  |  |
| Reflections used | 59000 | 6299 | 13776 | 19549 |
| Reflections used<br>for $R_{\text{free}}$ | 3022 | 308 | 711 | 989 |
| $R_{\text{cryst}}$<sup>e</sup> | 0.194 | 0.207 | 0.215 | 0.195 |
| $R_{\text{free}}$ | 0.228 | 0.276 | 0.260 | 0.227 |
| **Ramachandran**<br>**Plot** |  |  |  |  |
| Favoured regions<br>(%) | 98.0 | 93.9 | 97 | 99.0 |
| Allowed Regions<br>(%) | 1.8 | 5.7 | 2.6 | 0.7 |
| Outlier regions<br>(%) | 0.2 | 0.3 | 0.3 | 0.3 |

<sup>a</sup> Numbers in parentheses refer to the highest resolution shell
<sup>d</sup> $CC_{1/2}$ = as defined by Karplus and Diederichs (49)
<sup>e</sup> $R_{\text{cryst}} = \sum_{hkl} |F_o - F_c| / \sum_{hkl} |F_o|$ , where $F_o$ and $F_c$ are the observed and calculated structure factors, respectively. $R_{\text{free}}$ is the same as $R_{\text{cryst}}$ , but for 5% of the data randomly omitted from refinement (50).

#### Enzyme Kinetics

Test compounds and SARS-CoV-2 M^pro^ enzyme were incubated at a 1:1 molar ratio of 2 μM compound and 2 μM enzyme in assay buffer (20 mM Tris-HCl, pH 7.3, 100 mM NaCl, 1 mM EDTA, 5 mM TCEP and 0.1% BSA) for 20 minutes. The mixture was then diluted 50-fold into assay buffer followed by a transfer of 5 μl to wells of a black low volume 384-well assay plate. Enzyme activity was monitored on a BMG Pherastar at Ex/Em of 340 nm/460 nm after addition of 5 μl 60 μM peptide substrate (Dabcyl-KTSAVLQ**‖**SGFRKME-Edans) (*51*). Final reaction conditions were 20 nM enzyme with 20 nM compound and 30 μM peptide substrate. Data is expressed as fraction velocity using DMSO controls with and without M^pro^ enzyme.

Generation of Assay Ready Plates for Coronavirus M^pro^ and mammalian protease assays PF-07321332 was serially diluted by half-log in 100% DMSO 11 times with a top concentration of 3 mM or serially diluted by 2-fold in 100% DMSO 11 times with a top dose of 0.1 mM. A volume of 300 nl of each dilution was spotted into a separate plate ready for enzyme and substrate additions. The top dose of compound in the assay was 30 μM or 1 μM with the final DMSO concentration at 1%.

#### Biochemical Determination for Human Coronavirus M^pro^ Assays

The respective human coronavirus M^pro^ in assay buffer (20 mM Tris-HCl, pH 7.3, 100 mM NaCl, 1 mM EDTA, 5 mM TCEP) and 0.1% BSA was added to assay-ready plates containing compound. The enzymatic reaction was then immediately initiated with the addition of 5 μl substrate in assay buffer. Final concentrations of respective protease and substrate are shown in the table below. Initial rates were measured by following the fluorescence of the cleaved substrate using a Spectramax (Molecular Devices) fluorescence plate reader in the kinetic format.

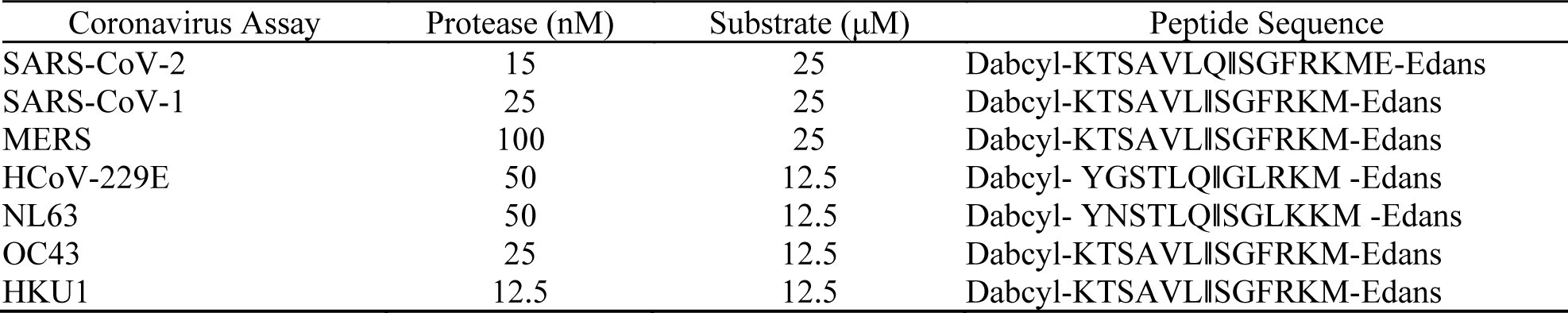

| Coronavirus Assay | Protease (nM) | Substrate (μM) | Peptide Sequence |
| --- | --- | --- | --- |
| SARS-CoV-2 | 15 | 25 | Dabcyl-KTSAVLQ$\parallel$SGFRKME-Edans |
| SARS-CoV-1 | 25 | 25 | Dabcyl-KTSAVL$\parallel$SGFRKM-Edans |
| MERS | 100 | 25 | Dabcyl-KTSAVL$\parallel$SGFRKM-Edans |
| HCoV-229E | 50 | 12.5 | Dabcyl- YGSTLQ$\parallel$GLRKM -Edans |
| NL63 | 50 | 12.5 | Dabcyl- YNSTLQ$\parallel$SGLKKM -Edans |
| OC43 | 25 | 12.5 | Dabcyl-KTSAVL$\parallel$SGFRKM-Edans |
| HKU1 | 12.5 | 12.5 | Dabcyl-KTSAVL$\parallel$SGFRKM-Edans |

PF-07321332 (**6**) was tested at final concentrations up to 30 μM in1% DMSO and compared to the broad-spectrum antiviral compound GC376 (*52*) which produced 100% inhibition at 30 μM. Control wells (0% inhibition) contained 1% DMSO with substrate and protease and did not contain compound. The reaction was allowed to progress for 60 minutes at 23 °C after which the plate was read on a Molecular Devices Spectramax M2e reader at an Ex/Em of 340 nm/490 nm.

#### Mammalian Protease Panel

The respective protease in assay buffer (50 mM Tris with 100 mM sodium chloride and Brij 35 at pH  8.0 except for cathepsin D pH  3.5 and HIV pH  5.5) was added to assay ready compound plates. The cathepsin L buffer was 400 mM sodium acetate pH 5.5 with 4 mM EDTA and 8 mM DTT. The enzymatic reaction was initiated with the addition of indicated substrate in assay buffer and proceeded at room temperature for 2 h. Final concentrations of respective protease and substrate are shown in the table below. Final DMSO concentration was below 1%. Initial rates were measured by following the fluorescence of the cleaved substrate using a Spectramax (Molecular Devices) fluorescence plate reader in the kinetic format.

| Protease/class | Enzyme (nM) | Substrate (μM) | Substrate |
| --- | --- | --- | --- |
| Caspase2/ cysteine | 10 | 5 | Ac-LEHD-AMC |
| CathepsinB/ cysteine | 1.2 | 15 | CBZArg-Arg-AMC |
| CathepsinD/ aspartyl | 1.0 | 2 | MCA-PLGL-Dap(Dnp)-AR-NH2 |
| CathepsinL/cysteine | 0.25 | 10 | Z-Phe-Arg-AMC |
| Chymotrypsin/ serine | 2.0 | 750 | Suc-AAP-AMC |
| Elastase/ serine | 0.6 | 10 | MeOSuc-AAPV-AMC |
| HIV-1/ aspartyl | 20 | 10 | Anaspec SensoLyte |
| Thrombin A/ serine | 0.01 | 10 | H-D-CHA -Ala-Arg-AMC.2AcOH |

#### Data Analysis for Mammalian and Coronavirus Protease Panels

Percent inhibition values were calculated based on control wells containing DMSO only (0% inhibition) and wells containing a control compound (100% inhibition). IC50 values were generated based on a four-parameter logistic fit model using ActivityBase software (IDBS). Percent activity values were calculated based on control wells containing no compound (100% activity) and wells containing the broad-spectrum antiviral compound GC376 (0% activity). Percent activity values were calculated based on control wells containing no compound (100% activity) and wells containing an internal Pfizer control compound (0% activity). Ki values were fit to the tight binding Morrison equation with fixed parameters for enzyme concentration, substrate concentration and the Km parameter using ActivityBase software (IDBS) indicated below

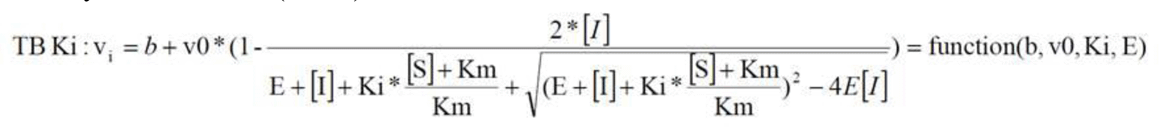

| Assay | Enzyme (nM) | Substrate (μM) | K<sub>m</sub> (μM) |
| --- | --- | --- | --- |
| 229E | 50 | 12.5 | 13.7 |
| HKU1 | 12.5 | 12.5 | 18.3 |
| MERS | 100 | 25 | 21.3 |
| NL63 | 50 | 12.5 | 13.5 |
| OC43 | 25 | 12.5 | 18.1 |
| SARS-CoV-1 | 25 | 25 | 39.3 |

#### Cellular Antiviral Activity

The ability of compounds to inhibit viral induced cytopathic effect (CPE) against human coronaviruses (SARS-CoV-1, SARS-CoV-2, hCoV-229E, MERS) was assessed by monitoring cell viability using two different assay endpoints in VeroE6, MRC-5 or Vero81 cells.

VeroE6 cells that are enriched for hACE2 expression were batched innoculated with SARS- CoV-2 (USA_WA1/2020) at a multiplicity of infection (MOI) of 0.002 in a BSL-3 lab (Southern Research Institute). Virus innoculated cells are then added to assay ready compound plates at a density of 4,000 cells/well in DMEM containing 2% heat inactivated FBS. Cells were incubated for 3 days at 37 °C with 5% CO2, a time at which virus induced CPE is 95% in the untreated, infected control conditions.

MRC-5 cells at a density of 20,000 cells/well were incubated overnight in MEM containing 5% FBS at 37 °C and 5% CO2. (Wuxi AppTech). Following addition of test compounds, HCoV- 229E virus (ATCC VR-740) (200 TCID50) was added at concentrations which correspond to a multiplicity of infection (MOI) of 0.0007 to MRC-5 cells. Cells were incubated for 3 days at 35 °C with 5% CO2.

Vero81 (ATCC CCL-81) cells were batched inoculated with MERS (EMC/2012) at M.O.I. ∼ 0.01 in a BSL-3 lab (Southern Research Institute). Virus inoculated cells were then added to assay ready compound plates at a density of 4,000 cells/well in DMEM containing 2% heat inactivated FBS. Following a 4 day incubation at 37 °C with 5% CO2, a time at which virus-induced CPE is 90 to 95% in the untreated, infected control conditions.

Cell viability was evaluated using Cell Titer-Glo (Promega), according to the manufacturer’s protocol, which quantitates ATP levels. Cytotoxicity of the compounds was assessed in parallel in assay ready compound plates with non-infected cells in a BSL-2 lab.

Test compound(s) were tested either alone or in the presence of the P-glycoprotein (P-gp) inhibitor, CP-100356 at indicated concentrations. The inclusion of CP-100356 was to assess if the test compound(s) are being effluxed out of cells due to endogenous expression of P-glycoprotein in the cell line. Percent effect at each concentration of test compound was calculated based on the values for the no virus control wells and virus containing control wells on each assay plate. The concentration required for a 50% response (EC50) value was determined from these data using a 4-parameter logistic model. EC50 curves were fit to a Hill slope of 3 when >3 and the top dose achieved ≥ 50% effect. If cytotoxicity was detected at greater than 30% effect, the corresponding concentration data was eliminated from the EC50 determination.

For cytotoxicity plates, a percent effect at each concentration of test compound was calculated based on the values for the cell only control wells and hyamine or no cell containing control wells on each assay plate. The CC50 value was calculated using a 4-parameter logistic model. A therapeutic index was then calculated by dividing the CC50 value by the EC50 value.

Antiviral activity of PF-07321332 (**6**) was evaluated in differentiated normal human bronchial epithelial (dNHBE) cells in a BSL-3 facility. The dNHBE cells (EpiAirway) were procured from MatTek Corporation (Ashland, MA) and were grown on trans-well inserts consisting of approximately 1.2 x 10^6^ cells in MatTek’s proprietary culture medium (AIR-100- MM) added to the basolateral side, with the apical side exposed to a humidified 5% CO2 environment at 37 °C. On day 1, dNHBE cells were infected with SARS-CoV-2 strain

USA-WA1/2020 at a MOI of approximately 0.0015 CCID50 per cell, and PF-07321332 treatment was carried out by inclusion of drug dilutions in basolateral culture media. At day 3 and day 5 , virus released into the apical compartment was harvested by the addition of 0.4ml culture media. The virus titer was then quantified by infecting Vero76 cells in a standard endpoint dilution assay and virus dose that was able to infect 50% of the cell cultures (CCID50 per ml) was calculated (*53*). To determine the EC50 and EC90, the CCID50/ml values were normalized to that of no drug control as a percentage of inhibition and plotted against compound concentration in GraphPad Prism software by using four-parameter logistic regression.

#### Mouse-Adapted SARS-CoV-2 Infection and Treatment Studies

The in vivo infection studies were performed in an animal biosafety level 3 (ABSL3) facility in the AAALAC-accredited Laboratory Animal Research Center at Utah State University.

Pharmacokinetics studies were performed in an animal biosafety level 2 (ABSL2) facility. The study procedures were conducted with approval by the Institutional Animal Care and Use Committee at Utah State University. A total of 24 BALB/c mice (Charles River, 8 week old female, n=6 mice/group) were divided into 4 groups: group 1: untreated, infected control; group 2: 300 mg/kg PF-07321332 (**6**); group 3: 1000 mg/kg PF-07321332 (**6**), and group 4: untreated, uninfected control (for pharmacokinetic analysis and normal weight). Mice were anesthetized by intraperitoneal (i.p.) injection of ketamine/xylazine (50 mg/kg/5 mg/kg) and inoculated intranasally (i.n.) with 1 x 10^5^ 50% cell culture infectious dose (CCID50) of SARS-CoV-2 MA10 (90 ml/nares). The mouse-adapted MA10 virus (*30*) was provided by Professor Ralph Baric (University of North Carolina). For oral (p.o.) administration, PF-07321332 (compound **6**, MTBE solvate form) was solubilized in 0.5% methylcellulose in water, containing 2% Tween80. Mice were dosed twice daily (BID) x 4 days beginning at 4 hours post infection. Mice were weighed daily starting at day 0 until end of study to measure infection-associated weight loss. At 4 days post infection (dpi), mice were euthanized by isoflurane inhalation. The lungs were collected and placed in 1 ml PBS and stored at –80 °C for evaluation of lung virus titers or collected for histopathology as described below. For virus titer assays, serial log10 dilutions of 1.0 ml lung tissue homogenates were performed in quadruplicate on confluent monolayers of Vero 76 cells seeded in 96-well microplates. The cells were incubated at 37 °C and 5% CO2 for 6 days and then scored for cytopathic effect (CPE) using a light microscope. Virus lung titer (CCID50/ml (Log10) was calculated by linear regression using the Reed-Muench method (*53*)

#### Lung Histopathology Assessment

To assess virus-induced damage to the lungs of SARS-CoV-2 MA10-infected mice, mice were euthanized at 4 dpi and lung lobes were collected for virus titer evaluation or left lobes were fixed in 4% paraformaldehyde at 4 °C for histopathology. Fixed lung lobes were shipped to an external histology laboratory (Histowiz, Inc (study 2) or M.D. Anderson (study 1)) for processing and blinded evaluation by an experienced veterinary pathologist and yielded similar results. Group samples (n=6) from study 2 were processed as one H&E-stained slide from each lung specimen. Each lung sample was evaluated using a semi-quantitative analysis using four parameters: perivascular inflammation, bronchial or bronchioloar epithelial degeneration or necrosis, bronchial or bronchiolar inflammation, and alveolar inflammation. A 5-point scoring system for assessment of epithelial degeneration/necrosis and inflammation was utilized (0-with normal limits; 1-mild; scattered cell necrosis/vacuolation, few/scattered inflammatory cells, 2- moderate; multifocal vacuolation or sloughed/necrotic cells, thin layer of inflammatory cells, 3- marked; multifocal/segmental necrosis, epithelial loss/effacement, thick layer of inflammatory cells, 4-severe; coalescing areas of necrosis; parenchymal effacement, confluent areas of inflammation. A total pathology score was calculated for each mouse by adding the individual histopathological scores.

#### Drug Metabolism Studies and Methods

Research was conducted on human tissue acquired from a third party that has been verified as compliant with Pfizer policies, including Institutional Review Board/Independent Ethics Committee approval. Phenylmethylsulfonyl fluoride-free intestinal microsomes were acquired to preserve serine hydrolase enzymatic activity. Rat intestinal microsomes (pool of 200, male Sprague Dawley,), monkey intestinal microsomes (pool of 6, male Cynomolgus,), human intestinal microsomes (pool of 9, male and female,), and human liver microsomes (HLM) (custom pool of 50 donors, male and female), were purchased from Sekisui XenoTech (Kansas City, KS) or BioIVT (Baltimore, MD). Recombinant human CYP enzymes were purchased from Corning (Glendale, Arizona). β-Nicotinamide adenine dinucleotide phosphate, reduced form (NADPH), potassium dihydrogen phosphate (monobasic), dipotassium hydrogen phosphate (dibasic), magnesium chloride, formic acid, sodium chloride, sodium hydroxide, hydrochloric acid, ketoconazole, and DMSO were obtained from Sigma Aldrich (St. Louis, MO). Acetonitrile (HPLC grade) was purchased from Fisher Scientific (Fair Lawn, NJ). High purity dosing excipients were purchased from the following sources: ethanol (200 proof anhydrous, meets US Pharmacopeia specifications) from Decon Laboratories Inc (King of Prussia, PA), PEG 400 (BioUltra 400) from Sigma, PEG 400 NF (Super Refined) and Tween 80 HP from Croda (Edison, NJ), methylcellulose A4M (premium) from Dow Chemical (Midland, MI), hydroxypropyl-β-cyclodextrin (HPCBD, Cavitron W7 HP7 Pharma Cyclodextrin) from Ashland (Columbus, Ohio), and Capmul MCM from Abitec (Janesville, WI).

#### Metabolic Stability of Compounds **1**–**5** in HLM

Substrate stocks (30 mM) were prepared in DMSO and diluted to 100-times the incubation concentration in 50% water 50% acetonitrile, for a final organic composition of 0.489% acetonitrile and 0.00334% DMSO in the incubations. Substrate (0.1 or 1 µM) was incubated in HLM (1 mg/ml) diluted in potassium phosphate buffer (100 mM, pH 7.4) supplemented with MgCl2 (3.3 mM) and NADPH (1.3 mM) in a final volume of 300 µl. Compound **1** incubations were conducted with 2 mg/ml human liver microsomes.  Incubations were conducted at 37 °C open to ambient air.  A no-NADPH control was carried out in parallel. Incubations were conducted in triplicate.  At various time points (typically 0.25, 2, 4, 6, 10, 20, 40 and 60 min), a 20 µl aliquot of incubate was removed and quenched in 100 µl of acetonitrile containing internal standard indomethacin (50 ng/ml).  Samples were vortexed, centrifuged (5 min, 2300 x g) and clean supernatant was diluted with an equal volume of water containing 0.2% formic acid.  Samples were directly analyzed by liquid chromatography tandem mass spectrometry (LC-MS/MS).  Analyst software (Sciex, Framingham, MA) was used to measure peak areas.  Peak area ratios of analyte to internal standard were calculated.  Substrate depletion half-life (t1/2) and intrinsic clearance (CLint) were calculated using E- WorkBook v10 (ID Business Solutions, Guildford, Surrey, UK).  The natural log of peak area ratios versus time were fitted using linear regression, the slope of which (*k*) was converted to  t1/2 values, where t1/2 = -0.693/*k*. To estimate in vitro CLint in HLM, the t1/2 for substrate depletion was scaled using the following equation (*54*): 

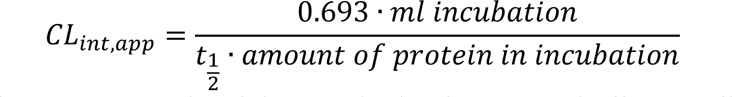

The incubation volume was 0.3 ml and the protein density was typically 1 mg liver microsomes/ml.

#### Impact of the Selective Cytochrome P450 (CYP)3A4 Inhibitor Ketoconazole on the CLint of PF- 07321332 (**6**) in HLM

Incubations were conducted in 100 mM potassium phosphate buffer (pH 7.4) containing MgCl2 (3.3 mM), NADPH (1.3 mM), HLM (1 mg/ml), and PF-07321332 (0.1 µM) at 37 °C for 0-80 min in the absence or presence of selective CYP3A inhibitor ketoconazole (1 μM). Incubations in the absence of NADPH were incubated for 0 – 60 min. The incubation volume was 0.4 ml. Reactions were terminated by transferring an aliquot (40 μl) of the incubation mixture to a solution of acetonitrile containing internal standard (25 ng/ml diclofenac , 160 μl). Quenched samples were centrifuged (2300 x g) for 5 min, followed by the transfer of supernatant (100 μl) to 96-well plates with 100 μl of H2O added to the samples. Incubations in the presence of NADPH were conducted in triplicate. Incubations in the absence of NADPH were conducted in duplicate. Samples were analyzed by LC-MS/MS for remaining PF-07321332 and CLint was estimated as described above.

#### Metabolic Stability of PF-07321332 (**6**) in Intestinal Microsomes

Stock solutions of **6** (4 mM) were prepared in MeCN and diluted to 100-times the incubation concentration in 50% water 50% acetonitrile, for a final organic composition of 0.489% acetonitrile and 0.00334% DMSO in the incubations. Compound **6** (0.1 or 1 μM) was incubated in rat, monkey, or human intestinal microsomes (1 mg/ml) diluted in potassium phosphate buffer (100 mM, pH 7.4) supplemented with MgCl2 (3.3 mM) and NADPH (1.3 mM) in a final volume of 300 µl. Incubations were conducted at 37 °C open to ambient air. A no-NADPH control was carried out in parallel. Incubations were conducted in triplicate. At various time points (typically 0, 5, 15, 30, 60, 90, and 120 min), a 30 µl aliquot of incubate was removed and quenched in 120 µl of acetonitrile containing internal standard diclofenac (25 ng/ml). Samples were vortexed, centrifuged (5 min, 2300 x g) and clean supernatant was diluted with an equal volume of water. Samples were directly analyzed for depletion of **6** by LC-MS/MS.

#### Plasma Protein Binding Determination for PF-07321332 (**6**)

On the day of each incubation, fresh blood in K2EDTA was collected from male Wistar-Hannover rats (*n*=5, pooled), male Cynomolgus monkeys (*n*=2, pooled), and humans (*n*=1 male and *n*=1 female, pooled). The blood was centrifuged for 10 min at 2500 x g and the plasma fraction was harvested. Plasma was spiked with a final concentration of 0.3, 1, 3, or 10 µM of **6** (final organic 1% DMSO). Fraction unbound in plasma (fu,p) was determined by equilibrium dialysis using an HTD 96 device assembled with 12-14k molecular weight cutoff membranes (HTDialysis, LLC, Gales Ferry, CT). Dialysis chambers were loaded with 150 µl plasma and 150 µl PBS in the donor and receiver chambers, respectively. The dialysis plate was sealed with a gas-permeable membrane and stored in a 37 °C water-jacketed incubator maintained at 75% relative humidity and 5% CO2, on a 100 rpm plate shaker. After a 6-hour incubation, samples were matrix-matched and quenched by protein precipitation, followed by LC-MS/MS analysis. A set of satellite samples was included to measure stability after a 6-hour incubation. Incubations were conducted with 12 replicates per concentration. fu,p was calculated by dividing the analyte concentration in the buffer sample by the signal in the donor sample, corrected for any dilution factors. All incubations had >70% analyte recovery and >70% stability in 6 h.

#### LC-MS/MS Analysis

LC-MS/MS analysis was performed using a Sciex Triple Quad 6500 mass spectrometer (Sciex, Framingham, MA), equipped with electrospray sources and Agilent 1290 binary pump (Santa Clara, CA). Aqueous mobile phase (A) was comprised of 0.1% formic acid in water and organic mobile phase (B) consisted of 0.1% formic acid in acetonitrile. Samples (10 µl) from various in vitro incubations were injected onto a Kinetex XB-C18 (2.1 x 50 mm, 1.7 µm) (Phenomenex, Torrance, CA) column at room temperature with a flow rate of 0.5 ml/min. The gradient program typically began with 10% initial mobile phase B held for 0.4 min, followed by a linear gradient to 60% B over 6.4 min, then to 95% over 0.2 min, held at 95% B for 0.5 min followed by re-equilibration to initial conditions for 0.5 min. The mass spectrometer was operated in multiple reaction monitoring mode, in positive detection mode, with the following mass transitions (Q1/Q3) and collision energies (CEs): Compound **1** 473/187 (CE 18), compound **2** 440/287 (CE 18), compound **3** 600/311 (CE 24), compound **4** 604/287 (CE 33), compound **5** 622/604 (CE 28), compound **6** 500/319 (CE 22), and indomethacin 358/139 (CE 27). To avoid artifacts arising from epimerization in samples from compound **5** samples, sample evaporation was avoided.

#### Metabolite Identification Studies in HLM and Recombinant Human CYP Enzymes

PF-07321332 (compound **6**, 10 μM) was incubated in pooled (mixed gender of 50) HLM (protein concentration = 2 mg/ml) and recombinant human CYP enzymes (CYP1A1, CYP1A2, CYP1B1, CYP2A6, CYP2B6, CYP2C8, CYP2C9, CYP2C18, CYP2C19, CYP2D6, CYP3A4, CYP3A5, CYP3A7, CYP2E1, CYP2J2, CYP4F2, 100 pmol/ml each) in 100 mM phosphate buffer (pH 7.4) containing NADPH (1.3 mM), MgCl2 (3.3 mM) at 37 °C. Incubations were carried out for 1 h and quenched with the addition of acetonitrile (0.6 ml). A second set of incubation mixtures were prepared where PF-07321332 was only added after quenching. These incubations served as controls to assess metabolite formation by HPLC-UV data as PF-07321332 has very poor UV absorbance and this step ensured a better delineation of PF-07321332-related material from other components in the incubation mixtures. The terminated incubation mixtures were centrifuged (1800 x g) for 5 min and the supernatants (0.6 ml) were evaporated to dryness using a Genevac evaporative centrifuge, and the residue was reconstituted in 50 μl of 1% formic acid containing 20% acetonitrile for analysis by ultra-high performance liquid chromatography-ultraviolet spectroscopy-high resolution mass spectrometry (UHPLC-UV-HRMS) analysis. Reconstituted samples were analyzed by UHPLC-UV-MS operated in positive ion mode using an Orbitrap Elite mass spectrometer in line with a Vanquish UHPLC-UV with cooled autoinjector (Thermofisher, Waltham, MA). Injection volumes were 5 to 15 μl, depending on the experimental run. A Kinetex XB C18 column (Agilent, Santa Clara, California) was used (2.1 x 100 mm, 2.6 μm) with a flow rate of 0.4 ml/min heated to 45 °C. Mobile phase A was comprised of 0.1% formic acid in water and mobile phase B was comprised of acetonitrile. The gradient system comprised of: initially, 5% B held for 0.5 min followed by a linear gradient to 70% B at 11 min, a second linear gradient to 95% B at 13 min, a 1 min wash at 95% B, and finally a 2 min re-equilibration period at 5% B. UV was monitored between 200 to 400 nm and chromatograms were reconstructed using λ at 200 nm. Mass spectral data were collected in positive ion mode.

#### Biosynthesis of PF-07321332 Metabolites for Structural Elucidation

PF-07321332 (25 μM) was incubated with human liver microsomes (2 mg/ml), MgCl2 (3.3 mM), and NADPH (1.3 mM) in a volume of 40 ml potassium phosphate buffer (100 mM, pH 7.4) at 37 °C for 55 min. The incubation mixture was quenched with acetonitrile (40 ml), centrifuged (1800 x g) for 5 min, and the solution was reduced in a Genevac vacuum centrifuge. To the remaining mixture was added formic acid (0.5 ml), acetonitrile (0.5 ml) and water up to a volume of 50 ml. This mixture was spun in a centrifuge (40000 x g) for 30 min and the supernatant was applied to a Polaris C18 column (Agilent, Santa Clara, CA) (4.6 x 250 mm; 5 μm) through a Jasco HPLC pump (Jasco Inc., Easton, MD) at 0.8 ml/min. After application, the column was moved to an Acquity HPLC-UV in line with a CTC Analytics fraction collector (Conquer Scientific, Poway, CA) and LTQ Velos mass spectrometer (Thermofisher, Waltham, MA). The material was eluted with a gradient consisting of mobile phase A (0.1% formic acid in water) and mobile phase B (acetonitrile) at 0.8 ml/min. The gradient commenced at 2% mobile phase B with a linear gradient to 15% mobile phase B at 5 min, a second linear gradient to 60% mobile phase B at 85 min, and a third linear gradient to 95% mobile phase B at 90 min. This composition was held for 9 min followed by a 10 min reequilibration period to initial conditions. The eluent was passed through the UV detector, then a splitter that directed the flow to the fraction collector and mass spectrometer in an approximate 15:1 ratio. Fractions were collected every 20 s and those containing metabolites of interest were evaluated for purity by UHPLC-UV-MS to facilitate pooling. Pooled fractions were evaporated in a vacuum centrifuge and residues evaluated by NMR spectroscopy for structural elucidation.

#### NMR Spectroscopy

Samples were dissolved in 0.045 ml of DMSO-d6 “100%” (Cambridge Isotope Laboratories, Andover, MA) and placed in a 1.7 mm NMR tube in a dry argon atmosphere. ^1^H and ^13^C spectra were referenced using residual DMSO-d6 (^1^H δ = 2.50 ppm relative to TMS, δ = 0.00, ^13^C δ = 39.50 ppm relative to TMS, δ = 0.00). NMR spectra were recorded on a Bruker Avance 600 MHz (Bruker BioSpin Corporation, Billerica, MA) controlled by Topspin V4.0 and equipped with a 1.7 mm TCI Cryo probe. 1D spectra were recorded using an approximate sweep width of 8400 Hz and a total recycle time of approximately 7 s. The resulting time-averaged free induction decays were transformed using an exponential line broadening of 1.0 Hz to enhance signal to noise. For through-space experiments (ROE or NOE) the sample temperature was raised to 340 °K. 2D data were recorded using the standard pulse sequences provided by Bruker. At minimum, a 1K x 128 data matrix was acquired using a minimum of 2 scans and 16 dummy scans with a spectral width of 10000 Hz in the f2 dimension. The 2D data sets were zero-filled to at least 1k data point. Post-acquisition data processing was performed with either Topspin V3.2 or MestReNova V9.1. A series of standard 1D and 2D NMR experiments were performed on each sample.

#### Preclinical Pharmacokinetics Studies

All activities involving animals were carried out in accordance with federal, state, local and institutional guidelines governing the use of laboratory animals in research in AAALAC accredited facilities and were reviewed and approved by Pfizer’s or Bioduro’s Institutional Animal Care and Use Committee.

#### Rat Pharmacokinetics

Rat pharmacokinetics studies were done at Pfizer (Groton, CT) or BioDuro Pharmaceutical Product Development Inc. (Shanghai, PRC); Jugular vein-cannulated male Wistar-Hannover rats were purchased from Charles River Laboratories, Inc. (Wilmington, MA) or Vital River (Beijing, China) and were typically 7-10 weeks of age at the time of dosing. During the pharmacokinetic studies all animals were housed individually. Access to food and water was provided ad libitum (i.e., subjects were dosed in the fed state). In the instances where oral (po) dose was administered in the fed state, subjects were fasted overnight and fed 4 h post-dose. Compounds were administered intravenously (iv) via the tail vein (n = 2 or 3) dosed as a solution (1 mg/kg, 1 ml/kg) or via po gavage as a solution or suspension (10 mg/kg, 10 ml/kg). Doses of compound **5** were prepared immediately before dosing. The composition of each dosing vehicle is provided in Table S1. Serial blood samples were collected via the jugular vein cannula at predetermined timepoints after dosing. Animals were monitored for pain or distress throughout the study, with at least daily monitoring during normal husbandry prior to study start. At the completion of the study, animals were euthanized by overdose of inhaled anesthesia followed by exsanguination. Blood samples were collected into tubes containing K2EDTA and stored on ice until centrifugation to obtain plasma, which was stored frozen at -20 °C or lower.

#### Monkey Pharmacokinetics

All procedures performed on Cynomolgus monkeys were in accordance with regulations and established guidelines and were reviewed and approved by an Institutional Animal Care and Use Committee through an ethical review process. Monkey studies were conducted at Pfizer (Groton, CT). Male Cynomolgus monkeys were purchased from Covance (Princeton, NJ), Charles River Laboratories, Inc. (Wilmington, MA), or Envigo Global Services (Indianapolis, IN); subjects 3-8 years of age were used in pharmacokinetics studies. For each study (n=2–3), compounds **5** and **6** were dosed iv by the saphenous vein (typically 1 mg/kg and 1 ml/kg) or via po gavage (typically 5–10 mg/kg, 5 ml/kg). Subjects were monitored for pain or distress throughout the study followed by at least daily monitoring while off study. The iv dosing vehicle was optimized such that the compounds were in solution and stable for at least 24 h. Doses of **5** were prepared immediately before dosing. The composition of each dosing vehicle is provided in Table S1. Serial blood samples were collected via the femoral vein before dosing at predefined time points post-dose. Blood samples were collected into K3EDTA treated collection tubes and were stored on wet ice prior to being centrifuged to obtain plasma, which was stored frozen at - 20 °C or lower.

#### LC-MS/MS Analysis of Plasma Samples

Plasma samples were processed using protein precipitation with 500:50 acetonitrile:methanol containing propranolol (50 ng/ml) as an internal standard followed by quantitation against a standard curve (0.1-2500 ng/ml) prepared in blank plasma. Quantitation of analyte in plasma samples was done using LC-MS/MS. Standard and quality control samples, prepared in blank plasma were extracted in the same manner as the in-life samples. Briefly, a Waters ACQUITY ultra performance liquid chromatography system (Waters, Milford, MA) coupled to an Sciex 6500 triple quadrupole mass spectrometer equipped with an electrospray ionization source was used. Chromatographic separation was accomplished using a Waters Acquity UPLC BEH C18 column (1.7 µm, 2.1 × 50 mm) maintained at either room temperature or 45 °C. The mobile phase (2 solvents gradient) was optimized to achieve good separation between the analytes. Typically, solvent A constituted of 0.025% formic acid and 1 mM ammonium acetate in water/acetonitrile (95:5 v/v), and solvent B included 0.025% formic acid and 1 mM ammonium acetate in water/acetonitrile (5:95 v/v). The gradient generally began at 3-30% B until about 1.2 min, followed by an increase to 50-65% B to 1.6 min, then decreased to 10-30% B until ∼1.7 – 1.9 min. MS/MS methods were the same as described for in vitro samples. Analyst 1.7 software was used for peak integration and standard curve regression. The protein precipitation and mobile phase solutions used for compound **1** contained 1% formic acid. To avoid artifacts of epimerization in samples from compound **5**, sample evaporation was avoided.

#### Pharmacokinetic Analysis

Pharmacokinetic parameters were calculated using noncompartmental analysis (Watson v.7.5, Thermo Scientific). The area under the plasma concentration-time curve from *t* = 0 to infinity (AUC0-∞) was estimated using the linear trapezoidal rule. In some instances, pharmacokinetic calculations were generated using the linear log-linear trapezoidal rule and C0 was calculated using the equation:

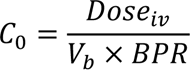

where Vb is the blood volume (rat, 69.0 ml/min/kg; monkey, 62.3 ml/kg) and BPR is the blood to plasma ratio. Plasma clearance (CLp) was calculated as:

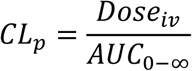

The terminal rate constant (kel) was calculated by linear regression of the terminal phase of the log-linear concentration-time curve and the terminal elimination t1/2 was calculated as:

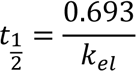

Apparent steady state distribution volume (Vdss) was determined by clearance multiplied by mean residence time. Oral bioavailability (F) was defined as:

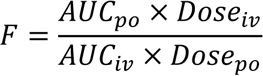

The fraction of the oral dose absorbed (Fa x Fg) was estimated using the equation (*24*):

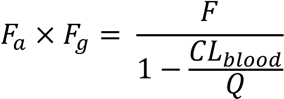

A hepatic blood flow (Q) of 70 ml/min/kg and 44 ml/min/kg was used for rats and monkeys, respectively (*55*). Blood clearance (CLblood) was calculated by dividing CLp by the blood-to-plasma ratio (ranging from 0.6–0.8) for the compounds in the respective preclinical species.

#### General Toxicity studies

Study designs and parameters evaluated in toxicology studies were consistent with accepted principles and practices as outlined in ICH, OECD guidelines, and national regulations (US FDA, European Community Directives, and Japan regulations). All definitive studies were conducted in accordance with US FDA GLP regulations in an OECD MAD member state. In vivo studies were conducted in accordance with the current guidelines for animal welfare (National Research Council Guide for the Care and Use of Laboratory Animals, 2011). The procedures used in these studies were reviewed and approved by the Institutional Animal Care and Use Committee.

#### Chronic Toxicity Studies with PF-07321332 (**6**)

Briefly, groups of male and female Wistar Han rats or Mauritian cynomolgus monkeys were administered vehicle [2% (v/v) Polysorbate 80 in 0.5% (w/v) of methylcellulose A4M in purified water], MTBE control [0.9% MTBE ( w/v) in 2% (v/v) Polysorbate 80 in 0.5% (w/v) of methylcellulose in purified water] or PF-07321332 in vehicle (group size, doses and dosing regimen are described in Table S10). Study evaluations included detailed clinical observations, body weights, food consumption, micronucleus assessment (rat only), electrocardiography (monkeys only), ophthalmology, clinical pathology (hematology, clinical chemistry, coagulation, urinalysis), toxicokinetics, gross and microscopic pathology. Adversity assessments and considerations for setting the no-observed-adverse-effect-levels included integrated evaluation of the incidence and severity of clinical and pathology findings.

#### In vivo Micronucleus Assessment in Rats

The potential of PF-07321332 to induce chromosome damage was determined by the increased frequency of micronucleated reticulocytes in peripheral blood samples from vehicle- and PF- 07321332-treated rats from the 2-week GLP toxicity study. The micronucleus portion of this study was designed using literature protocols (*56, 57*) and internationally accepted guidelines (*58, 59*). Briefly, peripheral blood was collected on Day 4 from the first 5 surviving animals/sex from the vehicle/MTBE and PF-07321332 MTBE co-solvate (60, 200, and 1000 mg/kg/day) dose groups, fixed and stained with fluorescent markers for red blood cells, platelets, and DNA, followed by quantitation of micronucleated reticulocytes and erythrocytes using flow cytometry (*60*).

#### Bacterial Reverse Mutation Assay

The mutagenic potential of PF-07321332 was evaluated by measuring the ability to induce reverse mutations at selected loci of several strains of Salmonella typhimurium (TA98, TA100, TA1535, and TA1537) and at the tryptophan locus of Escherichia coli strain WP2 uvrA in the presence and absence of metabolic activation (Aroclor 1254-induced rat liver S9/NADPH) using standard protocols (*61, 62*) and internationally accepted guidelines (*59, 63*). A concentration-related increase in the mean revertants per plate of at least one tester strain over a minimum of two increasing concentrations of PF-07321332 was to be considered as positive for mutagenic potential.

#### In Vitro Mammalian Cell Micronucleus Assay in TK6 Cells

The anuegenic and clastrogenic potential of PF-07321332 was evaluated, in the presence and the absence of metabolic activation (rat liver S9), based upon its ability to induce micronuclei in TK6 cells, a human lymphoblastoid cell line. The assay design is based on literature (*64, 65*) and internationally accepted guidelines (*59, 66*). Briefly, TK6 cells were incubated with (4 or 24 h) vehicle or PF-07321332 in the presence or absence of metabolic activation (Aroclor 1254- induced rat liver S9/NADPH), harvested, fixed, stained with acridine orange and scored for frequency of micronuclei in mononucleated cells by light microscopy.

#### Secondary Pharmacology General Methods for PF-07321332 (**6**) Screening

Secondary (off-target) pharmacology studies were conducted by Eurofins Cerep (Celle-Lévescault, France) on behalf of Pfizer Inc. The in vitro off-target pharmacology of PF- 07321332 was assessed at 100 µM in a broad target profiling panel which represents targets with known links to potential safety concerns and includes G-protein coupled receptors, ion channels, transporters, and enzymes according to established protocols.

#### FLIPR^®^ Calcium Assays

Cells used in the assay were stably transfected with the receptor of interest (adrenergic alpha1a, dopamine 1, histamine 1, muscarinic 1, muscarinic 3 and serotonin 2b). Activation of the receptor by an agonist in this assay system results in an increase in intracellular calcium levels which is measured using a calcium specific dye. Cells were plated at 7,500 cells per well (50 µl per well) in black walled clear bottomed 384-well plates 24 hours prior to running the assay.

Medium was removed from the plates and 80 µl of Hanks balanced salt solution (HBSS)/HEPES containing Calcium 5 dye (Molecular Devices, Sunnyvale CA, USA; Cat # R8186) and probenecid (1.25 mM) was added to each well and the plate was returned to the incubator for 1 hour to allow dye loading. Compound solution (10 µl) was added to each well by the FLIPR Tetra® instrument (Molecular Devices, Sunnyvale CA, USA) to measure agonist activity of the compound by measuring the change in fluorescence from baseline over a 60 second period (Excitation 470-495 nm; Emission 515-575 nm). Subsequently 10 µl of agonist (EC80 value) was added to each well by the FLIPR Tetra® instrument to evaluate antagonist activity, with the change in fluorescence from baseline being measured over a 60 second period.

#### Beta Arrestin Assays

The beta arrestin assay relies on enzyme fragment complementation with the respective stably transfected GPCR (adrenergic beta 2, cannabinoid 1 and mu opioid) being tagged with an inactive portion of the enzyme β-galactosidase and a co-transfected β-arrestin that is tagged with the complementary portion of β-galactosidase. Recruitment of β-arrestin to the GPCR, results in a functional enzyme that generates a chemiluminescent signal when substrate is added. Cells were plated at 5,000 cells per well (40 µl per well) in black walled clear bottomed 384-well plates 24 hours prior to running the assay. Medium was removed from the plates. For agonist studies 15 µl of HBSS/HEPES containing compound was added to the cells and the plate was incubated at room temperature for 90 min. For antagonist studies 15 µl of HBSS/HEPES containing compound was added to the cells and was incubated for 15 min prior to the addition of 15 µl of an EC80 concentration of agonist. The plate was subsequently incubated at room temperature for 90 min. Both assays were terminated by addition of 15 µl of a Beta-Glo® solution (Promega). Following an additional 30 min incubation the luminescence of each well was measured to determine the level of receptor activation.

#### Amine Transporter Assays

The amine transporter assay measures the ability of compounds to inhibit the activity of the norepinephrine (NET) dopamine (DAT) or serotonin (SERT) transporters by measuring the real time uptake of a dye labeled amine. HBSS/HEPES containing compound (5 µl) was added to the wells of black walled clear bottomed 384-well plate. Transporter dye (25 µl) (Molecular Devices, Sunnyvale CA, USA; Cat # R8174) was added to each well. Finally, 15,000 cells (20 µl) stably expressing the amine transporter of interest were added to each well and the plate is incubated at 37°C for 30 min (DAT) or 60 min (NET and SERT). The plate is transferred to the FLIPR Tetra® instrument and the fluorescence of each well was measured (Ex 470-495 nM; Em 515-575 nM). The level of fluorescence measured directly relates to the level of uptake of the dye labelled amine, with a reduction in levels being related to an inhibition of the respective transporter.

#### hERG Binding Assay

Human embryonic kidney (HEK) cells stably transfected with a doxycycline inducible plasmid expressing the hERG channel (Accession Number: NM_000238) were cultured in suspension in Ex-cell 293 Serum Free Medium containing fetal bovine serum (5% v/v), L-Glutamine (6 mM), Blasticidin (5 µg/ml) and Zeocin (600 µg/ml) at 37 °C in a humidified environment (5% CO2/95% air). hERG expression was induced by the addition of doxycycline (1 µg/ml) 48 h prior to harvesting by centrifugation. Cell pellets were resuspended in ice cold homogenization buffer (1 mM EDTA, 1 mM EGTA, 1 mM NaHCO3, and cOmplete™ protease Inhibitor cocktail). Cells were homogenized using a dounce homogenizer (20 strokes), and centrifuged (1,000xg) for 10 min at 4°C. The supernatant was transferred to a new tube and was centrifuged a second time (25,000xg) for 20 minutes at 4°C. The supernatant was discarded, and the pellet was resuspended in buffer (50 mM HEPES, 10 mM MgCl2, bovine serum albumin (0.2% w/v) and cOmplete™ protease inhibitor cocktail). The samples were adjusted to 5 mg/ml and frozen. For the assay, membrane aliquots were thawed on ice and diluted to 200 µg/ml in assay buffer (25 mM HEPES, 15 mM KCl, 1 mM MgCl2, and 0.05% (v/v) Pluronic F127). A Cy3B tagged N-desmethyl dofetilide ligand was prepared in the same assay buffer solution (5 nM). Compound or vehicle (DMSO) was spotted into each well of a black 384-well low-volume plate. Membrane homogenate (15 µL) and Cy3B tagged ligand (10 µL) were then added to each well and the plate was incubated at room temperature for 16 h. Fluorescence polarization measurements were made using an Envision plate reader (Perkin Elmer, Waltham, MA, USA) and mP values were used for analysis. Binding Ki values were determined using the Cheng-Prusoff equation (Ki = IC50/(1+L/Kd)), where L was the labelled ligand concentration in the assay (2 nM), and the Kd value (1.35 nM) the affinity constant for the labelled ligand (*67*).

#### GLP hERG Assay

GLP hERG studies were conducted at Charles River Laboratories Inc. (Cleveland, OH). In brief, human embryonic kidney (HEK) cells stably expressing the human Kv11.1 (hERG) channel were cultured in Dulbecco’s Modified Eagle Medium/Nutrient Mixture F-12 (DMEM/F-12) supplemented with 10% (v./v) fetal bovine serum, 100 U/ml penicillin G sodium, 100 µg/ml streptomycin sulfate and 500 µg/ml G418. Harvested cells were transferred to the recording chamber and superfused with vehicle control solution. Micropipette solution for whole cell patch clamp recordings was composed of (mM): potassium aspartate, 130; MgCl2, 5; EGTA, 5; ATP, 4; HEPES, 10; pH adjusted to 7.2 with KOH. The recording was performed at a temperature of 33 to 35 °C. Micropipettes for patch clamp recording were made from glass capillary tubing using a P 97 (Sutter Instruments, Novato, CA) or PC-10 (Narishige, Amityville, NY) micropipette puller. A commercial patch clamp amplifier (Axopatch 200B from Molecular Devices, San Jose, CA) was used for whole cell recordings. Before digitization, current records were low-pass filtered at one-fifth of the sampling frequency low-pass filtered at one-fifth of the sampling frequency. Cells were held at -80 mV. Onset and steady state inhibition of hERG potassium current were measured using a pulse pattern with fixed amplitudes (conditioning prepulse +20 mV for 1 s; repolarizing test ramp to 80 mV ( 0.5 V/s) repeated at 5 s intervals).

Each recording ended with a final application of a supramaximal concentration of the reference substance (E 4031, 500 nM) to assess the contribution of endogenous currents. The remaining uninhibited current was subtracted off-line digitally from the data to determine the potency of the test substance for hERG inhibition. One or more test article concentrations were applied sequentially (without washout between test substance concentrations) in ascending order, to each cell. Peak current was measured during the test ramp. A steady state was maintained for at least 20 s before applying test article or positive control. Peak current was measured until a new steady state was achieved.

#### Additional Ion Channel Assays

Chinese hamster ovary (CHO) cells stably expressing human Cav1.2/β2/α2δ1 calcium channel (Catalogue No. CT6004; Charles River Cleveland, OH, USA) were cultured in Ham’s F12 medium supplemented with fetal bovine serum (FBS; 10% (v/v)), G418 (0.25 mg/ml), hygromycin (0.25 mg/ml), zeocin (0.4 mg/ml), and blasticidin (0.01 mg/ml). On the day prior to cell harvest, tetracycline (1 µg/ml) was added to the media to induce channel expression and the calcium channel antagonist verapamil (15 µg/ml) was added to minimize calcium-induced cytotoxicity. CHO cells stably expressing the human Nav1.5 sodium channel (Catalogue No. CT6007; Charles River Cleveland, OH, USA) were cultured in Ham’s F12 media supplemented with 10% FBS (10% (v/v) and G418 (0.25 mg/ml).

Both cell lines were cultured at 37 °C in a humidified environment (5% CO2/95% air). On the day of the experiment, cells were harvested at 70-80% confluency by rinsing with Hank’s Balanced Salt Solution and incubating for 2 min in Accutase (Innovative Cell Technologies, San Diego, CA, USA). Cells were resuspended (2 million cells per ml) in CHO-S-SFM II serum-free medium supplemented with 20 mM HEPES and were allowed to recover for 45 min with constant stirring prior to electrophysiological measurements. All tissue culture media and reagents were obtained from Thermo Fisher (Waltham, MA, USA), unless otherwise stated.

Ionic currents were evaluated in the whole-cell configuration using the Qube384 automated planar patch clamp platform (Sophion Bioscience A/S, Ballerup, Denmark). QChip 384X plates, containing 10 patch clamp holes per well, were used to maximize success rate, which was routinely > 95%. For Cav1.2 experiments, the external solution was composed of (in mM): 137.9 NaCl, 5.3 KCl, 0.49 MgCl2, 10 CaCl2, 10 HEPES, 0.34 Na2HPO4, 4.16 NaHCO3, 0.41 MgSO4, 5.5 glucose, pH 7.4, 312 mOsm/kg. The internal solution contained (in mM): 27 CsF, 112 CsCl, 2 MgCl2, 10 EGTA, 10 HEPES, 2 Na2ATP, pH 7.2, 307 mOsm/kg. For Nav1.5 experiments, the external solution was composed of (in mM): 137.9 NaCl, 5.3 KCl, 0.49 MgCl2, 1.8 CaCl2, 10 HEPES, 0.34 Na2HPO4, 4.16 NaHCO3, 0.41 MgSO4, 5.5 glucose, pH 7.4, and osmolarity of 303 mOsm/kg. The internal solution contained (in mM): 92 CsF, 55 CsCl, 2 MgCl2, 5 EGTA, 5 HEPES, 1 MgATP, pH 7.2, 298 mOsm/kg. The osmolarity of the buffer was adjusted by the addition of sucrose as required.

The Cav1.2 current was elicited by a voltage step to 0 mV for 150 ms from a holding potential of -40 mV. Voltage steps were repeated at 0.05 Hz, and Cav1.2 amplitude was measured as the peak current at 0 mV. For the Nav1.5 current, from an initial holding potential of -80 mV, a 200 ms prepulse to -120 mV was used to homogenize channel inactivation, followed by a 40 ms step to a test potential of -15 mV. Membrane potential was further depolarized to +40 mV for 200 ms to completely inactivate the peak Nav1.5 current, followed by a ramp from +40 mV to -80 mV (-1.2 mV.ms). This voltage pattern was repeated at 0.2 Hz, with the Nav1.5 peak current defined as the maximum current during the step to -15 mV. All studies were conducted at 23°C.

Compounds were initially dissolved and diluted in DMSO, with a final dilution by the addition of external solution to generate final working concentrations. The final DMSO concentration in all experiments was 0.33% (v/v). Three vehicle periods each lasting 5 minutes were applied to establish a stable baseline, each well followed by the addition of increasing concentrations of test compound, with each exposure lasting 5 minutes. Patch clamp data were analyzed using Assay Software (Version 6.4.72; Sophion Bioscience A/S, Ballerup, Denmark). Current amplitudes were determined by averaging the last 4 currents under each test condition. The percent inhibition of each compound was determined by taking the ratio of current amplitude measured in the presence of various concentrations of the test compound (ICompound) versus the vehicle control current (IVehicle):

% Inhibition = [1-(ICompound/IVehicle)] * 100%.

A dose-response curve was generated IC50 value defined for each compound by fitting the data to a four-parameter logistical equation using the Sophion Analyzer software. The minimum response and slope were free fitted and maximum response was fixed to 100%.

#### Phosphodiesterase Assays

The phosphodiesterase (PDE) assays measure the conversion of 3’, 5’-[^3^H] cAMP to 5’-[^3^H] AMP (for PDE 3A1 and 4D3) or 3’, 5’-[^3^H] cGMP to 5’-[^3^H] GMP (for 5A1) by the relevant PDE enzyme subtype. Yttrium silicate (YSi) scintillation proximity (SPA) beads bind selectively to 5’-[^3^H] AMP or 5’-[^3^H] GMP, with the magnitude of radioactive counts being directly related to PDE enzymatic activity. The assay was performed in white walled opaque bottom 384-well plates. Test compound (1 μl) in dimethyl sulfoxide was added to each well. Enzyme solution was then added to each well in buffer (in mM: Trizma, 50 (pH7.5); MgCl2, 1.3 mM) containing Brij 35 (0.01% (v/v)). Subsequently, 20 µl of 3’,5’-[^3^H] cGMP (125 nM) or 20 µl of 3’,5’-[^3^H] cAMP (50 nM) was added to each well to start the reaction and the plate was incubated for 30 min at 25 °C. The reaction was terminated by the addition of 20 µl of PDE YSi SPA beads (Perkin Elmer, Waltham, MA). Following an additional 8 hour incubation period the plates were read on a MicroBeta radioactive plate counter (Perkin Elmer, Waltham, MA, USA) to determine radioactive counts per well.

#### Bromodomain-Containing Protein 4 (BRD4) Binding Assay

The BRD4 fluorescent polarization binding assay uses purified His-tagged BRD4 protein and its interaction with a Cy5 labelled small molecule probe that binds to the BRD4 site involved in the interaction with tetra-acetylated histone H4 peptide. In brief, the assay is performed in low volume black 384 well flat-bottomed polystyrene plates. Compound/vehicle or standard (5 µl) were added to wells followed by His-tagged BRD4 (10 µl; 40 nM final concentration in assay). Following a 15 min incubation at room temperature a proprietary Cy5-labelled probe molecule (5 µl; 2 nM final concentration in assay) was added. Following, an additional 16 h incubation at room temperature fluorescence polarization measurements were made using an Envision plate reader (Perkin Elmer, Waltham, MA, USA) and mP values were used for analysis.

#### Data Analysis

Agonist/antagonist curves were plotted from individual experiments, and EC50/IC50 values were determined using a four-parameter logistic fit. EC50 is defined as the concentration of the test article that produced a response that was equal to 50% of the maximal system response. IC50 is defined as the concentration of the test article that produced a 50% inhibition of a maximal response. An apparent KB value for antagonist activity was calculated using the following equation:

Apparent KB = IC50/(1+([A]/Agonist EC50) where the KB value is the dissociation constant of antagonist for the receptor, IC50 is the response produced by the test article in the presence of [A], the concentration of agonist used in the assay. Agonist EC50 is the EC50 value of the reference agonist used in the assay when tested alone. (*67*).

#### First-in-Human (FIH) Clinical Trial

##### Study design for the FIH clinical trial in healthy adult participants

The single ascending dose, which is a part of an ongoing multi-part FIH phase 1 study [NCT04756531], was conducted at the sponsor’s Clinical Research Unit in New Haven, CT, USA. The protocol was approved by an independent institutional review board and all participants provided informed consent before screening. The study was conducted in compliance with ethical principles of the Declaration of Helsinki and International Council for Harmonization Good Clinical Practice guidelines. All local regulatory requirements were followed. The design for the single ascending dose portion was investigator- and participant-blinded, sponsor-open, randomized, 4-period cross-over (with Period 3 and 4 being optional) in 2 interleaving cohorts with placebo substitution. The interleaving design enabled both within- and between-participant assessments.

Participants were required to be healthy adults aged 18–60 years, with a body mass index of 17.5–30.5 kg/m2 and a body weight of >50 kg. Exclusion criteria included history of clinically significant hematologic, renal, endocrine, pulmonary, gastrointestinal, cardiovascular, hepatic, psychiatric, neurologic, or allergic disease and conditions affecting drug absorption.

Randomization was performed using a sponsor-provided randomization schedule. A total of 13 participants were randomized in this part. At each dose level, 4 participants received PF- 07321332 and 2 participants received placebo. In a given participant, there was an interval of at least 5 days between dosing to allow for washout of PF-07321332 and review safety and pharmacokinetics data from each dose level before decisions were made on the next dose. Participants who discontinued for non-safety related reasons prior to completion of the study may have been replaced at the discretion of the principal investigator and Sponsor.

The starting dose of PF-07321332 150 mg alone was derived from nonclinical information on pharmacokinetics and metabolism of PF-07321332. Progression to the next dose level occurred if the last dosing paradigm (PF-07321332 alone or with RTV) was well tolerated and after satisfactory review of the available safety and pharmacokinetics data. Subsequent dosing paradigm was selected with projected (based on the pharmacokinetics data available the time of each dose escalation) mean exposure of <3.3-fold of the previous highest observed safe and tolerated mean exposure. In the PF-07321332/RTV co-administration dosing paradigm, each subject (active and placebo) received one tablet (100 mg) of RTV at -12 h, 0 h and 12 h. PF- 07321332 was administered as an oral suspension under fasted conditions at 0 h (minimum fast of ∼10 h prior to treatment). Data from two of the doses in this part, PF-07321332 150 mg alone and PF-07321332 250 mg with RTV, are presented in this manuscript.

Blood, to obtain plasma samples for pharmacokinetic assessments, was collected up to 72 hours post dose. Plasma samples were analyzed for PF-07321332 concentrations at Pfizer (Groton, CT) using a validated sensitive and specific LC-MS/MS method in compliance with the Sponsor SOPs. Plasma specimens were stored at approximately -70 °C until analysis and assayed within 150 days of established stability data generated during validation. Calibration standard responses were linear over the range of 10.0 to 50,000 ng/ml using a weighted (1/x^2^) linear least squares regression. Those samples with concentrations above the upper limits of quantification were adequately diluted into calibration range. The lower limit of quantification (LLOQ) for

PF-07321332 was 10.0 ng/ml. Clinical specimens with plasma PF-07321332 concentrations below the LLOQ were reported as “<10.0 ng/ml”. The between-day assay accuracy, expressed as percent relative error (%RE), for quality control (QC) concentrations, ranged from -3.7% to 2.0% for the low, medium, high, and diluted QC samples. Assay precision, expressed as the between-day percent coefficient of variation (%CV) of the mean estimated concentrations of QC samples was ≤5.9% for low (30.0 ng/ml), medium (1000 ng/ml), high (37,500 ng/ml), and diluted (100,000 ng/ml) concentrations.

---

**Figure S1.**
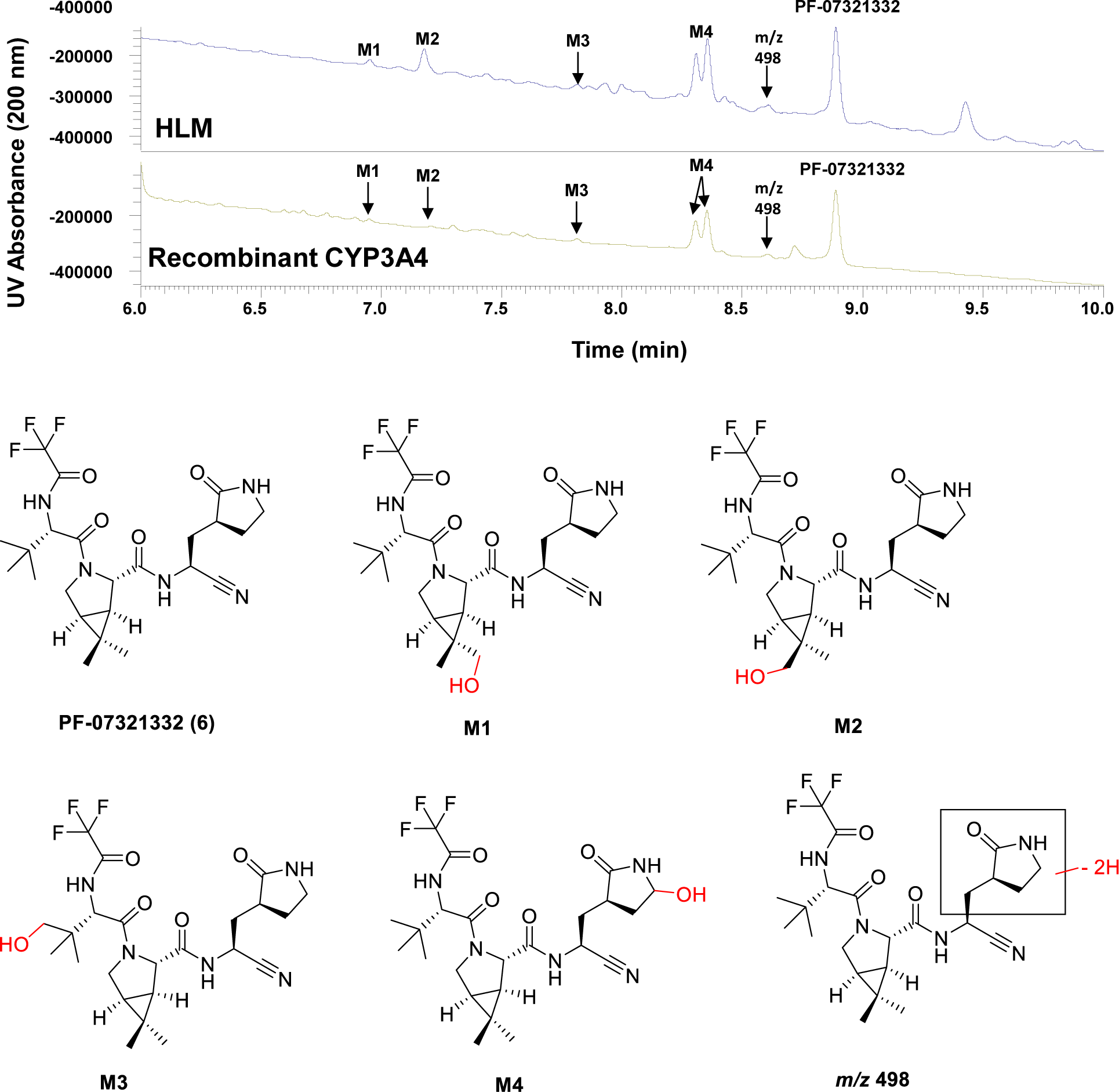
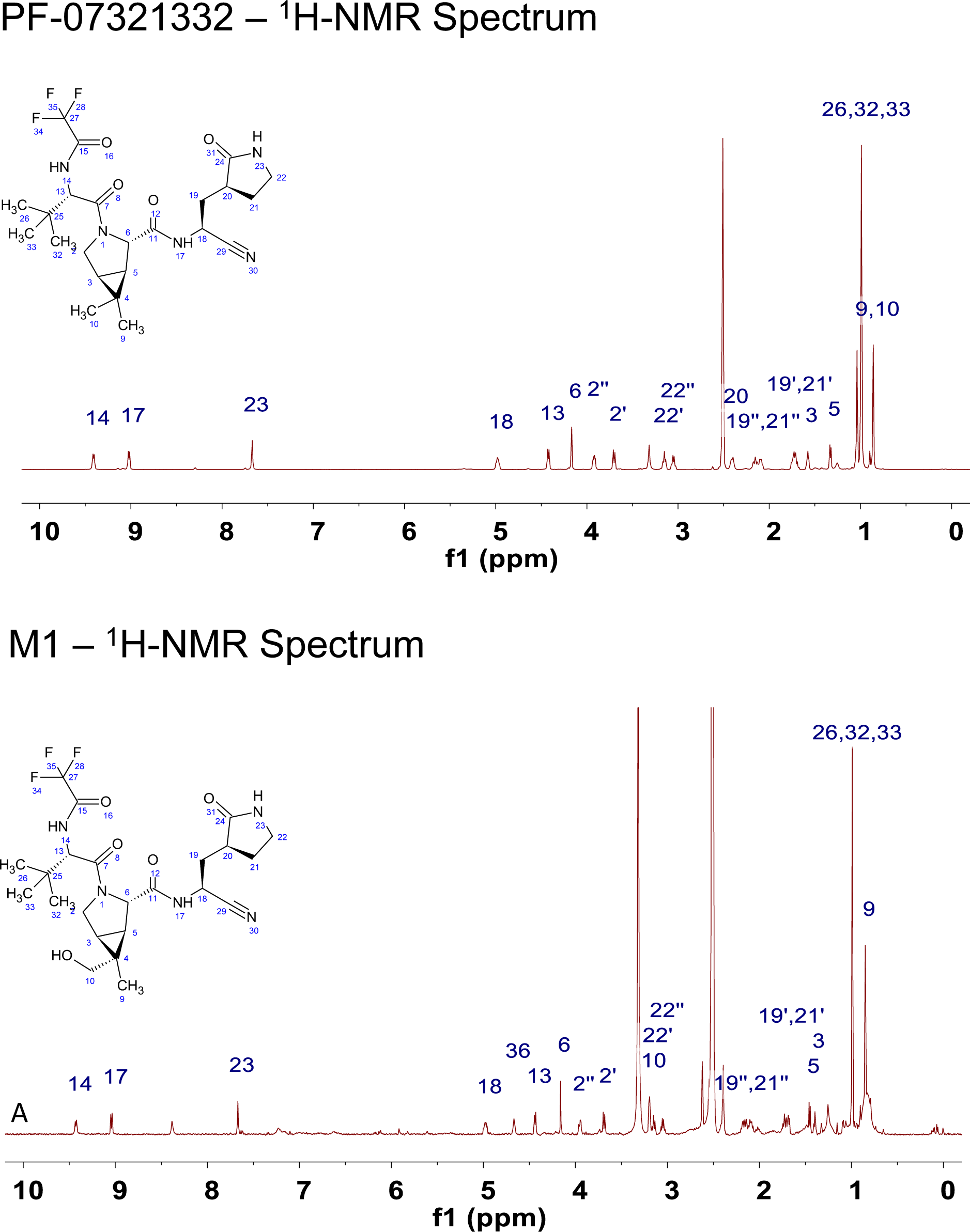
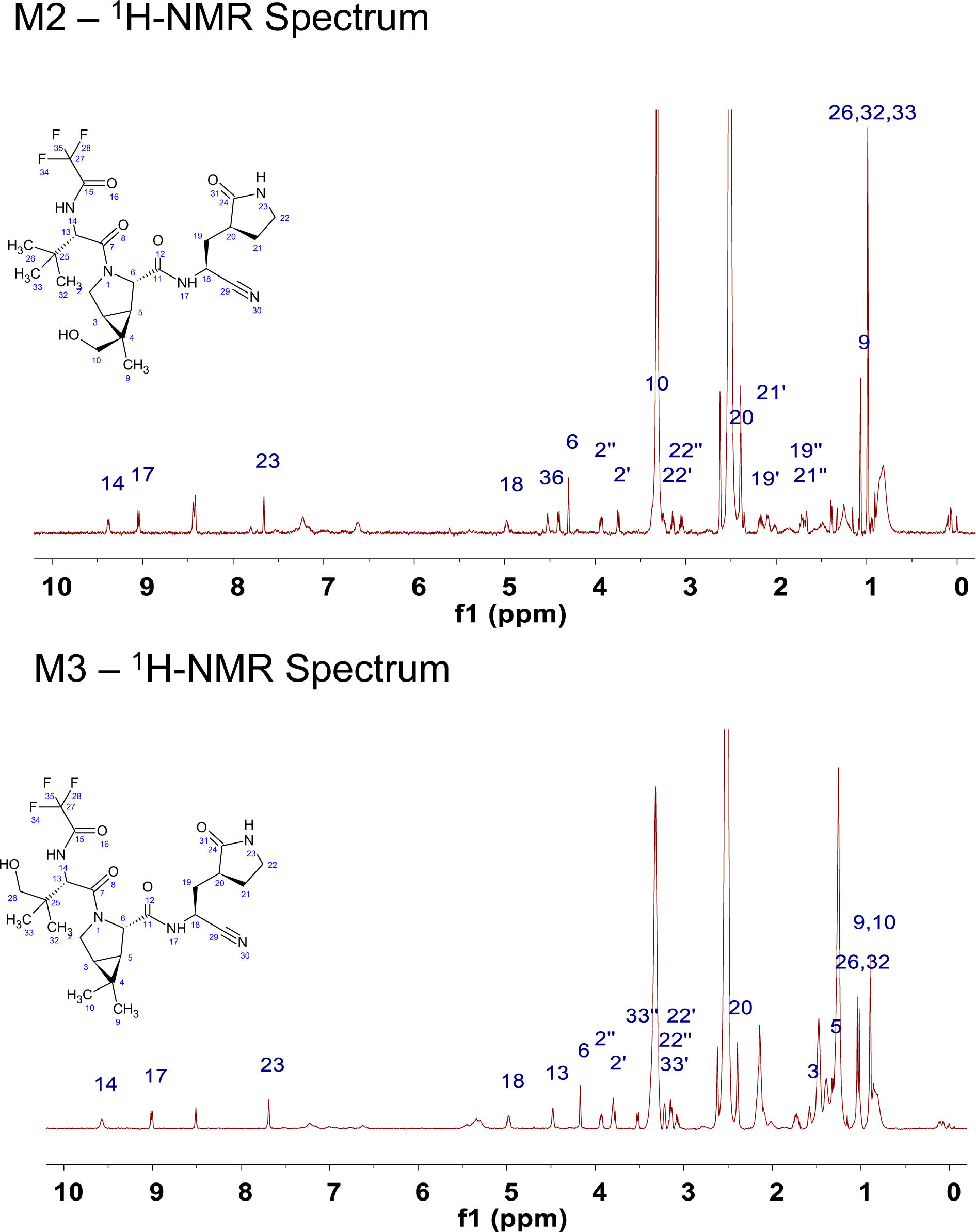
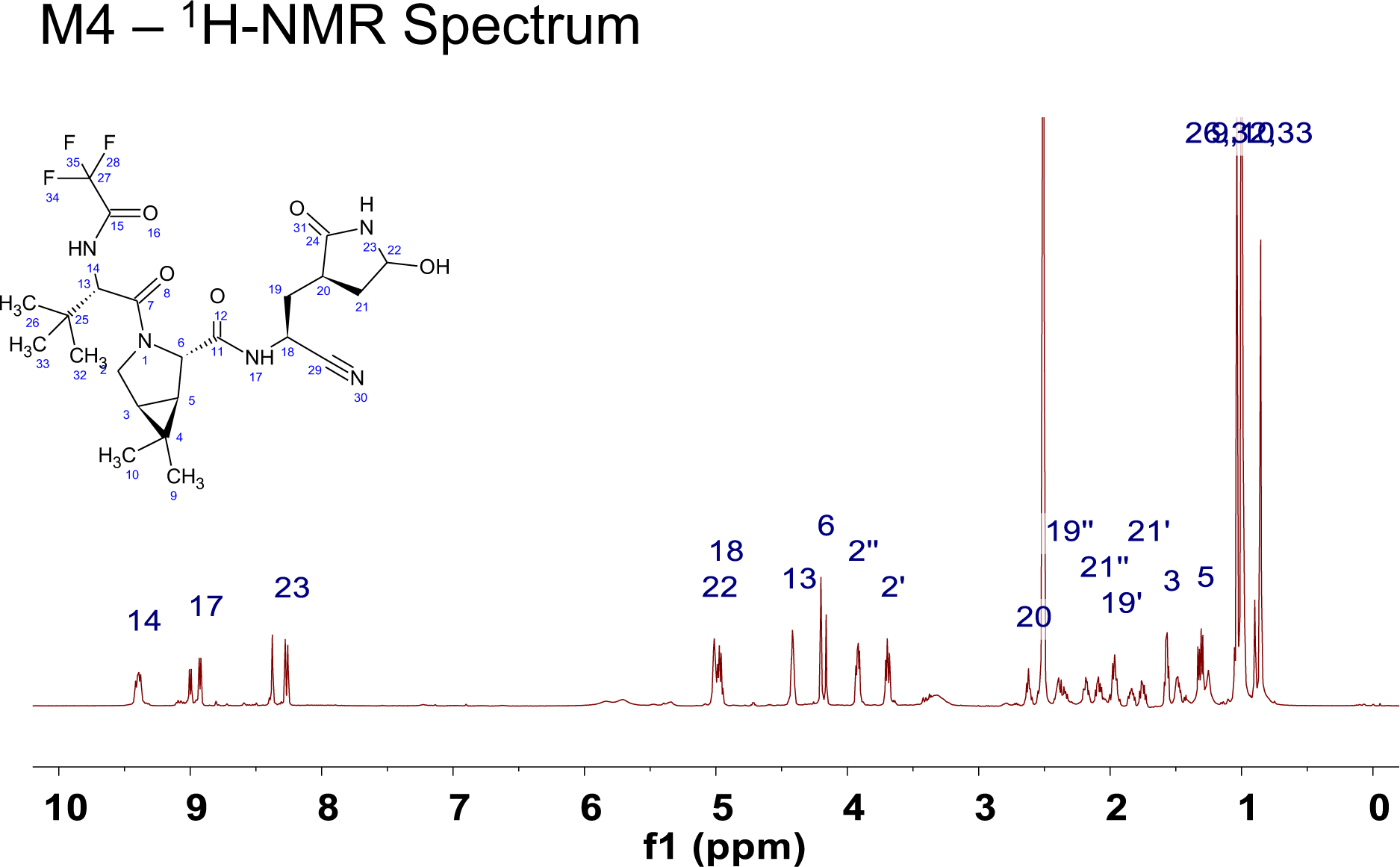
UHPLC-UV chromatograms of PF-07321332 (10 mM) incubations in NADPH-supplemented HLM and recombinant human CYP3A4. Structures of oxidative metabolites (M1–M4 and m/z 498) depicted were elucidated by ^1^H and ^13^C 1D and 2D NMR spectroscopy following their biosynthesis and purification from HLM. Metabolite M4 comprised of two closely eluting diastereomers. ^1^H NMR are shown for illustrative purposes.

**Table S1.** Preclinical pharmacokinetics of SARS-CoV2 M^pro^ inhibitors. ^a^Pharmacokinetic parameters were calculated from plasma concentration–time data and are reported as mean (± S.D. for n=3 and individual values for n=2). All pharmacokinetics were conducted in male gender of each species (Wistar-Han rats and cynomolgus monkey). Oral (po) studies were conducted in the fed state unless otherwise noted. For intravenous (iv) doses in rats, compounds were administered as solutions in 10% DMSO/30% PEG400/60% deionized water or 10% DMSO/50% PEG400/40% deionized water. For iv studies in monkeys, compound **1** was administered as a solution in 10% (v/v) PEG400/90% (v/v) of 23% (w/v) 2-hydroxypropyl-β- cyclodextrin in aqueous sodium phosphate buffer pH = 6.0, whereas compound **5** was administered as a solution in 10% PEG400/90% (v/v) of 23% (w/v) 2-hydroxypropyl-β- cyclodextrin in deionized water. For po studies in rats and monkeys, compound **1** was administered as a solution (2 mg/kg) or suspension (5 and 20 mg/kg) in 0.5% (w/v) aqueous methyl cellulose, whereas compounds **4** and **5** were administered as a solution in 0.5% (w/v) aqueous methyl cellulose containing 2% Tween80. For po studies in rats, compound **2** was administered as a suspension in 0.5% (w/v) aqueous methyl cellulose containing 2% Tween80. *^b^*po studies were conducted in the fasted state. *^c^*po formulation was 10% ethanol/10% capmul MCM/35% PEG400/45% Tween 80 which was preceded with 8 ml/kg oral of 1% (w/v) hydroxypropyl cellulose in deionized water immediately prior to dose. *^d^*Fa x Fg = fraction of the po dose absorbed. *^e^*AUC0-t is reported and t1/2 could not be determined (N.D.) due to a lack or measurable time points in the elimination phase.

| Compound | Species<sup>a</sup> | Dose<br>(mg/kg) | C<sub>max</sub><br>(ng/ml) | T<sub>max</sub> (h) | AUC<sub>0-∞</sub><br>(ng·h/ml) | CL<sub>p</sub><br>(ml/min/kg) | Vd<sub>ss</sub><br>(l/kg) | t<sub>1/2</sub><br>(h) | Oral F<br>(%) | F<sub>a</sub> x F<sub>g</sub><br>(%)<sup>d</sup> |
| --- | --- | --- | --- | --- | --- | --- | --- | --- | --- | --- |
| **1** | Rat | 2.0 (iv) | ---- | ---- | 1250 ± 146 | 27.0 ± 3.1 | 0.75 ± 0.24 | 0.72 ± 0.12 | ---- | ---- |
|  |  | 2.0 (po)<sup>b</sup> | 6.54 ± 2.40 | 0.33 ± 0.14 | 17.3 ± 9.5 | ---- | ---- | 1.7 ± 0.7 | 1.4 ± 0.8 | 3.3 |
|  |  | 20 (po)<sup>b</sup> | 71.6 ± 17.4 | 0.25 ± 0.00 | 162 ± 33 | ---- | ---- | 1.6 ± 0.4 | 1.3 ± 0.3 | 3.1 |
|  | Monkey | 1.0 (iv) | ---- | ---- | 583 (552, 614) | 28.7 (27.1, 30.2) | 1.4 (1.3, 1.5) | 1.2 (1.1, 1.3) | ---- | ---- |
|  |  | 5.0 (po)<sup>c</sup> | 0.404<br>(0.204, 0.605) | 1.0 (1.0, 1.0) | 0.466<br>(0.328, 0.603) | ---- | ---- | N.D. | < 0.05 | <1.0 |
| **2** | Rat | 1.0 (iv) | ---- | ---- | 427 (402, 451) | 39.3 (37.0, 41.5) | 0.91 (0.85, 0.97) | 0.62 (0.49, 0.74) | ---- | ---- |
|  |  | 10 (po) | 172 (152, 192) | 0.38 (0.25, 0.50) | 325 (315, 334) | ---- | ---- | 2.6 (1.8, 3.5) | 7.6 (7.4, 7.8) | 38 |
| **4** | Rat | 1.0 (iv) | ---- | ---- | 393 (350, 436) | 42.9 (38.2, 47.6) | 1.7 (1.5, 2.0) | 8.0 (7.8, 8.2) | ---- | ---- |
|  |  | 10 (po) | 411 (372, 449) | 0.38 (0.25, 0.50) | 410 (295, 524) | ---- | ---- | 3.8 (3.7, 3.9) | 10 (7.5, 13) | 84 |
| **5** | Rat | 1.0 (iv) | ---- | ---- | 538 (531, 544) | 31.0 (30.6, 31.4) | 2.2 (2.0, 2.3) | 6.8 (6.6, 7.0) | ---- | ---- |
|  |  | 10 (po) | 911 (711, 1110) | 1.0 (1.0, 1.0) | 2950 (2910, 2980) | ---- | ---- | 3.3 (2.9, 3.8) | 33 (33, 34) | 100 |
|  |  | 10 (po)<sup>c</sup> | 521 ± 67 | 0.67 ± 0.29 | 2310 ± 643 | ---- | ---- | 3.4 ± 0.8 | 54 ± 15 | 100 |
|  | Monkey | 1.0 (iv) | ---- | ---- | 841 (602, 682) | 26.0 (24.4, 27.6) | 0.58 (0.58, 0.59) | 1.1 (1.1, 1.1) | ---- | ---- |
|  |  | 10 (po) | 331 (112, 550) | 1.5 (1.0, 2.0) | 505 (265, 744) | ---- | ---- | 6.1 (5.7, 6.6) | 7.9 (4.1, 12) | 66 |

**Table S2.**
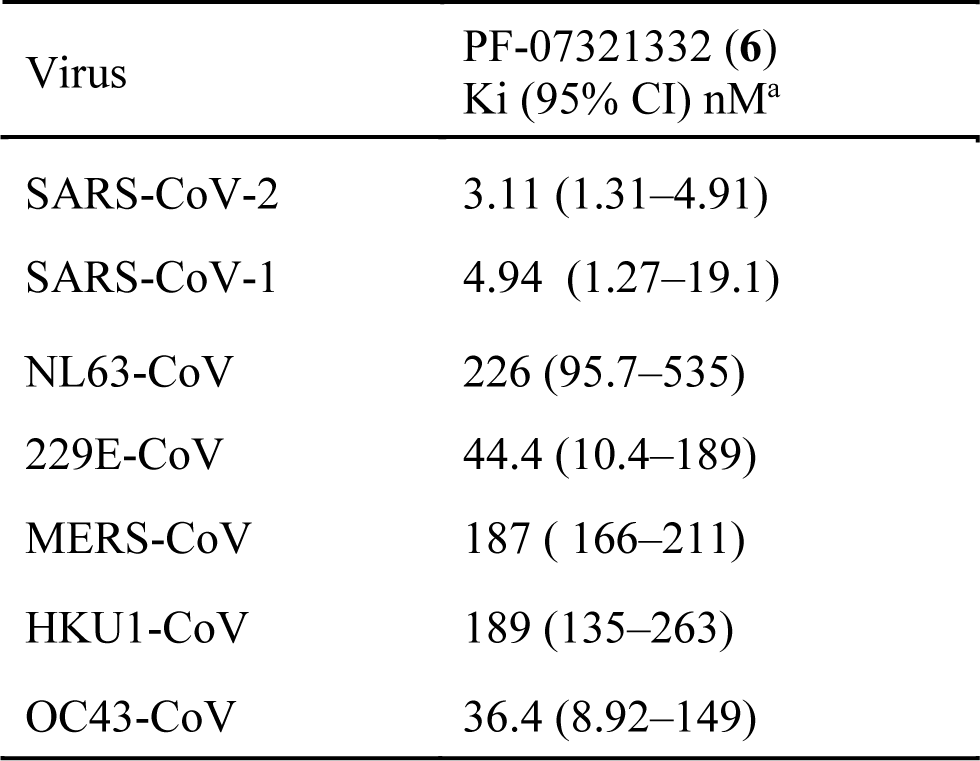
PF-07321332 (6) inhibition of human coronavirus main proteases. ^a^PF-07321332 was tested for its ability to inhibit proteolytic activity of M^Pro^ of SARS-CoV-2 as well as related coronaviruses in a FRET assay. PF-07321332 was tested up to 30 μM. Data are expressed as inhibitory constants (Ki) relative to controls with 0% inhibition wells containing 1% DMSO and full inhibition achieved by 30 μM of a broad-spectrum antiviral compound GC376. Data shown are the geometric mean and 95% confidence intervals (CI) from 3 independent experiments

**Table S3.** Selectivity of PF-07321332 (6) against mammalian and HIV proteases. The inhibitory activity of PF-07321332 was evaluated using FRET-based assay format at several mammalian cysteine (caspase 2, cathepsin L), serine (chymotrypsin, elastase, thrombin) and aspartyl (cathepsin B, cathepsin D, HIV-1) proteases at the indicated protease and substrate concentrations. Data shown represent at least two independent experiments.

| Protease | IC<sub>50</sub> μM |
| --- | --- |
| Human Cathepsin B | >100 |
| Bovine Chymotrypsin | >100 |
| Human Thrombin | >100 |
| Human Caspase 2 | >100 |
| Human Cathepsin D | >100 |
| Human Cathepsin L | >100 |
| Human Immunodeficiency Virus-1 | >100 |
| Human Elastase | >100 |

**Table S4.** PF-07321332 antiviral activity against SARS-CoV-1, 229E and MERS. PF-07321332 inhibited SARS-CoV-1 (N=2), h-CoV-229E (N=3) and MERS (N=1) induced CPE in Vero E6, MRC-5 and Vero 81 cells, respectively as measured by cell viability using ATP as a readout. A P-glycoprotein inhibitor, CP-100356 was added at 2 μM to inhibit the efflux of PF- 07321332 from Vero cells. Cytotoxicity of PF-07321332 was evaluated in non-infected cells and determined as CC50 was >100 μM in Vero E6 plus 2 μM EI (N=2, n=6), MRC-5 (N=3) and Vero 81 plus 2 μM EI (N=1, n=8) cells. Data are expressed as either % effective concentration (EC50 and EC90) relative controls to infected cells with DMSO and non-infected cells. Compound cytotoxicity was calculated relative to DMSO treated and a cytotoxic control compound. Data shown are the geometric mean and 95% confidence intervals (CI).

| Virus | Cell line | EC<sub>50</sub> nM<br>(95% CI) | EC<sub>90</sub> nM<br>(95% CI) | n |
| --- | --- | --- | --- | --- |
| SARS-CoV-1 | Vero E6 + 2 μM<br>CP-100356 | 151 , n=12<br>(108 to 212) | 317, n=12<br>(225 to 446) | 12 |
| 229E | MRC-5 | 190, n=3<br>(58.3 to 621) | 620, n=3<br>(166 to 2320) | 3 |
| MERS | Vero 81 + 1 μM<br>CP-100356 | 166, n=16<br>(145 to 191) | 351, n=16<br>(301 to 409) | 16 |

**Table S5.** Preclinical pharmacokinetics of PF-07321332 (6) ^a^Pharmacokinetic parameters were calculated from plasma concentration–time data and are reported as mean (± S.D. for n=3 and individual values for n=2). All pharmacokinetics were conducted in male gender of each species (Wistar-Han rats and cynomolgus monkey). Intravenous (iv) doses for PF-07321332 (**6**) were administered as a solution in 10% DMSO/30% PEG400/60% deionized water (rats) or 5% (v/v) PEG400:95% (v/v) of 23% 2-hydroxypropyl-β-cyclodextrin in aqueous sodium phosphate buffer pH = 6.0 (monkeys). ^b^Oral (po) pharmacokinetics studies were conducted in the fed state. Oral rat pharmacokinetics studies were conducted with crystalline PF-07321332 (**6**) anhydrous MTBE solvate or anhydrous ’Form 1’. Compound **6**-MTBE solvate and anhydrous ’Form 1’ forms were formulated as solutions in 10% ethanol/10% Capmul MCM/35% PEG400/45% Tween80 and 2% (v/v) Tween80/98% of 0.5% (w/v) methyl cellulose in deionized water, respectively. Rats received 8 ml/kg oral of 1% hydroxypropyl cellulose in deionized water immediately prior to the PF-07321332 (**6**)-MTBE solvate po dose. For po pharmacokinetics assessments in monkeys, crystalline MTBE solvate form of PF-07321332 (**6**) was administered as a solution in 2% (v/v) Tween80/98% of 0.5% (w/v) methyl cellulose in deionized water. ^c^AUC0-t. ^d^Fraction of the po dose absorbed. ^e^PF-07321332 (**6**) was dosed as the anhydrous MTBE solvate. ^f^PF-07321332 (**6**) was dosed as the anhydrous ’Form 1’.

| Species<sup>a</sup> | Dose (mg/kg) | C<sub>max</sub> (ng/ml) | T<sub>max</sub> (h) | AUC<sub>0-∞</sub> (ng·h/ml) | CL<sub>p</sub> (ml/min/kg) | Vd<sub>ss</sub> (l/kg) | t<sub>1/2</sub> (h) | Oral F (%) | F<sub>a</sub> × F<sub>g</sub> (%)<sup>d</sup> |
| --- | --- | --- | --- | --- | --- | --- | --- | --- | --- |
| Rat | 1.0 (iv) | ---- | ---- | 632 (523, 741) | 27.2 (22.5, 31.9) | 1.8 (1.4, 2.2) | 5.1 (3.3, 7.0) | ---- |  |
|  | 10 (po)<sup>b,e</sup> | 1290 (851, 1730) | 1.5 (1.0, 2.0) | 3190 (1890, 4480) | ---- | ---- | 4.0 (2.9, 5.1) | 50 (30, 71) | 95 |
|  | 10 (po)<sup>b,f</sup> | 1450 ± 373 | 0.25 ± 0.0 | 2170 ± 1180 | ---- | ---- | 2.8 ± 1.4 | 34 ± 19 | 65 |
| Monkey | 1.0 (iv) | ---- | ---- | 977 (965, 988) | 17.1 (16.9, 17.3) | 0.33 (0.31, 0.34) | 0.79 (0.74, 0.83) | ---- |  |
|  | 10 (po)<sup>b,e</sup> | 1450 (1050, 1850) | 0.25 (0.25, 0.25) | 831 (747, 914)<sup>c</sup> | ---- | ---- | ---- | 8.5 (7.7, 9.4) | 20 |

**Table S6.** In vitro metabolic stability of PF-07321332 (6) in intestinal microsomes from preclinical species and human. Metabolic stability was examined in intestinal microsomes (in the presence of NADPH) using a PF-07321332 (**6**) concentration of 1 μM. Incubations were conducted in duplicate or triplicate and mean stability data (half-life (t1/2) and intrinsic clearance (CLint) is depicted.

| Compound | Intestinal Microsomes | t<sub>1/2</sub> (min) | CL<sub>int</sub><br>(μl/min/mg) |
| --- | --- | --- | --- |
| PF-07321332 | Rat | >240 | <2.89 |
|  | Monkey | 20.5 | 33.8 |
|  | Human | >240 | <2.89 |

**Table S7.** Human liver microsomal stability of PF-07321332 (6) Metabolic stability of PF- 07321332 (0.1 μM) was examined in HLM (1 mg/ml protein concentration) in the absence or presence of CYP co-factor NADPH. To probe the involvement of CYP3A4 in the metabolic elimination of PF-07321332, NADPH-supplemented HLM were co-incubated with PF-07321332 (0.1 μM) and a selective CYP3A4/5 inhibitor ketoconazole (1 μM) was also included to the incubations. Incubations without NADPH were conducted in duplicate, all other incubations were conducted in triplicate and mean (SD) stability data (half-life (t1/2) and intrinsic clearance (CLint) is depicted in the table.

| Compound | Co-factor | CYP3A4 Inhibitor | t<sub>1/2</sub> (min) | CL<sub>int</sub> (μL/min/mg) |
| --- | --- | --- | --- | --- |
| PF-07321332<br>(6) | - NADPH | - | > 120 | < 5.78 |
|  | + NADPH | - | 28.2<br>(0.2) | 24.5 (0.2) |
|  | + NADPH | + Ketoconazole | > 160 | <4.33 |

**Table S8.**
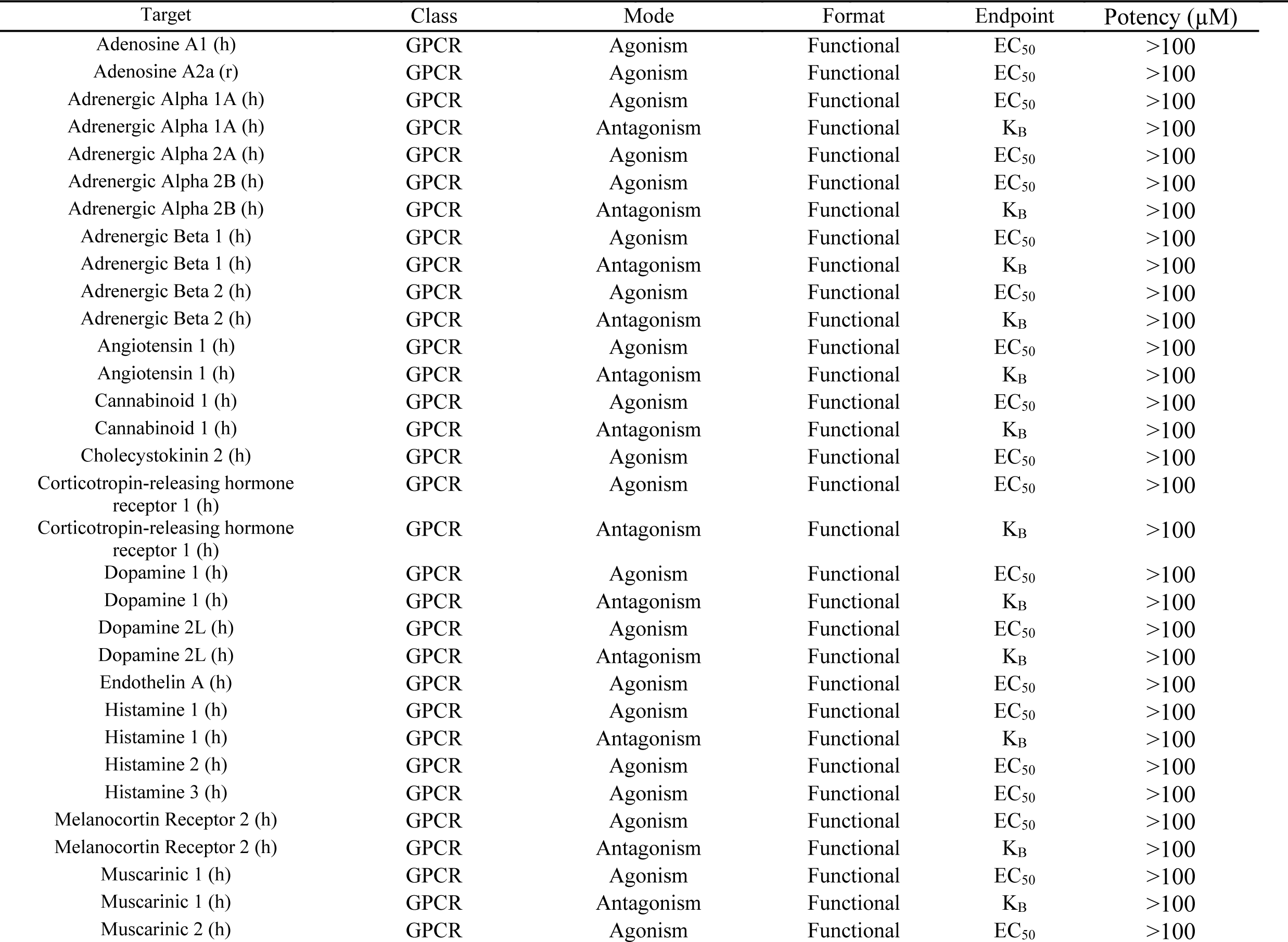
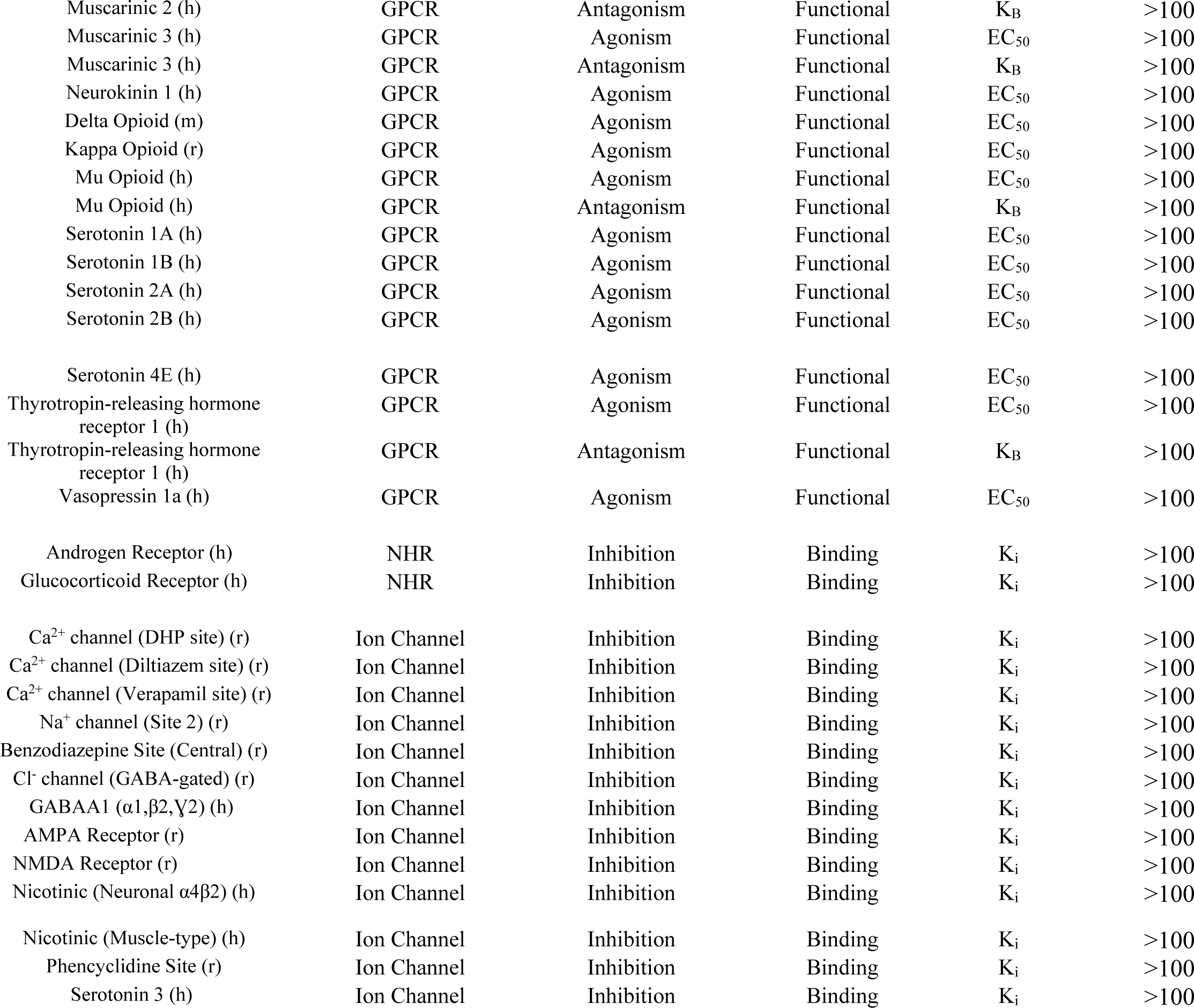
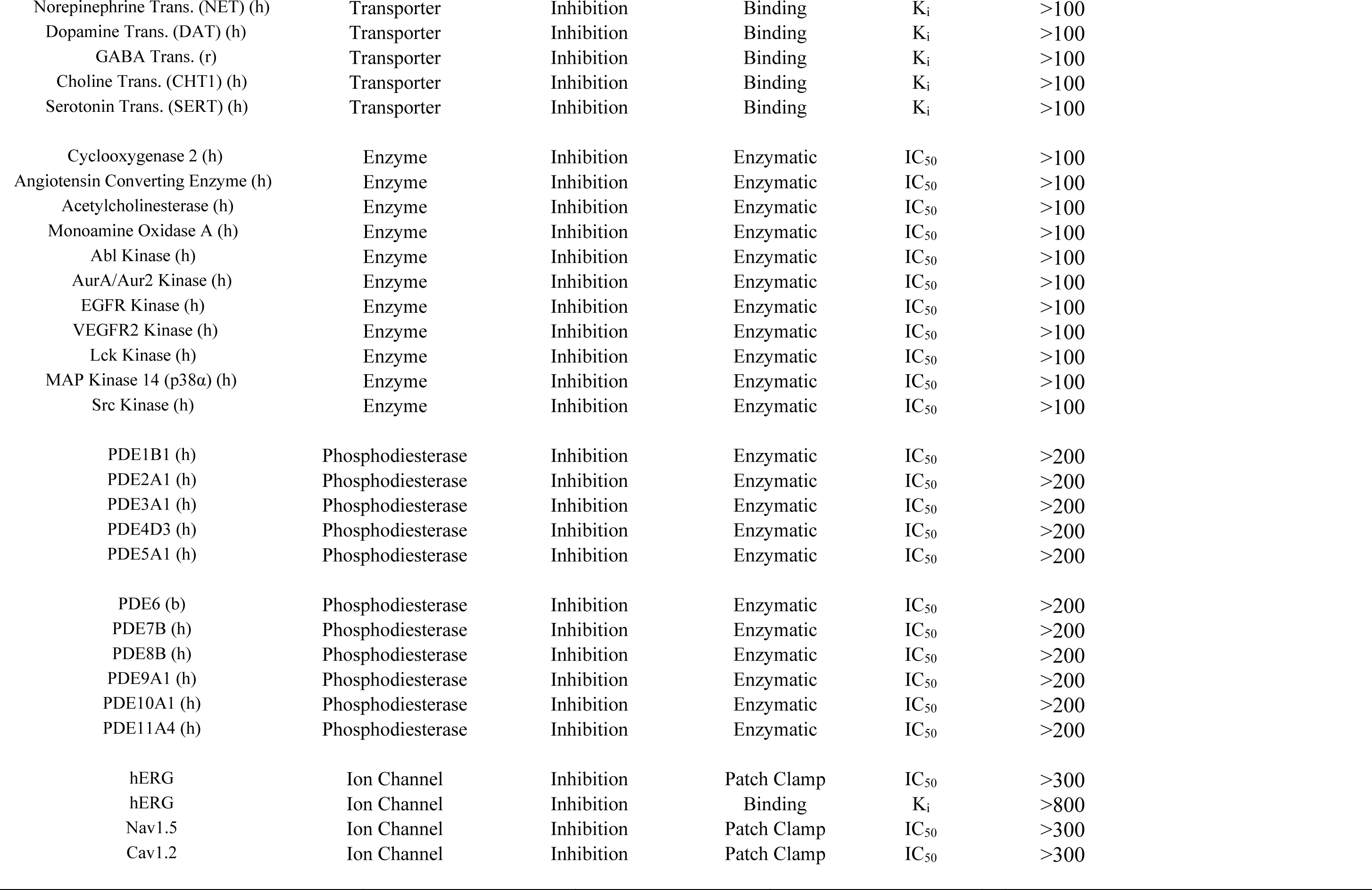
Off target pharmacology profile of PF-07321332 (6) Target species include human (h), rat (r), bovine (b) and mouse (m). Abbreviations: GPCR, G-Protein Coupled Receptor; NHR, Nuclear Hormone Receptor; EGFR, Endothelial Growth Factor Receptor; VEGFR, Vascular Endothelial Growth Factor Kinase; Lck, Lymphocyte-Specific Protein Tyrosine Kinase; MAPK14, Mitogen Activated Protein Kinase 14 (p38α).

| Target | Class | Mode | Format | Endpoint | Potency (μM) |
| --- | --- | --- | --- | --- | --- |
| Adenosine A1 (h) | GPCR | Agonism | Functional | EC<sub>50</sub> | >100 |
| Adenosine A2a (r) | GPCR | Agonism | Functional | EC<sub>50</sub> | >100 |
| Adrenergic Alpha 1A (h) | GPCR | Agonism | Functional | EC<sub>50</sub> | >100 |
| Adrenergic Alpha 1A (h) | GPCR | Antagonism | Functional | K<sub>B</sub> | >100 |
| Adrenergic Alpha 2A (h) | GPCR | Agonism | Functional | EC<sub>50</sub> | >100 |
| Adrenergic Alpha 2B (h) | GPCR | Agonism | Functional | EC<sub>50</sub> | >100 |
| Adrenergic Alpha 2B (h) | GPCR | Antagonism | Functional | K<sub>B</sub> | >100 |
| Adrenergic Beta 1 (h) | GPCR | Agonism | Functional | EC<sub>50</sub> | >100 |
| Adrenergic Beta 1 (h) | GPCR | Antagonism | Functional | K<sub>B</sub> | >100 |
| Adrenergic Beta 2 (h) | GPCR | Agonism | Functional | EC<sub>50</sub> | >100 |
| Adrenergic Beta 2 (h) | GPCR | Antagonism | Functional | K<sub>B</sub> | >100 |
| Angiotensin 1 (h) | GPCR | Agonism | Functional | EC<sub>50</sub> | >100 |
| Angiotensin 1 (h) | GPCR | Antagonism | Functional | K<sub>B</sub> | >100 |
| Cannabinoid 1 (h) | GPCR | Agonism | Functional | EC<sub>50</sub> | >100 |
| Cannabinoid 1 (h) | GPCR | Antagonism | Functional | K<sub>B</sub> | >100 |
| Cholecystokinin 2 (h) | GPCR | Agonism | Functional | EC<sub>50</sub> | >100 |
| Corticotropin-releasing hormone receptor 1 (h) | GPCR | Agonism | Functional | EC<sub>50</sub> | >100 |
| Corticotropin-releasing hormone receptor 1 (h) | GPCR | Antagonism | Functional | K<sub>B</sub> | >100 |
| Dopamine 1 (h) | GPCR | Agonism | Functional | EC<sub>50</sub> | >100 |
| Dopamine 1 (h) | GPCR | Antagonism | Functional | K<sub>B</sub> | >100 |
| Dopamine 2L (h) | GPCR | Agonism | Functional | EC<sub>50</sub> | >100 |
| Dopamine 2L (h) | GPCR | Antagonism | Functional | K<sub>B</sub> | >100 |
| Endothelin A (h) | GPCR | Agonism | Functional | EC<sub>50</sub> | >100 |
| Histamine 1 (h) | GPCR | Agonism | Functional | EC<sub>50</sub> | >100 |
| Histamine 1 (h) | GPCR | Antagonism | Functional | K<sub>B</sub> | >100 |
| Histamine 2 (h) | GPCR | Agonism | Functional | EC<sub>50</sub> | >100 |
| Histamine 3 (h) | GPCR | Agonism | Functional | EC<sub>50</sub> | >100 |
| Melanocortin Receptor 2 (h) | GPCR | Agonism | Functional | EC<sub>50</sub> | >100 |
| Melanocortin Receptor 2 (h) | GPCR | Antagonism | Functional | K<sub>B</sub> | >100 |
| Muscarinic 1 (h) | GPCR | Agonism | Functional | EC<sub>50</sub> | >100 |
| Muscarinic 1 (h) | GPCR | Antagonism | Functional | K<sub>B</sub> | >100 |
| Muscarinic 2 (h) | GPCR | Agonism | Functional | EC<sub>50</sub> | >100 |
| Muscarinic 2 (h) | GPCR | Antagonism | Functional | K<sub>B</sub> | >100 |
| Muscarinic 3 (h) | GPCR | Agonism | Functional | EC<sub>50</sub> | >100 |
| Muscarinic 3 (h) | GPCR | Antagonism | Functional | K<sub>B</sub> | >100 |
| Neurokinin 1 (h) | GPCR | Agonism | Functional | EC<sub>50</sub> | >100 |
| Delta Opioid (m) | GPCR | Agonism | Functional | EC<sub>50</sub> | >100 |
| Kappa Opioid (r) | GPCR | Agonism | Functional | EC<sub>50</sub> | >100 |
| Mu Opioid (h) | GPCR | Agonism | Functional | EC<sub>50</sub> | >100 |
| Mu Opioid (h) | GPCR | Antagonism | Functional | K<sub>B</sub> | >100 |
| Serotonin 1A (h) | GPCR | Agonism | Functional | EC<sub>50</sub> | >100 |
| Serotonin 1B (h) | GPCR | Agonism | Functional | EC<sub>50</sub> | >100 |
| Serotonin 2A (h) | GPCR | Agonism | Functional | EC<sub>50</sub> | >100 |
| Serotonin 2B (h) | GPCR | Agonism | Functional | EC<sub>50</sub> | >100 |
| Serotonin 4E (h) | GPCR | Agonism | Functional | EC<sub>50</sub> | >100 |
| Thyrotropin-releasing hormone receptor 1 (h) | GPCR | Agonism | Functional | EC<sub>50</sub> | >100 |
| Thyrotropin-releasing hormone receptor 1 (h) | GPCR | Antagonism | Functional | K<sub>B</sub> | >100 |
| Vasopressin 1a (h) | GPCR | Agonism | Functional | EC<sub>50</sub> | >100 |
| Androgen Receptor (h) | NHR | Inhibition | Binding | K<sub>i</sub> | >100 |
| Glucocorticoid Receptor (h) | NHR | Inhibition | Binding | K<sub>i</sub> | >100 |
| Ca<sup>2+</sup> channel (DHP site) (r) | Ion Channel | Inhibition | Binding | K<sub>i</sub> | >100 |
| Ca<sup>2+</sup> channel (Diltiazem site) (r) | Ion Channel | Inhibition | Binding | K<sub>i</sub> | >100 |
| Ca<sup>2+</sup> channel (Verapamil site) (r) | Ion Channel | Inhibition | Binding | K<sub>i</sub> | >100 |
| Na<sup>+</sup> channel (Site 2) (r) | Ion Channel | Inhibition | Binding | K<sub>i</sub> | >100 |
| Benzodiazepine Site (Central) (r) | Ion Channel | Inhibition | Binding | K<sub>i</sub> | >100 |
| Cl<sup>-</sup> channel (GABA-gated) (r) | Ion Channel | Inhibition | Binding | K<sub>i</sub> | >100 |
| GABAA1 (α1,β2,γ2) (h) | Ion Channel | Inhibition | Binding | K<sub>i</sub> | >100 |
| AMPA Receptor (r) | Ion Channel | Inhibition | Binding | K<sub>i</sub> | >100 |
| NMDA Receptor (r) | Ion Channel | Inhibition | Binding | K<sub>i</sub> | >100 |
| Nicotinic (Neuronal α4β2) (h) | Ion Channel | Inhibition | Binding | K<sub>i</sub> | >100 |
| Nicotinic (Muscle-type) (h) | Ion Channel | Inhibition | Binding | K<sub>i</sub> | >100 |
| Phencyclidine Site (r) | Ion Channel | Inhibition | Binding | K<sub>i</sub> | >100 |
| Serotonin 3 (h) | Ion Channel | Inhibition | Binding | K<sub>i</sub> | >100 |
| Norepinephrine Trans. (NET) (h) | Transporter | Inhibition | Binding | K<sub>i</sub> | >100 |
| Dopamine Trans. (DAT) (h) | Transporter | Inhibition | Binding | K<sub>i</sub> | >100 |
| GABA Trans. (r) | Transporter | Inhibition | Binding | K<sub>i</sub> | >100 |
| Choline Trans. (CHT1) (h) | Transporter | Inhibition | Binding | K<sub>i</sub> | >100 |
| Serotonin Trans. (SERT) (h) | Transporter | Inhibition | Binding | K<sub>i</sub> | >100 |
| Cyclooxygenase 2 (h) | Enzyme | Inhibition | Enzymatic | IC<sub>50</sub> | >100 |
| Angiotensin Converting Enzyme (h) | Enzyme | Inhibition | Enzymatic | IC<sub>50</sub> | >100 |
| Acetylcholinesterase (h) | Enzyme | Inhibition | Enzymatic | IC<sub>50</sub> | >100 |
| Monoamine Oxidase A (h) | Enzyme | Inhibition | Enzymatic | IC<sub>50</sub> | >100 |
| Abl Kinase (h) | Enzyme | Inhibition | Enzymatic | IC<sub>50</sub> | >100 |
| AurA/Aur2 Kinase (h) | Enzyme | Inhibition | Enzymatic | IC<sub>50</sub> | >100 |
| EGFR Kinase (h) | Enzyme | Inhibition | Enzymatic | IC<sub>50</sub> | >100 |
| VEGFR2 Kinase (h) | Enzyme | Inhibition | Enzymatic | IC<sub>50</sub> | >100 |
| Lck Kinase (h) | Enzyme | Inhibition | Enzymatic | IC<sub>50</sub> | >100 |
| MAP Kinase 14 (p38α) (h) | Enzyme | Inhibition | Enzymatic | IC<sub>50</sub> | >100 |
| Src Kinase (h) | Enzyme | Inhibition | Enzymatic | IC<sub>50</sub> | >100 |
| PDE1B1 (h) | Phosphodiesterase | Inhibition | Enzymatic | IC<sub>50</sub> | >200 |
| PDE2A1 (h) | Phosphodiesterase | Inhibition | Enzymatic | IC<sub>50</sub> | >200 |
| PDE3A1 (h) | Phosphodiesterase | Inhibition | Enzymatic | IC<sub>50</sub> | >200 |
| PDE4D3 (h) | Phosphodiesterase | Inhibition | Enzymatic | IC<sub>50</sub> | >200 |
| PDE5A1 (h) | Phosphodiesterase | Inhibition | Enzymatic | IC<sub>50</sub> | >200 |
| PDE6 (b) | Phosphodiesterase | Inhibition | Enzymatic | IC<sub>50</sub> | >200 |
| PDE7B (h) | Phosphodiesterase | Inhibition | Enzymatic | IC<sub>50</sub> | >200 |
| PDE8B (h) | Phosphodiesterase | Inhibition | Enzymatic | IC<sub>50</sub> | >200 |
| PDE9A1 (h) | Phosphodiesterase | Inhibition | Enzymatic | IC<sub>50</sub> | >200 |
| PDE10A1 (h) | Phosphodiesterase | Inhibition | Enzymatic | IC<sub>50</sub> | >200 |
| PDE11A4 (h) | Phosphodiesterase | Inhibition | Enzymatic | IC<sub>50</sub> | >200 |
| hERG | Ion Channel | Inhibition | Patch Clamp | IC<sub>50</sub> | >300 |
| hERG | Ion Channel | Inhibition | Binding | K<sub>i</sub> | >800 |
| Nav1.5 | Ion Channel | Inhibition | Patch Clamp | IC<sub>50</sub> | >300 |
| Cav1.2 | Ion Channel | Inhibition | Patch Clamp | IC<sub>50</sub> | >300 |

**Table S9.**
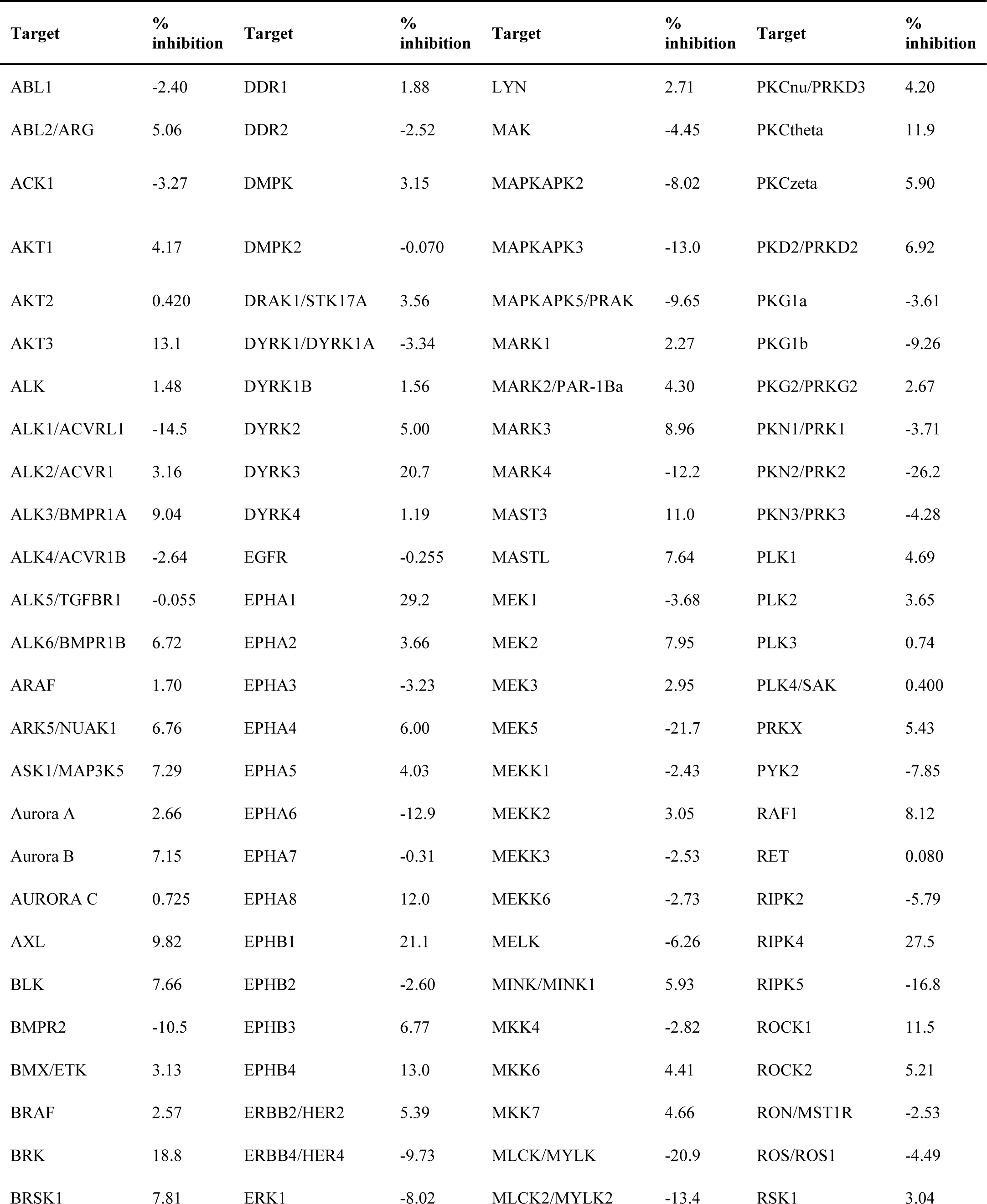
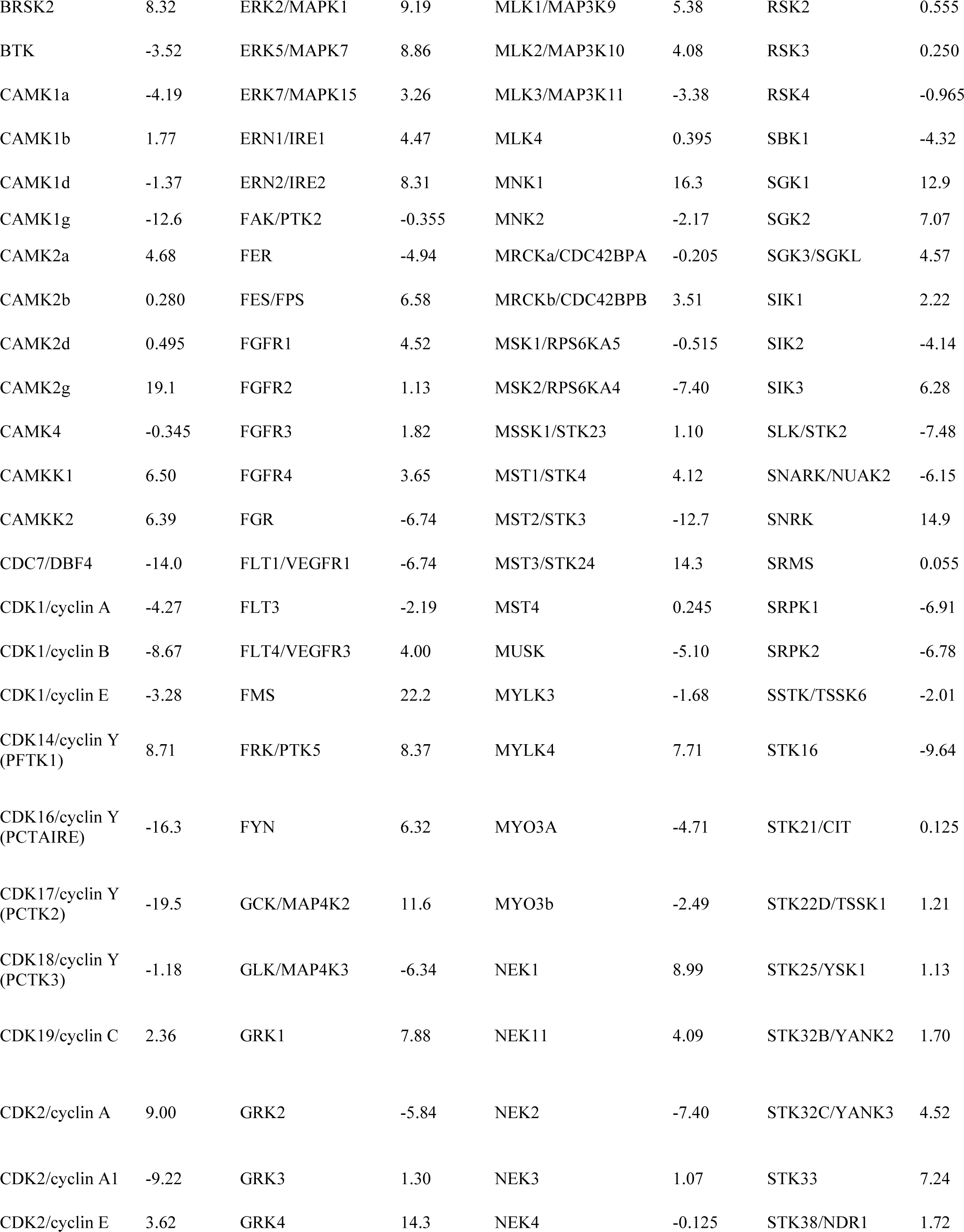
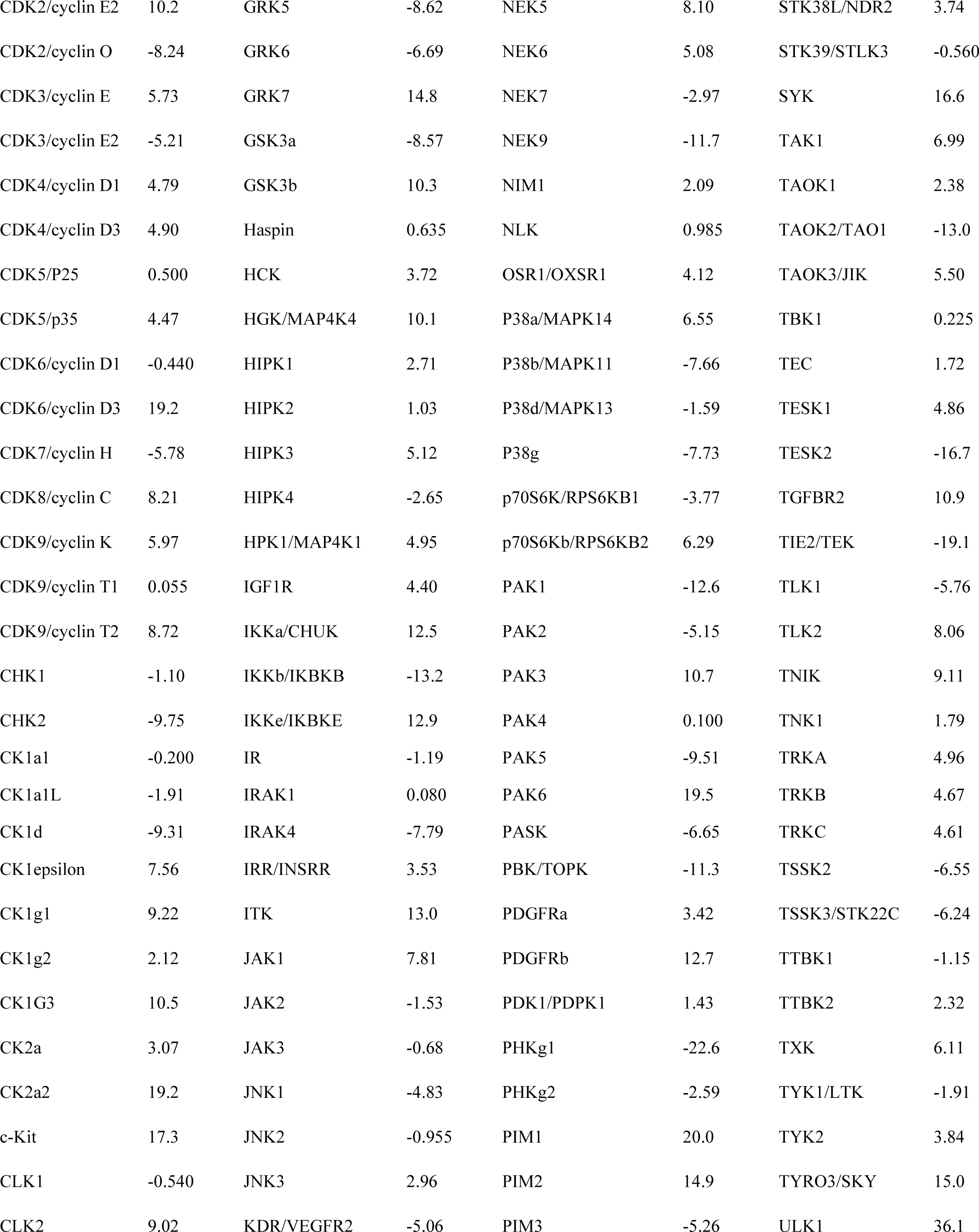
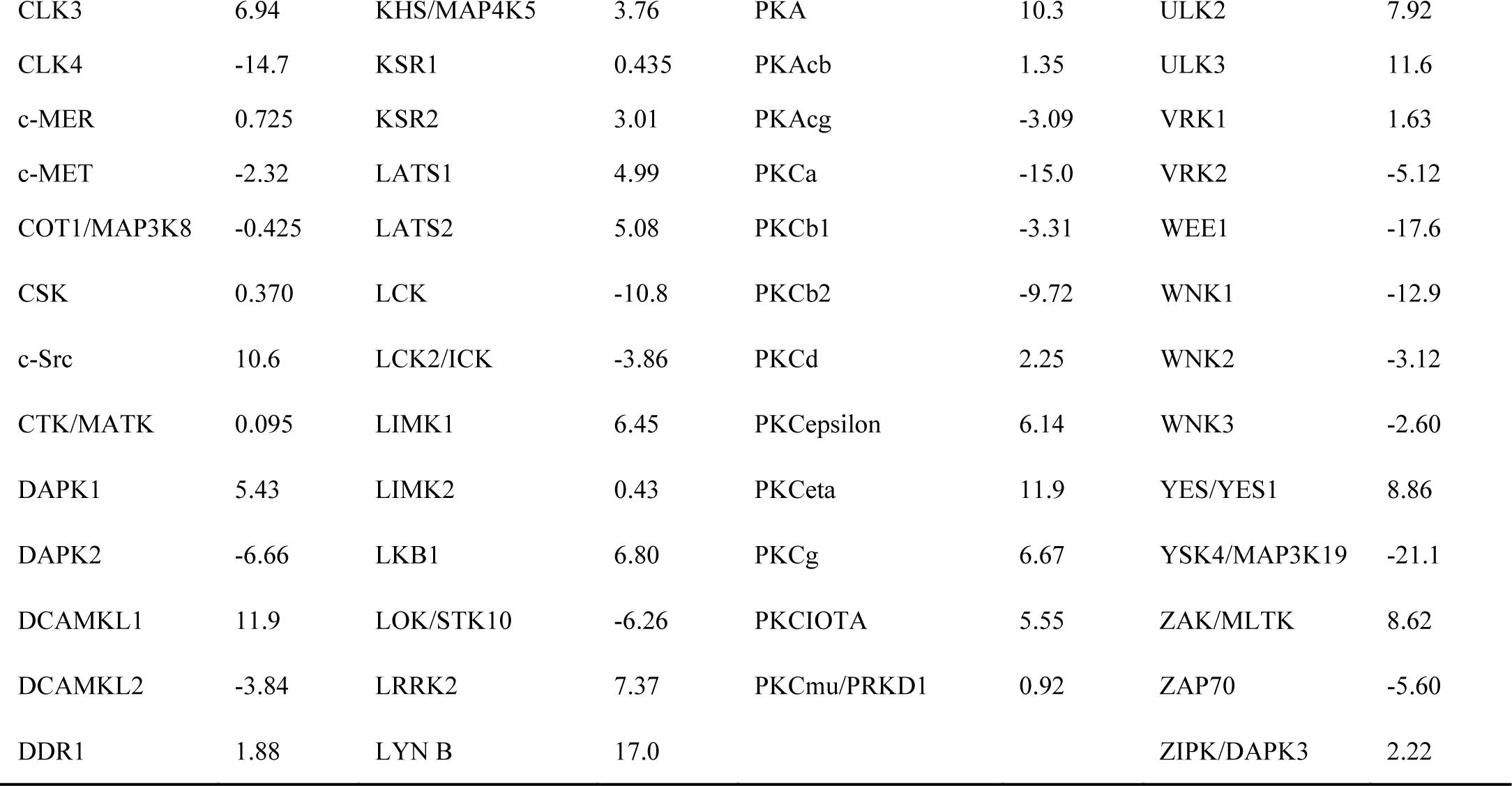
Kinase profile of PF-07321332 (6) Kinase inhibition studies were conducted by Reaction Biology (Malvern, PA, USA) on behalf of Pfizer Inc. The inhibitory activity of 10 µM PF-07321332 was assessed against a broad panel of kinases using the HotSpot™ kinase assay format in the presence of 10 µM ATP. Data are presented as the average of two technical replicates.

**Table S10.**
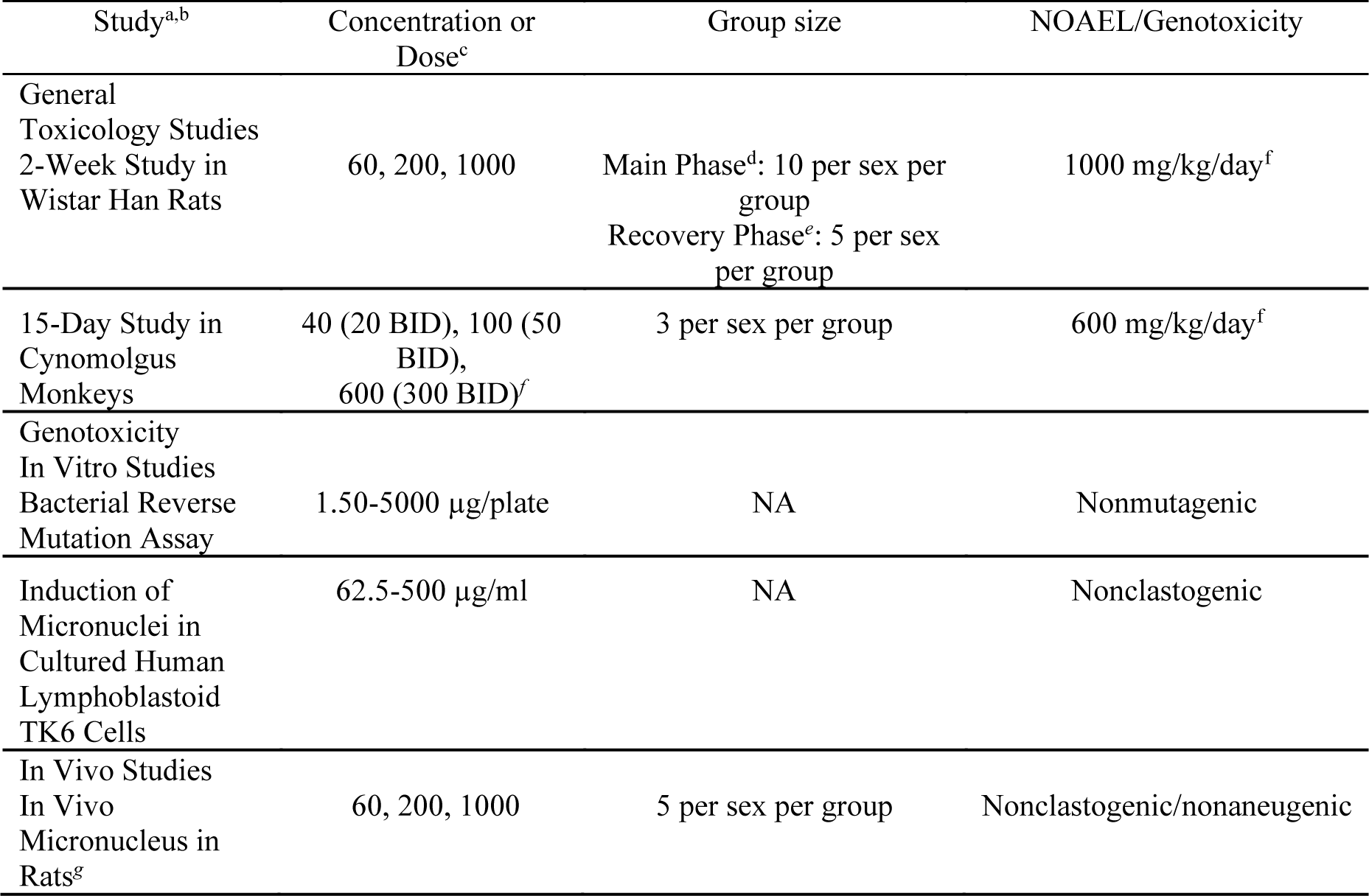
PF-07321332 (6) IND-enabling nonclinical safety studies. ^a^All GLP studies were conducted in an OECD MAD compliant member state. ^b^All in vivo studies were conducted with male and female animals; compound was administered orally. ^c^All doses expressed as mg/kg unless otherwise specified. ^d^Main phase rats were administered vehicle or PF-07321332 for 2weeks followed by scheduled necropsy. ^e^Recovery phase rats were administered vehicle or PF-07321332 for 2 weeks followed by a post dose phase of 2 weeks wherein no vehicle or drug was administered. At end of recovery phase, rats underwent scheduled necropsy. ^f^The interval between BID dosing sessions was approximately 6 hours. ^g^Conducted as part of the 2-week pivotal rat study. ^f^Associated plasma concentrations from the 2-week rat and monkey toxicity studies are presented in Table S11.

| Study<sup>a,b</sup> | Concentration or Dose<sup>c</sup> | Group size | NOAEL/Genotoxicity |
| --- | --- | --- | --- |
| General Toxicology Studies |  |  |  |
| 2-Week Study in Wistar Han Rats | 60, 200, 1000 | Main Phase<sup>d</sup>: 10 per sex per group<br>Recovery Phase<sup>e</sup>: 5 per sex per group | 1000 mg/kg/day<sup>f</sup> |
| 15-Day Study in Cynomolgus Monkeys | 40 (20 BID), 100 (50 BID),<br>600 (300 BID)<sup>f</sup> | 3 per sex per group | 600 mg/kg/day<sup>f</sup> |
| Genotoxicity In Vitro Studies |  |  |  |
| Bacterial Reverse Mutation Assay | 1.50-5000 µg/plate | NA | Nonmutagenic |
| Induction of Micronuclei in Cultured Human Lymphoblastoid TK6 Cells | 62.5-500 µg/ml | NA | Nonclastogenic |
| In Vivo Studies |  |  |  |
| In Vivo Micronucleus in Rats<sup>g</sup> | 60, 200, 1000 | 5 per sex per group | Nonclastogenic/nonaneugenic |

**Table S11.** Mean toxicokinetics of PF-07321332 (6) in rats and monkeys after 2-weeks of repeat dosing. ^a^AUC24 = Area under the concentration-time curve from time t=0 to t=24 h, Cmax = Maximum plasma concentration, Tmax = time at which Cmax was first observed. Toxicokinetics parameters are mean values (± standard deviation) combining male and female animals. MTBE solvate of crystalline PF-07321332 (**6**) was formulated in 2% (v/v) polysorbate 80 in 0.5% (w/v) methylcellulose. ^b^15 animals/sex/dose group (non-serial sampling at *n*=5/sex/dose group/timepoint). Animals were dosed once daily for 14 days. ^c^3 animals/sex/dose group. Animals were dosed orally twice daily for 15 days; the interval between dosing sessions was approximately 6 hours. Mean Tmax not reported due to twice daily dosing format. ^d^Unbound Cmax and unbound AUC24 values were estimated by multiplying the total values with the plasma unbound fraction (fu,p) of PF-07321332 (**6**) in rats (fu,p = 0.479) and monkeys (fu,p = 0.435). BID, twice daily; QD, once per day.

| Species<sup>a</sup> | Day | Daily Oral Dose<br>(mg/kg) | C<sub>max</sub> (μg/mL) |  | T<sub>max</sub> (h) | AUC<sub>24</sub><br>(μg·h/mL) |  |
| --- | --- | --- | --- | --- | --- | --- | --- |
|  |  |  | total | Unbound<sup>d</sup> |  | total | Unbound<sup>d</sup> |
| **Rat<sup>b</sup>** | 1 | 60 QD | 12.9 ± 1.9 | 6.18 | 0.5 ± 0.0 | 27.3 ± 2.4 | 13.1 |
|  |  | 200 QD | 37.0 ± 4.0 | 17.7 | 0.5 ± 0.0 | 291 ± 4 | 139 |
|  |  | 1000 QD | 62.1 ± 5.5 | 29.7 | 2.0 ± 1.5 | 796 ± 166 | 381 |
|  | 14 | 60 QD | 13.3 ± 5.0 | 6.37 | 0.5 ± 0.0 | 17.2 ± 5.1 | 8.24 |
|  |  | 200 QD | 27.1 ± 3.3 | 13.0 | 0.5 ± 0.0 | 80.5 ± 27.4 | 38.6 |
|  |  | 1000 QD | 51.5 ± 0.9 | 24.7 | 2.0 ± 0.0 | 292 ± 8 | 140 |
| **Monkey<sup>c</sup>** | 1 | 20 BID | 1.79 ± 0.89 | 0.779 | 1.3 ± 0.6 | 10.4 ± 5.7 | 4.52 |
|  |  | 50 BID | 11.3 ± 6.1 | 4.92 | 1.3 ± 1.3 | 84.2 ± 57.5 | 36.6 |
|  |  | 300 BID | 59.6 ± 11.8 | 25.9 | 3.0 ± 1.1 | 723 ± 152 | 315 |
|  | 15 | 20 BID | 2.42 ± 0.50 | 1.05 | 0.67 ± 0.26 | 9.61 ± 3.05 | 4.18 |
|  |  | 50 BID | 11.8 ± 6.6 | 5.13 | 0.92 ± 0.58 | 52.6 ± 28.8 | 22.9 |
|  |  | 300 BID | 106 ± 18 | 46.1 | 3.7 ± 0.8 | 1220 ± 224 | 531 |

## Notes

### Clinical Trial

NCT04756531

### Author Declarations

ADVARRA/IntegReview Ethical Review Board

